# Case Study: End User Development of an Agent-based Model of Malaria Transmission to Support the Design of Late-Life-Acting Insecticides for the Control of Malaria Transmission and Delay of the Evolution of Insecticide Resistance

**DOI:** 10.1101/2021.05.28.21257999

**Authors:** Jacob H Heintzelman, Gregory R Madey

## Abstract

We describe an end-user developed agent-based simulation of malaria transmission. The simulation’s development is a case study demonstrating an approach for non-technical investigators to easily develop useful simulations of complex public health problems. We focused on malaria transmission, a major global public health problem, and insecticide resistance (IR), a major problem affecting malaria control. Insecticides are used to reduce transmission of malaria caused by the *Plasmodium* parasite that is spread by the *Anopheles* mosquito. However, the emergence and spread of IR in a mosquito population can diminish the insecticide’s effectiveness. IR results from mutations that produce behavioral changes or biochemical changes (such as detoxification enhancement, target site alterations) in the mosquito population that provide resistance to the insecticide. Evolutionary selection for the IR traits reduces the effectiveness of an insecticide favoring the resistant mosquito population. It has been suggested that biopesticides, and specifically those that are Late Life Acting (LLA), could address this problem. LLA insecticides exploit *Plasmodium’s* approximate 10-day extrinsic incubation period in the mosquito vector, a delay that limits malaria transmission to older infected mosquitoes. Since the proposed LLA insecticide delays mosquito death until after the exposed mosquito has a chance to produce several broods of offspring, reducing the selective pressure for resistance, it delays IR and gives the insecticide longer effectivity. Such insecticides are designed to slow the evolution of IR thus maintaining their effectiveness for malaria control. For the IR problem, the simulation shows that an LLA insecticide could work as intended, but its operational characteristics are critical, primarily the mean-time-to-death after exposure and the associated standard deviation. We also demonstrate the simulation’s extensibility to other malaria control measures, including larval source control and policies to mitigate the spread of IR. The simulation was developed using NetLogo as a case study of a simple but useful approach to public health research.

## Introduction

It has recently been argued that modeling and simulation can be useful tools for public health research and innovation, but have yet to be used to their potential [1]. One recommended solution to this challenge is to increase the participation of the interested parties in the modeling and simulation development process, including the stakeholders, scientists, researchers and health professionals [1, 2]. Modeling and simulation can use many approaches, including differential equation-based or system dynamics approaches, statistical or data science approaches, and agent-based approaches. This study used the latter, the agent-based modeling (ABM) approach, which is highly suitable for end-user participation in the simulation process [3]. As faster computers and more intuitive programming tools become increasing available, and typical users become more comfortable with computing, such end-users can become more involved as the developers of simulations [4], including agent-based simulations [5]. We describe in this paper a case study of an end-user developed malaria transmission simulation, and demonstrate its usefulness for evaluating a potential solution to a major challenge for malaria control.

The use of chemical insecticides to control malaria vectors goes back at least to the 1940s and reported loss of their effectiveness caused by the vectors’ insecticide resistance (IR) goes back to the 1970’s [6, 7]. Malaria, a disease caused by the parasitic protist *Plasmodium*, is one of the major causes of death globally, especially in developing countries. There were 409,000 deaths and 229 million infections across the world from the disease in 2019 [8]. Mosquito vectors transmit malaria by taking a blood meal from an infectious human and, after an extrinsic incubation period (EIP), biting a susceptible human, releasing the parasite into their bloodstream. Transmission control can use chemical interventions to kill the mosquitoes that transmit the disease. These interventions primarily take the forms of insecticide treated bed-nets, which provide both a physical barrier and insecticide killing, and indoor residual spraying, which consists of an insecticide application inside human habitations. The efficacy of these interventions can be seen in the 27 countries which have improved to less than 100 yearly cases of malaria since 2010 [8].

However, these chemical interventions are losing their effectiveness due to the evolution of IR in the mosquitoes causing them to be less affected by the current chemicals [7, 9, 10]. Once IR emerges in a vector population, the insecticide interventions become effective at reducing the mosquito population and controlling malaria [11, 12]. To combat this loss of effectiveness, new interventions are proposed to replace those that are no longer effective. These interventions can be evaluated using mathematical and computer modeling to observe their effects before such interventions are designed, developed and produced [13–17]. Such model-based evaluations can help specify performance attributes for the proposed interventions before they enter more costly development and field evaluation. However, many computer models are complex and hard for anyone other than specialists to use and understand. They also can take a long time to develop or adapt, which can reduce their utility during the insecticide’s development. While there have been many agent-based malaria transmission models described in the literature [13, 15, 18–26], most are not free-open source, and experience shows that most users of the notable free-open source models actually reside in the organization that sponsored their development [13, 26].

We identify multiple challenges for the users of modeling and simulation of malaria transmission, including: 1) requirement of either strong mathematical or computer programming skills, 2) the traditional complexity of modeling and simulation, 3) long development time, 4) lack of ease of use and extensibility, 5) lack of transparency of assumptions and limitations, and often 6) the need for high performance computing resources.

One approach to address these six challenges is to develop models employing an easy-to-use ABM tool. These tools can simulate a sufficient level of needed problem complexity, while remaining simple enough to develop in a shorter time by a wider range of researchers compared to other modeling methods. These ABM tools enable relatively easy development of useful simulations of problems of interest and are accessible for use by non-technical investigators. Many reviews of these ABM tools, also called ABM platforms, are available [27–31]. Many are free-open source, require lower levels of development effort, and can be used for a wide range of applications [27].

This paper describes an ABM of malaria transmission developed using the popular simulation development tool NetLogo [32] demonstrating an approach for non-technical investigators to develop useful simulations of many complex problems. As a demonstration, we investigate a proposed insecticide that uses a novel mechanism to control the mosquito population and hence malaria transmission, while delaying the evolution of resistance to that intervention. The proposed intervention is a late-life-acting (LLA) insecticide [33, 34]. This intervention targets the problem of IR by being more selective in the timing between exposure and mortality than traditional insecticides for two key reasons. First, the *Plasmodium* parasite has an approximately 8 to 12-day EIP (the phase of the parasite’s life-cycle within the mosquito) meaning that the infectious mosquito does not need to die until at least 8-days after it has taken a blood-meal from an infectious person to block transmission of the disease. Second, this extra time also allows the mosquito to complete several gonotrophic cycles (i.e., the cycle of feeding, resting, and ovipositing), producing offspring with its genotype before they die, thus reducing the selective pressure on the resistant allele. Most insecticides kill the insect on or shortly after contact – referred to as Instant Acting (IA) – meaning that only the resistant insects are able to reproduce and pass on their genotype when exposed to the IA insecticide. An LLA insecticide slows the spread of IR by allowing the insecticide-susceptible mosquitoes to be more competitive at passing on their insecticide-susceptible genes, resulting in a longer-lasting insecticide-susceptible population.

One type of proposed LLA insecticide is based on using an entomopathogenic fungus – a biopesticide – which could provide the resistance delaying benefits of an LLA insecticide [33–40]. The application of these fungi has been evaluated in several field tests which have shown that the fungi can be applied effectively and work to kill mosquitoes in the field but have never been tried on a large enough scale to test their effect on malaria transmission [39, 41]. Mosquitoes are frequently in natural contact with fungi and do have ways to adapt to them in nature. Fungi can enter the mosquito from several points: ingestion, cutaneous openings, and forced introduction [37, 38]. Although their innate immune system is not capable of adapting to specific invaders [42], it is assumed that a mutation in the mosquitoes’ genome could confer a boosted immune system, causing the mosquito to become resistant to the fungal insecticide.

However, some potential drawbacks with LLA insecticides have been identified. The female mosquito may not be exposed to the insecticide until her second or third gonotrophic cycle so that she may still live long enough to cause an infectious bite. Even if the mosquito is exposed to the LLA insecticide during its first gonotrophic cycle, there may be a wide variability in the timing of the mosquito’s death, possibly resulting in reduced malaria control or a shortened delay of the emergence of IR. To examine such problems, we constructed an ABM of the malaria transmission cycle. We then applied this model by using it to perform a series of tests on hypothetical properties of an LLA insecticide [34]. This study utilized the simplicity, ease and speed of development of the NetLogo tool to quickly create a useful model.

The model presented in this paper is agent-based (sometimes called individual-based) because it explicitly represents in the simulation important elements of the modeled system (the agents or individual actors in the system). ABM is an alternative to mathematical modeling, which often models population dynamics using differential equations. ABM can simulate the actions and events in the life-cycle of each distinct individual agent, including its interactions with other agents, to simulate more detailed and representative behavior of the modeled system. In the model there are six types of agents: 1) the adult female mosquito agents, 2) the brood agents (representing the aquatic stages of mosquito cohorts), 3) the susceptible or exposed (but not yet infectious) human agents, 4) infectious human agents, 5) recovered human agents, and 6) the patch agents (the square spaces that make up the simulated world of the simulation). Each of the agents can have its own unique attributes, individual behaviors, and distinct interactions with the other agents. The malaria parasite was modeled as an attribute possessed by the mosquitoes and humans and not as an explicit agent.

This paper describes our results for two interrelated objectives: 1) a case study of how end users can develop agent-based simulation to be used by public health researchers to support their investigations, and 2) discover through simulation insights into the potential utility of LLA insecticides as a proposed strategy for addressing IR. The challenge of IR is not limited to malaria control and insights presented here may be applicable to other IR problem areas (e.g., other arthropod transmitted diseases and agricultural applications). The complete EMMIT (Extendible Model for Malaria Intervention Testing) program presented in this paper is included as a supporting information file and includes source code and documentation, permitting modifications and extension to other malaria control research topics. Additional supporting information files include 1) formal documentation using the ODD standard, 2) a table of simulation parameters, and 3) a compilation of calibration, verification, and validation experiments conducted on the model. The following sections present a description of a model of malaria transmission, the IA and LLA insecticide malaria control methods simulated in the model, our representation of the mosquitoes’ resistance genotypes, implementation details of the EMMIT program, and results and analysis.

## Model

The EMMIT program consists of mosquito agents, brood agents, three types of human agents, a simulated world for the agents to occupy consisting of discrete patches, villages, water patches representing breeding sites, and interventions designed to reduce the number of mosquito agents. The simulated world is conceptually divided into an aquatic habit (where the broods reside) and a terrestrial habitat (where the adult mosquitoes and humans reside).

### Mosquito Agents

We modeled the mosquitoes’ life cycle in two phases: the aquatic phase and the adult phase. In the aquatic phase of the life cycle, one brood agent represents all of the eggs associated with an oviposition. The brood agent represents all three aquatic stages (eggs, larvae, pupae) and has a lifespan equivalent to the sum of the time spent in each stage. For each gonotrophic cycle, the mosquitoes produce one brood. When each brood agent reaches the specified age, it is replaced by 15 - 50 juvenile female adult mosquito agents, representing the survivors of the aquatic life stage. These newly emerged mosquitoes move randomly for about two days, after which they are assumed to have mated, become mature female adult mosquitoes, and begin seeking a blood meal (the beginning of a gonotrophic cycle). After successfully taking a blood meal, a period of time passes representing the mosquito digesting the meal and the preparation of a brood of eggs. This is followed by search and selection of a water site for oviposition. After successful oviposition, the mosquito begins a new gonotrophic cycle. In each gonotrophic cycle the mosquito may be infected by the *Plasmodium* parasite. Male mosquitoes are not explicitly modeled as agents, but are implicitly modeled for determining the genotype of the offspring. See Fig 1.

**Fig 1.**
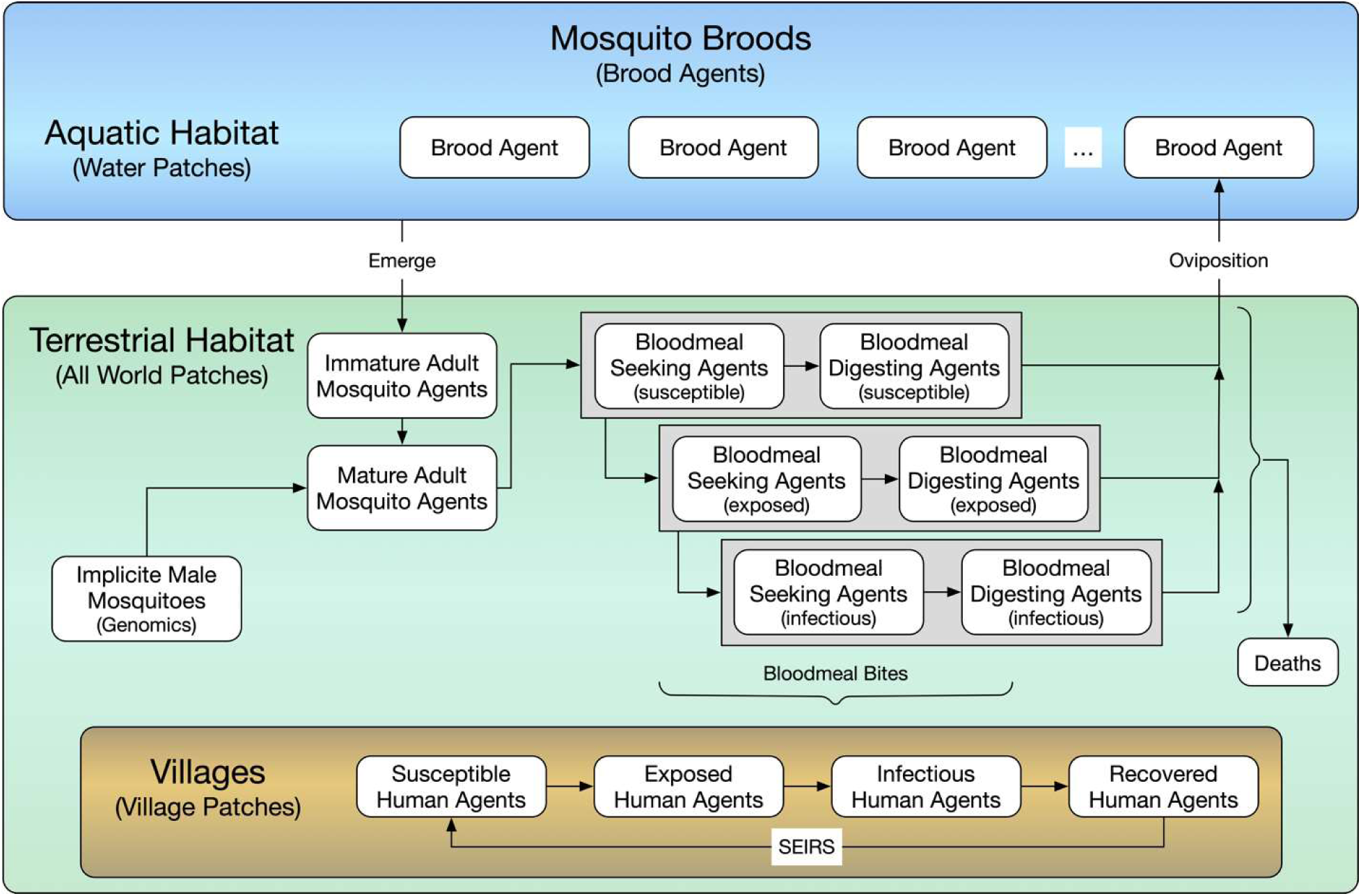
Modeled mosquito life cycle and human disease cycle. The simulated mosquitoes begin as a brood agent in the water patches of the simulated landscape until they emerge as immature mosquito agents wandering until they mature. Once matured (about two days later), the mosquito agents are assumed to have mated and begin searching for a blood meal. Once they take a blood meal, the simulated mosquitoes begin blood digesting then begin searching for a water patch to lay their eggs. If they bite an infectious person, they become exposed and then infectious. Once the simulated mosquito finds a water patch it creates a new brood agent and resumes the blood meal search. The human agents are *susceptible* until they receive a bite from an infectious mosquito, they then enter the *exposed* state, after which they transition to the *infectious* state. After the specified time, they transition from infectious to *recovered* and then back to *susceptible*.

The adult mosquitoes in the baseline model have a simple one percent hourly chance of mortality from natural causes (predators, age, environmental hazards, etc.). Since we only modeled the hours when the mosquitoes are active in our simulation (12-hours per day), this gave the mosquitoes a slightly less than 12% daily death rate causing an exponential decline in the mosquito population age structure. See Fig 4 in the Results section.

Malaria dynamics in the mosquito population were implemented using the Susceptible-Exposed-Infectious model. The mosquitoes are susceptible until they take a blood meal from an infectious human (with a given probability of infection), then they entered the exposed state with an EIP of 8-12 days [34, 43, 44], after which they became infectious for the rest of their lives.

The mosquito agents are able to bite one of the human agents when they are on the same patch and the mosquito is not gravid. When taking a blood meal from a human agent a separate procedure determines if the human they bite is infectious, and hence if they become infected. After the bite procedures, the mosquito agent is considered gravid, begins bloodmeal digesting, and seeking a water patch for egg laying. After a burn-in period (running the simulation until it reaches equilibrium), the simulation records the total number of bites, the hourly bites, the total bites that infect humans, and the hourly bites that infect humans.

The mosquito agents in our model move every timestep. If the non-gravid mosquitoes are within fifty patches of the center of either of the villages they move generally (although not directly) towards the village to take a blood meal. Outside of fifty patches from the villages, the mosquitoes move randomly. The gravid mosquitoes move randomly at a constant rate until they are within four patches of an open water patch. They then move to this patch and leave a brood agent there, and begin a new gonotrophic cycle searching for another blood meal.

### Humans Agents

Individual human agents all reside in one of two villages each represented in the model by a circle of patches. Human births and deaths are not modeled and the total number of human agents stays constant throughout the simulation’s duration. No additional attributes are assigned, such as co-morbidities, preventative drug administration, age, sex, or pregnancy. The human agents move randomly within the village patches. Some humans move linearly in and out their village to simulate time that members of the villages spend traveling, tending to crops, and other activities outside of the village. To simulate the human disease states in our model we assumed the SEIRS (Susceptible, Exposed, Infectious, Recovered, Susceptible) disease model (see Fig 1). We modeled the SEIRS states using three types of human agents: 1) *susceptible* (which includes *exposed*), 2) *infectious*, and 3) *recovered*. The human agents cycle between these states when receiving infectious bites and after a delay. The length that the human agent spends in each state is sampled randomly from a normal distribution. In the event of a bite, if the human is *susceptible* and the mosquito is infectious, then the human becomes *exposed*. After a delay to represent the time which the parasite spends in the human liver (the exo-erythrocytic cycle) and the human blood (erythrocytic cycle), the human becomes an *infectious* agent. If the bitten human is *infectious*, the mosquito becomes exposed but not infectious until after the EIP (the sporogonic cycle) [44]. If the human is *recovered*, they can receive infectious bites, but are considered immune and do not change states. Then, after a random delay, the recovered human agents switch to the *susceptible* state where they are can receive infectious bites and become *exposed* and then *infectious* again. The mean of the time spent in the recovered and infected states is an input value which can be set in the interface.

### Insecticides

Both LLA and IA insecticides are modeled. To model the use of the LLA insecticide, we randomly assigned patches across the villages to be treated with the insecticide with coverage percentage specified by the user. When the mosquitoes encountered these patches, they have a random chance of exposure to the insecticide. If they are exposed, they are assigned a time-to-death. This time-to-death is a normally distributed random number with adjustable mean and standard deviation used to evaluate the performance of the LLA. We evaluated multiple means from 7 to 11 days after exposure to evaluate the optimal delay time to death [37, 38]. We then evaluated multiple standard deviations to see how precise the timing of an LLA insecticide needed to be to reduce malaria transmission and simultaneously slow the spread of resistance. We modeled the effects of this LLA based on results from field tests of entomopathogenic fungi’s effect on mosquitoes [37, 38]. For example, after exposure to the LLA, we modeled a decrease in the mosquito’s flight speed, representing the growing burden of the fungal infection prior to death [40].

We also modeled an IA insecticide representative of those currently approved for use by the WHO [45]. This enabled us to calibrate our model using empirical studies conducted on these insecticides [46, 47]. This also allowed us to compare the effectiveness of approved IA insecticides to the LLA insecticide. The IA insecticide was implemented in the model in the same way as the LLA insecticide: patches of insecticide were randomly distributed around the villages and when the mosquitoes encountered them, they had a certain probability of exposure and death. The difference is that, for the IA insecticide, the mosquitoes die immediately after exposure.

Several variables are critical in determining the functionality of both insecticides. These include: the chance of exposure, the resistance dominance of the heterozygous genome [16, 17], the chance of death after exposure (IA), the mean and standard deviation of the kill time (LLA), the fitness cost on the resistant mosquitoes, and the insecticide coverage of the village. All of these variables can be set through the interface of the program.

### Source Control Extension

To demonstrate the flexibility and extensibility of the program, a simple source control feature was added to evaluate the reduction of breeding sites. When this feature is on, a variable percent of the water patches within the anulus of the village are removed to simulate the draining and removal of mosquito breeding sites near the villages. The distance that this intervention extends from the village is controlled by a slider on the user interface. The accuracy of this intervention is also controllable on the interface and can be set so that only a certain percent of patches in the intervention area are removed. The patches that become part of this intervention never become breeding sites again in the simulation. With this feature on, the number of people who leave the village is increased to simulate people going out to implement and maintain the source control measures. This extension was added with about 10 lines of code. See the S4 document for details on the results of this feature.

### Resistance Mitigation Policy Extension

A second demonstration the extensibility of our model was a feature which evaluates an adaptive policy for the managing the use of an insecticide as IR emerges. When activated, this feature controls the amount of insecticide treated patches in the model based on how prevalent the resistant allele is in the mosquito population. As resistance to the insecticide increases, this IR mitigation policy iteratively reduces the number of treated patches until no insecticide is applied (so that the selective pressure for resistance goes down). Then, it waits for the percent of resistant mosquitoes to return to a baseline level and reintroduces the insecticide at full strength. This feature relies on a high fitness cost of resistance to cause the resistant population to decrease after the insecticide is removed. This extension was added with about 35 lines of code. See the S4 document for details on the results of this feature.

### Agent Space

The agent space consists of 40,401 patches arranged in a 2-dimensional 201×201 grid. This is intended to represent 100m^2^ per patch for a simulated space of approximately 4 km^2^. The edges of the space wrap horizontally and vertically forming a torus. Two village areas (red) are set equidistant from the center of the space (see Fig 2). In these villages, the people move randomly. Water patches are randomly spread across the space controlled by a slider on the interface to specify their initial frequency. During the simulation, the water patch count decreased or increased seasonally with two peaks each year to simulate seasonal rainfall. Each water patch has a carrying capacity of three broods. The mosquitoes move randomly across the map unless they are within 50 patches (adjustable in the interface) of one of the villages in which case they move generally towards the village until they encounter a human and take a blood meal. After the blood meal, the mosquitoes move randomly out of the village until they find a water patch. If they are within 4 patches of a water patch, they move straight to the water where, and if their gonotrophic cycle is completed, they lay a brood of eggs and continue random flight.

**Fig 2.**
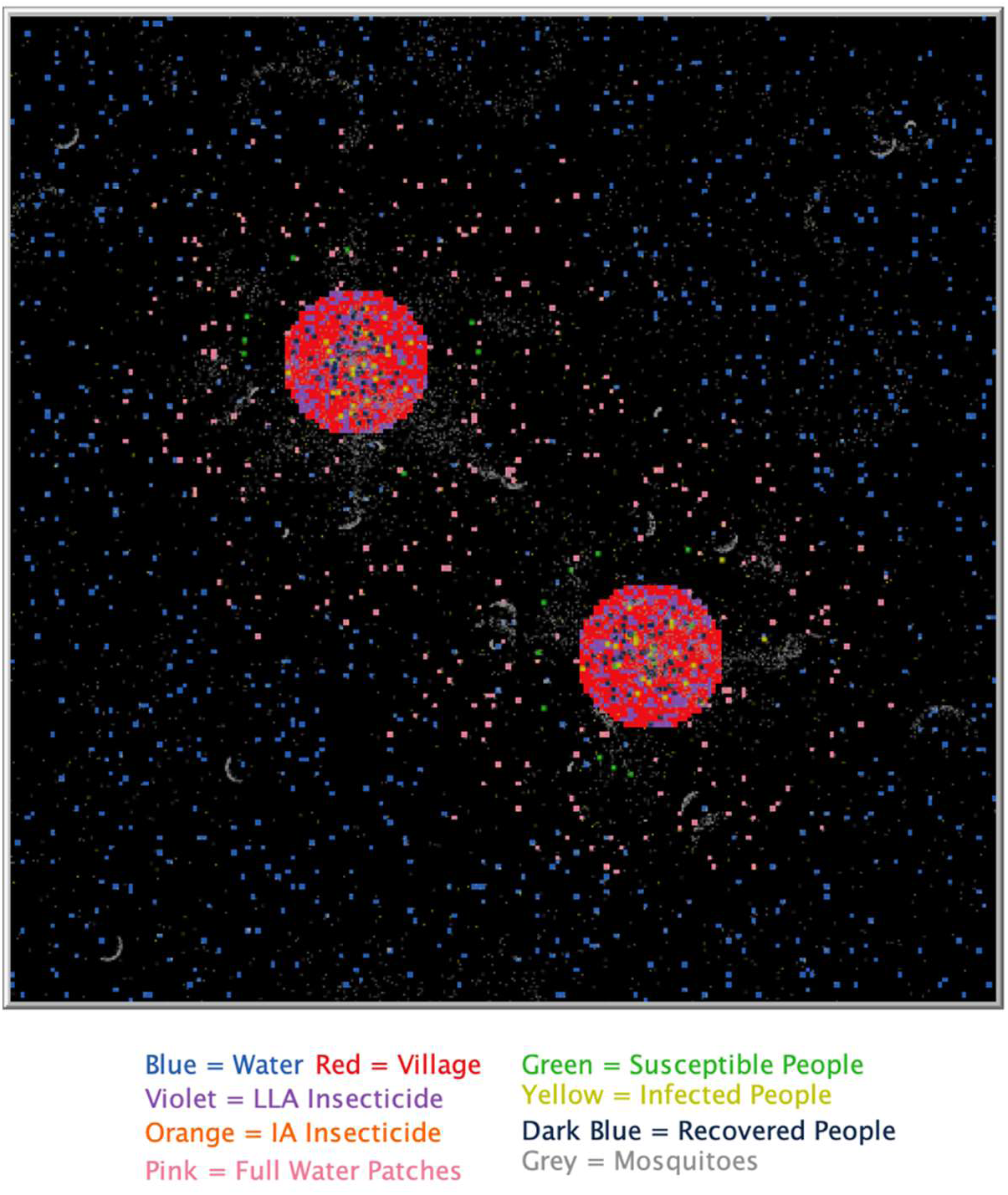
NetLogo representation of the agent-space. Screenshot showing two villages (red), patches with insecticide treatment, aquatic breeding sites, humans and mosquitoes. The black patches are empty, not including any insecticide, humans, breeding sites or mosquitoes.

### Resistance

The resistance to the LLA insecticide in the model was governed by two alleles giving three possible genotypes, susceptible (SS), partially resistant (SR), and resistant (RR) [16, 17]. We assumed that a single mutation was responsible for causing resistance for both of the insecticides (see Table 1). Since male mosquitoes were not explicitly modeled as agents in the program, the contribution of the male alleles to the spread of resistance was computed probabilistically based on the proportion of each genotype in the simulation. The resistant allele was introduced in the burn-in (a number of days at the beginning of a simulation run to let the model stabilize before collecting data – sometimes called warm-up) and maintained at up to 2% of the population to simulate a naturally occurring mutation. The initial percent of SR mosquitoes is a changeable variable in our model but we used 2% for consistency. Since the male mosquitoes were not simulated, the female mosquitoes determine the genotype of the male they have mated with by using the percentage of each genotype in the total population as the probability that they mate with each type of male. Once this had been evaluated, it was recorded in a variable which was passed from the female to any broods she created along with her own genotype. When a brood agent transitions to adult female mosquitoes, each new mosquito’s genotype is assigned using probabilities based on the genotypes of the mother and assumed father.

**Table 1.**
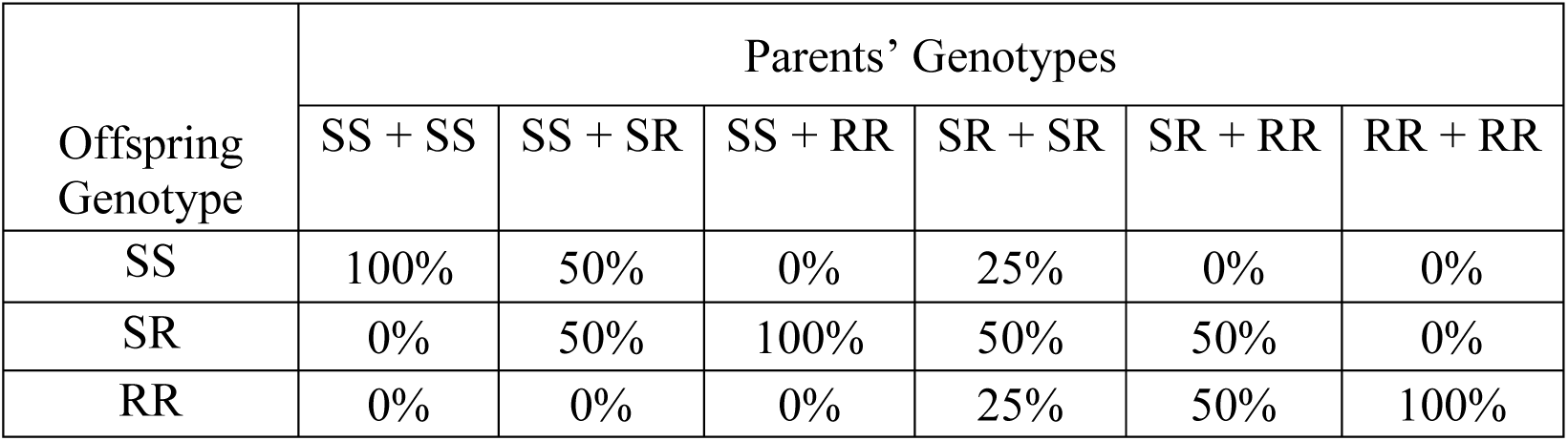
The assumed genetic outcomes for mosquito genotype parings. This table shows the assumed genetic outcomes of the combinations of each genotype. Each mosquito’s variables include a set of two genotypes, one it expresses which was assigned when it entered the system based on its parents’ genotypes, and one which it evaluates based randomly on the density of each genotype in the population. Both of these are combined to assign the genotypes of that mosquitoes’ offspring.

| Offspring Genotype | Parents' Genotypes |  |  |  |  |  |
| --- | --- | --- | --- | --- | --- | --- |
|  | SS + SS | SS + SR | SS + RR | SR + SR | SR + RR | RR + RR |
| SS | 100% | 50% | 0% | 25% | 0% | 0% |
| SR | 0% | 50% | 100% | 50% | 50% | 0% |
| RR | 0% | 0% | 0% | 25% | 50% | 100% |

The model includes an option for modeling a fitness cost of resistance implemented as a reduction in flight speed and an increased larval fatality for the homozygous resistant mosquitoes. The heterozygous resistant mosquitoes are also subject to the cost of resistance in the same ways but at a lower level than the homozygous resistant mosquitoes [17, 48–50]. The cost was assigned three levels: *High Cost, Low Cost*, and *No Cost*. Each of these levels acted on the same variables but gave them more or less extreme values. These values can easily be edited in the code by someone looking to experiment with different types of cost or resistance. These cost values were evaluated to test their effect on the spread of resistance. We used a variable called *Resistance-Dominance* [17] to determine how much the resistant allele was expressed for the heterozygous genotype mosquitoes. The *Resistance-Dominance* raged from 0 to 100 with 100 meaning that the resistant allele caused the heterozygous mosquito to be completely resistant and 0 causing complete susceptibility. For values in-between 0 and 100 the value that was chosen became the chance that the mosquito was affected by the insecticide upon exposure. For example, if the Resistance-Dominance was set to 50, the SR mosquitoes had a 50% chance of being affected by the implemented insecticide per encounter with an insecticide treated patch.

### Model Implementation

The model is implemented as a computer simulation using NetLogo 6.2.0. The NetLogo simulation tool is open source and freely available, running on the Windows, Macintosh and Linux operating systems [32]. A complete executable copy of the EMMIT program can be found in Supporting Information File S1 Program. The ODD (Overview, Design concepts and Details) protocol is employed to provide standardized and detailed documentation of the model [51–54] supporting experimental replication of results and a basis for model adaptation and extension by others. This documentation can be found in Supporting File S2 ODD Documentation. On opening the EMMIT program file in the NetLogo tool, the user can view the Interface_Tab (discussed in the next section), the InfoTab displaying model documentation and usage guidance, and the Code_Tab displaying the actual code with detailed line-by-line comments. As implemented, the program is highly modular, consisting of 39 procedures. Setup and initialization of the program takes place in 14 of those procedures and the remaining 25 procedures contain the code for execution of the simulation. About 40 global variables are used to control a simulation instance, of which about 20 can be set through the user interface using slider, switch and dropdown interface elements. The remaining global variables are set in the code but can be easy edited to take on different values. In the Code_Tab the global variable declarations and procedures (i.e., the functional code, excluding program comments) total approximately only 600 lines. These program statistics suggest that this program is an example of an end-user development, user-extensible agent-based simulation of malaria transmission, but capable of providing insight into complex questions about the disease’s transmission and control. A typical single simulation with an agent space consisting of 40,401 patches, as many as 50,000 mosquito agents and 200 human agents, running for up to 20 years, running on a single core on a consumer-grade computer (laptop or desktop) has a wall-clock run-time of approximately 5-8 hours. Simulation experiments consisting of hundreds of parameter-sweeps were run using the Behavior Space tool in NetLogo specific variable values, including ranges of values, as a parameter sweep. Each individual simulation runs on a separate core, with multiple simulations running in parallel on all available cores of a single computer (for example, 8 parallel runs on an 8-core machine). If needed, the Behavior Space tool also supports running multiple simulations on a cluster of computers. As each simulation is completed, the next one begins until all scheduled runs are completed. The output of each Behavior Space experiment is recorded to a CSV file with the specified output data from each run. A separate data tool can then be used to analyze and visualize the results.

### Interface and Simulations

A screen shot of the EMMIT program’s interface is shown in Fig 3. The interface of the program contains 25 input elements (sliders, switches and drop-down menus) that a user can select to set the associated 19 variable values, monitors for observing mosquito and human population values during run-time, dynamic charts, and a map of the simulated world and the agents on it. For interactive usage, a simulation is started by selecting the desired values in the input features, pushing the *Setup* button to initialize the agents, and then pushing the *Go* button to begin the simulation. We believe this interface is intuitive enough for most researchers to use and understand so that it can be used in many settings.

**Fig 3.**
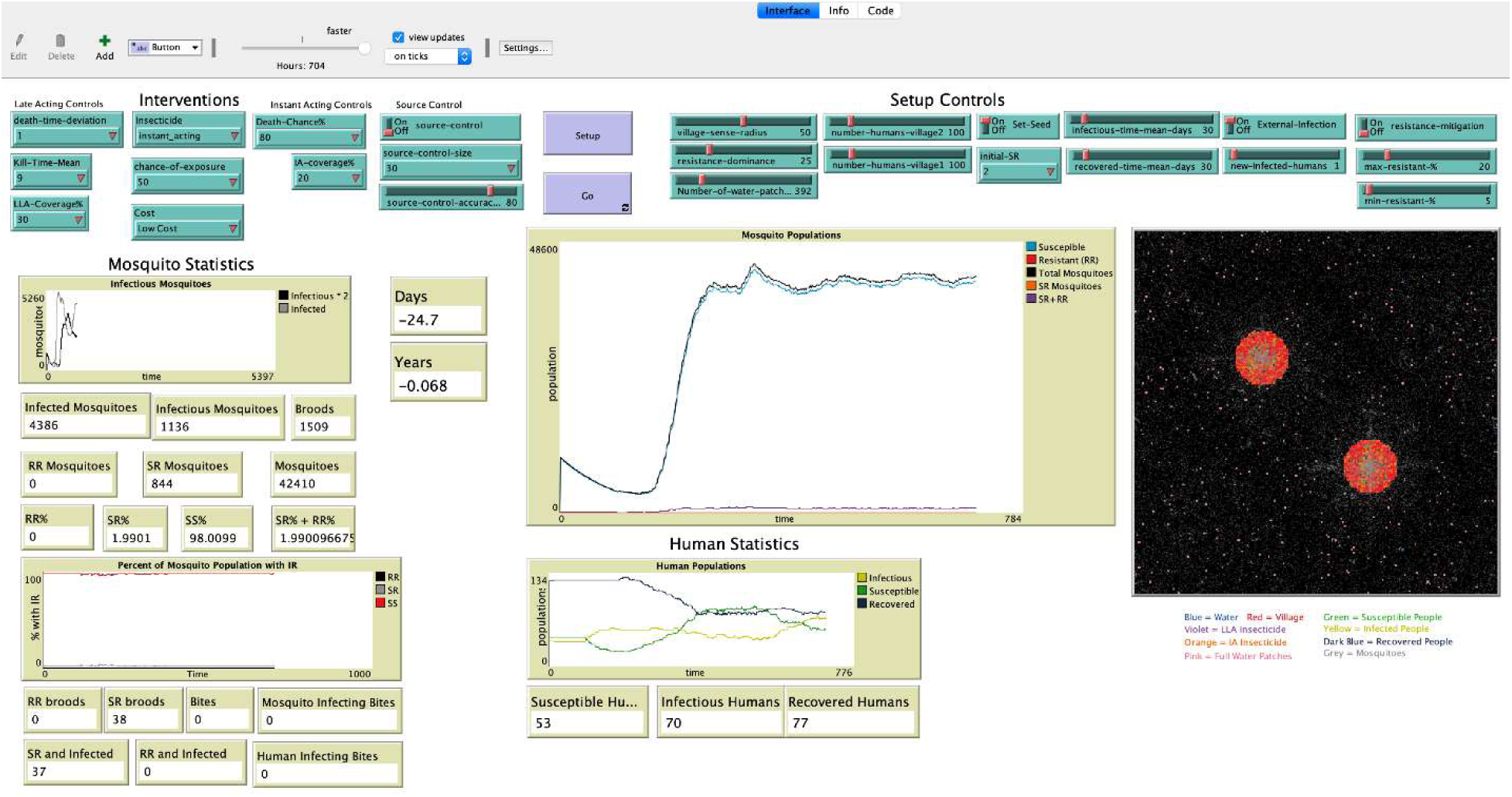
NetLogo user interface. Screen shot of the EMMIT program’s interface displays user-selectable inputs specifying malaria control interventions and simulation setup values, and program outputs including mosquito statistics and human statistics.

When the simulation begins, it has a 1000 timestep burn-in period to initialize the populations and then a one-year control period before the chosen intervention is applied. This control period enables the observation of a simulated baseline, prior to the introduction of a malaria control intervention. When running the simulation using the interactive interface, the simulation runs until the user stops it pressing again on the *Go* button.

Most of the simulations were run with a fixed seed for the random number generators to enable replication of results and comparison of simulations with different parameter settings. This removed variability in the simulation attributable to the starting point in the random sequence generated by the random number generator. Sensitivity analysis on different random seeds indicated that multiple runs are typically needed to properly determine the distributions of possible outcomes. More discussion on this is reported in the results section below.

### Model Parameters

Where applicable, we set simulation parameter values based on values reported in the literature. See Supporting File S3 Table of Model Variables for most of these used in the model. When a range of values are reported (perhaps dependent on temperature, mosquito or parasite species, etc., we typically modeled the uncertainty with random distributions. While most of these values are hard coded into the program, they can be easily edited in the Code_Tab view of NetLogo. As an example, one of these is the EIP which is reported as being between eight and twelve days [34, 43]. This value played an important role in the results and insights provided by our model and can be edited in the model code. As another calibration example, we were able to find WHO data to calibrate the kill rate of instant acting insecticide we simulated [12, 55]. We did test other values as well to allow for natural variation which may occur.

## Results

We ran many simulations and analyzed the output with six different (but overlapping) objectives: 1) calibration, 2) verification, 3) validation, 4) sensitivity analysis, 5) evaluation of the potential of LLA biopesticides for malaria control, and 6) demonstration of the model’s extensibility and use of simple end-user-developed agent-based modeling to investigate important public health research questions. Objectives 1-4 served the purpose of increasing our confidence in the model and objectives 5-6 provided the two contributions of this study. We discuss and provide examples of these below.

### Calibration, Verification, and Validation

Calibration of the model is the process of iteratively setting appropriate simulation parameters to achieve internally consistent and externally plausible outputs. For example, the simulated world is abstracted to be a toroidal grid of 100m^2^ patches of total size of approximately 4 km^2^. Movement of the mosquito in the simulation is specified by patches per time-step (where a time-step is specified to be one hour). This needs to be consistent with actual flight distances over time of the mosquitoes. For example, mosquito flight distances are reported to have typical values of a few hundred meters per day, but may occasionally be long distances [56, 57]. The size of our simulated world and typical flight distances are consistent with these reported values. Likewise, the natural mortality (pre-intervention) of the mosquitoes needs to be modeled, to properly address the research question investigated in this paper, i.e., the potential ability of a late-acting bio-pesticide to delay IR while controlling malaria transmission.

As described in more detail earlier, we assumed an average baseline mosquito mortality of 1% per hour for 12 hours per day, resulting in a daily mortality of slightly less than 12% per day on average. This value is consistent with reported survival rates of a mosquito in the wild [56, 57]. To calibrate this parameter value and its usage in the simulation, we recorded data from a running control simulation (no insecticide interventions) and tabulated the average age distribution of the mosquito population (see Fig 4) following the methods of Hancock, et. al. [11]. A good fit to the expected age distribution is observed demonstrating parameter calibration (also providing support for verification and validation of the simulation).

**Fig 4.**
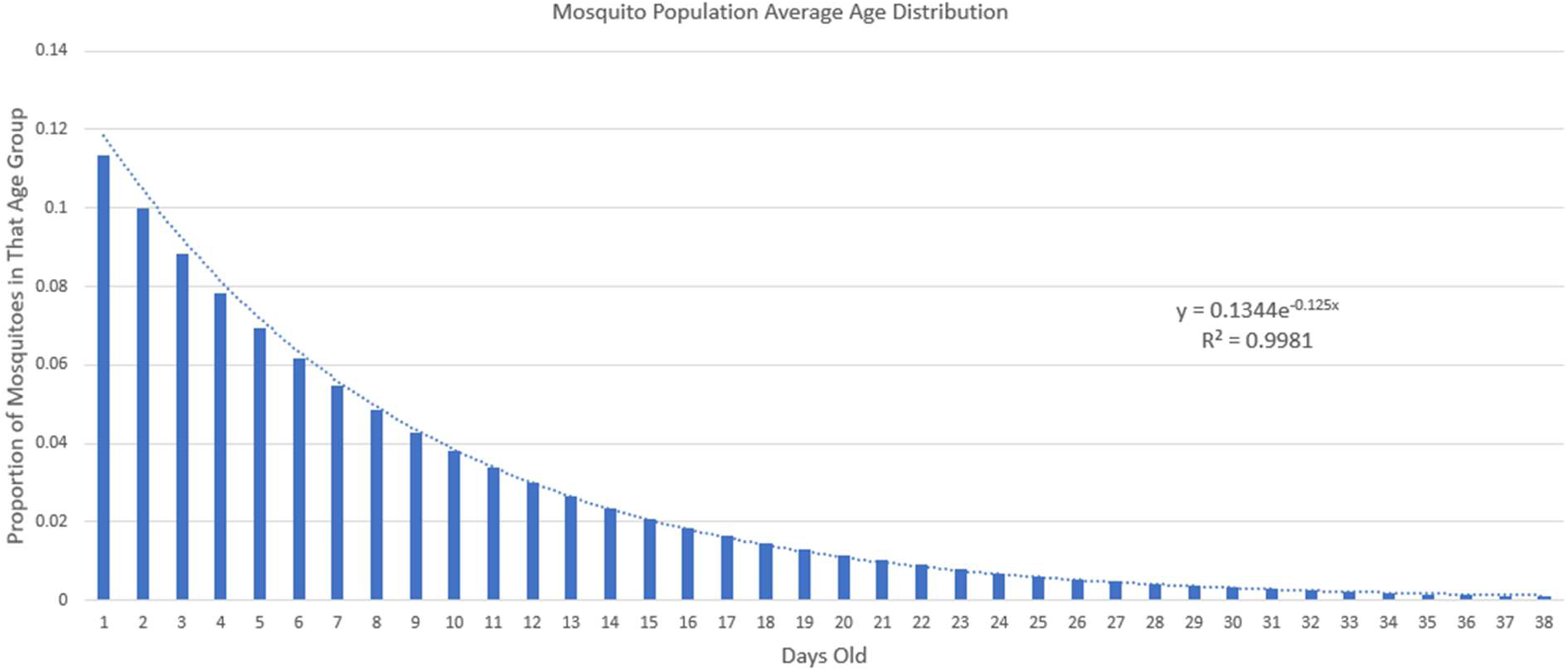
Mosquito Age Distribution. This graph shows the percent of the population in each daily age group. The data is an average of the population age structure over many days in a control test. This data confirms our intended age structure. In addition to the data, a fitted curve is plotted to show the similarity between the data and an exponential decline.

Verification checks if the simulation runs correctly, as intended by the modelers, primarily checking for coding and programming logic errors. Validation checks for the correctness of the design and specifications as defined by the researchers, primarily checking for reasonable emulation of the underlying phenomenon being investigated. A large literature describes methods for conducting verification and validation of ABMs. See Cooley [58], Xiang [59], and Sargent [60] as examples. To verify and validate the model described in this paper, we used multiple methods, including code walkthroughs, face validity, internal validity, input-output analysis, and sensitivity analysis. Since the NetLogo tool provides multiple types of interface elements (e.g., animations of agent movement, monitors displaying program variable values, graphical plots of time series – see Fig 3), face validity checks, internal validity checks, and input-output analysis was easily conducted. A face validity check on Fig 5 suggests that the underlying SEIRS disease model is properly implemented in the simulation. The number of humans in each state fluctuates stochastically in equilibrium around average levels. In Fig 6 we see support for the case that seasonality, insecticide treatments and IR are properly modeled and implemented in the model. Mosquito populations for three conditions are compared: baseline (no insecticides), IA insecticide introduced after 12 months, and LLA insecticide introduced after 12 months. Each of the three insecticide treatments was replicated 10 times with a different random starting seed. The thick lines in the plots are the average of all output sequences for each treatment. The IA insecticide is more effective initially, but resistance appears after a few years, while the LLA insecticide, while slightly less deadly, avoids resistance for the duration of the simulation run. This displays support for the verification and validation of the model. In these simulations we observe a minor dependence on the starting random generator seed, primary with the IA insecticide.

**Fig 5.**
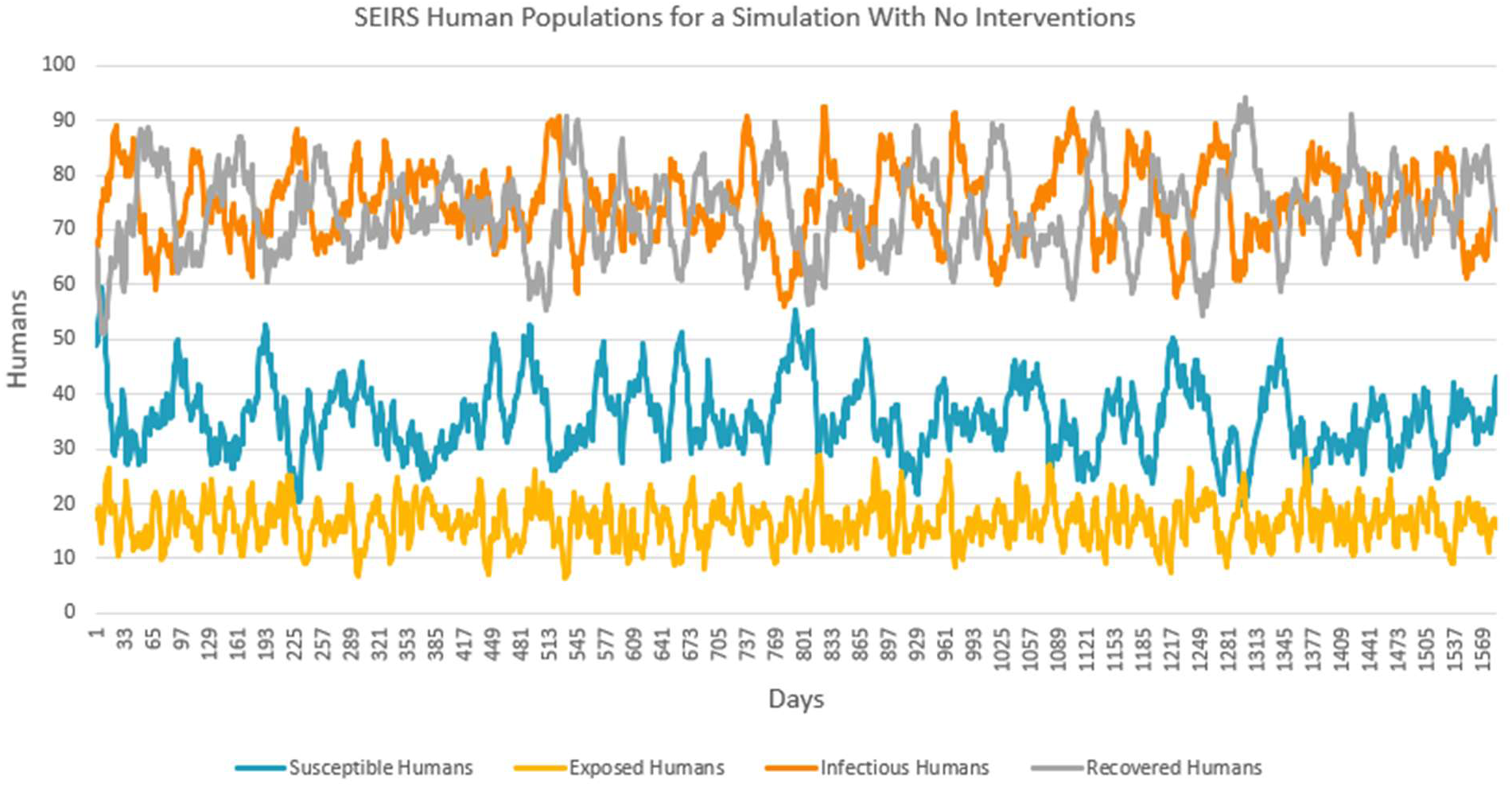
Validation of assumed SEIRS model implemented in the NetLogo simulation. People transition over 10 years through Susceptible, Infected, Recovered and Susceptible disease stages associated with an endemic disease such as malaria. See S4-Fig.1 in Supporting Information “S4 Figures Output” for parameter settings for this result.

**Fig 6.**
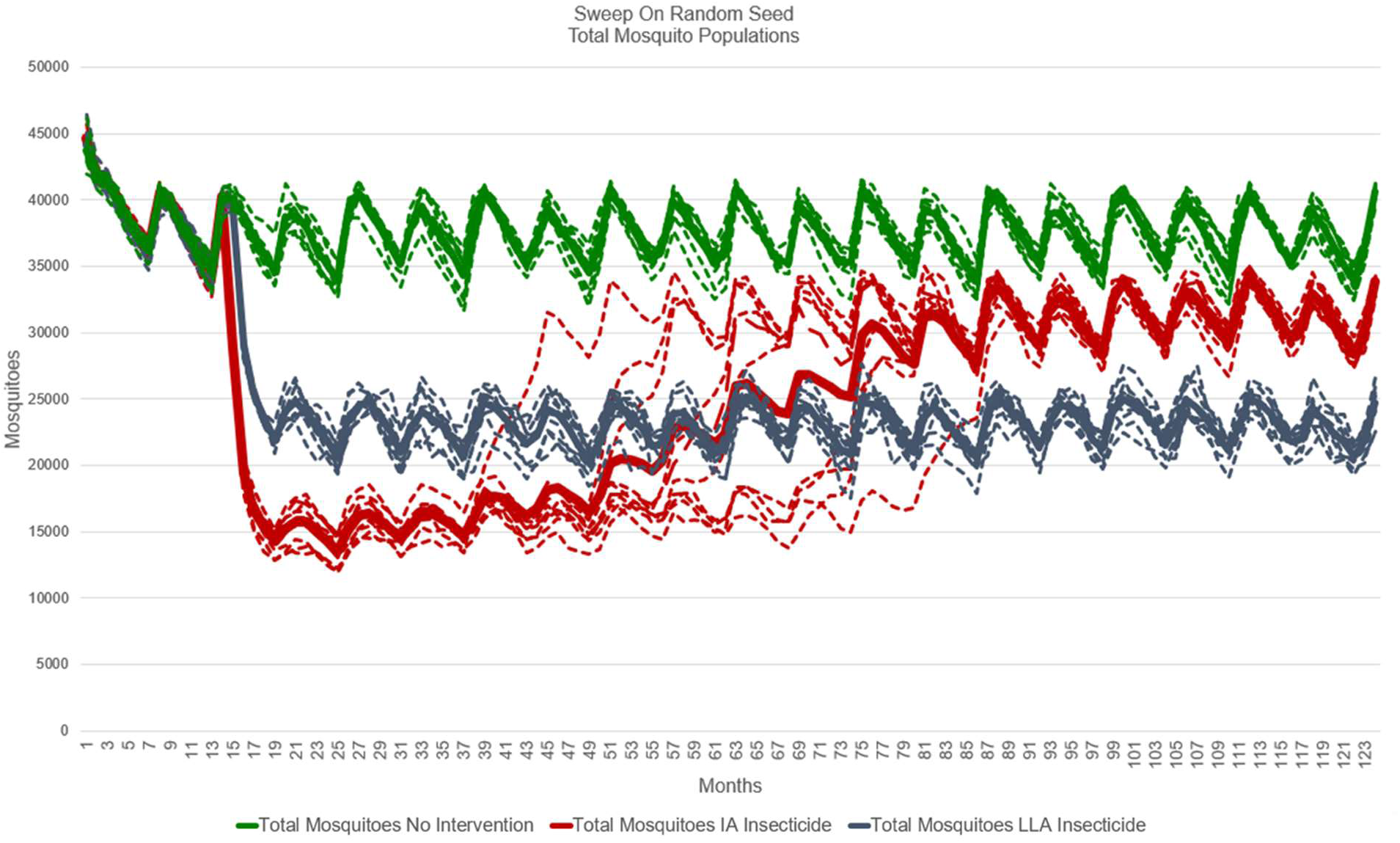
Validity check of simulations of three insecticide treatments: no insecticide, IA insecticide, and LLA insecticide. No intervention (no insecticide baseline) displays mosquito population varying with seasonality and stochasticity of mosquito deaths, births, and gonotrophic cycles. IA insecticide treatment displays initial large reduction in mosquito population, followed by evolving IR with mosquito population rising close to the baseline. The LLA insecticide treatment displays sustained reduction in mosquito population by avoiding evolving IR over an 8-year period. Each of the three insecticide treatments was replicated 10 times with a different random starting seed. Minor dependance on the starting seed is seen in the IA insecticide intervention between months 40 through 85.

One aspect of our model which required extensive calibration, verification, and validation was the genetics involved in the mosquito IR. Originally, we employed a simple method of transferring the resistant gene from a resistant female to all offspring. We refined this process to be more accurate through verification and validation to better represent the natural stochasticity of genetics. We implemented the two-allele genotype and a set of probabilities for how those genes were expressed described above. We also used the idea of resistance dominance from South and Hastings [17] to model multiple resistance levels for the heterozygous genotype. With this method, we were able to provide better analysis on the development of IR.

### Sensitivity Analysis

Sensitivity analysis of the simulation outputs was conducted for many of the important model parameters, including 1) sensitivity of the initial random number generator seed, 2) the delay in the timing of the mosquito’s death after exposure to LLA insecticide (*kill-time-mean*), 3) the variability in the timing of mosquitoes’ death after exposure to LLA insecticide (*death-time-deviation*), 4) the fitness cost of the resistance allele, 5) the resistance dominance associated with the SR genotype, 6) the effect of the area of source control (reduction of aquatic breeding sites) around the villages, 7) the chance of exposure to the LLA insecticide (probability range), 8) LLA insecticide coverage (percent of village patches with insecticide), 9) the size of the source reduction area, and 10) different resistance mitigation policies. A more extensive documentation and presentation of this analysis can be found in Supporting Information S4 Figures.

The results of a sensitivity analysis for two items are displayed in Fig 7: 1) variability of simulation output caused by the initial value of the random number generator seed, and 2) a sweep over multiple values of the LLA insecticide standard deviation parameter.

**Fig 7.**
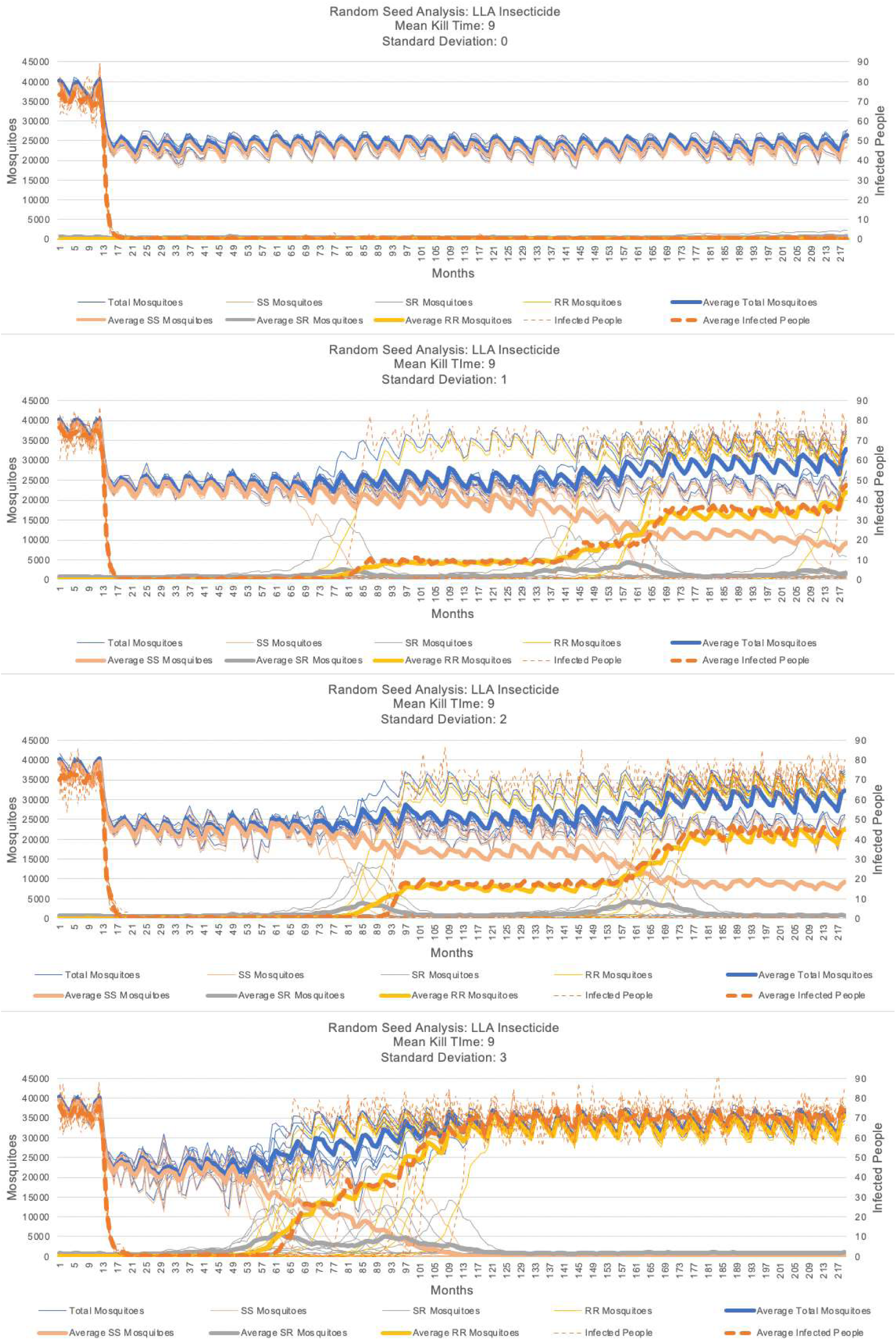
Sensitivity analysis on the Random Number Generator seed and the standard deviation for the kill-time-delay for the LLA insecticide. This figure displays the results of eight replications of the EMMIT program with the same settings but different random seeds. Individual run results are shown in thin lines and the averages of each output parameter are thick lines. See Supporting Information S4 Figures for the simulation parameters.

#### Sensitivity to Random Generator Seed

As discussed in the Model section above, the simulation expresses the randomness of many events in the transmission of malaria (e.g., probabilities of exposure to an insecticide, the probability that a mosquito blood meal results in an infection, the distribution of mortality of an LLA insecticide, etc.) with calls to a random number generator. As with most such random number generators, an initial seed is required to start a sequence of values, which then are used to compute probabilities in the simulation. Simulation output will typically vary based on the value of the initial seed and this needs to be accounted for in the interpretation of the results. See Martinez-Moyano and Macal [61]. Fig 7 presents the results of 8 runs of the EMMIT program for an LLA insecticide scenario, while all model parameters were held to the same values with only the starting seed changed. The sensitivity of simulation outputs based on the starting seed can be observed in the wide range of output time series. The thin graph lines present the simulation output for individual runs. The thick lines report the mean of the output time series over all the 8 runs of the simulation. We thus conclude that a valid interpretation of the simulation results will require such sensitivity analysis to avoid the possible situation of any one simulation run being an outlier and approximately 8 replications provide a good sample size for estimating the mean and variability of the simulation’s output sequences.

#### Sensitivity of Standard Deviation of LLA Insecticide Time to Death

In Fig 7, a sensitivity analysis on the standard deviation parameter is displayed for a simulation of the LLA insecticide, with mean time to death of 9-days (*kill-time-mean*), and an iteration of zero through three days for the standard deviation (*death-time-deviation*). For each standard deviation setting in the model, eight replications were run. Outputs are total mosquito population, numbers of each mosquito genotype (SS, SR, RR), and the number of infectious humans. As the standard deviation increases, it can be observed that the number of infectious humans increases because of the survival of older infectious mosquitoes. Also, as the standard deviation increases, we more frequently see an earlier appearance of IR. See Supporting Information S4 Figures for other sensitivity analyses.

### Results for Objective 1: Evaluating the Potential of LLA Insecticides

LLA insecticides have been proposed as a potential method to delay the emergence of IR and to provide another tool for malaria control managers to reduce transmission by reducing the mosquito population [33–39, 41]. It is argued that the delayed death of the mosquitoes after first exposure to the LLA insecticide can reduce the selective evolutionary pressures that result in IR by permitting the SS or SR genotypes time to return the susceptible allele to the population.

The first objective of the research reported in this paper is to provide (by way of simulation) insights into the required performance properties of such LLA insecticides in order to achieve the goal of delaying the emergence of IR and to identify other variables that affect the insecticide’s performance. Towards this objective, we have described 1) an assumed model of how malaria is transmitted between mosquito vectors and humans, 2) the emerging challenge of IR for malaria control, 3) the design and implementation of the EMMIT program, 4) the calibration, verification and validation of that program, and 5) and the use that program to explore, discover, and evaluate the effect of operational characteristics and assumptions about an LLA insecticide. See Supporting Information S4 Figures for an extensive report on this last item.

While there are many variables in the model, a few critical variables determine whether the LLA insecticide might be effective. An ideal set of values for these critical variables results in the LLA insecticide killing as many infected mosquitoes as possible prior to those mosquitoes becoming infectious, but with as long a time-to-death delay as possible to slow IR emergence. Analysis of simulation results (see Supporting File S4 Figures) suggests that several operational characteristics of the LLA insecticide, its application, and the nature of the mosquito’s resistance strongly influence the LLA insecticide’s potential success. These six critical variables are: 1) the delay between exposure and death of the mosquito (i.e., the Kill-Time-Mean), 2) the distribution of mosquito deaths around the mean delay-to-death (i.e., the standard deviation), 3) the LLA insecticide coverage, 4) the fitness-cost of resistance, 5) the chance of exposure, and 6) resistance dominance.

The best values of these critical variables that the EMMIT program predicts for a LLA insecticide are displayed in Table 2. The associated values at which LLA insecticide perform with either high or low success are listed in separate columns. These variables are synergistic with one another so there is an overlap in the categorization of some values. Finally, we note that these results are dependent on our assumptions of mosquito properties, LLA insecticide properties, malaria transmission properties, and the course of the disease in the human population. The assumptions in the current model will need to be calibrated before evaluation of a specific LLA insecticide, a specific vector species, and a specific environmental and public health setting.

**Table 2.** LLA Insecticide Critical Variables. Given all the assumptions built into the EMMIT program, six critical variables and their values associated with successful and less successful performance of the LLA insecticide are listed in this table. Successful performance here implies achieving the two simultaneous goals of a LLA insecticide: 1) killing mosquitoes before they can transmit malaria, and 2) doing it with as long as possible a delay so as to slow the emergence of IR. Some of the critical variables represent characteristics of IR in the mosquito population (cost-of-resistance and resistance-dominance) and the others are associated with the LLA insecticide’s application (LLA-Coverage-%) and its performance (kill-time-mean, death-time-deviation, and chance-of-exposure). Researchers wishing to use such a model to inform the development of a candidate LLA insecticide will need to calibrate all the assumptions of the model especially for these six critical variables.

| Critical Variables | Values With High Success | Values With Low Success |
| --- | --- | --- |
| Kill-Time-Mean | 7,8,9 | 10,11 |
| death-time-deviation | 0, 1, 2 | 2, 3 |
| LLA-Coverage% | 20, 25, 30, 40 | 10, 20, 50 |
| Cost | High Cost, Low Cost, No Cost | Low Cost, No Cost |
| chance-of-exposure | 50, 75, 100 | 25 |
| resistance-dominance | 0, 10, 20, 25, 30, 40, 50, 60 | 50, 60, 75, 80, 90, 100 |

In Fig 7 we display a sensitivity analysis of one of the most important variables, the distribution of mosquito deaths around the mean time from exposure to death (i.e., the *death-time-deviation*). A zero value for the death-time-deviation means the LLA insecticide kills all the mosquitoes exposed to it at precisely the specified time, ideally just before the mosquito becomes infectious and begins to transmit malaria. Values for the *death-time-deviation* greater than zero imply that the LLA insecticide kills the mosquitoes either earlier or later than the ideal mean kill time delay, diminishing its effectiveness both at reducing transmission and at slowing down the emergence of IR.

These results are primarily meaningful for a malaria transmission scenario close to the one we simulated. Researchers wishing to use such a model to inform the development of a candidate LLA insecticide will need to calibrate all the assumptions of the model especially for these six critical variables and confirm that our simplifying assumptions in the simulation are valid for their application.

### Results for Objective 2: Demonstration of an Approach for End-user Development of Computer Simulation in Support Public Health Research

The EMMIT program and investigation described in this paper demonstrate the feasibility of end-user/non-technical development of computer models and the ability to provide useful insights into public health research questions. Earlier in this paper we identified multiple modeling and simulation challenges, including: 1) requirement of either strong mathematical or computer programming skills, 2) the typical complexity of modeling and simulation, 3) long development time, 4) lack of ease of use, 5) simulation adaptation and extensibility, 6) transparency of assumptions and limitations, and 7) often the need for high performance computing resources. In Table 3 we summarize how the approach we describe in this paper addresses these challenges. For this study, we wished to demonstrate the relative ease of developing a model primarily focused on a specific public health topic. While the model presented here can be adapted and extended, as the research topic changes sufficiently from the motivating topic used to design the original model, modifications will become more difficult and a new design may be required.

**Table 3.** Modeling Challenges and How Approaches Address Those Challenges.

| **Modeling Challenges** | **How Approach Presented Addresses the Challenges** |
| --- | --- |
| Need for Strong Mathematical or Computer Programming Skills | Agent-based simulation and modeling using NetLogo is employed. Basic mathematical and programing skills are needed. The initial version of the model was developed by Author 1, a novice programmer (at the time) with only one semester of programming training in MATLAB. Author 2 helped define the public health problem to be modeled and assisted with the design of the conceptual model, but only contributed to the simulation with verification and validation of the results (primarily face-validity). |
| Complexity | The model contains only 600 lines of functional code (other lines in program are comments or demonstrations of extensibility). NetLogo (and other such simulation building tools) are widely available, free and open source in many cases, and can be learned and used in a short time. |
| Long Development Time | The initial version was developed in under 100 hours of effort by Author 1. That development time included both learning how to use the NetLogo tool and actual computer coding. (That said, much more additional time was eventually needed for extensions, verification, validation, and documentation, specifically for this paper). |
| Ease of Use & Transparency of Assumptions and Limitations | The graphical interface permits interactive simulation runs providing understanding of the behavior of the program and insights into its assumptions and limitations. The program also includes 1) tabs to view the model's documentation, 2) instructions on how to use the program, and 3) a full listing of the model's code and comments. |
| Extensibility and Flexibility | In addition to the model of LLA insecticides for addressing the malaria transmission control IR problem, two other interventions were added as a demonstration of the potential ease of extending the simulation. This required under 10 lines of code for the source control intervention and about 35 lines of code for the IR mitigation policy. |
| Need for High Performance Computing Resources | NetLogo and other such simulation development tools run on consumer-grade desktop/laptop computers. NetLogo programs can be run in "batch mode" using the Behavior Space tool on desktop and laptop computers utilizing all available cores, and if needed, can run in "headless mode" (ie, no user interface) from the command line on computer clusters. Modern consumer-grade computers have increased performance, often equivalent to high performance computing of a decade ago. |

## Discussion

In this study we designed, developed, verified, and validated a basic model of malaria transmission using NetLogo as a demonstration of how an easy-to-use ABM tool can be used to investigate specific public health questions. To demonstrate that meaningful research questions can be investigated with this approach, we presented the problem of IR and a proposed IA insecticide alternative – LLA insecticides – to evaluate how effective LLA insecticides could be at both controlling malaria and avoiding IR. We designed the EMMIT program such that its behavior and assumptions were transparent so that it can be adapted, used and understood by researchers from many disciplines, including those who regulate the production of new insecticides, researchers who engineer new insecticides, and public managers who design malaria control campaigns impacted by IR. We conducted comparison runs on a representative IA insecticide, such as those currently approved for use, and a hypothetical LLA insecticide. Two especially important parameters we evaluated were the *Kill-Time-Mean* (the mean delay-before-death after exposure to the LLA insecticide) and the standard deviation of the delay. Our analysis suggests that the more closely the timing of the LLA insecticide’s delay-to-death coincides with the end of the EIP, the better it was at preventing malaria transmission and avoiding insecticide resistance in the mosquito population. We also concluded that the hypothetical LLA insecticide we simulated performed better than the instant acting insecticide in most situations.

We made many simplifying assumptions in the model. We assumed that the mosquitoes have a constant baseline hourly mortality regardless of age or position on the map. While this is a very difficult parameter to calibrate, it could be improved by implementing some of the findings from [11]. Our simplification may result in a slight change in the selective pressure for IR and in the malaria transmission as Hancock [11] reported that mosquitoes may have a longer average lifespan than we modeled.

The two-allele mosquito genotype was also a simplification in our model as IR is often the result of many concurrent and interacting genotypic and phenotypic changes. An improvement to this simplification would be to increase the complexity of the modeled mosquito genotype and the fitness benefits and costs associated with that increase. This simplification may result in a change in the prevalence of IR, especially in the instant acting insecticide.

Another simplified component of our model was the mosquito movement. The mosquitoes move randomly until they enter the region of the map where they can sense the village (controlled by the *village-sense-radius* variable). This variable was not based on reported data but rather calibrated using face validity to find a suitable value. Future improvement might be to calibrate this variable more accurately to a specific region and mosquito population, and incorporate reported movement patterns for that geographic location.

Other simplifications that could be improved include: 1) male mosquitoes and their genotypes are abstracted in the simulation, 2) no birth or deaths of male mosquitoes are modeled, 3) no cost of resistance is included for the male mosquitoes, and 4) the cost of female mosquito IR is modeled by way of reduced travel distances per time step. These simplifications would need to be improved for more accurate representation of resistance cost in both the males and females or for studies of other interventions such as a sterile insect technique.

The humans in our model also have simple characteristics and behaviors. Most only move randomly within the village. A possible improvement to this would be to add human attributes such as age and gender specific differences for time spent in areas with a higher likelihood of infection, differences in bed-net usage, etc. We modeled a small number of humans who traveled out of the village. These people represented village members who may leave the village area for several days to perform agricultural, forestry or mining work away from their village. These people moved constantly in and out of the village in straight lines regardless of whether they were infected. This routine may not be representative of some populations and can be easily edited or disabled in the code of the model. We did model a feature where human agents were infected randomly, without mosquito interaction. This simulates humans receiving infections outside of the modeled region and bringing them back to their community, i.e., importation of external malaria cases [62]. We also did not model any malarial drug administration, prevention, and treatment. We did not model births, deaths, or age, sex, and pregnancy dependent malaria susceptibility in our model. This simplification may have effects on the malaria transmission rate in our simulation, depending on the specific questions being investigated.

We modeled the insecticides in our model so that they had a constant coverage rate and a constant performance level. Application details, reapplication, effectiveness decay, or delivery mechanism of the insecticides, (such as insecticide treated bed nets or indoor residual spraying) were not explicitly modeled. Such details should be relatively easy to add. In practice these performance parameters are difficult to achieve but may be closely approximated with frequent re-applications. For locations where reapplication is not an accessible option, an improvement to the model would be to have the chance of exposure to the insecticide decay over time to show the deterioration in the active amount of insecticide remaining.

Some remaining factors to explore which we did not investigate are: the length of time that the humans spend in the infected and recovered stages, the introduction of bed nets or other interventions, the combination of the insecticides with source control, the effect of changing temperatures, and varying the number of people in the simulation.

While NetLogo is the modeling tool used for this study, many other such tools are available [27, 29–31]. NetLogo is a relatively mature tool, with the benefit of many useful features supporting the programmer: an extensive tutorial and manual, tool studies, and many example models. Because it was originally designed for educational applications, inspired by the Logo project, and used the Lisp programming language in precursor implementations, some of NetLogo’s programming syntax contain curious legacy artifacts, such as agents are called turtles in the programming language. These legacy artifacts will seem strange to programmers trained in more contemporary programming languages, but should not be a problem for novice programmers. Given this minor issue, the tool is implemented in Java and Scala providing portability to most modern computer operating systems: Windows, Mac OS, and Linux.

## Conclusions

This paper presents the results of a study with two related objectives: 1) demonstrate how simple but useful models of public health questions can be rapidly build by skilled public health researchers, including those with limited mathematical modeling or programming skills (i.e., end-user-development), and 2) demonstrate this by simulating the performance of proposed LLA insecticides as an approach to address the major problem of IR for malaria control. A ready-to-run copy of the EMMIT program including graphical interface, usage information, and editable code is included. Supporting information also includes documentation using the ODD format for ABMs and an extensive report of calibration, verification, and validation simulation runs.

For this project, the agent-based modeling approach, employing the NetLogo tool, was used. The model presented in this paper was coded exclusively by one novice programmer (the first author), with the second author contributing by framing of the public health research problem and assisting with verification and validation. The modeling and simulation approach presented in this paper might be ideally suited to smaller or urgent public health investigations where resources are limited, but the potential insight provided by the simulation is needed quickly. For a study such as this one, the public health expert alone might be able to design and program the model in support of their investigation. For larger scale investigations where more time and resources are available, a team including both the public health experts and skilled mathematical modelers or programmers might be more appropriate. We suggest that the EMMIT program and investigation described in this paper demonstrate the feasibility of end-user/non-technical development of computer simulations and the ability to provide useful insights into public health research questions.

## Supporting information

S1 - Program

S2 - ODD Documentation

S3 - Table Model Variables

S4 - Supplemental Figures Output Plots

## Data Availability

All data related to this project can be found in the supporting files and at https://www.comses.net/codebases/2dbac15b-a67d-4ad0-9220-6cc12d2dc963/releases/1.0.0/

## Acknowledgments

We wish to acknowledge unpublished dissertation results based on the agent-based modeling of the LLA insecticide by Ying Zhou that inspired part of this study. We thank the guidance and support for this project from the Perkins Lab in the Department of Biological Sciences at the University of Notre Dame, specifically Alex Perkins and John Huber.

## Supporting Information

S1 Program

The EMMIT program, requires download of NetLogo from: https://ccl.northwestern.edu/netlogo/download.shtml

See system requirements at: https://ccl.northwestern.edu/netlogo/requirements.html (NLOGO/ZIP)

S2 ODD Documentation

The EMMIT program presented in this paper is documented below using the standard ODD format (Grimm2006, Grimm2010, Grimm2017, Grimm2020).

(PDF)

S3 Table Model Variables

Model variables (input and output), definitions, values, and data types.

(PDF)

S4 Supplemental Figures Output Plots

Supplementary simulation results with “behavior space” configuration (used to generate multiple batch simulation runs). Plots generated from CSV output files using Excel.

(PDF)

