## Supplementary material for "Case Study: End User Development of an Agent-based Model of Malaria Transmission to Support the Design of Late-Life-Acting Insecticides for the Control of Malaria Transmission and Delay of the Evolution of Insecticide Resistance": S1 - Program: The EMMIT Program.nlogo

````
;;;;;;;;;;;;;;;;;;;;;;;;;;;;;;;;;;;;;;;;;;;;;;;;;;;;;;;;;;;;;;;;;;;;;;;;;;;;;;;;;;;;;;;;;;;;;;;;;;;;;;;;;;;;;;;;;;;;;;;;;;;;;;;;;;;;;;;;;;;;;;;;;;;;;;;;;;;;;;;;;;;;;;;;;;;;;;;;;;;;;;;;
;;;;;;;;;;;;;;;;;;;;;;;;;;;;;;;;;;;;;;;;;;;;;;;;;;;;;;;;;;;;;;;;;;;;;;;;;;;;;;;;;;;;;;;;;;;;;;;;;;;;;;;;;;;;;;;;;;;;;;;;;;;;;;;;;;;;;;;;;;;;;;;;;;;;;;;;;;;;;;;;;;;;;;;;;;;;;;;;;;;;;;;;
;;;;;Jacob Heintzelman;;;;;;;;;;;;;;;;;;;;;;;;;;;;;;;;;;;;;;;;;;;;;;;;;;;;;;;;;;;;;;;;;;;;;;;;;;;;;;;;;;;;;;;;;;;;;;;;;;;;;;;;;;;;;;;;;;;;;;;;;;;;;;;;;;;;;;;;;;;;;;;;;;;;;;;;;;;;;;;;;;
;;;;;The EMMIT (Extendable Model for Malaria Intervention Testing) Program;;;;;;;;;;;;;;;;;;;;;;;;;;;;;;;;;;;;;;;;;;;;;;;;;;;;;;;;;;;;;;;;;;;;;;;;;;;;;;;;;;;;;;;;;;;;;;;;;;;;;;;;;;;;;;
;;;;;This program simulates the interactions between humanss and mosquitoes which spread malaria and a number of interventions for its control;;;;;;;;;;;;;;;;;;;;;;;;;;;;;;;;;;;;;;;;;;
;;;;;The possible interventions it simulates are a typical Instant Acting insecticide, a novel Late Life Acting insecticide, a larval source control;;;;;;;;;;;;;;;;;;;;;;;;;;;;;;;;;;;;
;;;;;program, and an insecticide management policy to reduce the spread of insecticide resistance;;;;;;;;;;;;;;;;;;;;;;;;;;;;;;;;;;;;;;;;;;;;;;;;;;;;;;;;;;;;;;;;;;;;;;;;;;;;;;;;;;;;;;;
;;;;;Input: use the setup controls and the Interventions choices on the interface to input the parameters;;;;;;;;;;;;;;;;;;;;;;;;;;;;;;;;;;;;;;;;;;;;;;;;;;;;;;;;;;;;;;;;;;;;;;;;;;;;;;;
;;;;;Output: view the plots and monitors on the running simulation to see how the model predicts each intervention will behave;;;;;;;;;;;;;;;;;;;;;;;;;;;;;;;;;;;;;;;;;;;;;;;;;;;;;;;;;;
;;;;;;;;;;;; You can also use the behaviorspace feature for multiple runs. The behaviorspace outputs a CSV file;;;;;;;;;;;;;;;;;;;;;;;;;;;;;;;;;;;;;;;;;;;;;;;;;;;;;;;;;;;;;;;;;;;;;;;;;
;;;;;;;;;;;;;;;;;;;;;;;;;;;;;;;;;;;;;;;;;;;;;;;;;;;;;;;;;;;;;;;;;;;;;;;;;;;;;;;;;;;;;;;;;;;;;;;;;;;;;;;;;;;;;;;;;;;;;;;;;;;;;;;;;;;;;;;;;;;;;;;;;;;;;;;;;;;;;;;;;;;;;;;;;;;;;;;;;;;;;;;;
;;;;;;;;;;;;;;;;;;;;;;;;;;;;;;;;;;;;;;;;;;;;;;;;;;;;;;;;;;;;;;;;;;;;;;;;;;;;;;;;;;;;;;;;;;;;;;;;;;;;;;;;;;;;;;;;;;;;;;;;;;;;;;;;;;;;;;;;;;;;;;;;;;;;;;;;;;;;;;;;;;;;;;;;;;;;;;;;;;;;;;;;
;;;;;Open Source Software:  The MIT License;;;;;;;;;;;;;;;;;;;;;;;;;;;;;;;;;;;;;;;;;;;;;;;;;;;;;;;;;;;;;;;;;;;;;;;;;;;;;;;;;;;;;;;;;;;;;;;;;;;;;;;;;;;;;;;;;;;;;;;;;;;;;;;;;;;;;;;;;;;;;
;;;;;Permission is hereby granted, free of charge, to any person obtaining a copy of this software and associated documentation files (the "Software"), to deal in the Software;;;;;;;;;
;;;;;without restriction, including without limitation the rights to use, copy, modify, merge, publish, distribute, sublicense, and/or sell copies of the Software, and to permit;;;;;;;
;;;;;persons to whom the Software is furnished to do so, subject to the following conditions: ;;;;;;;;;;;;;;;;;;;;;;;;;;;;;;;;;;;;;;;;;;;;;;;;;;;;;;;;;;;;;;;;;;;;;;;;;;;;;;;;;;;;;;;;;;
;;;;;                                                                                                                                        ;;;;;;;;;;;;;;;;;;;;;;;;;;;;;;;;;;;;;;;;;;;
;;;;;The above copyright notice and this permission notice shall be included in all copies or substantial portions of the Software.;;;;;;;;;;;;;;;;;;;;;;;;;;;;;;;;;;;;;;;;;;;;;;;;;;;;;
;;;;;                                                                                                                                        ;;;;;;;;;;;;;;;;;;;;;;;;;;;;;;;;;;;;;;;;;;;
;;;;;THE SOFTWARE IS PROVIDED "AS IS", WITHOUT WARRANTY OF ANY KIND, EXPRESS OR IMPLIED, INCLUDING BUT NOT LIMITED TO THE WARRANTIES OF MERCHANTABILITY, ;;;;;;;;;;;;;;;;;;;;;;;;;;;;;;;
;;;;;FITNESS FOR A PARTICULAR PURPOSE AND NONINFRINGEMENT. IN NO EVENT SHALL THE AUTHORS OR COPYRIGHT HOLDERS BE LIABLE FOR ANY CLAIM, DAMAGES OR OTHER LIABILITY, ;;;;;;;;;;;;;;;;;;;;;
;;;;;WHETHER IN AN ACTION OF CONTRACT, TORT OR OTHERWISE, ARISING FROM, OUT OF OR IN CONNECTION WITH THE SOFTWARE OR THE USE OR OTHER DEALINGS IN THE SOFTWARE.;;;;;;;;;;;;;;;;;;;;;;;;;
;;;;;;;;;;;;;;;;;;;;;;;;;;;;;;;;;;;;;;;;;;;;;;;;;;;;;;;;;;;;;;;;;;;;;;;;;;;;;;;;;;;;;;;;;;;;;;;;;;;;;;;;;;;;;;;;;;;;;;;;;;;;;;;;;;;;;;;;;;;;;;;;;;;;;;;;;;;;;;;;;;;;;;;;;;;;;;;;;;;;;;;;
;;;;;;;;;;;;;;;;;;;;;;;;;;;;;;;;;;;;;;;;;;;;;;;;;;;;;;;;;;;;;;;;;;;;;;;;;;;;;;;;;;;;;;;;;;;;;;;;;;;;;;;;;;;;;;;;;;;;;;;;;;;;;;;;;;;;;;;;;;;;;;;;;;;;;;;;;;;;;;;;;;;;;;;;;;;;;;;;;;;;;;;;
;;;;;;;;;;;;;;;;;;;;;;;;;;;;;;;;;;;;;;;;;;;;;;;;;;;;;;;;;;;;;;;;;;;;;;;;;;;;;;;;;;;;;;;;;;;;;;;;;;;;;;;;;;;;;;;;;;;;;;;;;;;;;;;;;;;;;;;;;;;;;;;;;;;;;;;;;;;;;;;;;;;;;;;;;;;;;;;;;;;;;;;;
;;;;;;;;;;;;;;;;;;;;;;;;;;;;;;;;;;;;;;;;;;;;;;;;;;;;;;;;;;;;;;;;;;;;;;;;;;;;;;;;;;;;;;;;;;;;;;;;;;;;;;;;;;;;;;;;;;;;;;;;;;;;;;;;;;;;;;;;;;;;;;;;;;;;;;;;;;;;;;;;;;;;;;;;;;;;;;;;;;;;;;;;


; assigns the variables belonging to the system and the individual agent-sets
globals [ days                           ; Counts the days the simulation has run for
          years                          ; Counts the years the simulation has run for
          RR-death-rate                  ; Defines the death rate for RR mosquitoes
          SR-death-rate                  ; Defines the death rate for SR mosquitoes
          RR-speed                       ; Defines the speed at which RR mosquitoes can travel
          SR-speed                       ; Defines the speed at which SR mosquitoes can travel
          ss                             ; The percent of the population with the SS genotype
          sr                             ; The percent of the population with the SR genotype
          rr                             ; The percent of the population with the RR genotype
          initial-recovered-humans       ; Defines the initial number of recovered humans (used only in the setup commands)
          initial-infectious-humans      ; Defines the initial number of infectious humans (used only in the setup commands)
          bites                          ; Counts the times the mosquitoes bite humans
          bites/hr                       ; Counts all the bites per hour
          infectious-bites               ; Counts the times the mosquitoes bite infectious humans during the simulation
          village1                       ; Defines the village patches of the first village
          village2                       ; Defines the village patches of the second village
          h-infectious-bites             ; Counts the bites on humans from infectious mosquitoes during the simulation
          infecting-bites/hr             ; Counts the bites on infectious humans which infect the mosquito
          mitigation-active              ; Defines the level of resistance mitigation which is active in the simulation
          treated-patches                ; Counts the number of insecticide treated patches at the beginning of the simulation
          count-water-in-area]           ; Counts the initial number of water patches in the source control area
  ;;;;;Globals in the interface;;;;;;;;;;;;;;;;;;;;;;;;;;;;;;;;;;;;;;;;;;;;;;;;;;;;;;;;;;;;;;;;;;;;;;;;;;;;;;;;;;;;;;;
        ;;Insecticide                    ; Defines which insecticide is being used
        ;;death-time-deviation           ; Defines the value of the standard deviation on the kill time of the Late Life Acting Insecticide (LLA)
        ;;LLA-Coverage%                  ; Defines the coverage rate over the village of the LLA
        ;;Death-Chance%                  ; Defines the probability that a mosquito will die after contact with the Instant Acting Insecticide (IA)
        ;;IA-coverage%                   ; Defines the coverage rate of the IA over the village
        ;;chance-of-exposure             ; Defines the chance that a mosquito will encounter the insecticide while on a treated patch for both insecticides
        ;;Cost                           ; Defines the level of fitness cost associated with resistance for both insecticides
        ;;source-control                 ; Boolean: defines if there is source control or not (i.e., source reduction of aquatic breeding sites)
        ;;source-control-size            ; Defines the size of the source control area
        ;;number-humans-village1         ; Defines the initial number of humans in the first village
        ;;number-humans-village2         ; Defines the initial number of humans in the second village
        ;;resistance-dominance           ; Defines the probability that an Individual SR mosquito will not be affected by either Insecticide
        ;;Number-of-water-patches        ; Defines the maximum (and initial) number of water patches
        ;;village-sense-radius           ; Defines the distance from the village at which the mosquitoes can detect the direction of the village
        ;;Set-Seed                       ; Boolean: defines whether the seed will be preset for the simulation or not
        ;;initial-SR                     ; Defines the maximum percent of SR mosquitoes that the system will allow during the burn in
        ;;Kill-Time-Mean                 ; Defines the mean of the standard deviation which determines how long after exposure to the LLA insecticide the mosquito will die
        ;;recovered-time-mean-days       ; Defines the average time (in days) that a recovered human will spend before becoming susceptible
        ;;infectious-time-mean-days      ; Defines the average time (in days) that an infectious human will spend before becoming Recovered
        ;;External-Infection             ; If this is true, every three months the number of humans defined by new-infected-humans become infected from outside the simulated world (Boolean)
        ;;new-infected-humans            ; Defines how many new humans become infected if External-Infection is on
        ;;source-control-accuracy        ; Defines what percent of the water patches the source control feature is capable of removing upon application

breed [ broods brood ]                   ; Represents the immature stages of the mosquitoes
breed [ humans human ]                   ; Susceptible humans (and infected but not infectious)
breed [ infectious a-infectious ]        ; infectious humans
breed [ recovered a-recovered ]          ; Recovered humans
breed [ mosquitoes mosquito ]            ; The mosquito agents

mosquitoes-own [ gravid                  ; Marks whether the mosquito has taken a blood meal
                 mAge                    ; Records the age of the mosquito
                 time-after-bite         ; Records the time since the mosquito took a blood meal
                 max-time-after-bite     ; Defines the time between when a mosquito takes a blood meal and when she is ready to lay her eggs
                 GCs                     ; Marks the number of Gonotrophic Cycles a mosquito has gone through
                 Infected-Time           ; Records the time since the mosquito picked up the plasmodium parasite  (Compared against EIP)
                 mInfectious             ; Shows that the mosquito is infectious  (Boolean)
                 EIP                     ; Defines the extrinsic incubation period for the mosquitoes (Random: Max 144, Min 96)
                 mGenotype               ; Recordes the genotype of each mosquito (SS, SR, RR)
                 mGenotypeM              ; Records the genotype of the father (Male) of each mosquito in the simulation
                 I-Exposed               ; Marks whether the mosquito has been exposed to the LLA insecticide
                 I-Exposed-time          ; Records how long the mosquito has been infected by the insecticide
                 mDeath-time ]           ; Defines when the mosquito will die from exposure to the LLA Insecticide, random normal

infectious-own [ time-since-bite         ; Records the time since an infecting bite (hours)
               sick-time                 ; Records how long the infectious human has been infected
               recovery                  ; Defines when the infectious humans will recover
               v1                        ; Marks the humans in the first village (boolean)
               v2                        ; Marks the humans in the second village (boolean)
               out                       ; Marks the humans who will leave the village
               r-time                    ; Counts how long the human has been recovered (hours)
               max-rtime ]               ; Defines how long the recovered human will be recovered before being susceptible

humans-own [ time-since-bite             ; Records the time since an infectious bite
             out                         ; Tells whether the human will eave the village
             v1                          ; Marks the humans in the first village (boolean)
             v2                          ; Marks the humans in the second village (boolean)
             sick-time                   ; Records how long the infectious human has been infected
             recovery                    ; Defines when the infectious human will recover
             r-time                      ; Counts how long the recovered human has been recovered
             max-rtime ]                 ; Defines how long the recovered human will be recovered

recovered-own [ r-time                   ; Counts the time the recovered human has been recovered
                out                      ; Marks the humans who leave the village
                v1                       ; Marks the humans in the first village (boolean)
                v2                       ; Marks the humans in the second village (boolean)
                time-since-bite          ; Records the time since an infecting bite
                sick-time                ; Records how long the infectious human has been infectious
                recovery                 ; Defines when the infectious humans will recover
                max-rtime]               ; Defines when the infectious human will recover

broods-own [ bAge                        ; Counts the age of the brood until it hatches; compared with bLifespan (hours)
             bLifespan                   ; Defines the amount time before a brood hatches (hours)
             mGenotype                   ; Marks the genotype of the mother of the brood (SS, SR, RR)
             mGenotypeM]                 ; Marks the genotype of the father (Male) of the brood

patches-own [ village1-scent             ; Marks the patches on which the mosquitoes can sense the closest village...
              village2-scent ]           ; used to direct the mosquitoes towards the village

;;;;;;;;;;;;;;;;;;;;;;;;;;;;;;;;;;;;;;;;;;;;;;;;;;;;;;;;;;;;;;;;;;;;;;;;;;;;;;;;;;;;;;;;;;;;;;;;;;;;;;;;;;;;;;;;;;;;;;;;;;;;;;;;;;;;;;;;;;;;;;;;;;;;;;;;;;;;;;;;;;;;;;;;;;;;;;;;;;;;;;;;;
;;;;;;;;;;;;;;;;;;;;;;;;;;;;;;;;;;;;;;;;;;;;;;;;;;;;;;;;;;;;;;;;;;;;;;;;;;;;;;;;;;;;;;;;;;;;;;;;;;;;;;;;;;;;;;;;;;;;;;;;;;;;;;;;;;;;;;;;;;;;;;;;;;;;;;;;;;;;;;;;;;;;;;;;;;;;;;;;;;;;;;;;;
;;;;;;;;;;;;;;;;;;;;;;;;;;;;;;;;;;;;;;;;;;;;;;;;;;Setup;;;;;;;;;;;;;;;;;;;;;;;;;;;;;;;;;;;;;;;;;;;;;;;;;;;;;;;;;;;;;;;;;;;;;;;;;;;;;;;;;;;;;;;;;;;;;;;;;;;;;;;;;;;;;;;;;;;;;;;;;;;;;;;;;;
;;;;;;;;;;;;;;;;;;;;;;;;;;;;;;;;;;;;;;;;;;;;;;;;;;;;;;;;;;;;;;;;;;;;;;;;;;;;;;;;;;;;;;;;;;;;;;;;;;;;;;;;;;;;;;;;;;;;;;;;;;;;;;;;;;;;;;;;;;;;;;;;;;;;;;;;;;;;;;;;;;;;;;;;;;;;;;;;;;;;;;;;;
;;;;;;;;;;;;;;;;;;;;;;;;;;;;;;;;;;;;;;;;;;;;;;;;;;;;;;;;;;;;;;;;;;;;;;;;;;;;;;;;;;;;;;;;;;;;;;;;;;;;;;;;;;;;;;;;;;;;;;;;;;;;;;;;;;;;;;;;;;;;;;;;;;;;;;;;;;;;;;;;;;;;;;;;;;;;;;;;;;;;;;;;;

to setup                                ; setup procedures called by the "setup" button
  clear-all                             ; resets the simulation from the last run
  reset-ticks
  if Set-Seed [random-seed 234543]      ; If the set-seed switch on the interface is on, this command sets the seed for the program's random number generator
  setup-turtles                         ; Initializes the turtle agents
  setup-patches                         ; Initializes the patch agents
end

to setup-turtles                        ; Procedures for initailizing the turtles
  setup-humans                             ; procedures for creating the humans agents
  setup-mosquitoes                         ; procedures for creating the mosquito agents
  setup-broods                             ; procedures for creating the brood agents
end

;;;;;;;;;;;;;;;;;;;;;;;;;;;;;;;;;;;;;;;;;;;;;;;;;;;;;;;;;;;;;;;;;;;;;;;;;;;;;;;;;;;;;;;;;;;;;;;;;;;;;;;;;;;;;;;;;;;;;;;;;;;;;;;;;;;;;;;;;;;;;;;;;;;;;;;;;;;;;;;;;;;;;;;;;;;;;;;;;;;;;;;;;;
;;;;;;;;;;;;;;;;;;;;;;;;;;;;;;;;;;;;;;;;;;;;;; Setup Turtles ;;;;;;;;;;;;;;;;;;;;;;;;;;;;;;;;;;;;;;;;;;;;;;;;;;;;;;;;;;;;;;;;;;;;;;;;;;;;;;;;;;;;;;;;;;;;;;;;;;;;;;;;;;;;;;;;;;;;;;;;;;;;;;
;;;;;;;;;;;;;;;;;;;;;;;;;;;;;;;;;;;;;;;;;;;;;;;;;;;;;;;;;;;;;;;;;;;;;;;;;;;;;;;;;;;;;;;;;;;;;;;;;;;;;;;;;;;;;;;;;;;;;;;;;;;;;;;;;;;;;;;;;;;;;;;;;;;;;;;;;;;;;;;;;;;;;;;;;;;;;;;;;;;;;;;;;;

to setup-mosquitoes                              ; Creates the mosquito agents and initializes their variables
  create-mosquitoes 10000                           ; create the mosquitoes
  [
    set shape "dot"                                 ; Sets the shape of the mosquitoes
    set size .3                                     ; Sets the size of the mosquitoes
    set mAge 0                                      ; Initializes the age counter
    set gravid false                                ; Marks the mosquito as not ready to lay eggs
    set color grey                                  ; Colors the mosquitoes grey
    setxy random-xcor random-ycor                   ; Distributes the mosquitoes randomly accross the map
    set max-time-after-bite 48                      ; Defines the gonotrophic cycle's length, is randomized for each mosquito later
    set GCs 0                                       ; Marks the mosquitoes as having no gonotrophic cycles
    set mGenotype 11                                ; 1 = susceptible 2 = resistant; so for now they are all susceptible
    set mInfectious false                           ; Make the mosquitoes not infectious...
    set Infected-Time 0                             ; Or infected with plasmodium...
    set I-Exposed false                             ; Or exposed to the LLA insecticide
  ]
   ask n-of 90 mosquitoes [ set gravid true ]       ; set an initial number of gravid mosquitoes to jumpstart the system
   ask n-of 568 mosquitoes [ set mInfectious true ] ; Infects some original mosquitoes for the Burn-In
   if initial-SR > 0 [                              ; if the initial-SR input is greater than 0
    ask n-of 44 mosquitoes [ set mGenotype 12 ]]    ; Makes some SR mosquitoes to begin with
   if Insecticide != "none" [                                                                          ; If there is an insecticide chosen...
    if Cost = "High Cost" [ set RR-death-rate 10 set RR-speed .7 set SR-death-rate 5 set SR-speed .8]  ; set the variables associated with the fitness cost setting chosen in the interface
    if Cost = "Low Cost" [ set RR-death-rate 5 set RR-speed .8 set SR-death-rate 2 set SR-speed .9]
    if Cost = "No Cost" [ set RR-death-rate 0 set RR-speed 1 set SR-death-rate 0 set SR-speed 1]]
end

to setup-broods                  ; initialize the broods' variables
  create-broods 15 [             ; create some initial broods
   set shape "default"           ; set the shape of the brood
   set bAge 1                    ; Initialize the brood's age
   set bLifespan 144             ; Initialize the brood's lifespan, will be randomized later
   setxy random-xcor random-ycor ; randomly distribute the broods
   set mGenotype 11              ; set the initial genotype
   set pcolor blue               ; make the patch the brood is on blue so they do not die right away
  ]
end

to setup-humans
  create-humans number-humans-village1       ; create the humans, then initialize their variables, all three types of human agents have the same variables so they can change breeds
  [
    set shape  "person"    ; sets the shape
    set color green        ; green = susceptible
    set size 1             ; sets size
    setxy 0 0              ; places the humans in the center of the first village
    set out false          ; Initializes a movement setting
    set v1 true            ; Marks the humans as being in village 1...
    set v2 false           ; not in village 2
  ]
  create-humans number-humans-village2    ; create the humans for village 2 and initialize their variables
  [
    set shape "person"     ; sets the shape
    set color green        ; green = susceptible
    set size 1             ; sets size
    setxy -60 60           ; places the humans in the center of the second village
    set out false          ; Initializes a movement setting
    set v2 true            ; Marks the humans as not being in village 2...
    set v1 false           ; not village 1
  ]
  ask one-of humans [ set initial-recovered-humans (.60 * (number-humans-village1 + number-humans-village2)) ; set how many initial recovered humans will be created
       set initial-infectious-humans (.19 * (number-humans-village1 + number-humans-village2) ) ]            ; set how many initial infectious humans will be created
  ask n-of initial-infectious-humans humans [ infect ]                                                       ; set some humans to infectious
  ask n-of initial-recovered-humans humans [ recover ]                                                       ; set some humans to recovered
  if-else source-control [                                                                                   ; if the source control feature is on,
    ask n-of 30 humans [ set out true]]                                                                      ; set 30 humans to leave the village (more than normal because they have to "drain the water patches")
    [ ask n-of 20 humans [ set out true]]                                                                    ; otherwise set 20 humans to travel in and out of the village
end

;;;;;;;;;;;;;;;;;;;;;;;;;;;;;;;;;;;;;;;;;;;;;;;;;;;;;;;;;;;;;;;;;;;;;;;;;;;;;;;;;;;;;;;;;;;;;;;;;;;;;;;;;;;;;;;;;;;;;;;;;;;;;;;;;;;;;;;;;;;;;;;;;;;;;;;;;;;;;;;;;;;;;;;;;;;;;;;;;;;;;;;;
;;;;;;;;;;;;;;;;;;;;;;;;;;;;;;;;;;;;;;;;;;;;;;;;;;;;;; setup patches ;;;;;;;;;;;;;;;;;;;;;;;;;;;;;;;;;;;;;;;;;;;;;;;;;;;;;;;;;;;;;;;;;;;;;;;;;;;;;;;;;;;;;;;;;;;;;;;;;;;;;;;;;;;;;;;;;;;
;;;;;;;;;;;;;;;;;;;;;;;;;;;;;;;;;;;;;;;;;;;;;;;;;;;;;;;;;;;;;;;;;;;;;;;;;;;;;;;;;;;;;;;;;;;;;;;;;;;;;;;;;;;;;;;;;;;;;;;;;;;;;;;;;;;;;;;;;;;;;;;;;;;;;;;;;;;;;;;;;;;;;;;;;;;;;;;;;;;;;;;;

to setup-patches
    setup-water          ; spreads the water patches across the map
  ask patches [          ; call the patches in the model
    setup-village1       ; define where the first village is and how far each patch is from it
    setup-village2       ; define where the second village is and how far each patch is from it
    set-red1             ; colors the first village red
    set-red2   ]         ; colors the second village red
  Setup-Insecticide      ; spreads the insecticide over the village
  set count-water-in-area (count patches with [   ; defines the number of water patches in the souce control area if that feature is activated
           pcolor = blue and distancexy -60 60 < 65 and distancexy -60 60 < distancexy 0 0 ] + count patches with [ pcolor = blue and distancexy 0 0 < 65 and distancexy 0 0 < distancexy -60 60 ])
end

to setup-village1                                           ; Sets the size of the village
  set village1 ( distancexy 0 0 ) < 15                      ; Defines the distace from the center where the village is
  set village1-scent village-sense-radius - distancexy 0 0  ; Gives the patches values which decrease as they get farther from the village which allows the mosquitoes to find the village
end

to setup-village2                                                      ; Sets the size of the village
  set village2 ( distancexy -60 60 ) < 15                              ; Defines the distace from the center where the village is
  set village2-scent village-sense-radius - distancexy -60 60          ; Gives the patches values which decrease as they get farther from the village which allows the mosquitoes to find the village
end

to set-red1                                                            ; Sets the color of the village 1 patches
  if village1 [ set pcolor red ]
end

to set-red2                                                            ; Sets the color of the village 2 patches
  if village2 [ set pcolor red ]
end

to Setup-Insecticide                    ; Defines the insecticides
  if insecticide = "instant_acting" [   ; If the instant acting insecticide is chosen,
     set-orange ]                       ; Spread it across the village
  if insecticide = "late_acting" [      ; If the late life acting insecticide insecticide is chosen,
    set-violet ]                        ; Spread it across the village
  ask n-of 1 patches [set treated-patches count patches with [pcolor = orange] + count patches with [pcolor = violet]] ; defines the number of insecticide treated patches for the resistance mitigation feature
end

to set-orange               ; Randomly spreads the instant acting insecticide on the villages
   ask n-of (count patches with [ pcolor = red ] * (IA-coverage% / 100)) patches with [ pcolor = red]  [ set pcolor orange]
end

to set-violet               ; Randomly spreads the LLA across the villages
   ask n-of (count patches with [ pcolor = red ] * (LLA-coverage% / 100)) patches with [ pcolor = red]  [ set pcolor violet]
end

to setup-water              ; Randomly spreads the water patches across the map
   ask n-of number-of-water-patches patches [ set pcolor blue ]
end

;;;;;;;;;;;;;;;;;;;;;;;;;;;;;;;;;;;;;;;;;;;;;;;;;;;;;;;;;;;;;;;;;;;;;;;;;;;;;;;;;;;;;;;;;;;;;;;;;;;;;;;;;;;;;;;;;;;;;;;;;;;;;;;;;;;;;;;;;;;;;;;;;;;;;;;;;;;;;;;;;;;;;;;;;;;;;;;;;;;;;;;;;
;;;;;;;;;;;;;;;;;;;;;;;;;;;;;;;;;;;;;;;;;;;;;;;;;;;;;;;;;;;;;;;;;;;;;;;;;;;;;;;;;;;;;;;;;;;;;;;;;;;;;;;;;;;;;;;;;;;;;;;;;;;;;;;;;;;;;;;;;;;;;;;;;;;;;;;;;;;;;;;;;;;;;;;;;;;;;;;;;;;;;;;;;
;;;;;;;;;;;;;;;;;;;;;;;;;;;;;;;;;;;;;;;;;;;;;;;;;;;;;;; go ;;;;;;;;;;;;;;;;;;;;;;;;;;;;;;;;;;;;;;;;;;;;;;;;;;;;;;;;;;;;;;;;;;;;;;;;;;;;;;;;;;;;;;;;;;;;;;;;;;;;;;;;;;;;;;;;;;;;;;;;;;;;;;
;;;;;;;;;;;;;;;;;;;;;;;;;;;;;;;;;;;;;;;;;;;;;;;;;;;;;;;;;;;;;;;;;;;;;;;;;;;;;;;;;;;;;;;;;;;;;;;;;;;;;;;;;;;;;;;;;;;;;;;;;;;;;;;;;;;;;;;;;;;;;;;;;;;;;;;;;;;;;;;;;;;;;;;;;;;;;;;;;;;;;;;;;
;;;;;;;;;;;;;;;;;;;;;;;;;;;;;;;;;;;;;;;;;;;;;;;;;;;;;;;;;;;;;;;;;;;;;;;;;;;;;;;;;;;;;;;;;;;;;;;;;;;;;;;;;;;;;;;;;;;;;;;;;;;;;;;;;;;;;;;;;;;;;;;;;;;;;;;;;;;;;;;;;;;;;;;;;;;;;;;;;;;;;;;;;

to go                                       ; These commands are called every tick
  set days (ticks / 12) - 83.3                    ; calculates the days global variable
  set years (days / 365)                          ; calculates the years global variable
  set infecting-bites/hr 0                        ; resets the bites/hour counter every simulation hour
  set bites/hr 0                                  ; resets the normbites/hour counter every hour
  if not any? mosquitoes and not any? broods [ stop ]                                ; stop the model if there are no mosquitoes
  set ss  (count mosquitoes with [ mGenotype = 11 ] / count mosquitoes) * 100        ; calculates the percentage of mosquitoes with the susceptible (SS) genotype
  set sr  (count mosquitoes with [ mGenotype = 12 ] / count mosquitoes) * 100        ; calculates the percentage of mosquitoes with the hetertozygous (SR) genotype
  set rr  (count mosquitoes with [ mGenotype = 22 ] / count mosquitoes) * 100        ; calculates the percentage of mosquitoes with the resistant (RR) genotype
     ;;;;;;;;;;;;;;;;;;;;;;;;;;;;;;;;;;;;;;;;;;;;;;;;;;;;;;;;;;;;;;;burn-in commands;;;;;;;;;;;;;;;;;;;;;;;;;;;;;;;;;;;;;;;;;;;;;;;;;;;;;;;;;;;;;;
 if ticks < (30 * recovered-time-mean-days) [                           ; this section helps initialize the human populations
    if count recovered / (count humans + count infectious) < .6 [       ; this checks if there are less than 60% recovered humans
      ask n-of 1 infectious [ recover ] ask n-of 2 humans [ recover ]]  ; and this shifts some of the other humans to recovered if there are not enough
          ]
     if rr < 8 [                      ; this section makes sure that there is at least the desired percentage of SR mosquitoes in the simulation
       if sr < (initial-SR - .2) [
          ask n-of 50 mosquitoes [ set mGenotype 12 ]] ]
    if ticks < 5380 [
      if rr < 8 [if sr > initial-SR [ ask n-of (count mosquitoes with [ mGenotype = 12] * .3) mosquitoes [ set mGenotype 11 ]]]]
     ;;;;;;;;;;;;;;;;;;;;;;;;;;;;;;;;;;;;;;;;;;;;;;;;;;;;;;;;;;;humans;;;;;;;;;;;;;;;;;;;;;;;;;;;;;;;;;;;;;;;;;;;;;;;;;;;;;;;;;;;;;;;;;;;;;;;;;;;;;;;;;;;;;;;;;;;;;;;;;;;;;;;;;;;;;;
  ask humans [                                   ; procedures for the susceptible humans in the simulation
    if-else out = false [                                          ; if the human is not one who leaves the village,
      move-humans-v1                                               ; move randomly in village 1
      move-humans-v2  ]                                            ; move randomly in village 2
    [ travel ]                                                     ; if they do leave the village use these commands
    if time-since-bite > 0 [                                       ; if the human has been bitten by an infectious mosquito,
      set time-since-bite time-since-bite + 1 ]                    ; count the time since they were bit
      if time-since-bite > 84 - random 24 [ infect ]               ; simulates the time the plasmodium parisite spends in the Exo-erythrocytic cycle
  ]
  if ticks > 1000 [if external-infection [ add-external-infection ]]  ; calls the code to add more infectious humans to simulate infections from outside sources

  ask recovered [                                ; provedures for the recovered humans in the simulation
   if-else out = false [
      move-humans-v1                                                                     ; move randomly in village 1
      move-humans-v2  ]                                                                  ; move randomly in village 2
     [travel]                                                                            ; move in and out of the village
      set r-time r-time + 1                                                              ; count the time they have been recovered
      if r-time = 2 [ set max-rtime random-normal (recovered-time-mean-days * 12) 120 ]  ; defines how long the recovered human will stay recovered for each recovered human
      if r-time > max-rtime  [ susceptible ]                                             ; if long enough, become susceptible

  ]
   ask infectious [                                ; procedures for the infectious humans in the simulation
      set sick-time sick-time + 1                                                            ; count the time the human has been sick
      if sick-time = 2 [ set recovery random-normal ( infectious-time-mean-days * 12 ) 180 ] ; at a certain point, define how long they will be sick
      if sick-time > recovery  [ recover ]                                                   ; if they have been sick long enough, recover
      if-else out = false [                                                                  ; if the humans do not go out of the village
      move-humans-v1                                                                         ; move randomly in village 1
      move-humans-v2]                                                                        ; move randomly in village 2
     [travel]                                                                                ; procedures for the humans chosen to leave the village
  ]
     ;;;;;;;;;;;;;;;;;;;;;;;;;;;;;;;;;;;;;;;;;;;;;;;;;;;;;;;;;;;;;;;;;;;;;;;;mosquitoes;;;;;;;;;;;;;;;;;;;;;;;;;;;;;;;;;;;;;;;;;;;;;;;;;;;;;;;;;;;;;;;;;;;;;;;;;;;;;;;;;;;;;;;;;;;;;
  ask mosquitoes [                                                ; mosquito procedures
    move-mosquitoes                                                             ; move
    set mAge mAge + 1                                                           ; get older
    if mAge = 2 [ set max-time-after-bite 49 - random 25                        ; defines the length of the gonotrophic cycle for each mosquito
      set EIP 145 - random 49                                                   ; defines the extrinsic incubation period for each mosquito
      if ticks > 500 [ mbreed]]                                                 ; calls the mbreed command after part of the burn-in
    bite-humans                                                                 ; mosquitoes bite a human on their patch
    get-infectious                                                              ; if that human is infectious, the mosquito becomes infected but not infectious yet
    bite-recovered                                                              ; defines how to bite a recovered human
    if Infected-Time > 0 [ set Infected-Time Infected-Time + 1 ]                ; if the mosquito has bitten an infectious human, count how long it has been infected
    if Infected-Time > EIP [ set mInfectious true  set Infected-Time 0 ]        ; if the mosquito has been infected long enough, make them infectious
    if gravid = true [                                                          ; if the mosquito is gravid,
      set time-after-bite time-after-bite + 1 ]                                 ; count how long it's been (mosquitoes can only hatch a brood if it has been two days since they bit someone)
    reproduce-mosquitoes                                                        ; make more mosquitoes
    if insecticide = "late_acting" [                                            ; if the late acting Insecticde is on,
      if mAge = 2 [death-time ]                                                 ; while the mosquitoes are young, define when they will die if they contact the LLA insecticide
      if I-Exposed = true [ set I-Exposed-Time I-Exposed-Time + 1]              ; count how long since the mosquito came in contact with the insecticide
      if I-Exposed-Time > mDeath-time [ die ] ]                                 ; mosquitoes die from the LLA after the time they have been infected is greater than their mDeath-Time
    death                                                                       ; mosquitoes have a chance of death every hour
    insecticide-1                                                               ; defines the effects of the instant acting insecticide
    insecticide-2                                                               ; defines the effects of the late acting insecticide
    if mInfectious = true [ set color yellow ]                                  ; marks the infectious mosquitoes
    if gravid = false [random-feed]   ]                                         ; the mosquitoes have a small chance of taking a blood meal outside of the humans

     ;;;;;;;;;;;;;;;;;;;;;;;;;;;;;;;;;;;;;;;;;;;;;;;;;;;;;;;;;;;;;;;;;;;;;;;;;;;;;;;;;;broods;;;;;;;;;;;;;;;;;;;;;;;;;;;;;;;;;;;;;;;;;;;;;;;;;;;;;;;;;;;;;;;;;;;;;;;;;;;;;;;;;;;;;;;
  ask broods [                                             ; commands for the brood agents
    set bAge bAge + 1                                           ; get older
    if bAge = 2 [                                               ; at a certain age, define the lifespan
      set bLifespan 144 - random 48 ]
    if bAge > bLifespan [                                       ; if old enough, develop into mosquitoes
      develop  ]
    if pcolor = black [ die ]                                   ; if the water patch turns black (dries up), die
]
     ;;;;;;;;;;;;;;;;;;;;;;;;;;;;;;;;;;;;;;;;;;;;;;;;;;;;;;;;;;;;;;;;;;;;;;;;;;;;;;;patches;;;;;;;;;;;;;;;;;;;;;;;;;;;;;;;;;;;;;;;;;;;;;;;;;;;;;;;;;;;;;;;;;;;;;;;;;;;;;;;;;;;;;;;;;
  ask patches [                                            ; commands for the patches in the model
    if pcolor = blue or pcolor = pink [                         ; if it is a water patch
      if-else count broods-here > 3 [                           ; if there are more than 3 broods on the patch
        set pcolor pink     ]                                   ; set the patch color green
       [set pcolor blue]]  ]                                    ; otherwise make the patch blue
    if resistance-mitigation [ask one-of patches[               ; if the resistance mitigation feature is active, run the code for it
       mitigate-resistance]]

  if source-control [                                           ; if the source control feature is on,
   if ticks = 5380 [                                            ; and the burn-in is over and the initial 2 control years are over,
     start-source-control                                       ; initialize the source control feature

  ]]
    if ticks > 1000 [                                                                     ; if the burn-in is over, start the water cycle
       ask n-of 4 patches with [                                                          ; select four patches to change
        pcolor != red and pcolor != orange and pcolor != violet and pcolor != red - 5] [  ; make sure these patches are not part of the villiage or the source control
            water-flow ] ]                                                                ; run the water cycle code
    ;;;;;;;;;;;;;;;;;;;;;;;;;;;;;;;;;;;;;;;;;;;;;;;;;;;;;;;;;;;;;;;;;;;;;;;;;;;;;;;;;;;;;;;;;;;;;;;;;;;;;;;;;;;;;;;;;;;;;;;;;;;;;;;;;;;;;;;;;;;;;;;;;;;;;;;;;;;;;;;;;;;;;;;;;;;;;;;;
  tick                   ; advance the clock
end

;;;;;;;;;;;;;;;;;;;;;;;;;;;;;;;;;;;;;;;;;;;;;;;;;;;;;;;;;;;;;;;;;;;;;;;;;;;;;;;;;;;;;;;;;;;;;;;;;;;;;;;;;;;;;;;;;;;;;;;;;;;;;;;;;;;;;;;;;;;;;;;;;;;;;;;;;;;;;;;;;;;;;;;;;;;;;;;;;;;;;;;;
;;;;;;;;;;;;;;;;;;;;;;;;;;;;;;;;;;;;;;;;;;;;;;;;;;;;;;;;;;;;;;;;;;;;;;;;;;;;;;;;;;;;;;;;;;;;;;;;;;;;;;;;;;;;;;;;;;;;;;;;;;;;;;;;;;;;;;;;;;;;;;;;;;;;;;;;;;;;;;;;;;;;;;;;;;;;;;;;;;;;;;;;
;;;;;;;;;;;;;;;;;;;;;;;;;;;;;;;;; Mosquito Procedures ;;;;;;;;;;;;;;;;;;;;;;;;;;;;;;;;;;;;;;;;;;;;;;;;;;;;;;;;;;;;;;;;;;;;;;;;;;;;;;;;;;;;;;;;;;;;;;;;;;;;;;;;;;;;;;;;;;;;;;;;;;;;;;;;;;
;;;;;;;;;;;;;;;;;;;;;;;;;;;;;;;;;;;;;;;;;;;;;;;;;;;;;;;;;;;;;;;;;;;;;;;;;;;;;;;;;;;;;;;;;;;;;;;;;;;;;;;;;;;;;;;;;;;;;;;;;;;;;;;;;;;;;;;;;;;;;;;;;;;;;;;;;;;;;;;;;;;;;;;;;;;;;;;;;;;;;;;;
;;;;;;;;;;;;;;;;;;;;;;;;;;;;;;;;;;;;;;;;;;;;;;;;;;;;;;;;;;;;;;;;;;;;;;;;;;;;;;;;;;;;;;;;;;;;;;;;;;;;;;;;;;;;;;;;;;;;;;;;;;;;;;;;;;;;;;;;;;;;;;;;;;;;;;;;;;;;;;;;;;;;;;;;;;;;;;;;;;;;;;;;

to death                                     ; mosquito procedure
    if random-float 100 < 1 [                        ; the mosquitoes have a 1 percent chance of death every hour
    die ]
end

to bite-humans                                       ; mosquito procedure
  if mAge > 24 [                                        ; only the adult mosquitoes bite, mosquitoes become adults 2 days after hatching
  if random 100 < 60 [                                  ; give the mosquitoes a chance of biting the humans on their patch
  if gravid = false [                                   ; the mosquitoes won't bite humans if they have recently
  let prey one-of humans-here                           ; grab a random human
  if prey != nobody  [                                  ; did we get one?  if so,
    if mInfectious = true [                             ; If the mosquito is infectious,
      ask prey [ set time-since-bite 1 ]                ; ask the human they bit to start the time-since-bit counter
      if ticks > 1000 [                                 ; and if it is after the burn-in,
        set h-infectious-bites h-infectious-bites + 1   ; count the bite as a human infecting bite
        set infecting-bites/hr infecting-bites/hr + 1]] ;count the bite in the human infecting bites per hour counter
    set gravid true                                     ; start the gonotrrophic cycle
    if ticks > 1000 [ set bites bites + 1               ; if the burn-in is over, count the bite
    set bites/hr bites/hr + 1]                          ; add the bite to the normal bites per hour counter
    set GCs GCs + 1                                     ; count how many Gonotrophic cycles the mosquito has been through
  ]]]
  ]
end

to bite-recovered                                ; mosquito procedure for biting recovered humans
 if mAge > 24 [                                        ; only the adult mosquitoes bite, mosquitoes become adults 2 days after hatching
 if random 100 < 60 [                                  ; give the mosquitoes a chance of successfully biting the humans on their patch
  if gravid = false [                                  ; the mosquitoes won't bite humans if they have recently
   let prey one-of recovered-here                      ; grab a human on that patch
   if prey != nobody  [                                ; if there are not no humans,
       set gravid true                                 ; start the gonotrophic cycle
       if ticks > 1000 [ set bites bites + 1           ; count the bite
        set bites/hr bites/hr + 1]                     ; add the bite to the normal bites per hour counter
       set GCs GCs + 1                                 ; count how many Gonotrophic cycles the mosquito has been through
    ]]
  ]]
end

to get-infectious                                       ; mosquito procedure for biting infectious humans
  if mAge > 24 [                                          ; only the adult mosquitoes bite, mosquitoes become adults 2 days after hatching
  if random 100 < 60 [                                    ; give the mosquito a chance of successfully biting a humans
      if gravid = false [                                 ; the mosquitoes won't bite humans if they have recently
         let prey one-of infectious-here                  ; grab a random infectious human
         if prey != nobody  [                             ; did we get one?  if so,
            set Infected-Time 2                           ; infect the mosquito
            set gravid true                               ; become gravid from eating
            set GCs GCs + 1                               ; count another gonotrophic cycle
            if ticks > 1000 [                             ; after the burn in
              set bites bites + 1                         ; count the bite
              set infectious-bites infectious-bites + 1 ] ; and count the infectious bites
  ]]]]
end

to mbreed                                            ; Mosquito procedure for determining the genotype of their male parent because they are not modeled explicitely
  let var random 100                                      ; choose a random number
  if-else var <= ss [ set mGenotypeM 11]                  ; if that number is less than the percent of SS mosquitoes set the male parent's genotype to be SS
    [ if-else var <= sr + ss [ set mGenotypeM 12 ]        ; if the number is less than the percent of SR mosquitoes set the male parent's genotype to be SR
      [ set mGenotypeM 22 ]]                              ; otherwise, set the genotype to be RR
end

to reproduce-mosquitoes                              ; mosquito procedure
 if pcolor = blue [                                       ; if the mosquito is on a water patch
   if-else count broods-here > 3 [fd 1][                  ; if there are too many other broods, move on
   if gravid = true  [                                    ; if they have bitten someone,
     if time-after-bite > max-time-after-bite [           ; and if they have been through their gonotrophic cycle...
          hatch-broods 1 [                                ; create a brood on the patch
            set bAge 1                                    ; initialize the brood's age
            set bLifespan 200 ]                           ; initialize the lifespan
        set gravid false                                  ; be ready to bite another human
        set time-after-bite 0                             ; be ready for another gonotrophic cycle
  ]]]]


end

to develop                                           ; brood procedure
  if-else ticks < 330 [                                   ; if in the burn-in stage...
    hatch-mosquitoes 80 [                                 ; develop into mosquitoes and initialize their variables
      set shape "dot"
      set mAge 0
      set gravid false
      set size .3
      set color grey
      ]]
 [  hatch-mosquitoes 50 - random 35 [                     ; after the burn in, hatch at most fifty mosquitoes and at least fifteen and initialize thier variables
      set shape "dot"
      set mAge 0
      set gravid false
      set color grey
  if ticks > 500 [                                        ; after some initial time, use these probabilities to determine the genotype of the mosquito
   if mGenotype = 11 [                                    ; for Mosquitoes with an SS mother...
      (ifelse
        mGenotypeM = 22 [                                 ; if the father was RR, become SR
          set mGenotype 12 ]
        mGenotypeM = 12  [
          ifelse random 100 < 50 [ set mGenotype 12 ] [set mGenotype 11 ]]         ; if the father was SR than the mosquito has 50% chance of being SR or SS
        [ set mGenotype 11 ]                                                       ; if both parents were SS then become SS
    )]
    if mGenotype = 12 [                                                              ; for mosquitoes with SR mothers...
        (ifelse
          mGenotypeM = 11 [
            if-else random 100 < 50 [ set mGenotype 12 ][ set mGenotype 11]]         ; if the father was SS then 50% of the offspring become SS and 50% SR
          mGenotypeM = 12 [
            if-else random 100 > 50 [ set mGenotype 12 ] [ if-else random 100 > 50 [ ; if the father was also SR, than 50% become SR, 25% SS, and 25% RR
              set mGenotype 11 ] [ set mGenotype 22 ]]]
         [ if-else random 100 < 50 [ set mGenotype 12 ] [ set mGenotype 22 ]]        ; If the father was RR then 50% become SR and 50% RR
     )]
    if mGenotype = 22 [                                                          ; if the mother was RR...
        (ifelse
          mGenotypeM = 22 [set mGenotype 22 ]                                    ; if both parents were RR, all offspring are RR
          mGenotypeM = 12 [
            if-else random 100 < 50 [ set mGenotype 12 ] [ set mGenotype 22 ]]   ; if the father was SR, 50% SR and 50% RR
          [ set mGenotype 12 ]                                                   ; if the father was SS all offspring become SR
    )]
    if mGenotype = 22 [                                     ; this section has to do with the cost of insecticide resistance
      if random 100 < RR-death-rate [                            ; a certain percentage of new RR mosquitoes die
          die]
   if mGenotype = 12 [
      if random 100 < SR-death-rate [                            ; a certain percentage of new SR mosquitoes die
          die ]]
    ]]]]
  die                                                            ; kill the brood after it has hatched into mosquitoes

end

to move-mosquitoes                                     ; mosquito procedures for moving
  if-else I-Exposed = true [                               ; if the mosquito is exposed to the LLA insecticide...
         fd ( cos (0.053 * I-Exposed-time * 12) ) ]        ; their movement decreases with time to simulate them having a fungal infection
   [ if mGenotype = 22 [fd RR-speed]                       ; if the mosquito is RR, move at the speed associated with the cost of being resistant
    if mGenotype = 12 [fd SR-speed]                        ; if the mosquito is SR, move at the speed associated with the cost of being SR
      if mGenotype = 11 [fd 1]                             ; if the genotype is SS, move forward a patch
  ]
    if-else gravid = false [                               ; if the mosquito is not gravid
      seek-humans]                                         ; move towards the village
  [if-else patch-ahead 4 = blue [ fd .5]                   ; if the mosquito is gravid, it moves towards any water patches more quickly if they are in front of it
     [rt random 45                                         ; otherwise it moves randomly forward
      lt random 45 ]]
end

to seek-humans                                                                ; mosquito procedure for finding the village
    let scent-ahead village-scent-at-angle   0                                      ; evaluate the distance to the village from the patch ahead
    let scent-right village-scent-at-angle  45                                      ; evaluate the distance to the village from the patch to the right
    let scent-left  village-scent-at-angle -45                                      ; evaluate the distance to the village from the patch to the left
   if-else (scent-left) < 0 and (scent-right) < 0 [ rt random 90 lt random 90 ] [   ; if the patches are too far away from the village, randomly move
    if (scent-right > scent-ahead) or (scent-left > scent-ahead) [                  ; if turning brings the mosquito closer to the village...
      if-else scent-right > scent-left                                              ; or if one direction is closer than the other,
          [ rt random 95 ]                                                          ; randomly turn in the direction which brings it closest to the direction of the village
          [ lt random 95 ] ]]
  if-else scent-left > 0 and scent-right > 0 [ fd 1.2 ][ fd 1]                      ; if the mosquito is getting close to a village, move faster
end

to-report village-scent-at-angle [angle]          ; reports which patches are closer to the village
  let p patch-right-and-ahead angle 1                        ; choose a patch to evaluate
  if p = nobody [ report 0 ]                                 ; if there is not a patch there, report 0
  if-else [village1-scent] of p > [village2-scent] of p [    ; if the patch is closer to one village than the other,
    report [village1-scent] of p]                            ; report the closest village
  [ report [village2-scent] of p]
end

to random-feed                                        ; gives the mosquitoes a random chance of finding a blood meal outside of the simulated humans
  if random-float 100 < .2 [ set gravid true ]        ; Mosquitoes have a .2% chance of taking a blood meal from a source not in the model every hour
end
;;;;;;;;;;;;;;;;;;;;;;;;;;;;;;;;;;;;;;;;;;;;;;;;;;;;;;;;;;;;;;;;;;;;;;;;;;;;;;;;;;;;;;;;;;;;;;;;;;;;;;;;;;;;;;;;;;;;;;;;;;;;;;;;;;;;;;;;;;;;;;;;;;;;;;;;;;;;;;;;;;;;;;;;;;;;;;;;;;;;;;;;;
;;;;;;;;;;;;;;;;;;;;;;;;;;;;;;;;;;;;;;;;;;;;;;;;;;;;;;;;;;;;;;;;;;;;;;;;;;;;;;;;;;;;;;;;;;;;;;;;;;;;;;;;;;;;;;;;;;;;;;;;;;;;;;;;;;;;;;;;;;;;;;;;;;;;;;;;;;;;;;;;;;;;;;;;;;;;;;;;;;;;;;;;;
;;;;;;;;;;;;;;;;;;;;;;;;;;;;;;;;;; Insecticide Procedures ;;;;;;;;;;;;;;;;;;;;;;;;;;;;;;;;;;;;;;;;;;;;;;;;;;;;;;;;;;;;;;;;;;;;;;;;;;;;;;;;;;;;;;;;;;;;;;;;;;;;;;;;;;;;;;;;;;;;;;;;;;;;;;;
;;;;;;;;;;;;;;;;;;;;;;;;;;;;;;;;;;;;;;;;;;;;;;;;;;;;;;;;;;;;;;;;;;;;;;;;;;;;;;;;;;;;;;;;;;;;;;;;;;;;;;;;;;;;;;;;;;;;;;;;;;;;;;;;;;;;;;;;;;;;;;;;;;;;;;;;;;;;;;;;;;;;;;;;;;;;;;;;;;;;;;;;;
;;;;;;;;;;;;;;;;;;;;;;;;;;;;;;;;;;;;;;;;;;;;;;;;;;;;;;;;;;;;;;;;;;;;;;;;;;;;;;;;;;;;;;;;;;;;;;;;;;;;;;;;;;;;;;;;;;;;;;;;;;;;;;;;;;;;;;;;;;;;;;;;;;;;;;;;;;;;;;;;;;;;;;;;;;;;;;;;;;;;;;;;;

to death-time                                                  ; sets the time of death for the mosquitoes effected by the late acting pesicide
  with-local-randomness [                                                    ; since there is a loop here, use local randomness
  loop [                                                                     ; the loop is used to make sure there are no negative values produced
    set mDeath-time (12 * random-normal Kill-Time-Mean death-time-deviation) ; defines the time that a mosquito will live before dieing from exposure to the LLA insecticide
    if mDeath-time > 0 [ stop ]                                              ; if the value is greater than 0, end the loop
  ]]
end

to insecticide-1                                          ; instant acting insecticide
  if ticks > 5380 [                                              ; after the burn in
    if pcolor = orange [                                         ; if the patch is orange (aka the patch has been treated with the IA insecticide)
        let t random 100                                         ; start a random variabe
        let t2 random 100                                        ; start another random variable
        let t3 random 100                                        ; start a new random variable
      if mGenotype = 11 [                                        ; if the mosquito is not resistant to the insecticide
        if t < chance-of-exposure [                              ; gives the probability that a mosquito is exposed to the insecticide
          if t2 < Death-Chance% [ die ]]]                        ; gives the probability that the insecticide kills the mosquito
      if mGenotype = 12 [                                        ; if the mosquito is SR
        if t < chance-of-exposure [                              ; gives the probability that the SR mosquito encounters the insecticide
            if t2 > resistance-dominance [                       ; gives the user a control over the effectiveness of the Resistant Allele by setting the resistance-dominance variable (higher resistance dominance makes it harder for the SR mosquito to die.)
              if t3 < Death-Chance% [die] ] ]]]]                 ; if the mosquito fits in the IA insecticide's death rate, it dies.
end

to insecticide-2                                          ; defines the effects of the late acting insecticide on the mosquitoes
  if ticks > 5380 [                                              ; after the burn in
   if pcolor = violet [                                          ; if the mosquito is on a purple patch (aka the patch has been treated with the LLA insecticide)
       let t random 100                                          ; set a random variable
       let t2 random 100                                         ; set another random variable
     if mGenotype = 11 [                                         ; if the mosquito is susceptible to the insecticide
        if t < chance-of-exposure [                              ; if the random variable is less than the chance of exposure...
          set I-Exposed true]]                                   ; become exposed the the LLA insecticide
     if mGenotype = 12 [                                         ; if the mosquito has the SR genotype
        if t < chance-of-exposure [                              ; if the random variable is less than the chance of exposure
        if t2 > resistance-dominance [                           ; if the variable is less than the resistance dominance variable...
          set I-Exposed true]   ] ]]  ]                          ; expose the mosquito to the LLA insecticide
end

;;;;;;;;;;;;;;;;;;;;;;;;;;;;;;;;;;;;;;;;;;;;;;;;;;;;;;;;;;;;;;;;;;;;;;;;;;;;;;;;;;;;;;;;;;;;;;;;;;;;;;;;;;;;;;;;;;;;;;;;;;;;;;;;;;;;;;;;;;;;;;;;;;;;;;;;;;;;;;;;;;;;;;;;;;;;;;;;;;;;;;;;;
;;;;;;;;;;;;;;;;;;;;;;;;;;;;;;;;;;;;;;;;;;;;;;;;;;;;;;;;;;;;;;;;;;;;;;;;;;;;;;;;;;;;;;;;;;;;;;;;;;;;;;;;;;;;;;;;;;;;;;;;;;;;;;;;;;;;;;;;;;;;;;;;;;;;;;;;;;;;;;;;;;;;;;;;;;;;;;;;;;;;;;;;;
;;;;;;;;;;;;;;;;;;;;;;;;;;;;;;;;;;;;;;;;;;;;;humans Procedures;;;;;;;;;;;;;;;;;;;;;;;;;;;;;;;;;;;;;;;;;;;;;;;;;;;;;;;;;;;;;;;;;;;;;;;;;;;;;;;;;;;;;;;;;;;;;;;;;;;;;;;;;;;;;;;;;;;;;;;;;;;
;;;;;;;;;;;;;;;;;;;;;;;;;;;;;;;;;;;;;;;;;;;;;;;;;;;;;;;;;;;;;;;;;;;;;;;;;;;;;;;;;;;;;;;;;;;;;;;;;;;;;;;;;;;;;;;;;;;;;;;;;;;;;;;;;;;;;;;;;;;;;;;;;;;;;;;;;;;;;;;;;;;;;;;;;;;;;;;;;;;;;;;;;
;;;;;;;;;;;;;;;;;;;;;;;;;;;;;;;;;;;;;;;;;;;;;;;;;;;;;;;;;;;;;;;;;;;;;;;;;;;;;;;;;;;;;;;;;;;;;;;;;;;;;;;;;;;;;;;;;;;;;;;;;;;;;;;;;;;;;;;;;;;;;;;;;;;;;;;;;;;;;;;;;;;;;;;;;;;;;;;;;;;;;;;;;

to move-humans-v1                           ; humans prosedure for moving
  if v1 = true [                                   ; for the humans in the first village
  rt random 180                                    ; turn randomly
  lt random 180
  if pcolor = black or pcolor = red - 5 [ home ]   ; if the patch is not in the village, return to the center of the village
    fd .5 ]                                        ; move forward
end

to move-humans-v2                           ; humans procedure for moving
  if v2 = true [                                   ; for the humans in the second village
  if-else pcolor = black or pcolor = red - 5 [     ; if the patch is not part of the village return to the center of the village
    setxy -60 60]
  [rt random 180                                   ; turn randomly
   lt random 180
    fd .5 ]]                                       ; move forward
end

to travel                                    ; humans procdure for moving in and out of the village
  if-else ticks mod 72 > 35 [fd -1 ]                    ; move in/out of the village for three days and back out/in for three days
  [ fd 1 ]
  if ticks mod 72 = 0 [ rt random 180 lt random 180 ]   ; when the humans are back in the center of the village, change their direction of movement
end

to infect                           ; humans procedure to become infectious
  set breed infectious                      ; change the breed and initialize the variables
  set color yellow - 1
  set sick-time 1
  set shape "person"
  set recovery 100
  set r-time 0
  set max-rtime 0
  set time-since-bite 0
end

to recover                          ; humans procedure to recover from being infectious
  set breed recovered                      ; change the breed and initialize the variables
  set color blue - 3
  set shape "person"
  set r-time 1
  set max-rtime 40
  set sick-time 0
  set recovery 0
  set time-since-bite 0
end

to susceptible                      ; humans procedure to become susceptible
  set color green                          ; change the breed and initialize the variables
  set breed humans
  set shape "person"
  set r-time 0
  set max-rtime 0
  set sick-time 0
  set recovery 0
  set time-since-bite 0
end

to add-external-infection                                              ; human procedure for external malaria infection
  if ticks mod ( 12 * 30 * 3) = 0 [                                           ; every three months
  if-else count humans with [ out = true] >= new-infected-humans [            ; if there are enough susceptible humans who leave the village
    ask n-of new-infected-humans humans with [ out = true] [infect]  ]        ; ask one of them to become infected from something outside the model
   [ask n-of new-infected-humans humans [ infect ]]]                          ; otherwise ask any of the susceptible humans to become infected
end

;;;;;;;;;;;;;;;;;;;;;;;;;;;;;;;;;;;;;;;;;;;;;;;;;;;;;;;;;;;;;;;;;;;;;;;;;;;;;;;;;;;;;;;;;;;;;;;;;;;;;;;;;;;;;;;;;;;;;;;;;;;;;;;;;;;;;;;;;;;;;;;;;;;;;;;;;;;;;;;;;;;;;;;;;;;;;;;;;;;;;;;;;;;;;;;
;;;;;;;;;;;;;;;;;;;;;;;;;;;;;;;;;;;;;;;;;;;;;;;;;;;;;;;;;;;;;;;;;;;;;;;;;;;;;;;;;;;;;;;;;;;;;;;;;;;;;;;;;;;;;;;;;;;;;;;;;;;;;;;;;;;;;;;;;;;;;;;;;;;;;;;;;;;;;;;;;;;;;;;;;;;;;;;;;;;;;;;;;;;;;;;
;;;;;;;;;;;;;;;;;;;;;;;;;;;;;;;;;;;;;;;;;;;;;;;;;;;;;;;;;;;;;;;;;;Patch Procedures;;;;;;;;;;;;;;;;;;;;;;;;;;;;;;;;;;;;;;;;;;;;;;;;;;;;;;;;;;;;;;;;;;;;;;;;;;;;;;;;;;;;;;;;;;;;;;;;;;;;;;;;;;;;;;;;
;;;;;;;;;;;;;;;;;;;;;;;;;;;;;;;;;;;;;;;;;;;;;;;;;;;;;;;;;;;;;;;;;;;;;;;;;;;;;;;;;;;;;;;;;;;;;;;;;;;;;;;;;;;;;;;;;;;;;;;;;;;;;;;;;;;;;;;;;;;;;;;;;;;;;;;;;;;;;;;;;;;;;;;;;;;;;;;;;;;;;;;;;;;;;;;
;;;;;;;;;;;;;;;;;;;;;;;;;;;;;;;;;;;;;;;;;;;;;;;;;;;;;;;;;;;;;;;;;;;;;;;;;;;;;;;;;;;;;;;;;;;;;;;;;;;;;;;;;;;;;;;;;;;;;;;;;;;;;;;;;;;;;;;;;;;;;;;;;;;;;;;;;;;;;;;;;;;;;;;;;;;;;;;;;;;;;;;;;;;;;;;
to water-flow                                                                                  ; patch procedure for choosing water patches. Run by 4 patches per hour
   if-else (ticks - 1000) mod 4380 > 1460 and (ticks - 1000) mod 4380 < 1825 [                       ; evaluates if it is the first rainy season (April - May)
      if count patches with [ pcolor = blue or pcolor = pink ] <= number-of-water-patches [          ; makes sure there are not too many water patches
       if random 100 < 4 [                                                                           ; gives the patches a 4% chance of becoming water
          set pcolor blue ]]                                                                         ; sets the patch blue to indicate it is water
         ]
  [ if-else (ticks - 1000) mod 4380 > 3650 and (ticks - 1000) mod 4380 < 4015 [                      ; evaluates if it is the second rainy season (October - November).
    if count patches with [ pcolor = blue or pcolor = pink ] <= number-of-water-patches [            ; if it is the rainy season, set a limit on the ammount of water
      if random 100 < 6 [                                                                            ; gives the patch a 6% chance of becoming water
          set pcolor blue ]]]                                                                        ; sets the patch blue to show it is water
       [  if pcolor = blue or pcolor = pink [                                                        ; If it is not the rainy season, if it is a water patch...
                 set pcolor black ]                                                                  ; make it a land patch
  ]]
end

to start-source-control                                                                                  ; patch procedure for setting the source control
  ask patches with [ (distancexy 0 0) > 15 and (distancexy 0 0) < 14 + source-control-size ] [                       ; get the patches in the source control area of the first village
    set pcolor red - 5 ]                                                                                             ; set their color so they will not be part of the water cycle
  ask patches with [ (distancexy -60 60) > 15 and (distancexy -60 60) < 14 + source-control-size ] [                 ; get the patches in the source control area of the second village
    set pcolor red - 5 ]                                                                                             ; set their color so they will not be part of the water cycle
  if source-control-accuracy-% > 0 [                                                                                 ; if the souce control interventin is set to be not completely accurate
        ask up-to-n-of ((1 - source-control-accuracy-% / 100) * count-water-in-area) patches with [pcolor = red - 5] ; ask a number of the patches in the intervention area defined by the accuracy percent
            [ set pcolor blue ]]                                                                                     ; to remain water patches and stay in the water cycle.
end

to mitigate-resistance                                                                               ; This code operates the resistance mitigation feature
  if min-resistant-% >= (.25 * max-resistant-%) [ set min-resistant-% (max-resistant-% * .125)]        ; make sure that the minimum resistant percent is less than the lowest level of resistance mitgation
  if ticks > 1000 [                                                                                    ; after the burn-in
    if (sr + rr) > (.25 * max-resistant-%) [                                                           ; if the Insecticide resistance (IR) reaches the first threshold...
     if mitigation-active = 0 [                                                                        ; if this level has not already been activated...
      ask n-of (.25 * treated-patches) patches with [ pcolor = violet or pcolor = orange ] [           ; find a quarter of the insecticide treated patches...
        set pcolor red                                                                                 ; and turn them into regular village patches
          set mitigation-active 1]]]                                                                   ; set the mitigation level to the first level
    if (sr + rr) > (.5 * max-resistant-%) [                                                            ; if the Insecticide resistance (IR) reaches the second threshold...
     if mitigation-active = 1 [                                                                        ; if this level has not already been activated...
      ask n-of (.25 * treated-patches) patches with [ pcolor = violet or pcolor = orange ] [           ; find a quarter of the remaining insecticide treated patches...
        set pcolor red                                                                                 ; and turn them into regular village patches
          set mitigation-active 2]]]                                                                   ; set the mitigation level to the second level
    if (sr + rr) > (.75 * max-resistant-%) [                                                           ; if the Insecticide resistance (IR) reaches the third threshold...
     if mitigation-active = 2 [                                                                        ; if this level has not already been activated...
      ask up-to-n-of (.25 * treated-patches) patches with [ pcolor = violet or pcolor = orange ] [     ; find a quarter of the remaining insecticide treated patches...
        set pcolor red                                                                                 ; and turn them into regular village patches
          set mitigation-active 3]]]                                                                   ; set the mitigation level to the third level
    if (sr + rr) >= max-resistant-% [                                                                  ; if the Insecticide resistance (IR) reaches the final threshold...
     if mitigation-active = 3 [                                                                        ; if this level has not already been activated...
      ask patches with [ pcolor = violet or pcolor = orange ] [                                        ; find the remaining insecticide treated patches...
        set pcolor red                                                                                 ; and turn them into regular village patches
          set mitigation-active 4]]]                                                                   ; set the mitigation level to the fourth level
  ]
  if ticks > 1000 [                                                                                                                                   ; after the burn-in
    if mitigation-active != 0 [                                                                                                                       ; if the resistance mitigation feature has been activated
     if (sr + rr) < min-resistant-% [                                                                                                                 ; if the IR is below the minimum threshold
        ask up-to-n-of ((treated-patches / 2) - count patches with [pcolor = violet or pcolor = orange and distancexy -60 60 < 15 ]) patches with [   ; take the original number of treated patches
          distancexy -60 60 < 15 and pcolor = red ] [                                                                                                 ; in the first village...
            if-else Insecticide = "late_acting" [set pcolor violet] [set pcolor orange]]                                                              ; and re-apply the insecticide to them
        ask up-to-n-of ((treated-patches / 2) - count patches with [pcolor = violet or pcolor = orange and distancexy 0 0 < 15]) patches with [       ; take the original number of treated patches
          distancexy 0 0 < 15 and pcolor = red] [                                                                                                     ; in the second villiage
           if-else insecticide =  "late_acting" [set pcolor violet] [set pcolor orange]]                                                              ; and re-apply the insecticide to them
        set mitigation-active 0]]                                                                                                                     ; report that there is no mitigation active in the simulation
  ]

end
@#$#@#$#@
GRAPHICS-WINDOW
1393
182
1847
637
-1
-1
2.22
1
14
1
1
1
0
1
1
1
-130
70
-70
130
1
1
1
Hours
50.0

SLIDER
1015
83
1197
116
number-humans-village1
number-humans-village1
5
600
100.0
1
1
NIL
HORIZONTAL

BUTTON
670
40
780
94
Setup
setup
NIL
1
T
OBSERVER
NIL
NIL
NIL
NIL
1

BUTTON
670
115
779
167
Go
go
T
1
T
OBSERVER
NIL
NIL
NIL
NIL
0

PLOT
649
182
1374
550
Mosquito Populations
time
population
0.0
500.0
0.0
40000.0
true
true
"" ""
PENS
"Suscepible" 1.0 0 -13791810 true "" "plot count mosquitoes with [ mGenotype = 11 ]"
"Resistant (RR)" 1.0 0 -2674135 true "" "plot count mosquitoes with [ mGenotype = 22 ]"
"Total Mosquitoes" 1.0 0 -16777216 true "" "plot count mosquitoes"
"SR Mosquitoes" 1.0 0 -955883 true "" "plot count mosquitoes with [ mGenotype = 12 ]"
"SR+RR" 1.0 0 -10141563 true "" "plot count mosquitoes with [ mGenotype = 12] + count mosquitoes with [ mGenotype = 22]"

MONITOR
650
746
800
811
Susceptible Humans
count humans
3
1
16

SLIDER
825
115
1008
148
Number-of-water-patches
Number-of-water-patches
50
1800
392.0
1
1
NIL
HORIZONTAL

MONITOR
360
388
440
445
Broods
count broods
0
1
14

MONITOR
335
458
440
515
Mosquitoes
count mosquitoes
17
1
14

MONITOR
482
243
601
316
Days
days
1
1
18

MONITOR
231
748
315
805
Bites
bites
0
1
14

CHOOSER
469
80
644
125
source-control-size
source-control-size
5 10 15 20 25 30 35 40 45 50
5

SWITCH
469
43
643
76
source-control
source-control
1
1
-1000

MONITOR
810
746
970
811
Infectious Humans
count infectious
0
1
16

MONITOR
189
388
353
445
Infectious Mosquitoes
count mosquitoes with [ mInfectious = true ]
0
1
14

MONITOR
319
749
497
806
Mosquito Infecting Bites
infectious-bites
0
1
14

CHOOSER
15
40
154
85
death-time-deviation
death-time-deviation
0 1 2 2.5 3 4 5 6 7 8 9 10
1

PLOT
25
242
434
375
Infectious Mosquitoes
time
mosquitoes
0.0
5397.0
0.0
200.0
true
true
"" ""
PENS
"Infectious * 2" 1.0 0 -16777216 true "" "plot count mosquitoes with [ mInfectious = true ] * 2"
"Infected" 1.0 0 -7500403 true "" "plot count mosquitoes with [ Infected-Time > 1 ]"

MONITOR
28
386
188
443
Infected Mosquitoes
count mosquitoes with [ Infected-Time > 0 ]
17
1
14

MONITOR
313
815
498
872
Human Infecting Bites 
h-infectious-bites
0
1
14

PLOT
650
589
1135
739
Human Populations
time
populations
0.0
100.0
0.0
100.0
true
true
"" ""
PENS
"Infectious" 1.0 0 -4079321 true "" "plot count infectious"
"Susceptible" 1.0 0 -12087248 true "" "plot count humans"
"Recovered" 1.0 0 -15390905 true "" "plot count recovered"

MONITOR
966
746
1133
811
Recovered Humans
count recovered
17
1
16

TEXTBOX
173
13
363
40
Interventions
20
0.0
1

TEXTBOX
135
215
324
242
Mosquito Statistics
20
0.0
1

TEXTBOX
825
559
1014
586
Human Statistics
20
0.0
1

TEXTBOX
1097
13
1286
40
Setup Controls
20
0.0
1

CHOOSER
15
143
112
188
LLA-Coverage%
LLA-Coverage%
5 10 15 20 25 30 40 50 60 70 75 80 90 100
5

MONITOR
28
458
147
515
 RR Mosquitoes
count mosquitoes with [ mGenotype = 22 ]
17
1
14

CHOOSER
313
42
452
87
Death-Chance%
Death-Chance%
100 90 85 80 70 75 60 50 40 30 25 20 10
3

MONITOR
28
523
118
580
RR%
100 * (count mosquitoes with [ mGenotype = 22 ] / count mosquitoes )
4
1
14

MONITOR
482
324
601
397
Years
years
3
1
18

CHOOSER
360
92
453
137
IA-coverage%
IA-coverage%
10 20 30 40 50 60 70 80 90
1

SLIDER
825
78
1008
111
resistance-dominance
resistance-dominance
0
100
25.0
1
1
NIL
HORIZONTAL

MONITOR
178
458
301
515
SR Mosquitoes
count mosquitoes with [ mGenotype = 12 ]
17
1
14

MONITOR
225
525
314
582
SS%
count mosquitoes with [ mGenotype = 11 ] / count mosquitoes * 100
5
1
14

MONITOR
129
525
209
582
SR%
count mosquitoes with [ mGenotype = 12 ] / count mosquitoes * 100
5
1
14

MONITOR
31
748
128
805
RR broods
count broods with [ mGenotype = 22 ]
17
1
14

MONITOR
130
748
228
805
SR broods
count broods with [ mGenotype = 12 ]
17
1
14

CHOOSER
165
40
304
85
Insecticide
Insecticide
"none" "late_acting" "instant_acting"
2

TEXTBOX
335
25
523
48
Instant Acting Controls
11
0.0
1

TEXTBOX
20
24
208
47
Late Acting Controls
11
0.0
1

CHOOSER
163
97
302
142
chance-of-exposure
chance-of-exposure
10 20 25 30 40 50 60 70 75 80 85 90 95 100
5

CHOOSER
163
154
302
199
Cost
Cost
"High Cost" "Low Cost" "No Cost"
1

SWITCH
1203
42
1305
75
Set-Seed
Set-Seed
0
1
-1000

SLIDER
825
43
1007
76
village-sense-radius
village-sense-radius
0
100
50.0
1
1
NIL
HORIZONTAL

TEXTBOX
489
25
677
48
Source Control
11
0.0
1

CHOOSER
1203
84
1307
129
initial-SR
initial-SR
0 0.25 0.5 0.75 1 1.25 1.5 1.75 2 2.25 2.5 2.75 3
8

CHOOSER
15
92
115
137
Kill-Time-Mean
Kill-Time-Mean
6 7 8 9 10 11 12 13 14
3

TEXTBOX
1458
655
1646
678
Blue = Water
11
105.0
1

TEXTBOX
1535
655
1723
678
Red = Village
11
15.0
1

TEXTBOX
1458
670
1646
693
Violet = LLA Insecticide
11
115.0
1

TEXTBOX
1458
686
1646
709
Orange = IA Insecticide
11
25.0
1

TEXTBOX
1629
699
1817
722
Grey = Mosquitoes
11
5.0
1

TEXTBOX
1629
655
1817
678
Green = Susceptible People
11
55.0
1

TEXTBOX
1629
669
1817
692
Yellow = Infected People
11
44.0
1

TEXTBOX
1629
685
1817
708
Dark Blue = Recovered People
11
102.0
1

TEXTBOX
1458
703
1646
726
Pink = Full Water Patches
11
135.0
1

SLIDER
1014
43
1196
76
number-humans-village2
number-humans-village2
5
600
100.0
1
1
NIL
HORIZONTAL

SLIDER
1313
85
1500
118
recovered-time-mean-days
recovered-time-mean-days
0
300
30.0
1
1
NIL
HORIZONTAL

SLIDER
1310
40
1502
73
infectious-time-mean-days
infectious-time-mean-days
0
300
30.0
1
1
NIL
HORIZONTAL

SWITCH
1504
42
1657
75
External-Infection
External-Infection
0
1
-1000

SLIDER
1505
84
1657
117
new-infected-humans
new-infected-humans
0
20
1.0
1
1
NIL
HORIZONTAL

MONITOR
31
812
161
869
SR and Infected
count mosquitoes with [ mInfectious = true and mGenotype = 12]
17
1
14

MONITOR
169
812
306
869
RR and Infected
count mosquitoes with [ mInfectious = true and mGenotype = 22]
17
1
14

SLIDER
1669
85
1842
118
max-resistant-%
max-resistant-%
0
100
20.0
1
1
NIL
HORIZONTAL

SWITCH
1668
44
1842
77
resistance-mitigation
resistance-mitigation
1
1
-1000

SLIDER
1670
127
1843
160
min-resistant-%
min-resistant-%
0
100
5.0
1
1
NIL
HORIZONTAL

MONITOR
336
525
439
582
SR% + RR%
100 * (count mosquitoes with [ mGenotype = 22 ] / count mosquitoes ) + 100 * (count mosquitoes with [ mGenotype = 12 ] / count mosquitoes )
17
1
14

SLIDER
468
129
646
162
source-control-accuracy-%
source-control-accuracy-%
0
100
80.0
1
1
NIL
HORIZONTAL

PLOT
28
588
496
741
Percent of Mosquito Population with IR
Time
% with IR
0.0
1000.0
0.0
100.0
true
true
"" ""
PENS
"RR" 1.0 0 -16777216 true "" "plot 100 * ((count mosquitoes with [ mGenotype = 22 ] + .00001) / (count mosquitoes + .0001)  )"
"SR" 1.0 0 -7500403 true "" "plot 100 * ((count mosquitoes with [ mGenotype = 12 ] + .0001) / (count mosquitoes + .0001))"
"SS" 1.0 0 -2674135 true "" "plot 100 * ((count mosquitoes with [ mGenotype = 11 ] + .0001) / (count mosquitoes + .0001))"

@#$#@#$#@
# EMMIT Program (Extendable Model for Malaria Intervention Testing)

## What is it?
This model simulates the malaria transmission cycle for a small, rural village setting. It simulates four types of agents: humans, mosquitoes, broods (aquatic mosquitoes), and patches (landscape). This model also simulates the application of several interventions to disrupt the malaria cycle: a traditional instant acting (IA) insecticide, a novel Late Life Acting (LLA) insecticide, and source control. The simulation consists of two villages where the people move and a surrounding space where the mosquitoes move between searching for people to take a blood meal and water patches to lay their eggs. 

## How To Use It
To use this model, 

  1. Choose the desired setup controls
  2. Choose the desired interventions and their specifications
  3. Hit the 'setup' button
  4. Hit the 'go' button

## Terminology
There are several important terms to know to understand this model

   * Insecticide Resistance - any development in an insect which makes it immune to the effects of an insecticide. Often conferred genetically.
   * Source Control - in our model source control is the process of draining standing water to remove the possibility of mosquitoes using it for oviposition.
   * Resistance Dominance - resistance dominance is used to describe how much resistance the resistant allele conferres to the heterozygous genotype mosquitoes. A value of 0 means that SR mosquitoes are completely susceptible, 50 means that they have a 50% chance of being affected, and 100 means that they are completely resistant. 
   * Kill Time Mean - the LLA insecticide has a normally distributed killing effect which is controlled by the Kill Time Mean and the Death-time Deviation which set the mean and standard deviation of the kill time. 
   * Set-Seed - by setting the seed for the random number generators, the simulation becomes reproducible as long as the same seed is used each time. 
   * Resistance mitigation - is a feature we developed which strategically removes the insecticide from the simulation as the IR grows in the mosquito population to reduce the selective pressure for IR. 
   * Village Sense radius - controls how far away from the village the mosquitoes can direct themselves towards the village. 
   * Change of Exposure - defines how likely it is that a mosquito encounters the insecticide when it crosses an insecticide treated patch. The higher the value, the more likely. 
   * EIP - the extrinsic incubation period is the span of time it takes for a mosquito to become infectious after taking an infected blood meal. This is often 8 to 12 days. (note how this time is longer than the average mosquito lifespan)
   * Exo-Erythrocytic Cycle - the time it takes for an infected human to become infectious. 
   * Allele - a section of a genome which confreres a specific trait.
   * If you are curious about other variables or terms check the comments in the code tab.

## Suggestions

  * To increase the number of mosquitoes, increase the water patches with the "Number-of-water-patches" slider on the interface.
  * To change the amount of malaria in the model, experiment with the "infectious-time-mean-days" and "recovered-time-mean-days" sliders on the interface.
  * To make the simulation run faster try disabling the "view updates" check-box in the top ribbon.
  * The "Days" and "Years" reporters will display as negative until the burn-in is complete and the simulation is ready to report data.
  * If you are curious about the value of a variable, you can use the "show" or "count" command in the command center to see the value. (ex: ```show infecting-bites/hr```, ```count mosquitoes with [ mGenotype = 12]```)

## Output

  * The significant outputs of this model are
    * The proportions of the humans in each of the SEIRS categories 
    * The number or percent of resistant mosquitoes
    * The number of infectious mosquitoes
    * The length of time before resistant mosquitoes developed 

## Biology

  * The important biological information for this model was found on the CDC webpage on malaria (https://www.cdc.gov/parasites/malaria/index.html)
  * The significant biological information...
    * The mosquitoes have a lifespan defined by a 1% hourly mortality
    * Only three broods can be in a water patch at a time
    * This model only simulates the evening to night hours (12 hour days)
    * The Mosquitos' gonotrophic cycles take 2 days
    * It takes at 10-14 days for the mosquito eggs to grow to adults
    * There are three types of humans: susceptible, Infectious, and Recovered
    * The extrinsic incubation period is 8-12 days 
    * The exo-erythrocytic cycle takes place over 7 days after which the human transitions from susceptible to infectious

## Late Life Acting Insecticide
The Late Life Acting (LLA) insecticide is a new type of insecticide proposed by Read et al which seeks to avoid the challenges of insecticide resistance (IR). Whereas classic insecticides kill mosquitoes on contact, the proposed LLA insecticide kills mosquitoes several days after contact so that there is less pressure to develop resistance. We modeled this insecticide with a variable mean and standard deviation so the user can experiment with many possible combinations. 


## Important Code

 * This code defines how the brood agents hatch into individual mosquito agents and determine their genotype based on their parent's genotypes. Notice how each mosquito takes into account both their father's genotype and mother's genotype and then uses random probability to determine their own genotype based on the possibilities.
```
to develop                                                ; Brood commands               
  if-else ticks < 330 [                                   ; during initialization        
    hatch-mosquitoes 80 [                                 
      set shape "dot"
      set mAge 0
      set gravid false
      set size .3
      set color grey
      ]]
 [  hatch-mosquitoes 50 - random 35 [                     ; after the burn in, hatch at most fifty mosquitoes and at least fifteen and initialize thier variables
      set shape "dot"
      set mAge 0
      set gravid false
      set color grey
  if ticks > 500 [                                        ; after some initial time, use these probabilities to determine the genotype of the mosquito
   if mGenotype = 11 [                                    ; for Mosquitoes with an SS mother...
      (ifelse
        mGenotypeM = 22 [                                 ; if the father was RR, become SR
          set mGenotype 12 ]
        mGenotypeM = 12  [
          ifelse random 100 < 50 [ set mGenotype 12 ] [set mGenotype 11 ]]         ; if the father was SR than the mosquito has 50% chance of being SR or SS
        [ set mGenotype 11 ]                                                       ; if both parents were SS then become SS
    )]
    if mGenotype = 12 [                                                              ; for mosquitoes with SR mothers...
        (ifelse
          mGenotypeM = 11 [
            if-else random 100 < 50 [ set mGenotype 12 ][ set mGenotype 11]]         ; if the father was SS then 50% of the offspring become SS and 50% SR
          mGenotypeM = 12 [
            if-else random 100 > 50 [ set mGenotype 12 ] [ if-else random 100 > 50 [ ; if the father was also SR, than 50% become SR, 25% SS, and 25% RR
              set mGenotype 11 ] [ set mGenotype 22 ]]]
         [ if-else random 100 < 50 [ set mGenotype 12 ] [ set mGenotype 22 ]]        ; If the father was RR then 50% become SR and 50% RR
     )]
    if mGenotype = 22 [                                                          ; if the mother was RR...
        (ifelse
          mGenotypeM = 22 [set mGenotype 22 ]                                    ; if both parents were RR, all offspring are RR
          mGenotypeM = 12 [
            if-else random 100 < 50 [ set mGenotype 12 ] [ set mGenotype 22 ]]   ; if the father was SR, 50% SR and 50% RR
          [ set mGenotype 12 ]                                                   ; if the father was SS all offspring become SR
    )]
    if mGenotype = 22 [                                     ; this section has to do with the cost of insecticide resistance
      if random 100 < RR-death-rate [                            ; a certain percentage of new RR mosquitoes die
          die]
   if mGenotype = 12 [
      if random 100 < SR-death-rate [                            ; a certain percentage of new SR mosquitoes die
          die ]]
    ]]]]
  die                                                            ; kill the brood after it has hatched into mosquitoes

end`
```


  
  * This code determines the length of time after exposure to the LLA insecticide before the mosquito will die from the insecticide. It uses on a normal distribution with a mean and standard deviation defined in the interface. It uses a loop to filter out any values which are less than zero. 

```
to death-time                                                  ; sets the time of death for the mosquitoes effected by the late acting pesicide
  with-local-randomness [                                                    ; since there is a loop here, use local randomness
  loop [                                                                     ; the loop is used to make sure there are no negative values produced
    set mDeath-time (12 * random-normal Kill-Time-Mean death-time-deviation) ; defines the time that a mosquito will live before dieing from exposure to the LLA insecticide
    if mDeath-time > 0 [ stop ]                                              ; if the value is greater than 0, end the loop
  ]]
end
```


 * This code causes there to be a seasonal pattern in the number of water patches in the model. 
```
to water-flow                                                                                  ; patch procedure for choosing water patches. Run by 4 patches per hour
   if-else (ticks - 1000) mod 4380 > 1460 and (ticks - 1000) mod 4380 < 1825 [                       ; evaluates if it is the first rainy season (April - May)
      if count patches with [ pcolor = blue or pcolor = pink ] <= number-of-water-patches [          ; makes sure there are not too many water patches
       if random 100 < 4 [                                                                           ; gives the patches a 4% chance of becoming water
          set pcolor blue ]]                                                                         ; sets the patch blue to indicate it is water
         ]
  [ if-else (ticks - 1000) mod 4380 > 3650 and (ticks - 1000) mod 4380 < 4015 [                      ; evaluates if it is the second rainy season (October - November).
    if count patches with [ pcolor = blue or pcolor = pink ] <= number-of-water-patches [            ; if it is the rainy season, set a limit on the ammount of water
      if random 100 < 6 [                                                                            ; gives the patch a 6% chance of becoming water
          set pcolor blue ]]]                                                                        ; sets the patch blue to show it is water
       [  if pcolor = blue or pcolor = pink [                                                        ; If it is not the rainy season, if it is a water patch...
                 set pcolor black ]                                                                  ; make it a land patch
  ]]
end
```


 * This code defines the costs of resistance differs between the SR and RR mosquitoes. It can be edited to experiment with different costs of resistance.
```
   if Insecticide != "none" [                                                                          ; If there is an insecticide chosen...
    if Cost = "High Cost" [ set RR-death-rate 10 set RR-speed .7 set SR-death-rate 5 set SR-speed .8]  ; set the variables associated with the fitness cost setting chosen in the interface
    if Cost = "Low Cost" [ set RR-death-rate 5 set RR-speed .8 set SR-death-rate 2 set SR-speed .9]
    if Cost = "No Cost" [ set RR-death-rate 0 set RR-speed 1 set SR-death-rate 0 set SR-speed 1]]
```

## Open Source
Open Source Software:  The MIT License

Permission is hereby granted, free of charge, to any person obtaining a copy of this software and associated documentation files (the "Software"), to deal in the Software without restriction, including without limitation the rights to use, copy, modify, merge, publish, distribute, sublicense, and/or sell copies of the Software, and to permit persons to whom the Software is furnished to do so, subject to the following conditions:

The above copyright notice and this permission notice shall be included in all copies or substantial portions of the Software.

THE SOFTWARE IS PROVIDED "AS IS", WITHOUT WARRANTY OF ANY KIND, EXPRESS OR IMPLIED, INCLUDING BUT NOT LIMITED TO THE WARRANTIES OF MERCHANTABILITY, FITNESS FOR A PARTICULAR PURPOSE AND NONINFRINGEMENT. IN NO EVENT SHALL THE AUTHORS OR COPYRIGHT HOLDERS BE LIABLE FOR ANY CLAIM, DAMAGES OR OTHER LIABILITY, WHETHER IN AN ACTION OF CONTRACT, TORT OR OTHERWISE, ARISING FROM, OUT OF OR IN CONNECTION WITH THE SOFTWARE OR THE USE OR OTHER DEALINGS IN THE SOFTWARE.


## Literature

Thomas, M.B & Read, A.F., (2007). Can fungal biopesticides control malaria? Nature Microbiology Reviews, 5: 377-383.


Zhou, Y. (2013). A Case Study of collecting PDA-based geo-tagged malaria-related survey data, an agent-based entomological model and two applications . PhD Dissertation, Notre Dame: Computer Science and Engineering.


Wilensky, U. (1999). NetLogo. Center for Connected Learning and Computer-Based Modeling, Northwestern University. Evanston, IL. http://ccl.northwestern.edu/netlogo/. 


Malaria. (2020, July 16). Retrieved from Centers for Disease Control and Prevention: https://www.cdc.gov/parasites/malaria/index.html
@#$#@#$#@
default
true
0
Polygon -7500403 true true 150 5 40 250 150 205 260 250

airplane
true
0
Polygon -7500403 true true 150 0 135 15 120 60 120 105 15 165 15 195 120 180 135 240 105 270 120 285 150 270 180 285 210 270 165 240 180 180 285 195 285 165 180 105 180 60 165 15

airplane 2
true
0
Polygon -7500403 true true 150 26 135 30 120 60 120 90 18 105 15 135 120 150 120 165 135 210 135 225 150 285 165 225 165 210 180 165 180 150 285 135 282 105 180 90 180 60 165 30
Line -7500403 false 120 30 180 30
Polygon -7500403 true true 105 255 120 240 180 240 195 255 180 270 120 270

arrow
true
0
Polygon -7500403 true true 150 0 0 150 105 150 105 293 195 293 195 150 300 150

bee 2
true
0
Polygon -1184463 true false 195 150 105 150 90 165 90 225 105 270 135 300 165 300 195 270 210 225 210 165 195 150
Rectangle -16777216 true false 90 165 212 185
Polygon -16777216 true false 90 207 90 226 210 226 210 207
Polygon -16777216 true false 103 266 198 266 203 246 96 246
Polygon -6459832 true false 120 150 105 135 105 75 120 60 180 60 195 75 195 135 180 150
Polygon -6459832 true false 150 15 120 30 120 60 180 60 180 30
Circle -16777216 true false 105 30 30
Circle -16777216 true false 165 30 30
Polygon -7500403 true true 120 90 75 105 15 90 30 75 120 75
Polygon -16777216 false false 120 75 30 75 15 90 75 105 120 90
Polygon -7500403 true true 180 75 180 90 225 105 285 90 270 75
Polygon -16777216 false false 180 75 270 75 285 90 225 105 180 90
Polygon -7500403 true true 180 75 180 90 195 105 240 195 270 210 285 210 285 150 255 105
Polygon -16777216 false false 180 75 255 105 285 150 285 210 270 210 240 195 195 105 180 90
Polygon -7500403 true true 120 75 45 105 15 150 15 210 30 210 60 195 105 105 120 90
Polygon -16777216 false false 120 75 45 105 15 150 15 210 30 210 60 195 105 105 120 90
Polygon -16777216 true false 135 300 165 300 180 285 120 285

box
false
0
Polygon -7500403 true true 150 285 285 225 285 75 150 135
Polygon -7500403 true true 150 135 15 75 150 15 285 75
Polygon -7500403 true true 15 75 15 225 150 285 150 135
Line -16777216 false 150 285 150 135
Line -16777216 false 150 135 15 75
Line -16777216 false 150 135 285 75

bug
true
0
Circle -7500403 true true 96 182 108
Circle -7500403 true true 110 127 80
Circle -7500403 true true 110 75 80
Line -7500403 true 150 100 80 30
Line -7500403 true 150 100 220 30

butterfly
true
0
Polygon -7500403 true true 150 165 209 199 225 225 225 255 195 270 165 255 150 240
Polygon -7500403 true true 150 165 89 198 75 225 75 255 105 270 135 255 150 240
Polygon -7500403 true true 139 148 100 105 55 90 25 90 10 105 10 135 25 180 40 195 85 194 139 163
Polygon -7500403 true true 162 150 200 105 245 90 275 90 290 105 290 135 275 180 260 195 215 195 162 165
Polygon -16777216 true false 150 255 135 225 120 150 135 120 150 105 165 120 180 150 165 225
Circle -16777216 true false 135 90 30
Line -16777216 false 150 105 195 60
Line -16777216 false 150 105 105 60

car
false
0
Polygon -7500403 true true 300 180 279 164 261 144 240 135 226 132 213 106 203 84 185 63 159 50 135 50 75 60 0 150 0 165 0 225 300 225 300 180
Circle -16777216 true false 180 180 90
Circle -16777216 true false 30 180 90
Polygon -16777216 true false 162 80 132 78 134 135 209 135 194 105 189 96 180 89
Circle -7500403 true true 47 195 58
Circle -7500403 true true 195 195 58

circle
false
0
Circle -7500403 true true 0 0 300

circle 2
false
0
Circle -7500403 true true 0 0 300
Circle -16777216 true false 30 30 240

cow
false
0
Polygon -7500403 true true 200 193 197 249 179 249 177 196 166 187 140 189 93 191 78 179 72 211 49 209 48 181 37 149 25 120 25 89 45 72 103 84 179 75 198 76 252 64 272 81 293 103 285 121 255 121 242 118 224 167
Polygon -7500403 true true 73 210 86 251 62 249 48 208
Polygon -7500403 true true 25 114 16 195 9 204 23 213 25 200 39 123

cylinder
false
0
Circle -7500403 true true 0 0 300

dot
false
0
Circle -7500403 true true 90 90 120

face happy
false
0
Circle -7500403 true true 8 8 285
Circle -16777216 true false 60 75 60
Circle -16777216 true false 180 75 60
Polygon -16777216 true false 150 255 90 239 62 213 47 191 67 179 90 203 109 218 150 225 192 218 210 203 227 181 251 194 236 217 212 240

face neutral
false
0
Circle -7500403 true true 8 7 285
Circle -16777216 true false 60 75 60
Circle -16777216 true false 180 75 60
Rectangle -16777216 true false 60 195 240 225

face sad
false
0
Circle -7500403 true true 8 8 285
Circle -16777216 true false 60 75 60
Circle -16777216 true false 180 75 60
Polygon -16777216 true false 150 168 90 184 62 210 47 232 67 244 90 220 109 205 150 198 192 205 210 220 227 242 251 229 236 206 212 183

fish
false
0
Polygon -1 true false 44 131 21 87 15 86 0 120 15 150 0 180 13 214 20 212 45 166
Polygon -1 true false 135 195 119 235 95 218 76 210 46 204 60 165
Polygon -1 true false 75 45 83 77 71 103 86 114 166 78 135 60
Polygon -7500403 true true 30 136 151 77 226 81 280 119 292 146 292 160 287 170 270 195 195 210 151 212 30 166
Circle -16777216 true false 215 106 30

flag
false
0
Rectangle -7500403 true true 60 15 75 300
Polygon -7500403 true true 90 150 270 90 90 30
Line -7500403 true 75 135 90 135
Line -7500403 true 75 45 90 45

flower
false
0
Polygon -10899396 true false 135 120 165 165 180 210 180 240 150 300 165 300 195 240 195 195 165 135
Circle -7500403 true true 85 132 38
Circle -7500403 true true 130 147 38
Circle -7500403 true true 192 85 38
Circle -7500403 true true 85 40 38
Circle -7500403 true true 177 40 38
Circle -7500403 true true 177 132 38
Circle -7500403 true true 70 85 38
Circle -7500403 true true 130 25 38
Circle -7500403 true true 96 51 108
Circle -16777216 true false 113 68 74
Polygon -10899396 true false 189 233 219 188 249 173 279 188 234 218
Polygon -10899396 true false 180 255 150 210 105 210 75 240 135 240

house
false
0
Rectangle -7500403 true true 45 120 255 285
Rectangle -16777216 true false 120 210 180 285
Polygon -7500403 true true 15 120 150 15 285 120
Line -16777216 false 30 120 270 120

leaf
false
0
Polygon -7500403 true true 150 210 135 195 120 210 60 210 30 195 60 180 60 165 15 135 30 120 15 105 40 104 45 90 60 90 90 105 105 120 120 120 105 60 120 60 135 30 150 15 165 30 180 60 195 60 180 120 195 120 210 105 240 90 255 90 263 104 285 105 270 120 285 135 240 165 240 180 270 195 240 210 180 210 165 195
Polygon -7500403 true true 135 195 135 240 120 255 105 255 105 285 135 285 165 240 165 195

line
true
0
Line -7500403 true 150 0 150 300

line half
true
0
Line -7500403 true 150 0 150 150

monster
false
0
Polygon -7500403 true true 75 150 90 195 210 195 225 150 255 120 255 45 180 0 120 0 45 45 45 120
Circle -16777216 true false 165 60 60
Circle -16777216 true false 75 60 60
Polygon -7500403 true true 225 150 285 195 285 285 255 300 255 210 180 165
Polygon -7500403 true true 75 150 15 195 15 285 45 300 45 210 120 165
Polygon -7500403 true true 210 210 225 285 195 285 165 165
Polygon -7500403 true true 90 210 75 285 105 285 135 165
Rectangle -7500403 true true 135 165 165 270

pentagon
false
0
Polygon -7500403 true true 150 15 15 120 60 285 240 285 285 120

person
false
0
Circle -7500403 true true 110 5 80
Polygon -7500403 true true 105 90 120 195 90 285 105 300 135 300 150 225 165 300 195 300 210 285 180 195 195 90
Rectangle -7500403 true true 127 79 172 94
Polygon -7500403 true true 195 90 240 150 225 180 165 105
Polygon -7500403 true true 105 90 60 150 75 180 135 105

plant
false
0
Rectangle -7500403 true true 135 90 165 300
Polygon -7500403 true true 135 255 90 210 45 195 75 255 135 285
Polygon -7500403 true true 165 255 210 210 255 195 225 255 165 285
Polygon -7500403 true true 135 180 90 135 45 120 75 180 135 210
Polygon -7500403 true true 165 180 165 210 225 180 255 120 210 135
Polygon -7500403 true true 135 105 90 60 45 45 75 105 135 135
Polygon -7500403 true true 165 105 165 135 225 105 255 45 210 60
Polygon -7500403 true true 135 90 120 45 150 15 180 45 165 90

sheep
false
15
Circle -1 true true 203 65 88
Circle -1 true true 70 65 162
Circle -1 true true 150 105 120
Polygon -7500403 true false 218 120 240 165 255 165 278 120
Circle -7500403 true false 214 72 67
Rectangle -1 true true 164 223 179 298
Polygon -1 true true 45 285 30 285 30 240 15 195 45 210
Circle -1 true true 3 83 150
Rectangle -1 true true 65 221 80 296
Polygon -1 true true 195 285 210 285 210 240 240 210 195 210
Polygon -7500403 true false 276 85 285 105 302 99 294 83
Polygon -7500403 true false 219 85 210 105 193 99 201 83

skeeter
true
7
Circle -14835848 true true 90 88 122
Circle -14835848 true true 192 116 67
Rectangle -7500403 false false 150 45 180 105
Rectangle -14835848 true true 150 45 180 105
Rectangle -14835848 true true 150 180 180 255
Line -7500403 false 285 150 255 150
Circle -14835848 true true 135 60 30
Circle -7500403 false false 135 210 30
Circle -14835848 true true 135 210 30
Line -7500403 false 195 105 210 75
Line -7500403 false 195 195 210 225
Line -7500403 false 135 90 120 60
Line -7500403 false 120 105 90 75
Line -7500403 false 105 180 75 210
Line -7500403 false 135 210 120 240

square
false
0
Rectangle -7500403 true true 30 30 270 270

square 2
false
0
Rectangle -7500403 true true 30 30 270 270
Rectangle -16777216 true false 60 60 240 240

star
false
0
Polygon -7500403 true true 151 1 185 108 298 108 207 175 242 282 151 216 59 282 94 175 3 108 116 108

target
false
0
Circle -7500403 true true 0 0 300
Circle -16777216 true false 30 30 240
Circle -7500403 true true 60 60 180
Circle -16777216 true false 90 90 120
Circle -7500403 true true 120 120 60

tree
false
0
Circle -7500403 true true 118 3 94
Rectangle -6459832 true false 120 195 180 300
Circle -7500403 true true 65 21 108
Circle -7500403 true true 116 41 127
Circle -7500403 true true 45 90 120
Circle -7500403 true true 104 74 152

triangle
false
0
Polygon -7500403 true true 150 30 15 255 285 255

triangle 2
false
0
Polygon -7500403 true true 150 30 15 255 285 255
Polygon -16777216 true false 151 99 225 223 75 224

truck
false
0
Rectangle -7500403 true true 4 45 195 187
Polygon -7500403 true true 296 193 296 150 259 134 244 104 208 104 207 194
Rectangle -1 true false 195 60 195 105
Polygon -16777216 true false 238 112 252 141 219 141 218 112
Circle -16777216 true false 234 174 42
Rectangle -7500403 true true 181 185 214 194
Circle -16777216 true false 144 174 42
Circle -16777216 true false 24 174 42
Circle -7500403 false true 24 174 42
Circle -7500403 false true 144 174 42
Circle -7500403 false true 234 174 42

turtle
true
0
Polygon -10899396 true false 215 204 240 233 246 254 228 266 215 252 193 210
Polygon -10899396 true false 195 90 225 75 245 75 260 89 269 108 261 124 240 105 225 105 210 105
Polygon -10899396 true false 105 90 75 75 55 75 40 89 31 108 39 124 60 105 75 105 90 105
Polygon -10899396 true false 132 85 134 64 107 51 108 17 150 2 192 18 192 52 169 65 172 87
Polygon -10899396 true false 85 204 60 233 54 254 72 266 85 252 107 210
Polygon -7500403 true true 119 75 179 75 209 101 224 135 220 225 175 261 128 261 81 224 74 135 88 99

wheel
false
0
Circle -7500403 true true 3 3 294
Circle -16777216 true false 30 30 240
Line -7500403 true 150 285 150 15
Line -7500403 true 15 150 285 150
Circle -7500403 true true 120 120 60
Line -7500403 true 216 40 79 269
Line -7500403 true 40 84 269 221
Line -7500403 true 40 216 269 79
Line -7500403 true 84 40 221 269

wolf
false
0
Polygon -16777216 true false 253 133 245 131 245 133
Polygon -7500403 true true 2 194 13 197 30 191 38 193 38 205 20 226 20 257 27 265 38 266 40 260 31 253 31 230 60 206 68 198 75 209 66 228 65 243 82 261 84 268 100 267 103 261 77 239 79 231 100 207 98 196 119 201 143 202 160 195 166 210 172 213 173 238 167 251 160 248 154 265 169 264 178 247 186 240 198 260 200 271 217 271 219 262 207 258 195 230 192 198 210 184 227 164 242 144 259 145 284 151 277 141 293 140 299 134 297 127 273 119 270 105
Polygon -7500403 true true -1 195 14 180 36 166 40 153 53 140 82 131 134 133 159 126 188 115 227 108 236 102 238 98 268 86 269 92 281 87 269 103 269 113

x
false
0
Polygon -7500403 true true 270 75 225 30 30 225 75 270
Polygon -7500403 true true 30 75 75 30 270 225 225 270
@#$#@#$#@
NetLogo 6.2.0
@#$#@#$#@
set model-version "sheep-wolves-grass"
set show-energy? false
setup
repeat 75 [ go ]
@#$#@#$#@
@#$#@#$#@
<experiments>
  <experiment name="Final Figure 7 Seed Sweep" repetitions="8" runMetricsEveryStep="true">
    <setup>setup</setup>
    <go>go</go>
    <timeLimit steps="79840"/>
    <metric>count mosquitoes</metric>
    <metric>count mosquitoes with [ mGenotype = 11 ]</metric>
    <metric>count mosquitoes with [ mGenotype = 12 ]</metric>
    <metric>count mosquitoes with [ mGenotype = 22 ]</metric>
    <metric>count infectious</metric>
    <metric>count humans</metric>
    <metric>count recovered</metric>
    <enumeratedValueSet variable="number-humans-village1">
      <value value="100"/>
    </enumeratedValueSet>
    <enumeratedValueSet variable="chance-of-exposure">
      <value value="50"/>
    </enumeratedValueSet>
    <enumeratedValueSet variable="number-humans-village2">
      <value value="100"/>
    </enumeratedValueSet>
    <enumeratedValueSet variable="Set-Seed">
      <value value="false"/>
    </enumeratedValueSet>
    <enumeratedValueSet variable="recovered-time-mean-days">
      <value value="30"/>
    </enumeratedValueSet>
    <enumeratedValueSet variable="new-infected-humans">
      <value value="1"/>
    </enumeratedValueSet>
    <enumeratedValueSet variable="resistance-dominance">
      <value value="50"/>
    </enumeratedValueSet>
    <enumeratedValueSet variable="initial-SR">
      <value value="2"/>
    </enumeratedValueSet>
    <enumeratedValueSet variable="source-control">
      <value value="false"/>
    </enumeratedValueSet>
    <enumeratedValueSet variable="IA-coverage%">
      <value value="30"/>
    </enumeratedValueSet>
    <enumeratedValueSet variable="death-time-deviation">
      <value value="0"/>
      <value value="1"/>
      <value value="2"/>
      <value value="3"/>
    </enumeratedValueSet>
    <enumeratedValueSet variable="Insecticide">
      <value value="&quot;late_acting&quot;"/>
    </enumeratedValueSet>
    <enumeratedValueSet variable="infectious-time-mean-days">
      <value value="30"/>
    </enumeratedValueSet>
    <enumeratedValueSet variable="source-control-accuracy-%">
      <value value="80"/>
    </enumeratedValueSet>
    <enumeratedValueSet variable="Cost">
      <value value="&quot;Low Cost&quot;"/>
    </enumeratedValueSet>
    <enumeratedValueSet variable="source-control-size">
      <value value="30"/>
    </enumeratedValueSet>
    <enumeratedValueSet variable="max-resistant-%">
      <value value="20"/>
    </enumeratedValueSet>
    <enumeratedValueSet variable="min-resistant-%">
      <value value="5"/>
    </enumeratedValueSet>
    <enumeratedValueSet variable="LLA-Coverage%">
      <value value="30"/>
    </enumeratedValueSet>
    <enumeratedValueSet variable="resistance-mitigation">
      <value value="false"/>
    </enumeratedValueSet>
    <enumeratedValueSet variable="External-Infection">
      <value value="true"/>
    </enumeratedValueSet>
    <enumeratedValueSet variable="Kill-Time-Mean">
      <value value="9"/>
    </enumeratedValueSet>
    <enumeratedValueSet variable="Death-Chance%">
      <value value="80"/>
    </enumeratedValueSet>
    <enumeratedValueSet variable="village-sense-radius">
      <value value="50"/>
    </enumeratedValueSet>
    <enumeratedValueSet variable="Number-of-water-patches">
      <value value="392"/>
    </enumeratedValueSet>
  </experiment>
  <experiment name="SEIRS Humans" repetitions="1" runMetricsEveryStep="true">
    <setup>setup</setup>
    <go>go</go>
    <timeLimit steps="20000"/>
    <metric>count humans with [ time-since-bite = 0 ]</metric>
    <metric>count humans with [ time-since-bite &gt; 0 ]</metric>
    <metric>count infectious</metric>
    <metric>count recovered</metric>
    <enumeratedValueSet variable="Insecticide">
      <value value="&quot;none&quot;"/>
    </enumeratedValueSet>
    <enumeratedValueSet variable="Kill-Time-Mean">
      <value value="8"/>
    </enumeratedValueSet>
    <enumeratedValueSet variable="death-time-deviation">
      <value value="1"/>
    </enumeratedValueSet>
    <enumeratedValueSet variable="Death-Chance%">
      <value value="80"/>
    </enumeratedValueSet>
    <enumeratedValueSet variable="chance-of-exposure">
      <value value="75"/>
    </enumeratedValueSet>
    <enumeratedValueSet variable="resistance-dominance">
      <value value="80"/>
    </enumeratedValueSet>
    <enumeratedValueSet variable="Set-Seed">
      <value value="true"/>
    </enumeratedValueSet>
    <enumeratedValueSet variable="Cost">
      <value value="&quot;No Cost&quot;"/>
    </enumeratedValueSet>
    <enumeratedValueSet variable="IA-coverage%">
      <value value="30"/>
    </enumeratedValueSet>
    <enumeratedValueSet variable="LLA-Coverage%">
      <value value="30"/>
    </enumeratedValueSet>
    <enumeratedValueSet variable="number-humans-village1">
      <value value="100"/>
    </enumeratedValueSet>
    <enumeratedValueSet variable="number-humans-village2">
      <value value="100"/>
    </enumeratedValueSet>
    <enumeratedValueSet variable="new-infected-humans">
      <value value="1"/>
    </enumeratedValueSet>
    <enumeratedValueSet variable="initial-SR">
      <value value="2"/>
    </enumeratedValueSet>
    <enumeratedValueSet variable="source-control">
      <value value="false"/>
    </enumeratedValueSet>
    <enumeratedValueSet variable="infectious-time-mean-days">
      <value value="30"/>
    </enumeratedValueSet>
    <enumeratedValueSet variable="recovered-time-mean-days">
      <value value="30"/>
    </enumeratedValueSet>
    <enumeratedValueSet variable="source-control-accuracy-%">
      <value value="80"/>
    </enumeratedValueSet>
    <enumeratedValueSet variable="source-control-size">
      <value value="30"/>
    </enumeratedValueSet>
    <enumeratedValueSet variable="max-resistant-%">
      <value value="20"/>
    </enumeratedValueSet>
    <enumeratedValueSet variable="resistance-mitigation">
      <value value="false"/>
    </enumeratedValueSet>
    <enumeratedValueSet variable="External-Infection">
      <value value="true"/>
    </enumeratedValueSet>
    <enumeratedValueSet variable="min-resistant-%">
      <value value="5"/>
    </enumeratedValueSet>
    <enumeratedValueSet variable="village-sense-radius">
      <value value="50"/>
    </enumeratedValueSet>
    <enumeratedValueSet variable="Number-of-water-patches">
      <value value="392"/>
    </enumeratedValueSet>
  </experiment>
  <experiment name="ages_measure" repetitions="1" runMetricsEveryStep="true">
    <setup>setup</setup>
    <go>go</go>
    <timeLimit steps="10000"/>
    <metric>count mosquitoes with [ mAge &lt; 12 ]</metric>
    <metric>count mosquitoes with [ mAge &gt; 12 and mAge &lt;= 24 ]</metric>
    <metric>count mosquitoes with [ mAge &gt; 24 and mAge &lt;= 36 ]</metric>
    <metric>count mosquitoes with [ mAge &gt; 36 and mAge &lt;= 48 ]</metric>
    <metric>count mosquitoes with [ mAge &gt; 48 and mAge &lt;= 60 ]</metric>
    <metric>count mosquitoes with [ mAge &gt; 60 and mAge &lt;= 72 ]</metric>
    <metric>count mosquitoes with [ mAge &gt; 72 and mAge &lt;= 84 ]</metric>
    <metric>count mosquitoes with [ mAge &gt; 84 and mAge &lt;= 96 ]</metric>
    <metric>count mosquitoes with [ mAge &gt; 96 and mAge &lt;= 108 ]</metric>
    <metric>count mosquitoes with [ mAge &gt; 108 and mAge &lt;= 120 ]</metric>
    <metric>count mosquitoes with [ mAge &gt; 120 and mAge &lt;= 132 ]</metric>
    <metric>count mosquitoes with [ mAge &gt; 132 and mAge &lt;= 144 ]</metric>
    <metric>count mosquitoes with [ mAge &gt; 144 and mAge &lt;= 156 ]</metric>
    <metric>count mosquitoes with [ mAge &gt; 156 and mAge &lt;= 168 ]</metric>
    <metric>count mosquitoes with [ mAge &gt; 168 and mAge &lt;= 180 ]</metric>
    <metric>count mosquitoes with [ mAge &gt; 180 and mAge &lt;= 192 ]</metric>
    <metric>count mosquitoes with [ mAge &gt; 192 and mAge &lt;= 204 ]</metric>
    <metric>count mosquitoes with [ mAge &gt; 204 and mAge &lt;= 216 ]</metric>
    <metric>count mosquitoes with [ mAge &gt; 216 and mAge &lt;= 228 ]</metric>
    <metric>count mosquitoes with [ mAge &gt; 228 and mAge &lt;= 240 ]</metric>
    <metric>count mosquitoes with [ mAge &gt; 240 and mAge &lt;= 252 ]</metric>
    <metric>count mosquitoes with [ mAge &gt; 252 and mAge &lt;= 264 ]</metric>
    <metric>count mosquitoes with [ mAge &gt; 264 and mAge &lt;= 276 ]</metric>
    <metric>count mosquitoes with [ mAge &gt; 278 and mAge &lt;= 290 ]</metric>
    <metric>count mosquitoes with [ mAge &gt; 290 and mAge &lt;= 302 ]</metric>
    <metric>count mosquitoes with [ mAge &gt; 302 and mAge &lt;= 314 ]</metric>
    <metric>count mosquitoes with [ mAge &gt; 314 and mAge &lt;= 326 ]</metric>
    <metric>count mosquitoes with [ mAge &gt; 326 and mAge &lt;= 338 ]</metric>
    <metric>count mosquitoes with [ mAge &gt; 338 and mAge &lt;= 350 ]</metric>
    <metric>count mosquitoes with [ mAge &gt; 350 and mAge &lt;= 362 ]</metric>
    <metric>count mosquitoes with [ mAge &gt; 362 and mAge &lt;= 374 ]</metric>
    <metric>count mosquitoes with [ mAge &gt; 374 and mAge &lt;= 386 ]</metric>
    <metric>count mosquitoes with [ mAge &gt; 386 and mAge &lt;= 398 ]</metric>
    <metric>count mosquitoes with [ mAge &gt; 410 and mAge &lt;= 422 ]</metric>
    <metric>count mosquitoes with [ mAge &gt; 434 and mAge &lt;= 446 ]</metric>
    <metric>count mosquitoes with [ mAge &gt; 446 and mAge &lt;= 458 ]</metric>
    <metric>count mosquitoes with [ mAge &gt; 458 and mAge &lt;= 470 ]</metric>
    <metric>count mosquitoes with [ mAge &gt; 470 and mAge &lt;= 482 ]</metric>
    <metric>count mosquitoes with [ mAge &gt; 482 ]</metric>
    <metric>count mosquitoes</metric>
    <enumeratedValueSet variable="Insecticide">
      <value value="&quot;none&quot;"/>
    </enumeratedValueSet>
    <enumeratedValueSet variable="Kill-Time-Mean">
      <value value="8"/>
    </enumeratedValueSet>
    <enumeratedValueSet variable="death-time-deviation">
      <value value="1"/>
    </enumeratedValueSet>
    <enumeratedValueSet variable="LLA-Coverage%">
      <value value="30"/>
    </enumeratedValueSet>
    <enumeratedValueSet variable="Death-Chance%">
      <value value="80"/>
    </enumeratedValueSet>
    <enumeratedValueSet variable="IA-coverage%">
      <value value="30"/>
    </enumeratedValueSet>
    <enumeratedValueSet variable="Set-Seed">
      <value value="true"/>
    </enumeratedValueSet>
    <enumeratedValueSet variable="resistance-dominance">
      <value value="80"/>
    </enumeratedValueSet>
    <enumeratedValueSet variable="Cost">
      <value value="&quot;No Cost&quot;"/>
    </enumeratedValueSet>
    <enumeratedValueSet variable="chance-of-exposure">
      <value value="75"/>
    </enumeratedValueSet>
    <enumeratedValueSet variable="source-control">
      <value value="false"/>
    </enumeratedValueSet>
    <enumeratedValueSet variable="source-control-size">
      <value value="30"/>
    </enumeratedValueSet>
    <enumeratedValueSet variable="source-control-accuracy-%">
      <value value="80"/>
    </enumeratedValueSet>
    <enumeratedValueSet variable="number-humans-village1">
      <value value="100"/>
    </enumeratedValueSet>
    <enumeratedValueSet variable="number-humans-village2">
      <value value="100"/>
    </enumeratedValueSet>
    <enumeratedValueSet variable="recovered-time-mean-days">
      <value value="30"/>
    </enumeratedValueSet>
    <enumeratedValueSet variable="infectious-time-mean-days">
      <value value="30"/>
    </enumeratedValueSet>
    <enumeratedValueSet variable="resistance-mitigation">
      <value value="false"/>
    </enumeratedValueSet>
    <enumeratedValueSet variable="max-resistant-%">
      <value value="20"/>
    </enumeratedValueSet>
    <enumeratedValueSet variable="min-resistant-%">
      <value value="5"/>
    </enumeratedValueSet>
    <enumeratedValueSet variable="initial-SR">
      <value value="2"/>
    </enumeratedValueSet>
    <enumeratedValueSet variable="External-Infection">
      <value value="true"/>
    </enumeratedValueSet>
    <enumeratedValueSet variable="new-infected-humans">
      <value value="1"/>
    </enumeratedValueSet>
    <enumeratedValueSet variable="village-sense-radius">
      <value value="50"/>
    </enumeratedValueSet>
    <enumeratedValueSet variable="Number-of-water-patches">
      <value value="392"/>
    </enumeratedValueSet>
  </experiment>
  <experiment name="Total Populations Random Seed Test" repetitions="10" runMetricsEveryStep="true">
    <setup>setup</setup>
    <go>go</go>
    <timeLimit steps="44800"/>
    <metric>count mosquitoes</metric>
    <metric>count mosquitoes with [ mGenotype = 11 ]</metric>
    <metric>count mosquitoes with [ mGenotype = 12 ]</metric>
    <metric>count mosquitoes with [ mGenotype = 22 ]</metric>
    <metric>count infectious</metric>
    <metric>count humans</metric>
    <metric>count recovered</metric>
    <enumeratedValueSet variable="Insecticide">
      <value value="&quot;instant_acting&quot;"/>
      <value value="&quot;late_acting&quot;"/>
      <value value="&quot;none&quot;"/>
    </enumeratedValueSet>
    <enumeratedValueSet variable="Kill-Time-Mean">
      <value value="9"/>
    </enumeratedValueSet>
    <enumeratedValueSet variable="death-time-deviation">
      <value value="1"/>
    </enumeratedValueSet>
    <enumeratedValueSet variable="LLA-Coverage%">
      <value value="30"/>
    </enumeratedValueSet>
    <enumeratedValueSet variable="Death-Chance%">
      <value value="80"/>
    </enumeratedValueSet>
    <enumeratedValueSet variable="IA-coverage%">
      <value value="10"/>
    </enumeratedValueSet>
    <enumeratedValueSet variable="Set-Seed">
      <value value="false"/>
    </enumeratedValueSet>
    <enumeratedValueSet variable="resistance-dominance">
      <value value="25"/>
    </enumeratedValueSet>
    <enumeratedValueSet variable="Cost">
      <value value="&quot;High Cost&quot;"/>
    </enumeratedValueSet>
    <enumeratedValueSet variable="chance-of-exposure">
      <value value="75"/>
    </enumeratedValueSet>
    <enumeratedValueSet variable="source-control">
      <value value="false"/>
    </enumeratedValueSet>
    <enumeratedValueSet variable="source-control-size">
      <value value="30"/>
    </enumeratedValueSet>
    <enumeratedValueSet variable="source-control-accuracy-%">
      <value value="80"/>
    </enumeratedValueSet>
    <enumeratedValueSet variable="number-humans-village1">
      <value value="100"/>
    </enumeratedValueSet>
    <enumeratedValueSet variable="number-humans-village2">
      <value value="100"/>
    </enumeratedValueSet>
    <enumeratedValueSet variable="recovered-time-mean-days">
      <value value="30"/>
    </enumeratedValueSet>
    <enumeratedValueSet variable="infectious-time-mean-days">
      <value value="30"/>
    </enumeratedValueSet>
    <enumeratedValueSet variable="resistance-mitigation">
      <value value="false"/>
    </enumeratedValueSet>
    <enumeratedValueSet variable="max-resistant-%">
      <value value="20"/>
    </enumeratedValueSet>
    <enumeratedValueSet variable="min-resistant-%">
      <value value="5"/>
    </enumeratedValueSet>
    <enumeratedValueSet variable="initial-SR">
      <value value="2"/>
    </enumeratedValueSet>
    <enumeratedValueSet variable="External-Infection">
      <value value="true"/>
    </enumeratedValueSet>
    <enumeratedValueSet variable="new-infected-humans">
      <value value="1"/>
    </enumeratedValueSet>
    <enumeratedValueSet variable="village-sense-radius">
      <value value="50"/>
    </enumeratedValueSet>
    <enumeratedValueSet variable="Number-of-water-patches">
      <value value="392"/>
    </enumeratedValueSet>
  </experiment>
  <experiment name="Instant Acting Test" repetitions="1" runMetricsEveryStep="true">
    <setup>setup</setup>
    <go>go</go>
    <timeLimit steps="44800"/>
    <metric>count mosquitoes</metric>
    <metric>count mosquitoes with [ mGenotype = 11 ]</metric>
    <metric>count mosquitoes with [ mGenotype = 12 ]</metric>
    <metric>count mosquitoes with [ mGenotype = 22 ]</metric>
    <metric>count infectious</metric>
    <metric>count humans</metric>
    <metric>count recovered</metric>
    <enumeratedValueSet variable="Insecticide">
      <value value="&quot;instant_acting&quot;"/>
    </enumeratedValueSet>
    <enumeratedValueSet variable="Kill-Time-Mean">
      <value value="9"/>
    </enumeratedValueSet>
    <enumeratedValueSet variable="death-time-deviation">
      <value value="1"/>
    </enumeratedValueSet>
    <enumeratedValueSet variable="LLA-Coverage%">
      <value value="30"/>
    </enumeratedValueSet>
    <enumeratedValueSet variable="Death-Chance%">
      <value value="80"/>
    </enumeratedValueSet>
    <enumeratedValueSet variable="IA-coverage%">
      <value value="20"/>
    </enumeratedValueSet>
    <enumeratedValueSet variable="Set-Seed">
      <value value="true"/>
    </enumeratedValueSet>
    <enumeratedValueSet variable="resistance-dominance">
      <value value="25"/>
      <value value="50"/>
      <value value="75"/>
    </enumeratedValueSet>
    <enumeratedValueSet variable="Cost">
      <value value="&quot;Low Cost&quot;"/>
      <value value="&quot;High Cost&quot;"/>
    </enumeratedValueSet>
    <enumeratedValueSet variable="chance-of-exposure">
      <value value="50"/>
      <value value="75"/>
    </enumeratedValueSet>
    <enumeratedValueSet variable="source-control">
      <value value="false"/>
    </enumeratedValueSet>
    <enumeratedValueSet variable="source-control-size">
      <value value="30"/>
    </enumeratedValueSet>
    <enumeratedValueSet variable="source-control-accuracy-%">
      <value value="80"/>
    </enumeratedValueSet>
    <enumeratedValueSet variable="number-humans-village1">
      <value value="100"/>
    </enumeratedValueSet>
    <enumeratedValueSet variable="number-humans-village2">
      <value value="100"/>
    </enumeratedValueSet>
    <enumeratedValueSet variable="recovered-time-mean-days">
      <value value="30"/>
    </enumeratedValueSet>
    <enumeratedValueSet variable="infectious-time-mean-days">
      <value value="30"/>
    </enumeratedValueSet>
    <enumeratedValueSet variable="resistance-mitigation">
      <value value="false"/>
    </enumeratedValueSet>
    <enumeratedValueSet variable="max-resistant-%">
      <value value="20"/>
    </enumeratedValueSet>
    <enumeratedValueSet variable="min-resistant-%">
      <value value="5"/>
    </enumeratedValueSet>
    <enumeratedValueSet variable="initial-SR">
      <value value="2"/>
    </enumeratedValueSet>
    <enumeratedValueSet variable="External-Infection">
      <value value="true"/>
    </enumeratedValueSet>
    <enumeratedValueSet variable="new-infected-humans">
      <value value="1"/>
    </enumeratedValueSet>
    <enumeratedValueSet variable="village-sense-radius">
      <value value="50"/>
    </enumeratedValueSet>
    <enumeratedValueSet variable="Number-of-water-patches">
      <value value="392"/>
    </enumeratedValueSet>
  </experiment>
  <experiment name="COE Sweep LLA" repetitions="1" runMetricsEveryStep="true">
    <setup>setup</setup>
    <go>go</go>
    <timeLimit steps="44800"/>
    <metric>count mosquitoes</metric>
    <metric>count mosquitoes with [ mGenotype = 11 ]</metric>
    <metric>count mosquitoes with [ mGenotype = 12 ]</metric>
    <metric>count mosquitoes with [ mGenotype = 22 ]</metric>
    <metric>count infectious</metric>
    <metric>count humans</metric>
    <metric>count recovered</metric>
    <enumeratedValueSet variable="Insecticide">
      <value value="&quot;late_acting&quot;"/>
    </enumeratedValueSet>
    <enumeratedValueSet variable="Kill-Time-Mean">
      <value value="9"/>
    </enumeratedValueSet>
    <enumeratedValueSet variable="death-time-deviation">
      <value value="0"/>
      <value value="1"/>
      <value value="2"/>
    </enumeratedValueSet>
    <enumeratedValueSet variable="LLA-Coverage%">
      <value value="30"/>
    </enumeratedValueSet>
    <enumeratedValueSet variable="Death-Chance%">
      <value value="80"/>
    </enumeratedValueSet>
    <enumeratedValueSet variable="IA-coverage%">
      <value value="20"/>
    </enumeratedValueSet>
    <enumeratedValueSet variable="Set-Seed">
      <value value="true"/>
    </enumeratedValueSet>
    <enumeratedValueSet variable="resistance-dominance">
      <value value="25"/>
    </enumeratedValueSet>
    <enumeratedValueSet variable="Cost">
      <value value="&quot;No Cost&quot;"/>
    </enumeratedValueSet>
    <enumeratedValueSet variable="chance-of-exposure">
      <value value="25"/>
      <value value="50"/>
      <value value="75"/>
      <value value="100"/>
    </enumeratedValueSet>
    <enumeratedValueSet variable="source-control">
      <value value="false"/>
    </enumeratedValueSet>
    <enumeratedValueSet variable="source-control-size">
      <value value="30"/>
    </enumeratedValueSet>
    <enumeratedValueSet variable="source-control-accuracy-%">
      <value value="80"/>
    </enumeratedValueSet>
    <enumeratedValueSet variable="number-humans-village1">
      <value value="100"/>
    </enumeratedValueSet>
    <enumeratedValueSet variable="number-humans-village2">
      <value value="100"/>
    </enumeratedValueSet>
    <enumeratedValueSet variable="recovered-time-mean-days">
      <value value="30"/>
    </enumeratedValueSet>
    <enumeratedValueSet variable="infectious-time-mean-days">
      <value value="30"/>
    </enumeratedValueSet>
    <enumeratedValueSet variable="resistance-mitigation">
      <value value="false"/>
    </enumeratedValueSet>
    <enumeratedValueSet variable="max-resistant-%">
      <value value="20"/>
    </enumeratedValueSet>
    <enumeratedValueSet variable="min-resistant-%">
      <value value="5"/>
    </enumeratedValueSet>
    <enumeratedValueSet variable="initial-SR">
      <value value="2"/>
    </enumeratedValueSet>
    <enumeratedValueSet variable="External-Infection">
      <value value="true"/>
    </enumeratedValueSet>
    <enumeratedValueSet variable="new-infected-humans">
      <value value="1"/>
    </enumeratedValueSet>
    <enumeratedValueSet variable="village-sense-radius">
      <value value="50"/>
    </enumeratedValueSet>
    <enumeratedValueSet variable="Number-of-water-patches">
      <value value="392"/>
    </enumeratedValueSet>
  </experiment>
  <experiment name="Coverage% sweep LLA" repetitions="1" runMetricsEveryStep="true">
    <setup>setup</setup>
    <go>go</go>
    <timeLimit steps="44800"/>
    <metric>count mosquitoes</metric>
    <metric>count mosquitoes with [ mGenotype = 11 ]</metric>
    <metric>count mosquitoes with [ mGenotype = 12 ]</metric>
    <metric>count mosquitoes with [ mGenotype = 22 ]</metric>
    <metric>count infectious</metric>
    <metric>count humans</metric>
    <metric>count recovered</metric>
    <enumeratedValueSet variable="Insecticide">
      <value value="&quot;late_acting&quot;"/>
    </enumeratedValueSet>
    <enumeratedValueSet variable="Kill-Time-Mean">
      <value value="9"/>
    </enumeratedValueSet>
    <enumeratedValueSet variable="death-time-deviation">
      <value value="1"/>
    </enumeratedValueSet>
    <enumeratedValueSet variable="LLA-Coverage%">
      <value value="10"/>
      <value value="20"/>
      <value value="30"/>
      <value value="40"/>
    </enumeratedValueSet>
    <enumeratedValueSet variable="Death-Chance%">
      <value value="80"/>
    </enumeratedValueSet>
    <enumeratedValueSet variable="IA-coverage%">
      <value value="20"/>
    </enumeratedValueSet>
    <enumeratedValueSet variable="Set-Seed">
      <value value="true"/>
    </enumeratedValueSet>
    <enumeratedValueSet variable="resistance-dominance">
      <value value="25"/>
    </enumeratedValueSet>
    <enumeratedValueSet variable="Cost">
      <value value="&quot;No Cost&quot;"/>
    </enumeratedValueSet>
    <enumeratedValueSet variable="chance-of-exposure">
      <value value="75"/>
    </enumeratedValueSet>
    <enumeratedValueSet variable="source-control">
      <value value="false"/>
    </enumeratedValueSet>
    <enumeratedValueSet variable="source-control-size">
      <value value="30"/>
    </enumeratedValueSet>
    <enumeratedValueSet variable="source-control-accuracy-%">
      <value value="80"/>
    </enumeratedValueSet>
    <enumeratedValueSet variable="number-humans-village1">
      <value value="100"/>
    </enumeratedValueSet>
    <enumeratedValueSet variable="number-humans-village2">
      <value value="100"/>
    </enumeratedValueSet>
    <enumeratedValueSet variable="recovered-time-mean-days">
      <value value="30"/>
    </enumeratedValueSet>
    <enumeratedValueSet variable="infectious-time-mean-days">
      <value value="30"/>
    </enumeratedValueSet>
    <enumeratedValueSet variable="resistance-mitigation">
      <value value="false"/>
    </enumeratedValueSet>
    <enumeratedValueSet variable="max-resistant-%">
      <value value="20"/>
    </enumeratedValueSet>
    <enumeratedValueSet variable="min-resistant-%">
      <value value="5"/>
    </enumeratedValueSet>
    <enumeratedValueSet variable="initial-SR">
      <value value="2"/>
    </enumeratedValueSet>
    <enumeratedValueSet variable="External-Infection">
      <value value="true"/>
    </enumeratedValueSet>
    <enumeratedValueSet variable="new-infected-humans">
      <value value="1"/>
    </enumeratedValueSet>
    <enumeratedValueSet variable="village-sense-radius">
      <value value="50"/>
    </enumeratedValueSet>
    <enumeratedValueSet variable="Number-of-water-patches">
      <value value="392"/>
    </enumeratedValueSet>
  </experiment>
  <experiment name="Standard Deviation Vs Kill Time Mean" repetitions="1" runMetricsEveryStep="true">
    <setup>setup</setup>
    <go>go</go>
    <timeLimit steps="44800"/>
    <metric>count mosquitoes</metric>
    <metric>count mosquitoes with [ mGenotype = 11 ]</metric>
    <metric>count mosquitoes with [ mGenotype = 12 ]</metric>
    <metric>count mosquitoes with [ mGenotype = 22 ]</metric>
    <metric>count infectious</metric>
    <metric>count humans</metric>
    <metric>count recovered</metric>
    <enumeratedValueSet variable="Insecticide">
      <value value="&quot;late_acting&quot;"/>
    </enumeratedValueSet>
    <enumeratedValueSet variable="Kill-Time-Mean">
      <value value="7"/>
      <value value="8"/>
      <value value="9"/>
      <value value="10"/>
      <value value="11"/>
    </enumeratedValueSet>
    <enumeratedValueSet variable="death-time-deviation">
      <value value="0"/>
      <value value="1"/>
      <value value="2"/>
      <value value="3"/>
    </enumeratedValueSet>
    <enumeratedValueSet variable="LLA-Coverage%">
      <value value="30"/>
    </enumeratedValueSet>
    <enumeratedValueSet variable="Death-Chance%">
      <value value="80"/>
    </enumeratedValueSet>
    <enumeratedValueSet variable="IA-coverage%">
      <value value="20"/>
    </enumeratedValueSet>
    <enumeratedValueSet variable="Set-Seed">
      <value value="true"/>
    </enumeratedValueSet>
    <enumeratedValueSet variable="resistance-dominance">
      <value value="25"/>
    </enumeratedValueSet>
    <enumeratedValueSet variable="Cost">
      <value value="&quot;No Cost&quot;"/>
    </enumeratedValueSet>
    <enumeratedValueSet variable="chance-of-exposure">
      <value value="75"/>
    </enumeratedValueSet>
    <enumeratedValueSet variable="source-control">
      <value value="false"/>
    </enumeratedValueSet>
    <enumeratedValueSet variable="source-control-size">
      <value value="30"/>
    </enumeratedValueSet>
    <enumeratedValueSet variable="source-control-accuracy-%">
      <value value="80"/>
    </enumeratedValueSet>
    <enumeratedValueSet variable="number-humans-village1">
      <value value="100"/>
    </enumeratedValueSet>
    <enumeratedValueSet variable="number-humans-village2">
      <value value="100"/>
    </enumeratedValueSet>
    <enumeratedValueSet variable="recovered-time-mean-days">
      <value value="30"/>
    </enumeratedValueSet>
    <enumeratedValueSet variable="infectious-time-mean-days">
      <value value="30"/>
    </enumeratedValueSet>
    <enumeratedValueSet variable="resistance-mitigation">
      <value value="false"/>
    </enumeratedValueSet>
    <enumeratedValueSet variable="max-resistant-%">
      <value value="20"/>
    </enumeratedValueSet>
    <enumeratedValueSet variable="min-resistant-%">
      <value value="5"/>
    </enumeratedValueSet>
    <enumeratedValueSet variable="initial-SR">
      <value value="2"/>
    </enumeratedValueSet>
    <enumeratedValueSet variable="External-Infection">
      <value value="true"/>
    </enumeratedValueSet>
    <enumeratedValueSet variable="new-infected-humans">
      <value value="1"/>
    </enumeratedValueSet>
    <enumeratedValueSet variable="village-sense-radius">
      <value value="50"/>
    </enumeratedValueSet>
    <enumeratedValueSet variable="Number-of-water-patches">
      <value value="392"/>
    </enumeratedValueSet>
  </experiment>
  <experiment name="Kill Time Delay 7 random seed follow-up experiment" repetitions="3" runMetricsEveryStep="true">
    <setup>setup</setup>
    <go>go</go>
    <timeLimit steps="44800"/>
    <metric>count mosquitoes</metric>
    <metric>count mosquitoes with [ mGenotype = 11 ]</metric>
    <metric>count mosquitoes with [ mGenotype = 12 ]</metric>
    <metric>count mosquitoes with [ mGenotype = 22 ]</metric>
    <metric>count infectious</metric>
    <metric>count humans</metric>
    <metric>count recovered</metric>
    <enumeratedValueSet variable="Insecticide">
      <value value="&quot;late_acting&quot;"/>
    </enumeratedValueSet>
    <enumeratedValueSet variable="Kill-Time-Mean">
      <value value="7"/>
    </enumeratedValueSet>
    <enumeratedValueSet variable="death-time-deviation">
      <value value="0"/>
      <value value="1"/>
      <value value="2"/>
      <value value="3"/>
    </enumeratedValueSet>
    <enumeratedValueSet variable="LLA-Coverage%">
      <value value="30"/>
    </enumeratedValueSet>
    <enumeratedValueSet variable="Death-Chance%">
      <value value="80"/>
    </enumeratedValueSet>
    <enumeratedValueSet variable="IA-coverage%">
      <value value="20"/>
    </enumeratedValueSet>
    <enumeratedValueSet variable="Set-Seed">
      <value value="false"/>
    </enumeratedValueSet>
    <enumeratedValueSet variable="resistance-dominance">
      <value value="25"/>
    </enumeratedValueSet>
    <enumeratedValueSet variable="Cost">
      <value value="&quot;No Cost&quot;"/>
    </enumeratedValueSet>
    <enumeratedValueSet variable="chance-of-exposure">
      <value value="75"/>
    </enumeratedValueSet>
    <enumeratedValueSet variable="source-control">
      <value value="false"/>
    </enumeratedValueSet>
    <enumeratedValueSet variable="source-control-size">
      <value value="30"/>
    </enumeratedValueSet>
    <enumeratedValueSet variable="source-control-accuracy-%">
      <value value="80"/>
    </enumeratedValueSet>
    <enumeratedValueSet variable="number-humans-village1">
      <value value="100"/>
    </enumeratedValueSet>
    <enumeratedValueSet variable="number-humans-village2">
      <value value="100"/>
    </enumeratedValueSet>
    <enumeratedValueSet variable="recovered-time-mean-days">
      <value value="30"/>
    </enumeratedValueSet>
    <enumeratedValueSet variable="infectious-time-mean-days">
      <value value="30"/>
    </enumeratedValueSet>
    <enumeratedValueSet variable="resistance-mitigation">
      <value value="false"/>
    </enumeratedValueSet>
    <enumeratedValueSet variable="max-resistant-%">
      <value value="20"/>
    </enumeratedValueSet>
    <enumeratedValueSet variable="min-resistant-%">
      <value value="5"/>
    </enumeratedValueSet>
    <enumeratedValueSet variable="initial-SR">
      <value value="2"/>
    </enumeratedValueSet>
    <enumeratedValueSet variable="External-Infection">
      <value value="true"/>
    </enumeratedValueSet>
    <enumeratedValueSet variable="new-infected-humans">
      <value value="1"/>
    </enumeratedValueSet>
    <enumeratedValueSet variable="village-sense-radius">
      <value value="50"/>
    </enumeratedValueSet>
    <enumeratedValueSet variable="Number-of-water-patches">
      <value value="392"/>
    </enumeratedValueSet>
  </experiment>
  <experiment name="Resistance Dominance sweep LLA" repetitions="1" runMetricsEveryStep="true">
    <setup>setup</setup>
    <go>go</go>
    <timeLimit steps="44800"/>
    <metric>count mosquitoes</metric>
    <metric>count mosquitoes with [ mGenotype = 11 ]</metric>
    <metric>count mosquitoes with [ mGenotype = 12 ]</metric>
    <metric>count mosquitoes with [ mGenotype = 22 ]</metric>
    <metric>count infectious</metric>
    <metric>count humans</metric>
    <metric>count recovered</metric>
    <enumeratedValueSet variable="Insecticide">
      <value value="&quot;late_acting&quot;"/>
    </enumeratedValueSet>
    <enumeratedValueSet variable="Kill-Time-Mean">
      <value value="8"/>
      <value value="9"/>
      <value value="10"/>
    </enumeratedValueSet>
    <enumeratedValueSet variable="death-time-deviation">
      <value value="1"/>
    </enumeratedValueSet>
    <enumeratedValueSet variable="LLA-Coverage%">
      <value value="30"/>
    </enumeratedValueSet>
    <enumeratedValueSet variable="Death-Chance%">
      <value value="80"/>
    </enumeratedValueSet>
    <enumeratedValueSet variable="IA-coverage%">
      <value value="20"/>
    </enumeratedValueSet>
    <enumeratedValueSet variable="Set-Seed">
      <value value="true"/>
    </enumeratedValueSet>
    <enumeratedValueSet variable="resistance-dominance">
      <value value="25"/>
      <value value="50"/>
      <value value="75"/>
      <value value="100"/>
    </enumeratedValueSet>
    <enumeratedValueSet variable="Cost">
      <value value="&quot;Low Cost&quot;"/>
    </enumeratedValueSet>
    <enumeratedValueSet variable="chance-of-exposure">
      <value value="50"/>
      <value value="75"/>
    </enumeratedValueSet>
    <enumeratedValueSet variable="source-control">
      <value value="false"/>
    </enumeratedValueSet>
    <enumeratedValueSet variable="source-control-size">
      <value value="30"/>
    </enumeratedValueSet>
    <enumeratedValueSet variable="source-control-accuracy-%">
      <value value="80"/>
    </enumeratedValueSet>
    <enumeratedValueSet variable="number-humans-village1">
      <value value="100"/>
    </enumeratedValueSet>
    <enumeratedValueSet variable="number-humans-village2">
      <value value="100"/>
    </enumeratedValueSet>
    <enumeratedValueSet variable="recovered-time-mean-days">
      <value value="30"/>
    </enumeratedValueSet>
    <enumeratedValueSet variable="infectious-time-mean-days">
      <value value="30"/>
    </enumeratedValueSet>
    <enumeratedValueSet variable="resistance-mitigation">
      <value value="false"/>
    </enumeratedValueSet>
    <enumeratedValueSet variable="max-resistant-%">
      <value value="20"/>
    </enumeratedValueSet>
    <enumeratedValueSet variable="min-resistant-%">
      <value value="5"/>
    </enumeratedValueSet>
    <enumeratedValueSet variable="initial-SR">
      <value value="2"/>
    </enumeratedValueSet>
    <enumeratedValueSet variable="External-Infection">
      <value value="true"/>
    </enumeratedValueSet>
    <enumeratedValueSet variable="new-infected-humans">
      <value value="1"/>
    </enumeratedValueSet>
    <enumeratedValueSet variable="village-sense-radius">
      <value value="50"/>
    </enumeratedValueSet>
    <enumeratedValueSet variable="Number-of-water-patches">
      <value value="392"/>
    </enumeratedValueSet>
  </experiment>
  <experiment name="Random Seed sensitivity analysis LLA" repetitions="10" runMetricsEveryStep="true">
    <setup>setup</setup>
    <go>go</go>
    <timeLimit steps="44800"/>
    <metric>count mosquitoes</metric>
    <metric>count mosquitoes with [ mGenotype = 11 ]</metric>
    <metric>count mosquitoes with [ mGenotype = 12 ]</metric>
    <metric>count mosquitoes with [ mGenotype = 22 ]</metric>
    <metric>count infectious</metric>
    <metric>count humans</metric>
    <metric>count recovered</metric>
    <enumeratedValueSet variable="Insecticide">
      <value value="&quot;late_acting&quot;"/>
    </enumeratedValueSet>
    <enumeratedValueSet variable="Kill-Time-Mean">
      <value value="9"/>
    </enumeratedValueSet>
    <enumeratedValueSet variable="death-time-deviation">
      <value value="1"/>
    </enumeratedValueSet>
    <enumeratedValueSet variable="LLA-Coverage%">
      <value value="30"/>
    </enumeratedValueSet>
    <enumeratedValueSet variable="Death-Chance%">
      <value value="80"/>
    </enumeratedValueSet>
    <enumeratedValueSet variable="IA-coverage%">
      <value value="20"/>
    </enumeratedValueSet>
    <enumeratedValueSet variable="Set-Seed">
      <value value="false"/>
    </enumeratedValueSet>
    <enumeratedValueSet variable="resistance-dominance">
      <value value="70"/>
    </enumeratedValueSet>
    <enumeratedValueSet variable="Cost">
      <value value="&quot;Low Cost&quot;"/>
    </enumeratedValueSet>
    <enumeratedValueSet variable="chance-of-exposure">
      <value value="75"/>
    </enumeratedValueSet>
    <enumeratedValueSet variable="source-control">
      <value value="false"/>
    </enumeratedValueSet>
    <enumeratedValueSet variable="source-control-size">
      <value value="30"/>
    </enumeratedValueSet>
    <enumeratedValueSet variable="source-control-accuracy-%">
      <value value="80"/>
    </enumeratedValueSet>
    <enumeratedValueSet variable="number-humans-village1">
      <value value="100"/>
    </enumeratedValueSet>
    <enumeratedValueSet variable="number-humans-village2">
      <value value="100"/>
    </enumeratedValueSet>
    <enumeratedValueSet variable="recovered-time-mean-days">
      <value value="30"/>
    </enumeratedValueSet>
    <enumeratedValueSet variable="infectious-time-mean-days">
      <value value="30"/>
    </enumeratedValueSet>
    <enumeratedValueSet variable="resistance-mitigation">
      <value value="false"/>
    </enumeratedValueSet>
    <enumeratedValueSet variable="max-resistant-%">
      <value value="20"/>
    </enumeratedValueSet>
    <enumeratedValueSet variable="min-resistant-%">
      <value value="5"/>
    </enumeratedValueSet>
    <enumeratedValueSet variable="initial-SR">
      <value value="2"/>
    </enumeratedValueSet>
    <enumeratedValueSet variable="External-Infection">
      <value value="true"/>
    </enumeratedValueSet>
    <enumeratedValueSet variable="new-infected-humans">
      <value value="1"/>
    </enumeratedValueSet>
    <enumeratedValueSet variable="village-sense-radius">
      <value value="50"/>
    </enumeratedValueSet>
    <enumeratedValueSet variable="Number-of-water-patches">
      <value value="392"/>
    </enumeratedValueSet>
  </experiment>
  <experiment name="Source Control experiment" repetitions="1" runMetricsEveryStep="true">
    <setup>setup</setup>
    <go>go</go>
    <timeLimit steps="44800"/>
    <metric>count mosquitoes</metric>
    <metric>count mosquitoes with [ mGenotype = 11 ]</metric>
    <metric>count mosquitoes with [ mGenotype = 12 ]</metric>
    <metric>count mosquitoes with [ mGenotype = 22 ]</metric>
    <metric>count infectious</metric>
    <metric>count humans</metric>
    <metric>count recovered</metric>
    <enumeratedValueSet variable="Insecticide">
      <value value="&quot;none&quot;"/>
    </enumeratedValueSet>
    <enumeratedValueSet variable="Kill-Time-Mean">
      <value value="9"/>
    </enumeratedValueSet>
    <enumeratedValueSet variable="death-time-deviation">
      <value value="1"/>
    </enumeratedValueSet>
    <enumeratedValueSet variable="LLA-Coverage%">
      <value value="10"/>
    </enumeratedValueSet>
    <enumeratedValueSet variable="Death-Chance%">
      <value value="80"/>
    </enumeratedValueSet>
    <enumeratedValueSet variable="IA-coverage%">
      <value value="20"/>
    </enumeratedValueSet>
    <enumeratedValueSet variable="Set-Seed">
      <value value="true"/>
    </enumeratedValueSet>
    <enumeratedValueSet variable="resistance-dominance">
      <value value="25"/>
    </enumeratedValueSet>
    <enumeratedValueSet variable="Cost">
      <value value="&quot;No Cost&quot;"/>
    </enumeratedValueSet>
    <enumeratedValueSet variable="chance-of-exposure">
      <value value="75"/>
    </enumeratedValueSet>
    <enumeratedValueSet variable="source-control">
      <value value="true"/>
    </enumeratedValueSet>
    <enumeratedValueSet variable="source-control-size">
      <value value="10"/>
      <value value="20"/>
      <value value="30"/>
      <value value="40"/>
      <value value="50"/>
    </enumeratedValueSet>
    <enumeratedValueSet variable="source-control-accuracy-%">
      <value value="80"/>
    </enumeratedValueSet>
    <enumeratedValueSet variable="number-humans-village1">
      <value value="100"/>
    </enumeratedValueSet>
    <enumeratedValueSet variable="number-humans-village2">
      <value value="100"/>
    </enumeratedValueSet>
    <enumeratedValueSet variable="recovered-time-mean-days">
      <value value="30"/>
    </enumeratedValueSet>
    <enumeratedValueSet variable="infectious-time-mean-days">
      <value value="30"/>
    </enumeratedValueSet>
    <enumeratedValueSet variable="resistance-mitigation">
      <value value="false"/>
    </enumeratedValueSet>
    <enumeratedValueSet variable="max-resistant-%">
      <value value="20"/>
    </enumeratedValueSet>
    <enumeratedValueSet variable="min-resistant-%">
      <value value="5"/>
    </enumeratedValueSet>
    <enumeratedValueSet variable="initial-SR">
      <value value="2"/>
    </enumeratedValueSet>
    <enumeratedValueSet variable="External-Infection">
      <value value="true"/>
    </enumeratedValueSet>
    <enumeratedValueSet variable="new-infected-humans">
      <value value="1"/>
    </enumeratedValueSet>
    <enumeratedValueSet variable="village-sense-radius">
      <value value="50"/>
    </enumeratedValueSet>
    <enumeratedValueSet variable="Number-of-water-patches">
      <value value="392"/>
    </enumeratedValueSet>
  </experiment>
  <experiment name="Resistance Mitigation Policy experiment" repetitions="1" runMetricsEveryStep="true">
    <setup>setup</setup>
    <go>go</go>
    <timeLimit steps="53560"/>
    <metric>count mosquitoes</metric>
    <metric>count mosquitoes with [ mGenotype = 11 ]</metric>
    <metric>count mosquitoes with [ mGenotype = 12 ]</metric>
    <metric>count mosquitoes with [ mGenotype = 22 ]</metric>
    <metric>count infectious</metric>
    <metric>count humans</metric>
    <metric>count recovered</metric>
    <enumeratedValueSet variable="Insecticide">
      <value value="&quot;late_acting&quot;"/>
    </enumeratedValueSet>
    <enumeratedValueSet variable="Kill-Time-Mean">
      <value value="8"/>
    </enumeratedValueSet>
    <enumeratedValueSet variable="death-time-deviation">
      <value value="1"/>
    </enumeratedValueSet>
    <enumeratedValueSet variable="LLA-Coverage%">
      <value value="30"/>
    </enumeratedValueSet>
    <enumeratedValueSet variable="Death-Chance%">
      <value value="80"/>
    </enumeratedValueSet>
    <enumeratedValueSet variable="IA-coverage%">
      <value value="20"/>
    </enumeratedValueSet>
    <enumeratedValueSet variable="Set-Seed">
      <value value="true"/>
    </enumeratedValueSet>
    <enumeratedValueSet variable="resistance-dominance">
      <value value="80"/>
    </enumeratedValueSet>
    <enumeratedValueSet variable="Cost">
      <value value="&quot;High Cost&quot;"/>
    </enumeratedValueSet>
    <enumeratedValueSet variable="chance-of-exposure">
      <value value="75"/>
    </enumeratedValueSet>
    <enumeratedValueSet variable="source-control">
      <value value="false"/>
    </enumeratedValueSet>
    <enumeratedValueSet variable="source-control-size">
      <value value="30"/>
    </enumeratedValueSet>
    <enumeratedValueSet variable="source-control-accuracy-%">
      <value value="80"/>
    </enumeratedValueSet>
    <enumeratedValueSet variable="number-humans-village1">
      <value value="100"/>
    </enumeratedValueSet>
    <enumeratedValueSet variable="number-humans-village2">
      <value value="100"/>
    </enumeratedValueSet>
    <enumeratedValueSet variable="recovered-time-mean-days">
      <value value="30"/>
    </enumeratedValueSet>
    <enumeratedValueSet variable="infectious-time-mean-days">
      <value value="30"/>
    </enumeratedValueSet>
    <enumeratedValueSet variable="resistance-mitigation">
      <value value="true"/>
    </enumeratedValueSet>
    <enumeratedValueSet variable="max-resistant-%">
      <value value="20"/>
      <value value="30"/>
      <value value="40"/>
      <value value="50"/>
    </enumeratedValueSet>
    <enumeratedValueSet variable="min-resistant-%">
      <value value="5"/>
    </enumeratedValueSet>
    <enumeratedValueSet variable="initial-SR">
      <value value="2"/>
    </enumeratedValueSet>
    <enumeratedValueSet variable="External-Infection">
      <value value="true"/>
    </enumeratedValueSet>
    <enumeratedValueSet variable="new-infected-humans">
      <value value="1"/>
    </enumeratedValueSet>
    <enumeratedValueSet variable="village-sense-radius">
      <value value="50"/>
    </enumeratedValueSet>
    <enumeratedValueSet variable="Number-of-water-patches">
      <value value="392"/>
    </enumeratedValueSet>
  </experiment>
</experiments>
@#$#@#$#@
@#$#@#$#@
default
0.0
-0.2 0 0.0 1.0
0.0 1 1.0 0.0
0.2 0 0.0 1.0
link direction
true
0
Line -7500403 true 150 150 90 180
Line -7500403 true 150 150 210 180
@#$#@#$#@
1
@#$#@#$#@
````
