## Supplementary material for "Case Study: End User Development of an Agent-based Model of Malaria Transmission to Support the Design of Late-Life-Acting Insecticides for the Control of Malaria Transmission and Delay of the Evolution of Insecticide Resistance": S2 - ODD Documentation

The simulation presented in this paper is documented below using the standard ODD format (Grimm2006, Grimm2010, Grimm2017, Grimm2020). The implementation of the model as a computer simulation using NetLogo 6.2.0 can be found in supporting file S1 Program.

1. Purpose
   - 1.1. To support the design of the late-acting insecticides, which are yet to be developed.
   - 1.2. Evaluation and comparison of the effectiveness of two insecticides types for malaria control with instant-acting and late-life-acting killing properties.
     - 1.2.1. Delay of IR (insecticide resistance)
     - 1.2.2. Effect on transmission
     - 1.2.3. Interaction with other control methods, such as source control
   - 1.3. Demonstrate that agent-based models built on easy-to-use tool-kits such as NetLogo, can be rapidly developed, deployed, and validated by non-computer scientists (end-user-developers).
2. Entities, state variables, and scales
   - 2.1. Six types of entities
     - 2.1.1. Mosquitoes – female mosquito agents
     - 2.1.2. Broods – a cohort agent of mosquito eggs (laid by a female per one gonotrophic cycle) and the following aquatic life stages, prior to emergence as individual mosquitoes
     - 2.1.3. Susceptible people – human agents susceptible to malaria, or exposed to an infectious bite but not yet infectious
     - 2.1.4. Infectious people – human agents infectious after a bite from an infectious mosquito and able to infect mosquitoes
     - 2.1.5. Recovered people – human agents who have cleared the malaria parasite and have immunity for a period of time (after which they become susceptible)
     - 2.1.6. Patches – agents representing "patches" of the simulated world that the other agents can occupy; the patches form a grid with horizontal and vertical wrapping
   - 2.2. State Variables for Each Entity (for additional variable documentation, see the other supporting documents)
     - 2.2.1. Global variables (not specific to any agentset but can be set by any agent or in the interface)
       - 2.2.1.1. <u>Days</u>: Counts the days the simulation has run for
       - 2.2.1.2. years: Counts the years the simulation has run for
       - 2.2.1.3. <u>RR-death-rate</u>: Defines the death rate for RR mosquitoes
       - 2.2.1.4. <u>SR-death-rate</u>: Defines the death rate for SR mosquitoes
       - 2.2.1.5. <u>RR-speed</u>: Defines the speed at which RR mosquitoes can travel
       - 2.2.1.6. <u>SR-speed</u>: Defines the speed at which SR mosquitoes can travel
       - 2.2.1.7. <u>ss</u>: The percent of the population with the SS genotype
       - 2.2.1.8. <u>sr</u>: The percent of the population with the SR genotype

2.2.1.9. rr: The percent of the population with the RR genotype
2.2.1.10. initial-recovered-people: Defines the initial number of recovered people (used only in the setup commands)
2.2.1.11. initial-infectious-people: Defines the initial number of infectious people (used only in the setup commands)
2.2.1.12. bites: Counts the times the mosquitoes bite people
2.2.1.13. normbites/hr: Counts the total bites per hour
2.2.1.14. infectious-bites: Counts the times the mosquitoes bite infectious people during the simulation
2.2.1.15. village1: Defines the village patches of the first village
2.2.1.16. village2: Defines the village patches of the second village
2.2.1.17. h-infectious-bites: Counts the bites on people from infectious mosquitoes during the simulation
2.2.1.18. infecting-bites/hr: Counts the bites on people from infectious mosquitoes per hour
2.2.1.19. mitigation-active: Defines the level of resistance mitigation which is active in the simulation
2.2.1.20. treated-patches: Counts the number of insecticide-treated patches at the beginning of the simulation
2.2.1.21. count-water-in-area: Counts the initial number of water patches in the source control area
2.2.1.22. Insecticide: Defines which insecticide is being used
2.2.1.23. death-time-deviation: Defines the value of the standard deviation on the kill time of the Late Life Acting Insecticide (LLA)
2.2.1.24. LLA-Coverage%: Defines the coverage rate over the village of the LLA
2.2.1.25. Death-Chance%: Defines the probability that a mosquito will die after contact with the Instant Acting Insecticide (IA)
2.2.1.26. IA-coverage%: Defines the coverage rate of the IA over the village
2.2.1.27. chance-of-exposure: Defines the chance that a mosquito will encounter the insecticide while on a treated patch for both insecticides
2.2.1.28. Cost: Defines the level of fitness cost associated with resistance for both insecticides
2.2.1.29. source-control: Boolean: defines if there is source control or not (i.e., source reduction of aquatic breeding sites)
2.2.1.30. source-control-size: Defines the size of the source control area
2.2.1.31. number-humans-village1: Defines the initial number of humans in the first village
2.2.1.32. number-humans-village2: Defines the initial number of humans in the second village

2.2.1.33. resistance-dominance: Defines the probability that an Individual SR mosquito will not be affected by either Insecticide

2.2.1.34. Number-of-water-patches: Defines the maximum (and initial) number of water patches

2.2.1.35. village-sense-radius: Defines the distance from the village at which the mosquitoes can detect the direction of the village

2.2.1.36. Set-Seed: Boolean: defines whether the seed will be preset for the simulation or not

2.2.1.37. initial-SR: Defines the maximum percent of SR mosquitoes that the system will allow during the burn in

2.2.1.38. Kill-Time-Mean: Defines the mean of the standard deviation which determines how long after exposure to the LLA insecticide the mosquito will die

2.2.1.39. recovered-time-mean-days: Defines the average time (in days) that a recovered human will spend before becoming susceptible

2.2.1.40. infectious-time-mean-days: Defines the average time (in days) that an infectious human will spend before becoming Recovered

2.2.1.41. External-Infection: If this is true, every three months the number of humans defined by new-infected-humans become infected from outside the simulated world (Boolean)

2.2.1.42. new-infected-humans: Defines how many new humans become infected if External-Infection is on

2.2.1.43. source-control-accuracy: Defines what percent of the water patches the source control feature is capable of removing upon application

2.2.2. Mosquitoes

2.2.2.1. gravid: Marks whether the mosquito has taken a blood meal

2.2.2.2. mAge: Records the age of the mosquito

2.2.2.3. time-after-bite: Records the time since the mosquito took a blood meal

2.2.2.4. GCs: Marks the number of Gonotrophic Cycles a mosquito has gone through

2.2.2.5. timeAfterGC1: Records the time since the first Gonotrophic Cycle

2.2.2.6. Infected-Time: Records the time since the mosquito picked up the plasmodium parasite (Compared against EIP)

2.2.2.7. mInfectious: Shows if the mosquito is infectious (Boolean)

2.2.2.8. mGenotype: Records the genotype of each mosquito (SS, SR, RR)

2.2.2.9. mGenotypeM: Records the genotype of the father (Male) of each mosquito in the simulation

2.2.2.10. I-Exposed: Marks whether the mosquito has been exposed to the LLA insecticide

2.2.2.11. I-Exposed-time: Records how long the mosquito has been infected by the insecticide

2.2.2.12. mDeath-time: Defines when the mosquito will die from exposure to the LLA Insecticide, random normal

2.2.3. The Three Types of Human Entities (Susceptible, Infectious, Recovered). All share the same variables.

2.2.3.1. time-since-bite: Records the time since an infecting bite (hours)

2.2.3.2. sick-time: Records how long the infectious person has been infectious.

2.2.3.3. v1: Marks the people in the first village

2.2.3.4. v2: Marks the people in the second village

2.2.3.5. out: Marks the people who will leave the village

2.2.3.6. r-time: Counts how long the person has been recovered (hours)

2.2.4. Brood Entities

2.2.4.1. bAge: Counts the age of the brood until it hatches; (Hours)

2.2.4.2. BLifespan: Defines the amount time before a brood "hatches" (hours) new adult mosquitoes

2.2.4.3. mGenotype: Marks the genotype of the mother of the brood (SS, SR, RR)

2.2.4.4. mGenotypeM: Marks the genotype of the father (Male) of the brood

2.2.5. Patches Entities

2.2.5.1. village1-scent: Marks the patches on which the mosquitoes can sense the first village

2.2.5.2. village2-scent: Marks the patches on which the mosquitoes can sense the second village

2.3. Scales

2.3.1. The space of the model is approximately $100m^2$ per patch. There are 40,401 patches arranged in a 201x201 grid making an approximately 4 $km^2$ map.

2.3.2. The space is toroidal, with horizontal and vertical edges wrapping.

2.3.3. The time in the model progresses in discrete timesteps each representing 1 hour. We used twelve-hour days to capture only the night hours that the mosquito is active (*Anopheles gambiae*). Most simulations were run for eight years or, counting the 1000 timestep burn-in period, 44800 hours.

3. Process overview and scheduling

3.1. Movement

3.1.1. The mosquitoes move forward randomly until they are within a certain distance from the village at which point, they move towards the village. Their path is still randomized but they are able to determine which of the three patches ahead of them (the one in front of them and the two diagonals) is closer to the village and they move that direction. Once they reach the village, they move until they take a blood meal or die. Once they have a blood meal they move randomly away from the village until they find a water patch where they lay a brood. They then resume their search for a blood meal. If there is a cost associated with resistance in the model, the mosquitoes with a resistant allele move more slowly, otherwise all the mosquitoes move at the same speed.

3.1.2. The humans move randomly within the village except for twenty or thirty humans which move in and out of either village linearly to simulate travel.

3.1.3. The broods and patches do not move

3.2. The state variables are updated after each timestep.

3.3. Time is modeled as discrete timesteps (1 tick = 1 hour)

4. Design concepts

4.1. Basic principles

4.1.1. For this model we relied heavily on the well documented principles of Insecticide Resistance (IR) emergence, the *Plasmodium falciparum* life cycle, the malaria disease in humans, and the aquatic life stages of the mosquito broods. The model uses these principles to give insight on the relationship between interventions and the emergence of IR and the transmission of malaria.

4.1.2. The malaria transmission cycle in the model begins with infectious humans who are bitten by mosquitoes who then become infected. After the extrinsic incubation period (EIP), the mosquito becomes infectious and any humans it bites afterward become infected. After time for the exo-erythrocytic cycle (liver-stage of parasite in human) the bitten humans become infectious – able to infect new mosquitoes.

4.2. Emergence

4.2.1. Insecticide Resistance is modeled as an emergent characteristic of the mosquito population influenced by the mosquito's genotype, the dominance of the resistance allele, the fitness cost of the resistance allele, the insecticide effectiveness, and other minor factors.

4.2.2. The transmission and endemicity of malaria in the humans are also emergent outcomes in the model, resulting from the interactions of mosquitoes, humans, insecticide interventions, and other factors.

4.3. Adaptation

4.3.1. The mosquito population has the ability to adapt to the insecticides by developing IR. This is done by using mendelian style genetics to pass resistant alleles from parent to offspring generations.

4.4. Objectives

4.4.1. The motivating design objective was to evaluate the potential of late-life-acting insecticides.

4.4.2. A second objective was to discover and conduct sensitivity analysis of critical simulation variables that might influence the success of the new insecticide type.

4.4.3. A third objective was a case study to determine if a novice programmer (an end-user-programmer) could develop a high quality and useful simulation using agent-based modeling using NetLogo

- 4.5. Learning
  - 4.5.1. N/A
- 4.6. Prediction
  - 4.6.1. N/A
- 4.7. Sensing
  - 4.7.1. The mosquitoes are able to sense the distance they are from the nearest village up to a certain number of patches away.
  - 4.7.2. The mosquitoes can also detect the number of broods in each water patch so that there are not too many broods in each water patch.
  - 4.7.3. The mosquitoes can sense a water patch a certain distance away from them when they are gravid.
- 4.8. Interaction
  - 4.8.1. The mosquitoes interact with the human agents by "biting" them when they are on the same patch. If the mosquito is not infectious this has no effect on the human. If the mosquito is infectious then the human becomes infected.
  - 4.8.2. The mosquitoes interact with the broods by laying (ovipositing) them in water patches and defining some of their variables. Each brood then hatches back into multiple mosquitoes after a time. These mosquitoes can have variations in their genotypes within the same brood.
  - 4.8.3. The broods and humans do not interact.
  - 4.8.4. Each entity interacts with the patches by evaluating their color. The humans interact with the "village" patches by determining whether they are in the "red" area to stay within the village. The mosquitoes interact with the "water" patches by determining whether they are blue so they can lay a brood of eggs there. The mosquitoes also interact with the patches to determine how far from the village they are. The broods interact with the patches to evaluate whether their blue "water" patch has turned black indicating that their water has dried up and they should die.
  - 4.8.5. The patches also interact with the broods by changing color depending on how many broods are on a "water" patch. If the water patch turns black (dries up), the brood dies.
- 4.9. Stochasticity
  - 4.9.1. There are many stochastic behaviors in the model such as the movement of the human and mosquito agents and the setting of variables whose values are not precise in nature.
- 4.10. Collectives
  - 4.10.1. The brood agents represent a collective of the aquatic stages of a whole brood of eggs laid by a single mosquito. Each brood agent "hatches" into multiple mosquitoes.
- 4.11. Observation
  - 4.11.1. The outcomes we measured to evaluate our simulations are the number of mosquitoes, the number of mosquitoes with each genotype, the infectious mosquitoes, the number of susceptible, infectious and recovered people, the number of infecting bites, and the total number of bites per hour.

5. Initialization

5.1. This model is initialized by the *Setup* command on the interface. This command creates each type of agent and initializes their variables. The patches (squares on a 201x201 grid) are created and colored according to what they represent. Two thirty patch diameter red circles are created to represent the villages located at (0,0) and (-60,60). A number of patches (defined by the *number-of-water-patches* slider) turn blue and are randomly distributed across the map to represent water. Each patch is also assigned a value determined by how far away from the village they are (used for mosquito movement). The following steps are used to populate the map.

5.1.1. The human agents are created in the center of each village and they are divided evenly between villages.

5.1.1.1. The number of humans is controlled by the *initial-number-humans* slider on the interface. Fourteen percent of the people are set to the recovered breed, forty-five percent are set to the infectious breed, and the rest are the susceptible breed. Twenty of the humans are given the *out* command which causes them to move in and out of the village in straight lines to simulate travel.

5.1.2. 1000 mosquitoes are created and spread across the map randomly.

5.1.2.1. All the mosquito variables are set to zero except for:

5.1.2.1.1. The genotype: 99.956% SS, .044% SR.

5.1.2.1.2. The gonotrophic cycle: 4 days (This value is randomized for each mosquito during the simulation)

5.1.2.1.3. 90 mosquitoes are gravid

5.1.2.1.4. 568 mosquitoes are infectious

5.1.3. 15 broods are created by the *setup* command to initialize the brood variables.

5.1.3.1. These broods are given an age of 1, the SS genotype and a lifespan of 144 ticks which is randomized later.

5.2. Burn in period

5.2.1. Ticks = 0: begin of burn in (negative starting time in days)

5.2.2. Ticks = 1000 (burn in period): begin of actual model runs (day 0)

5.2.3. The simulation runs for one year after the burn in before any interventions are applied to give a baseline comparison to each run.

6. Input Data

6.1. This model does not use data from outside sources.

7. Submodels

7.1. Humans:

7.1.1. Most humans remain at a fixed location in their home village, but a subset moves out of their home village and return home on a six-day cycle to reflect workers leaving to work outside the village.

7.1.2. All humans cycle through a SEIRS disease model (Susceptible, Exposed, Infectious, Recovered, Susceptible). Susceptible humans, when bitten by an infectious mosquito, have a chance of becoming infected, and then progress through the infectious, recovered (immune), and back to the susceptible states. In the program susceptible humans are instances of the human breed. We modeled the infected people as still part of the susceptible breed until they become infectious and move to the infectious breed. The recovered humans are instances of the recovered breed.

7.2. Broods: Broods are composites of all mosquito eggs (and subsequent aquatic stages) associated with one female egg-laying event (oviposition).

7.3. Mosquitoes:

7.3.1. Adult and juvenile mosquitoes are instances of the mosquito breed. All simulated mosquitoes are female. They are associated with a specific brood, inheriting their genotype from that brood at the time of birth. The female mosquitoes spend their adult life in a gonotrophic cycle of (1) blood meal seeking, (2) biting a human, and (3) oviposition, continuing with randomized movement until death. Death can occur from hourly mortality, or because of the killing effect of an insecticide, either instant-acting (IA) or late-life-acting (LLA). When biting an infectious human, the mosquito has a chance of becoming infected, followed by randomized extrinsic incubation period of 8-12 days before she becomes infectious. After the incubation period, the humans it bites have a chance of becoming infected.
