## Supplementary material for "Case Study: End User Development of an Agent-based Model of Malaria Transmission to Support the Design of Late-Life-Acting Insecticides for the Control of Malaria Transmission and Delay of the Evolution of Insecticide Resistance": S3 - Table Model Variables

### Supporting File S3 Table Model Variables

| Variables | Descriptions | Values/Data Types |
|---|---|---|

#### Global Variables

| | | |
|---|---|---|
| Days | Counts the days the simulation has been running | floating point |
| Years | Counts the years the simulation has been running | floating point |
| SS | The percent of the mosquito population with the SS genotype | floating point |
| SR | The percent of the mosquito population with the SR genotype | floating point |
| RR | The percent of the mosquito population with the RR genotype | floating point |
| RR-death-rate | Defines the death rate for RR mosquitoes | 0, 5, 10 |
| SR-death-rate | Defines the death rate for SR mosquitoes | 0, 2, 5 |
| RR-speed | Defines the speed at which RR mosquitoes can travel | 1, 0.8, 0.7 |
| SR-speed | Defines the speed at which SR mosquitoes can travel | 1, 0.9, 0.8 |
| Initial-recovered-humans | Defines the initial number of recovered humans (used only in the setup commands) | .14 * total humans |
| Initial-infectious-humans | Defines the initial number of infectious humans (used only in the setup commands) | .45 * total humans |
| bites | Counts all the times the mosquitoes bite people | Integer |
| Bites/hr | Counts the total bites per hour | Integer |
| Infectious-bites | Counts the times the mosquitoes bite infectious people during the simulation | Integer |
| Village1 | Defines the village patches of the first village | True/False |
| Village2 | Defines the village patches of the second village | True/False |
| h-infectious-bites | Counts the bites on people from infectious mosquitoes during the simulation | Integer |

| | | |
|---|---|---|
| Infecting-bites/hr | Counts the bites on infectious people which infect the mosquito per hour | Integer |
| insecticide | Defines which insecticide is being used | Instant-Acting, Late-Acting, None |
| death-time-deviation | Defines the value of the standard deviation on the kill time of the Late Life Acting (LLA) Insecticide | 0, 1, 2, 3, 4, 5, 6, 7, 8, 9, 10 |
| LLA-Coverage% | Defines the coverage rate over the village of the LLA insecticide | 5, 10, 15, 20, 25, 30, 40, 50, 60, 70, 75, 80, 90, 100 |
| Death-Chance% | Defines the probability that a mosquito will die after contact with the Instant Acting (IA) Insecticide | 10, 20, 25, 30, 40, 50, 60, 70, 75, 80, 85, 90, 100 |
| IA-coverage% | Defines the coverage rate of the IA insecticide over the village | 5, 10, 15, 20, 25, 30, 40, 50, 60, 70, 75, 80, 90, 100 |
| chance-of-exposure | Defines the chance that a mosquito will encounter the insecticide while on a treated patch for both insecticides | 5, 10, 15, 20, 25, 30, 40, 50, 60, 70, 75, 80, 90, 95, 100 |
| cost | Defines the level of fitness cost associated with resistance for both insecticides | High Cost, Low Cost, No Cost |
| source-control | Defines if the source control feature has been activated for the simulation. | True/False |
| source-control-size | Defines the size of the source control area | 5, 10, 15, 20, 25, 30, 35, 40, 45, 50 |
| number-humans-village1 | Defines the number of people in the first village | 0-600 |
| number-humans-village2 | Defines the number of people in the second village | 0-600 |
| resistance-dominance | Defines the probability that an Individual SR mosquito will not be affected by either Insecticide | 0-100 |
| number-of-water-patches | Defines the maximum (and initial) number of water patches in the simulation | 50-1800 |
| village-sense-radius | Defines the distance from the village at which the mosquitoes can detect the direction of the village | 0-100 |
| Set-Seed | Defines whether the seed will be preset for the simulation or not | True/False |
| Initial-SR | Defines baseline percent of the mosquito population that is maintained as SR in the simulation | 0, 0.25, 0.5, 0.75, 1, 1.25, 1.5, 1.75, 2, 2.25, 2.5, 2.75, 3 |
| Kill-Time-Mean | Defines the mean of the standard deviation which determines how long after exposure | 6-14 |

| | | |
|---|---|---|
| | to the LLA insecticide the mosquito will die (days) | |
| recovered-time-mean-days | Defines the average time (in days) that a recovered person will spend before becoming susceptible | 0-300 |
| infectious-time-mean-days | Defines the average time (in days) that an infectious person will spend before becoming Recovered | 0-300 |
| External-Infection | If this is true, every three months the number of people defined by new-infected-humans become infected from outside the simulated world | True/False |
| new-infected-humans | Defines how many new humans become infected at each interval if External-Infection is on | 0-20 |
| source-control-accuracy | Defines what percent of the water patches the source control feature is capable of removing upon application | 0-100 |

#### Mosquito Variables

| | | |
|---|---|---|
| gravid | Marks whether the mosquito has taken a blood meal | True/False |
| mAge | Records the age of the mosquito | Integer |
| time-after-bite | Records the time since the mosquito took a blood meal | Integer |
| max-time-after-bite | Defines the time between when a mosquito takes a blood meal and when she is ready to lay her eggs | Random: max-48 min-24 |
| GCs | Marks the number of Gonotrophic Cycles a mosquito has gone through | Integer |
| Infected-Time | Records the time (hours) since the mosquito picked up the plasmodium parasite (Compared against EIP) | Integer |
| mInfectious | Shows that the mosquito is infectious | True/False |
| EIP | Defines the extrinsic incubation period for the mosquitoes | Random: Max-144, Min-96 |
| mGenotype | Records the genotype of each mosquito | SS, SR, RR |
| mGenotypeM | Records the genotype of the father (Male) of each mosquito in the simulation | SS, SR, RR |

| | | |
|---|---|---|
| I-Exposed | Marks whether the mosquito has been exposed to the LLA insecticide | True/False |
| I-Exposed-time | Records how long the mosquito has been infected by the insecticide | Integer |
| mDeath-time | Defines when the mosquito will die from exposure to the LLA Insecticide, random normal | random normal mean: *Kill-Time-Mean* Standard Deviation: *death-time-deviation* |

#### Humans Variables (same for all three types of human agents)

| | | |
|---|---|---|
| time-since-bite | Records the time since an infecting bite (hours) | Integer |
| sick-time | Records how long the infectious person has been infectious (hours) | Integer |
| recovery | Defines how long before infectious people become recovered (hours) | Random-Normal mean: *recovered-time-mean-days* Standard Deviation: 180 |
| v1 | Marks the Humans in the first village | True/False |
| v2 | Marks the Humans in the second village | True/False |
| out | Marks the humans who will leave the village | True/False |
| r-time | Counts how long the person has been recovered (hours) | Integer |
| max-rtime | Defines how long the recovered person will be recovered before being susceptible | Random-Normal Mean: *recovered-time-mean-days* Standard Deviation: 120 |

#### Brood Variables

| | | |
|---|---|---|
| bAge | Counts the age of the brood until it hatches; compared with *bLifespan* (hours) | Integer |
| bLifespan | Defines the amount time before a brood hatch (hours) | random-normal max-144 min-98 |
| mGenotype | Marks the genotype of the mother of the brood | SS, SR, RR |
| mGenotypeM | Marks the genotype of the father (Male) of the brood | SS, SR, RR |

#### Patch Variables

| | | |
|---|---|---|
| village1-scent & village2-scent | Marks the patches on which the mosquitoes can sense the closest village. Used to direct the mosquitoes towards the village | Gradient |
