## Supplementary material for "Case Study: End User Development of an Agent-based Model of Malaria Transmission to Support the Design of Late-Life-Acting Insecticides for the Control of Malaria Transmission and Delay of the Evolution of Insecticide Resistance": S4 - Supplemental Figures Output Plots

### Supporting Information: S4 Figures – Simulation Parameter Settings and Output Plots

The figures in this supporting information file present various simulation outputs generated by the NetLogo program, EMMIT (Extendable Model for Malaria Intervention Trials), that is provided in the Supporting Information file – "S1 Program". Our figures help demonstrate the validity of the simulation and served as detailed examples of how the simulation could be used and modified by other researchers. We present findings from three malaria control scenarios, no interventions (control), Instant Acting (IA) insecticides, and Late Life Acting (LLA) insecticides. Two additional investigations are presented as examples on how the simulation can be expanded to evaluate additional malaria control efforts: larval source control, and an evaluation of policy to mitigate the impact of resistance. Each figure, or group of figures, below is accompanied by a screen shot of the NetLogo simulation experiment settings (the BehaviorSpace settings) used to generate the plots presented in the figures. BehaviorSpace is a NetLogo feature that allows users to "batch" test combinations of several variables at a time. To utilize it, the user inputs a value or values for each of the parameters and then NetLogo runs each combination of values and reports the results in a CSV file, which are then plotted as the figures below using a graphing program (Excel in our case). The figures presented below were generated using this method.

It is important to know some of the assumptions we have implemented in the production of these figures. First, we have assumed that there is a constant population of heterozygous genotype mosquitoes in the population which account for up to 2% of the mosquito population at the beginning of each simulation. This represents the natural occurrence of resistance caused by genetic mutations in the wild. The amount of these mosquitoes to begin the simulation is controlled by the "initial-SR" variable. Significant changes in this variable (on the order of multiple percentages) can cause the development of resistance to happen more or less rapidly. Second, we implemented a feature that introduces new infected humans to the simulation (by infecting one of the susceptible humans) at a controlled rate, normally of one new infected person per three months. The number of infected humans introduced is controlled by the "new-infected-humans" variable. We did this to reproduce the way in which sometimes infected humans travel and introduce infections in other areas. Without this feature, an effective insecticide can create a state where there are no infected humans for the rest of the simulation, which is not always accurate and not as useful for evaluating the insecticide with the simulation. Third, we ran each simulation with a one-year control period before any of the interventions were applied. This effect can be seen in the plots of the total mosquito population which frequently decreases after twelve months due to the application of the insecticide. Fourth, we created a setting to determine the mean time that humans would spend infected or recovered.

These numbers were then used to set the mean of a normally distributed random number generator to determine how long they would remain in either state. We ran our experiments with thirty days as the mean for both the time infected and the time recovered but this can be changed to fit a more specific region's data. Fifth, we implemented an oscillation in the number of water patches in our simulation to represent the natural seasonality in the availability of water in nature. This results in the fluctuation in the number of total mosquitoes dependent on the amount of aquatic breeding sites which are available for ovipositioning, and larva and pupae development. Finally, most of the timings presented here are based on assumptions about the durations of the mosquitoes' gonotrophic cycle, the assumed hourly death rate of the mosquitoes from natural causes, the hourly distance traveled by each mosquito when searching for a blood meal or a breeding site, etc. These are calibration parameters that would need to be "tuned" for a specific malaria transmission setting.

### Figures

#### Verification and Validation – Human SIR populations

Many verification, and validation (V&V) trials of the malaria transmission program were run during program development. In most cases, face-validity was the primary test of correctness of the program. While the samples listed in this section are explicitly used for verification and validation purposes, all subsequent results implicitly contribute to the confidence in the correctness of the program. The BehaviorSpace settings for this experiment are shown in S4-Fig. 1. The corresponding chart (see S4-Fig. 2). shows the results for the susceptible, exposed, infected, recovered and susceptible (SEIRS) states in the human population in a simulation with no interventions applied.

Notes:

- Both the “insecticide” and “source-control” variables are inactivated so that there are no interventions in the simulation.
- The “infected-time” and “recovered-time” variables were set to 30 days to accelerate the run time. Changing these parameters results in a different equilibrium in the SEIRS states.

Observations:

- Note how it displays the endemic steady-state of malaria in the model. This is the expected epidemiological condition for the tested malaria scenarios. Despite random fluctuations, there is a persistent population of infected humans in the simulation. This plot is used later to compare the effects of insecticide interventions, to how the system behaves without such interventions. A couple variable settings are important to note.

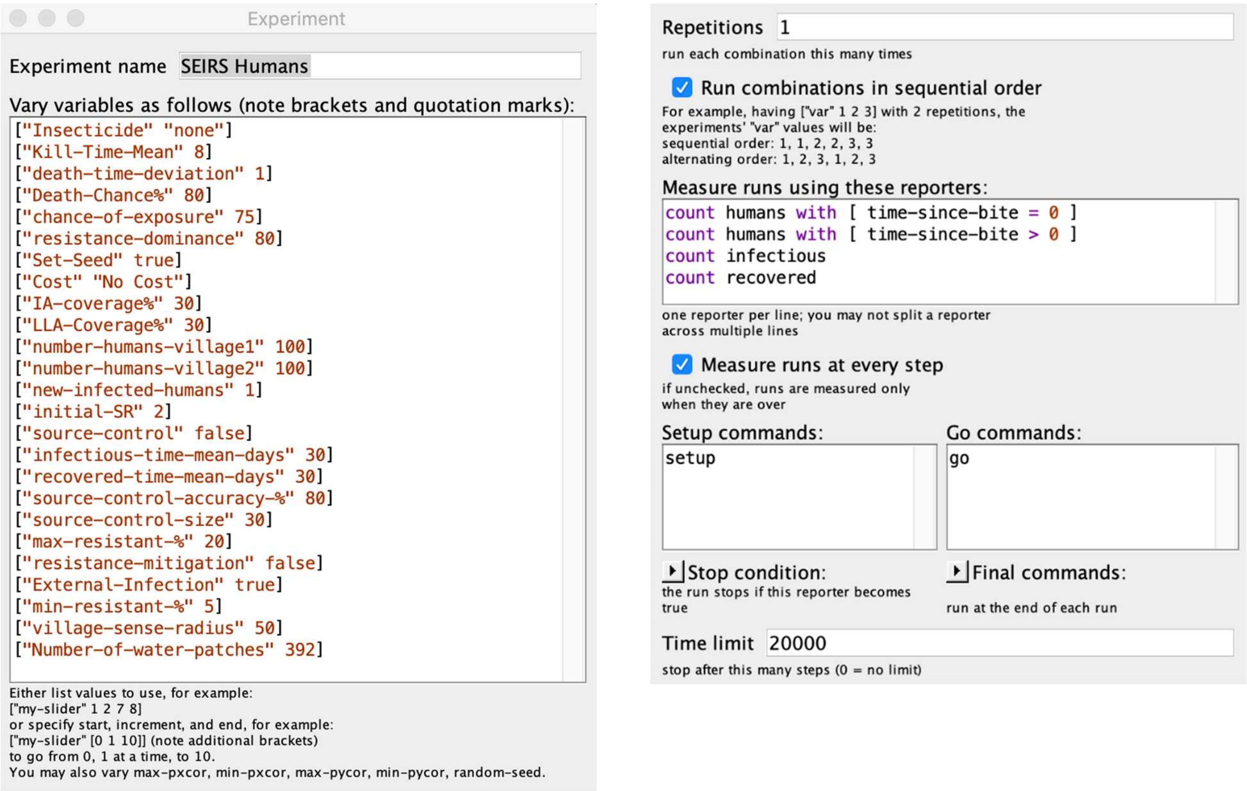

**S4-Fig. 1. Simulation experiment settings: V&V on human and mosquito populations.** Face validity checks on human and mosquito populations while model is running with no mosquito control interventions.

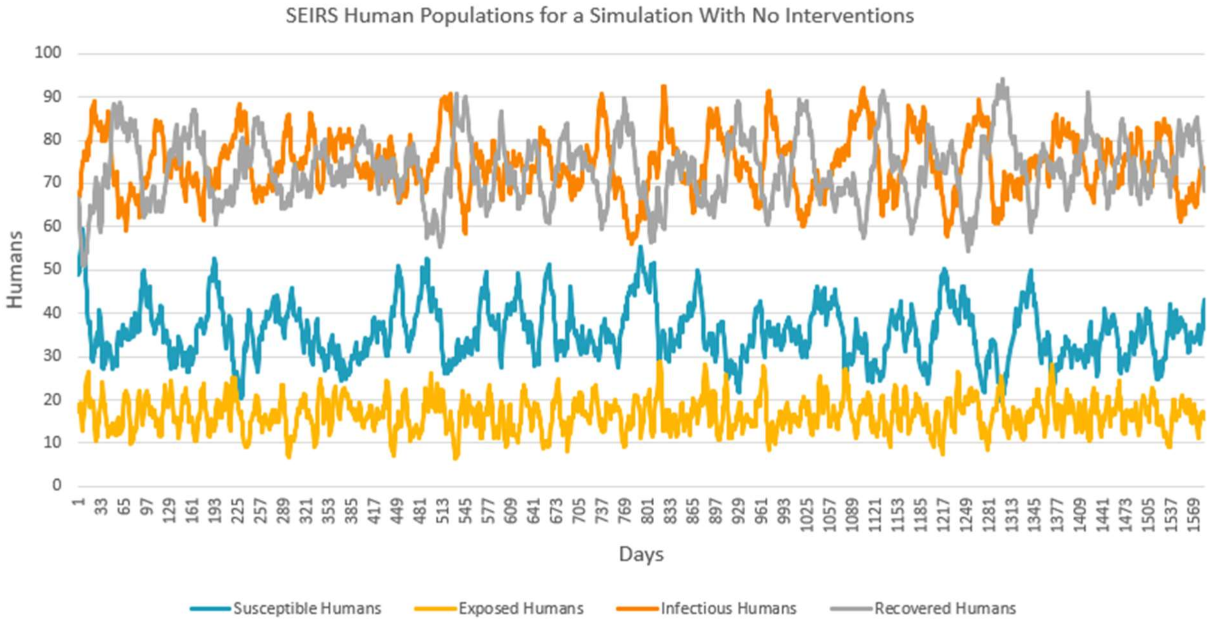

**S4-Fig. 2. Number of humans in each of the SEIRS states.**
Note how it displays the endemic steady-state of malaria in the model. Despite random fluctuations, there is a persistent population of infected humans in the simulation. The proportion of humans in each SEIRS state can be adjusted in the model's code

### Verification and Validation – Mosquito populations

This plot shows an example of the mosquito populations with no interventions applied. The lack or SR or RR genotype mosquitoes shows that the development of resistant mosquitoes is not a result of something other than insecticide application in our model and only remain at the baseline levels which we use for testing and comparison with the insecticide interventions. For results see S4-Fig. 3.

Observations:

- The periodic variation in the total mosquito population is due to seasonality caused the modeled rainfall seasons.
- Observe in S4-Fig. 3, since no insecticide is in use, there is no emergence of insecticide resistance as indicated by only the baseline number of mosquitoes with an R allele (SR or RR).

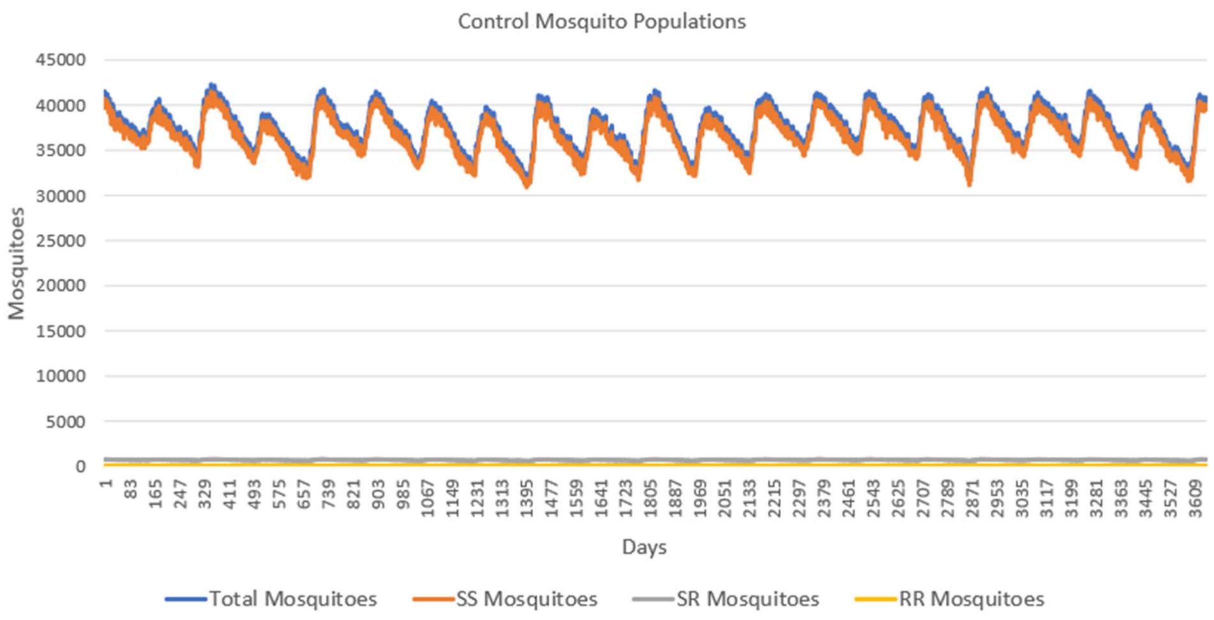

**S4-Fig. 3. Mosquito genotype populations with no insecticide interventions.**
Note: no emergence of insecticide resistance (no mosquitoes with SR or RR genotypes), since for this experiment, there was no use of an insecticide intervention.

#### Verification and Validation – Mosquito age distribution

While there is evidence for age-dependent mortality for the female mosquitoes, we made the simplifying assumption that mortality remains constant with age. This experiment shows the proportion of the population in each daily age group. Each box represents the proportion of the mosquito population within each day of survival (ex: the first box represents the mosquitoes within one day old). The data is an average of the population age structure over many days in a control test. This data confirms our intended age structure, i.e., that associated with constant mortality. In addition to the data, a fitted curve is plotted to show the similarity between the simulation's population data and an exponential decline. The BehaviorSpace settings for this experiment are shown in figure S4-Fig. 4 and the mosquito age distribution is shown in figure S4-Fig. 5. This experiment provides support for the verification and validation of the assumed constant mortality of the mosquitoes in the simulation.

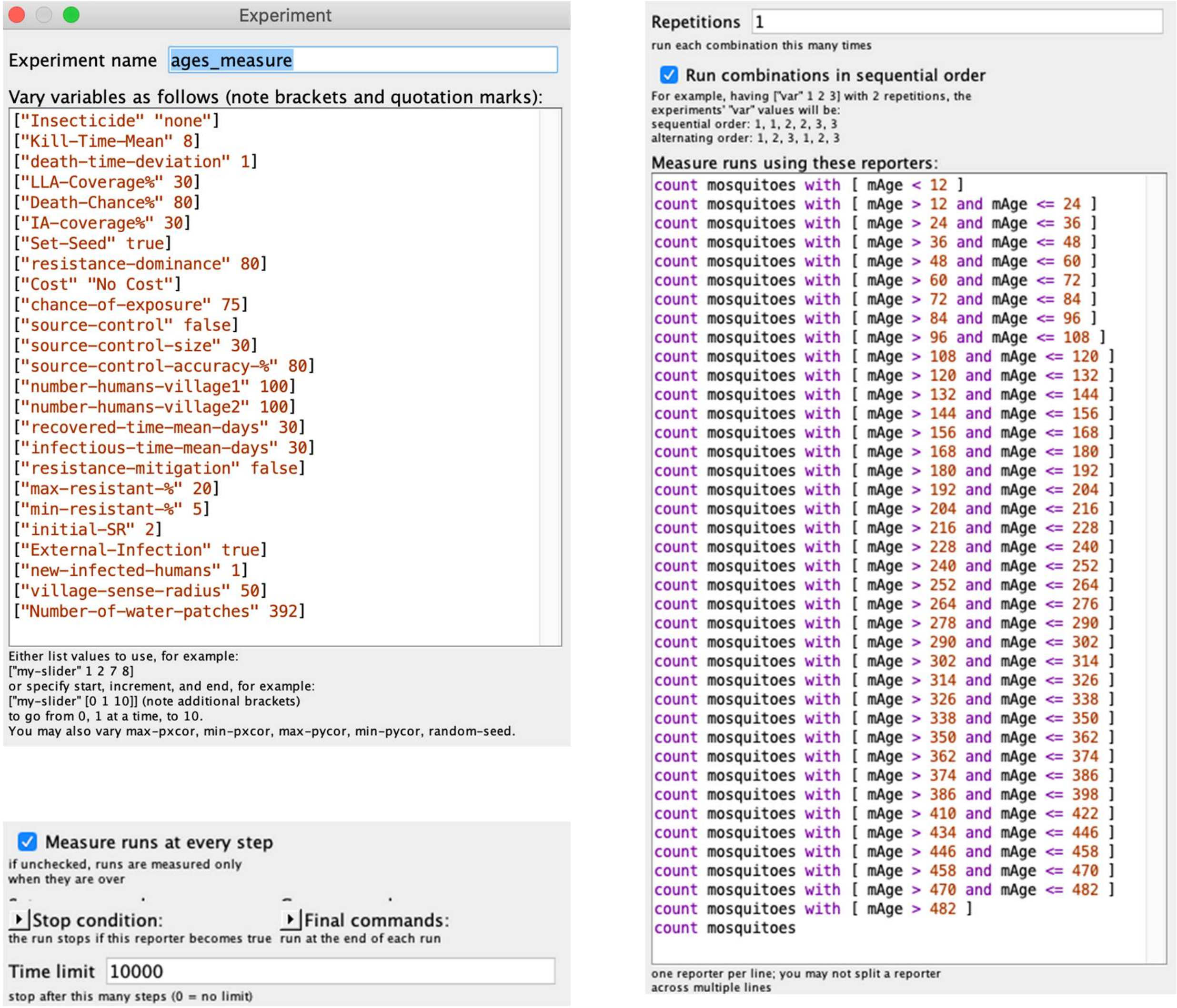

**S4-Fig. 4. Experimental Settings – Mosquito Population Age Distribution.**

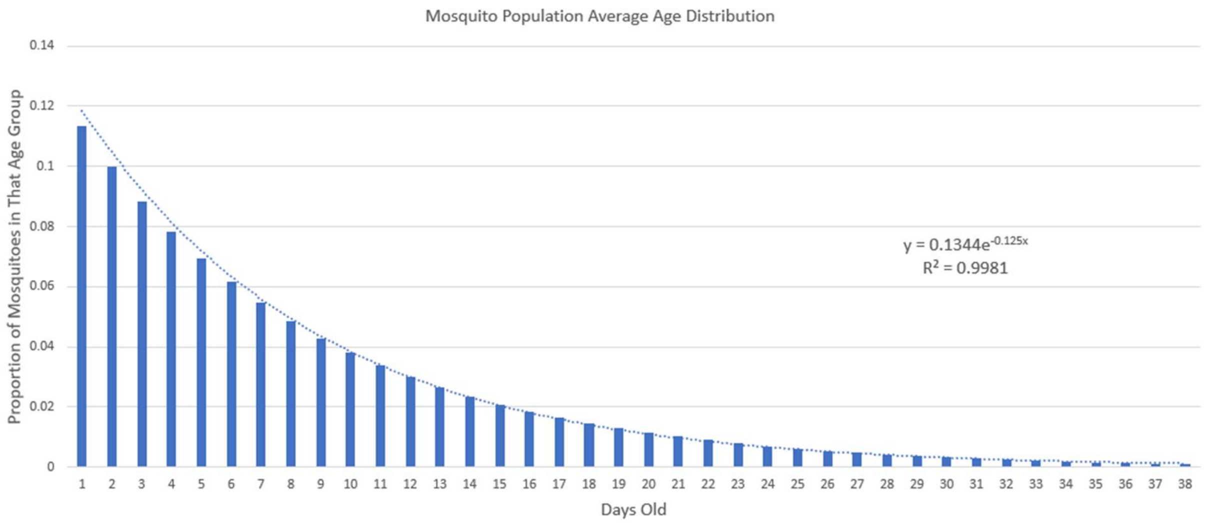

**S4-Fig. 5. Mosquito Population Average Age Distribution.**

#### Sensitivity Analysis to the Initial Random Generator Seed

To account for: 1) the uncertainty around the values of many simulation parameters, and 2) the natural randomness in malaria transmission, our simulation makes extensive use of random distributions, including uniform-discrete, uniform-continuous, and normal distributions. The (pseudo-) random number generator used by NetLogo is the Mersenne Twister and uses a seed to start the generation of a sequence of (pseudo-) random numbers. Often simulation results are dependant on the starting seed and one run of the simulation can provide misleading results if examined alone. Should this be the case, repeated iterations of the simulation are needed to obtain the range and distribution of possible outcomes. This experiment investigates the sensitivity of the starting seed for the random number generator. Ten replications of the simulation were run (each with a different starting seed) for each of three scenarios: 1) no insecticide intervention, 2) IA insecticide intervention, and 3) LLA insecticide intervention. A total of 30 runs displaying the mosquito populations are plotted, color coded by scenario (10 runs for each scenario). The BehaviorSpace experimental setup is displayed in S4-Fig. 6. The mean of the ten runs for each scenario is also plotted in bold with the same color, showing the variability around the mean – and hence sensitivity – to choice of seed. See S4-Fig. 7.

Observations:

- Observe that only the IA insecticide intervention scenario displays insecticide resistance (as can be seen by the return of the mosquito population to close to the original level as insecticide resistance spreads throughout the mosquito population). The onset of resistance (indicated by increasing mosquito population) is sensitive to random seed, starting as early as month 38 and as late as month 80.
- The other two scenarios are relatively uneffected by the choice of random seed. Thus for robust comparisons of experimental results, replications may be needed. To keep the

figures displayed later in this document from being too cluttered we usually display one run (i.e., one random number generator seed).

- Proper analysis and interpretation of the results will often require multiple experimental runs with different seeds. Additional sensitivity tests of the choice of initial random number generator seed are presented in figures S4-Fig. 29, S4-Fig. 40 and S4-Fig. 43.

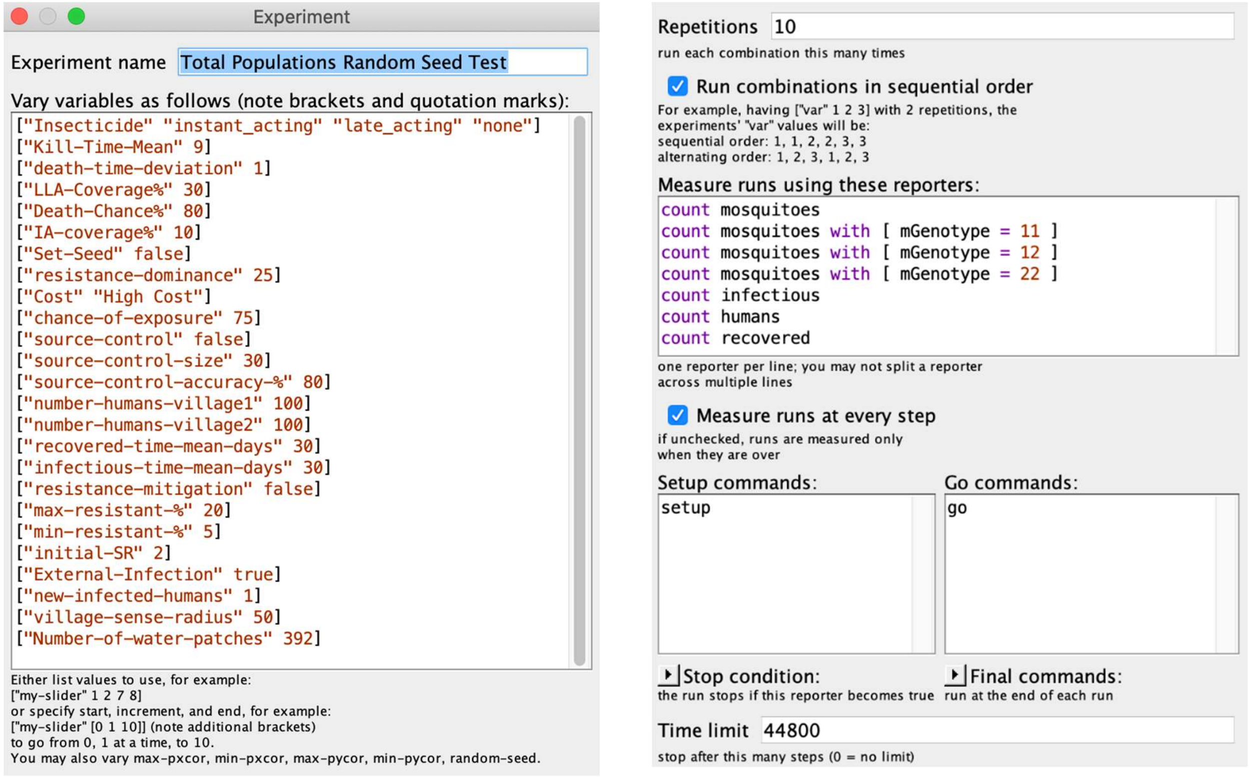

**S4-Fig. 6. Experiment settings – sensitivity analysis on random number seed.**
Ten replications for each of three scenarios: no interventions, IA insecticide, and LLA insecticide for a total of 30 runs.

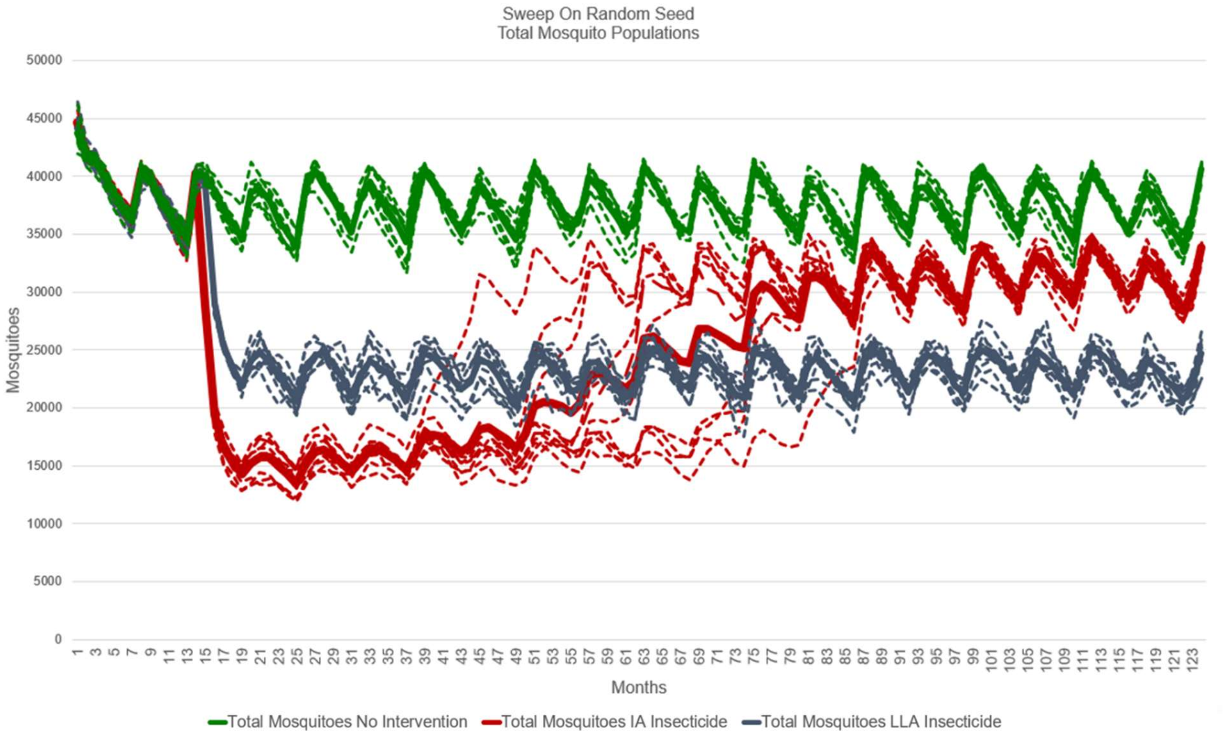

**S4-Fig. 7. Sensitivity analysis on the random number generator seed.**
Note: The no intervention and the LLA insecticide intervention scenarios are not significantly affected by the random number generator seed, while the IA insecticide intervention scenario is affected by the random number generator seed.

#### Sensitivity Analysis and Verification & Validation – Evaluating the Effects of Resistance Dominance and Resistance Cost on IA Insecticide

The following experiment evaluates two levels of "chance-of-exposure" to the IA insecticide, (50% and 75%, but only 75% is plotted), three levels of resistance dominance (0, 25, and 50), three levels of resistance cost (none, low and high). The BehaviorSpace experimental setup is displayed in S4-Fig. 8. The resulting plots are displayed in figures S4-Fig. 9 though S4-Fig. 12.

Notes:

- In S4-Fig. 9 the results of a sweep on three levels of resistance cost (none, low, high) are displayed for 75% chance-of-exposure and zero resistance dominance. With zero resistance dominance, there is no advantage given to the SR genotype and no resistance population emerges as would be expected, thus supporting face validity of the simulation. Likewise, resistance cost is not a factor, as no resistance population emerges.
- In S4-Fig. 10 the results of a sweep on three levels of resistance cost (none, low, high) are displayed for 75% chance-of-exposure and 25% resistance dominance. Because of the

25% resistance dominance setting, we see the first emergence of the SR mosquitoes around month 70 for the case of no resistance cost. In the other plots for the cases of low or high resistance cost, no SR mosquitoes emerge. These results appear to be trending as expected, thus supporting face validity.

- In S4-Fig. 11 the results of a sweep on three levels of resistance cost (none, low, high) are displayed for 75% chance-of-exposure and 50% resistance dominance. Because of the 50% resistance dominance setting, we see the emergence of the SR mosquitoes much sooner, around month 20 and the subsequent emergence and domination of the RR mosquitoes (for the case of no resistance cost). In the other plots: i) for the case of low resistance cost, SR mosquitoes emerge later (around month 26) and ii) for the case of high resistance cost, no SR mosquitoes emerge. These results appear to be trending as expected, thus supporting face validity of the simulation.
- Finally, in S4-Fig. 12, the results of a sweep on three levels of resistance cost (none, low, high) are displayed for 75% chance-of-exposure and 75% resistance dominance. Because of the high resistance dominance (75%) we observe the very early emergence of the SR mosquitoes (around month 16) for the case of no resistance cost. Likewise we observe the emergence of SR mosquitoes, but somewhat later caused by the increasing cost of resistance (low and high cost). These results appear to be trending as expected, thus supporting face validity of the simulation.

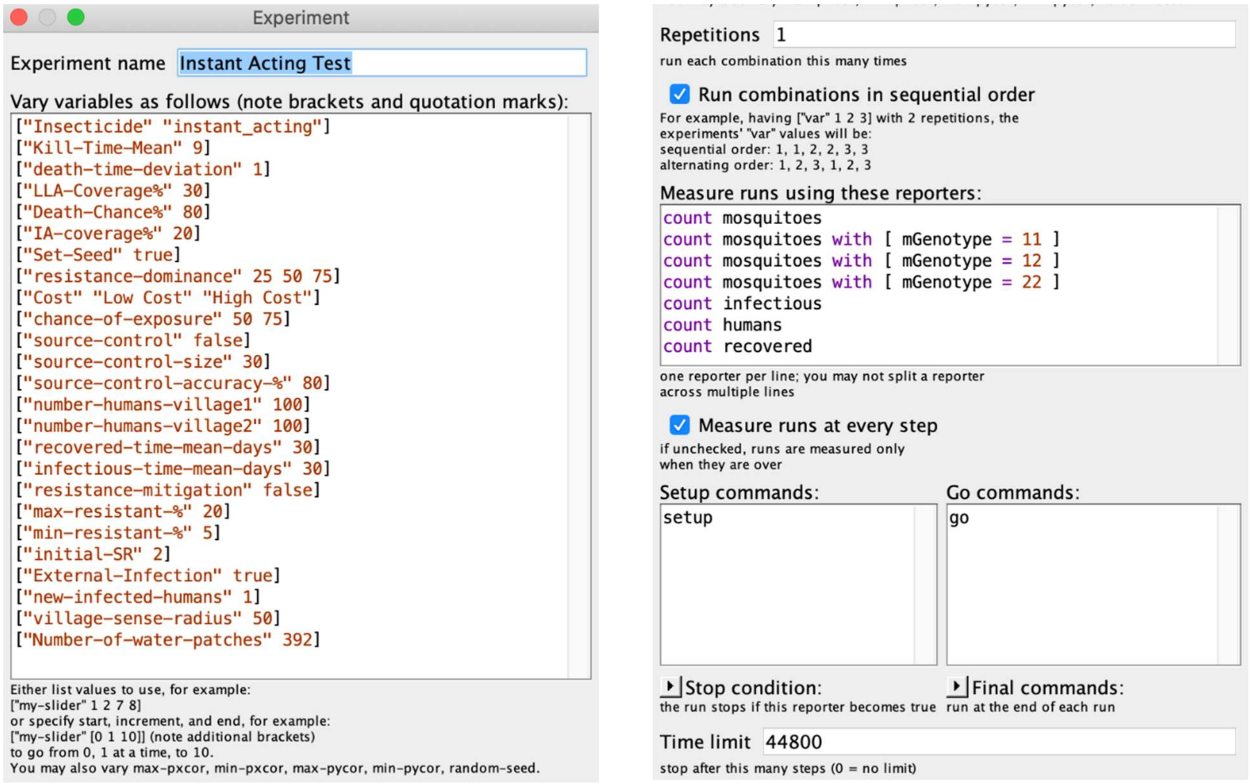

**S4-Fig. 8. Sensitivity and V&V sweeps on IA insecticide parameters.**

Evaluation of the effects of different levels of resistance dominance and resistance cost.

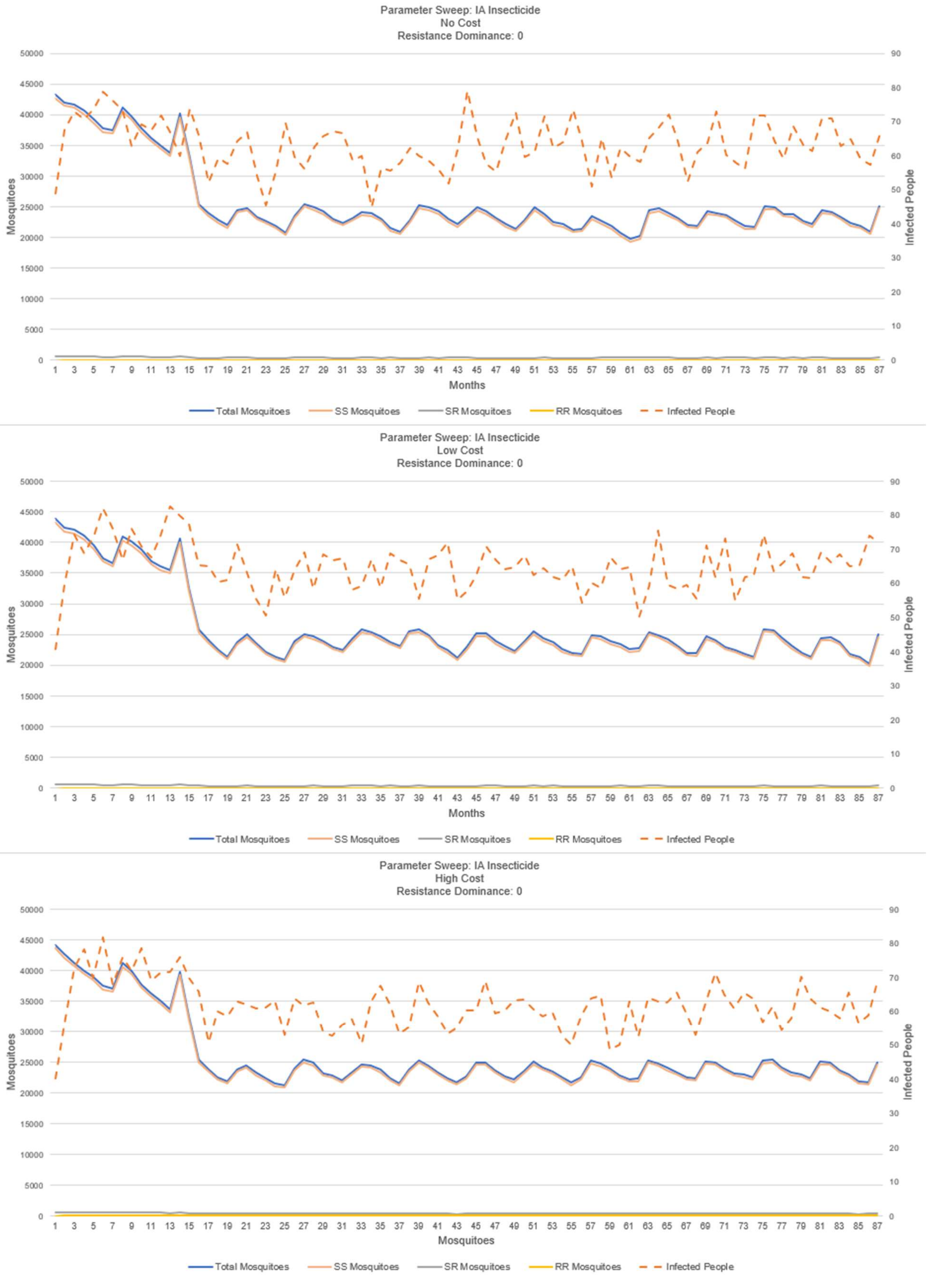

**S4-Fig. 9. IA insecticide: sweep on resistance cost – 0% resistance dominance.**

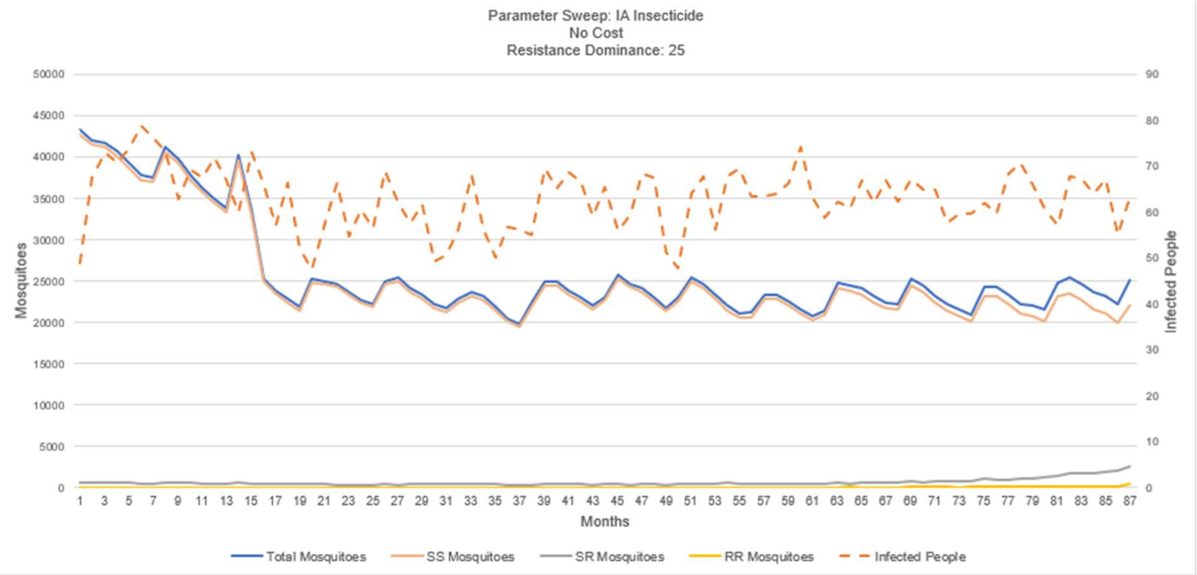

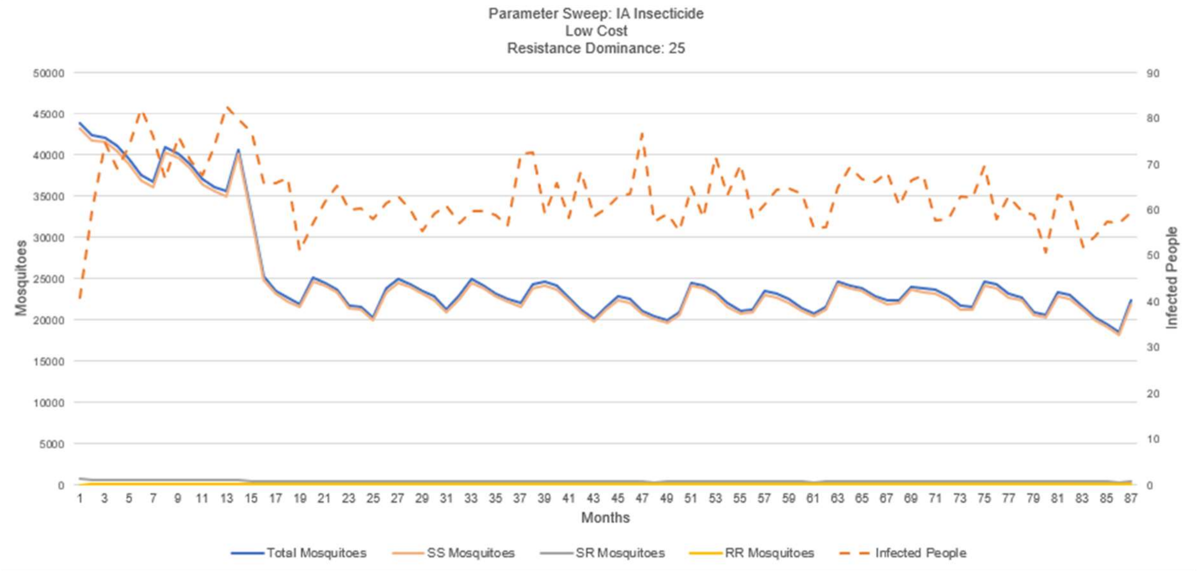

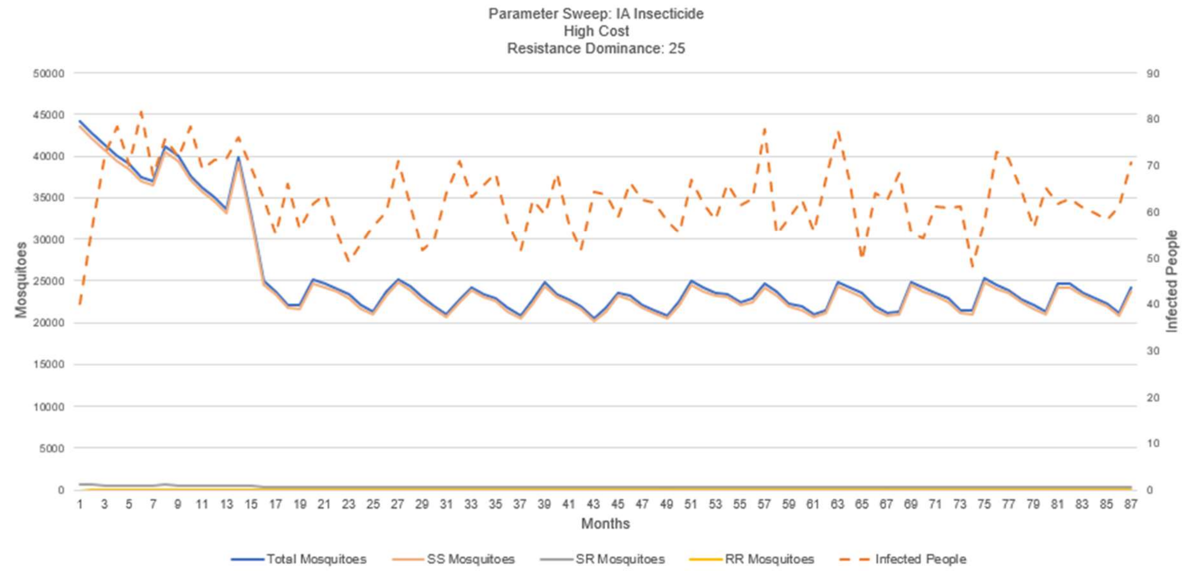

**S4-Fig. 10. IA insecticide: sweep on resistance cost – 25% resistance dominance.**

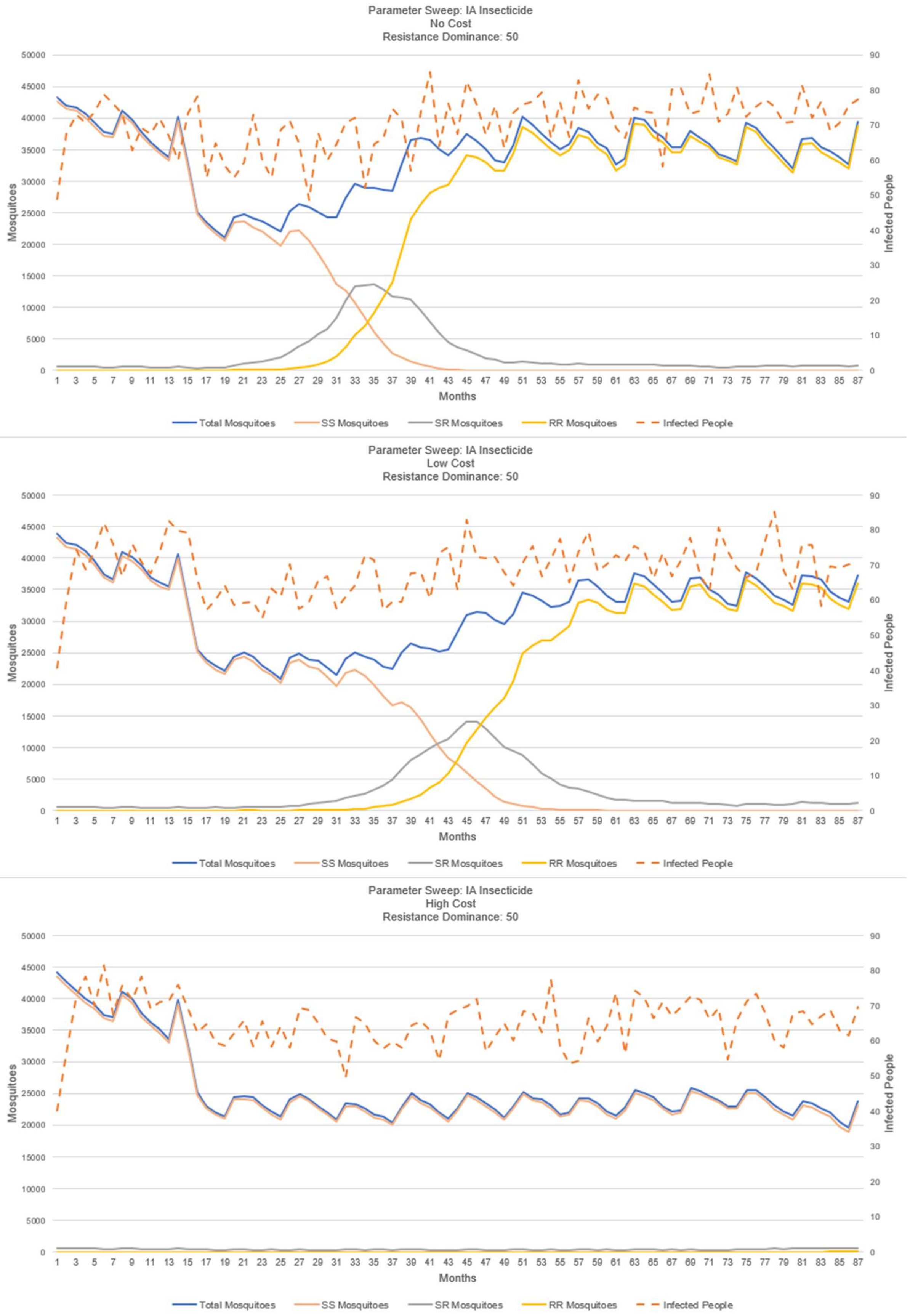

**S4-Fig. 11. IA insecticide: sweep on resistance cost – 50% resistance dominance.**

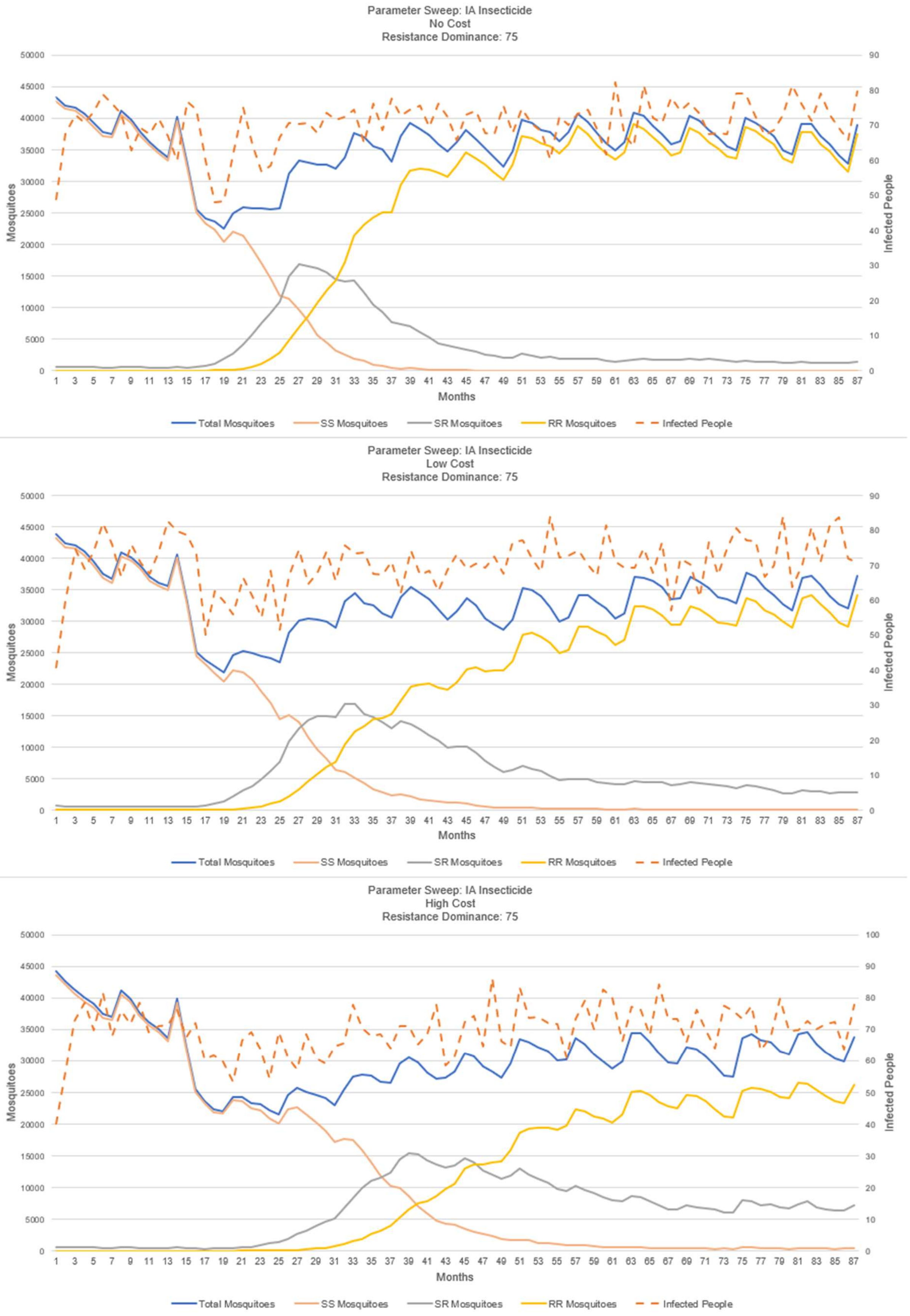

**S4-Fig. 12. IA insecticide: sweep on resistance cost – 75% resistance dominance.**

### Sensitivity Analysis and Verification & Validation Runs – Sweep on Chance-of-Exposure and Standard Deviation

These figures display the results from a parameter sweep on the variables "Chance-of-Exposure" and "Standard Deviation" (SD). They are organized by the value of the "Chance of Exposure". The BehaviorSpace experimental settings are displayed in S4-Fig. 13. The results are presented in figures S4-Fig. 14 through S4-Fig. 17.

Notes:

- The mean-kill-time for this test was 9 days and there was no cost of resistance.
- Coverage was set at 30%
- In each figure we sweep on SD: zero, one and two days.
- Each figure has chance-of-exposure set to one value: 25%, 50%, 75%, and 100%
- Importation of external cases is simulated, resulting in small random numbers of human infections over the time period of the simulation.

Observations:

- One key thing to notice from these results is that as the chance that the mosquito is exposed to the insecticide goes up, the malaria transmission goes down.
- Another noticeable result is that as the standard deviation value increases the time to resistance decreases and the LLA insecticide's ability to inhibit the spread of malaria decreases.
- One variable that interacts with and has similar effects to the "Chance of Exposure" variable is "LLA-Coverage%" which controls how much of the villages are insecticide treated patches. In this test, only 30% of the villages were covered with the insecticide. As you raise the coverage percent, it effectively increases the chance of exposure as the mosquitoes encounter a treated patch more frequently.
- Parameter changes that could result in improved performance of the insecticide (in the simulation) at lower chances of exposure include: 1) increasing the cost of resistance, "Cost"; 2) raising the insecticide coverage percentage, "LLA-Coverage%"; 3) lowering the dominance of the resistant allele, "resistance-dominance"; and 4) raising the accuracy of the insecticide around an ideal kill time "death-time-deviation" and "Kill-Time-Mean" respectively.

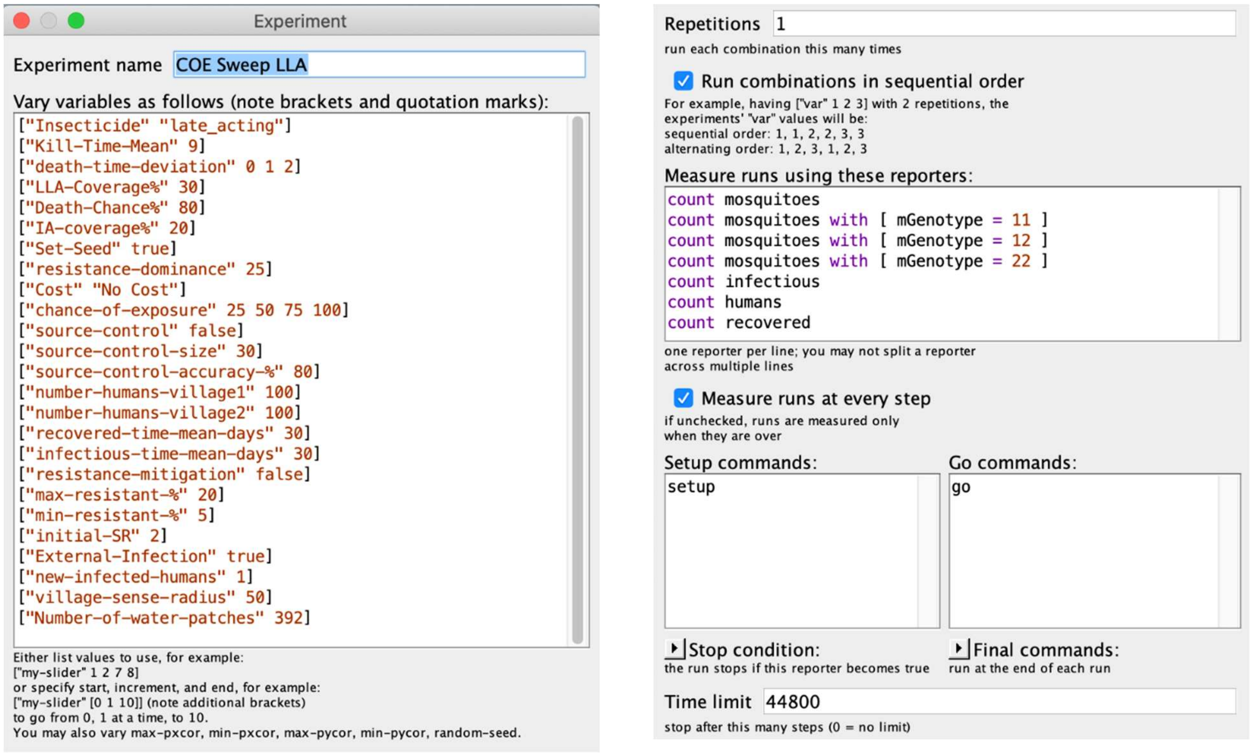

**S4-Fig. 13. Experiment settings for SD and Chance of Exposure Sweeps.**

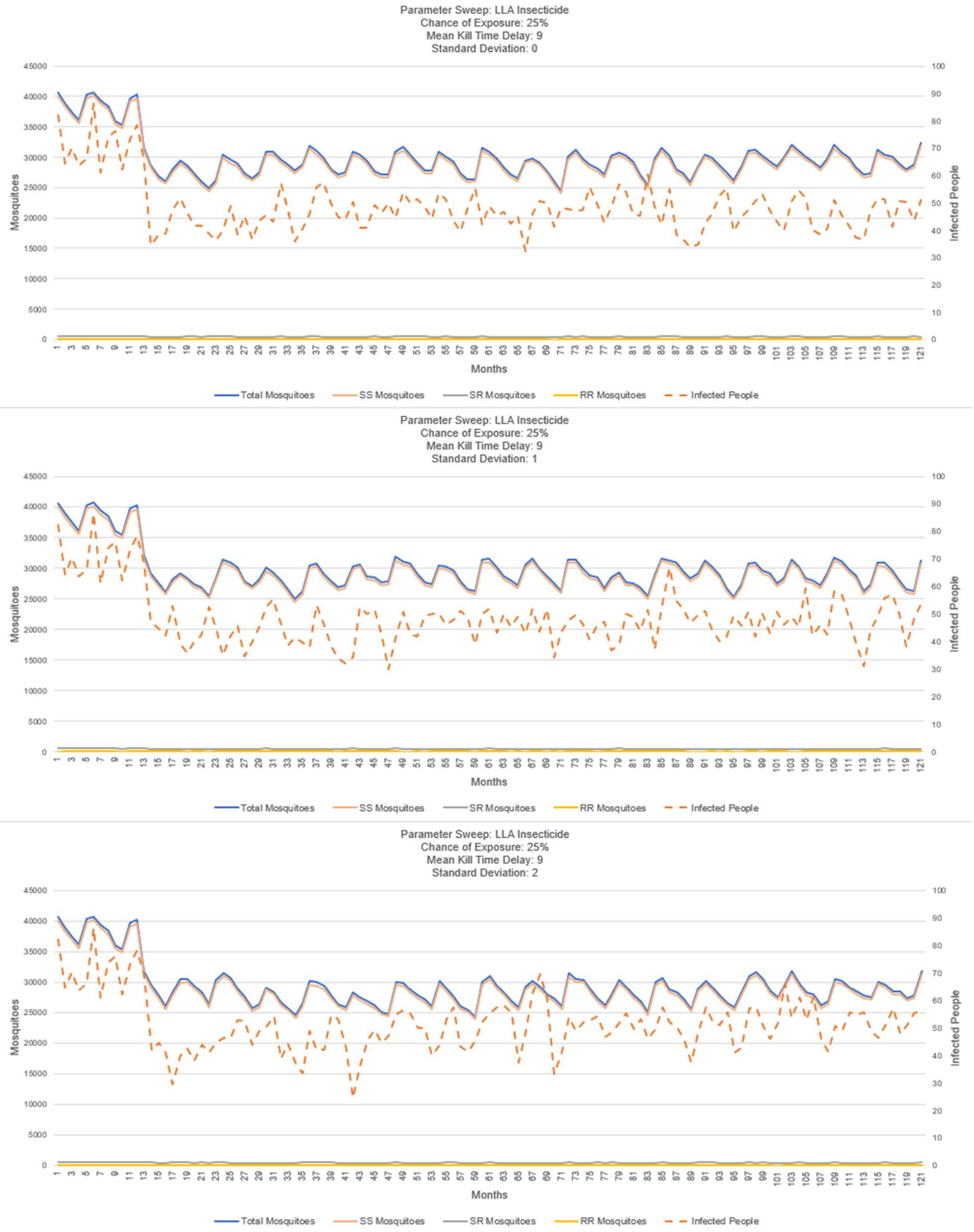

**S4-Fig. 14. LLA insecticide: sweep on SD – mean kill-delay 9 days, exposure chance 25%.**

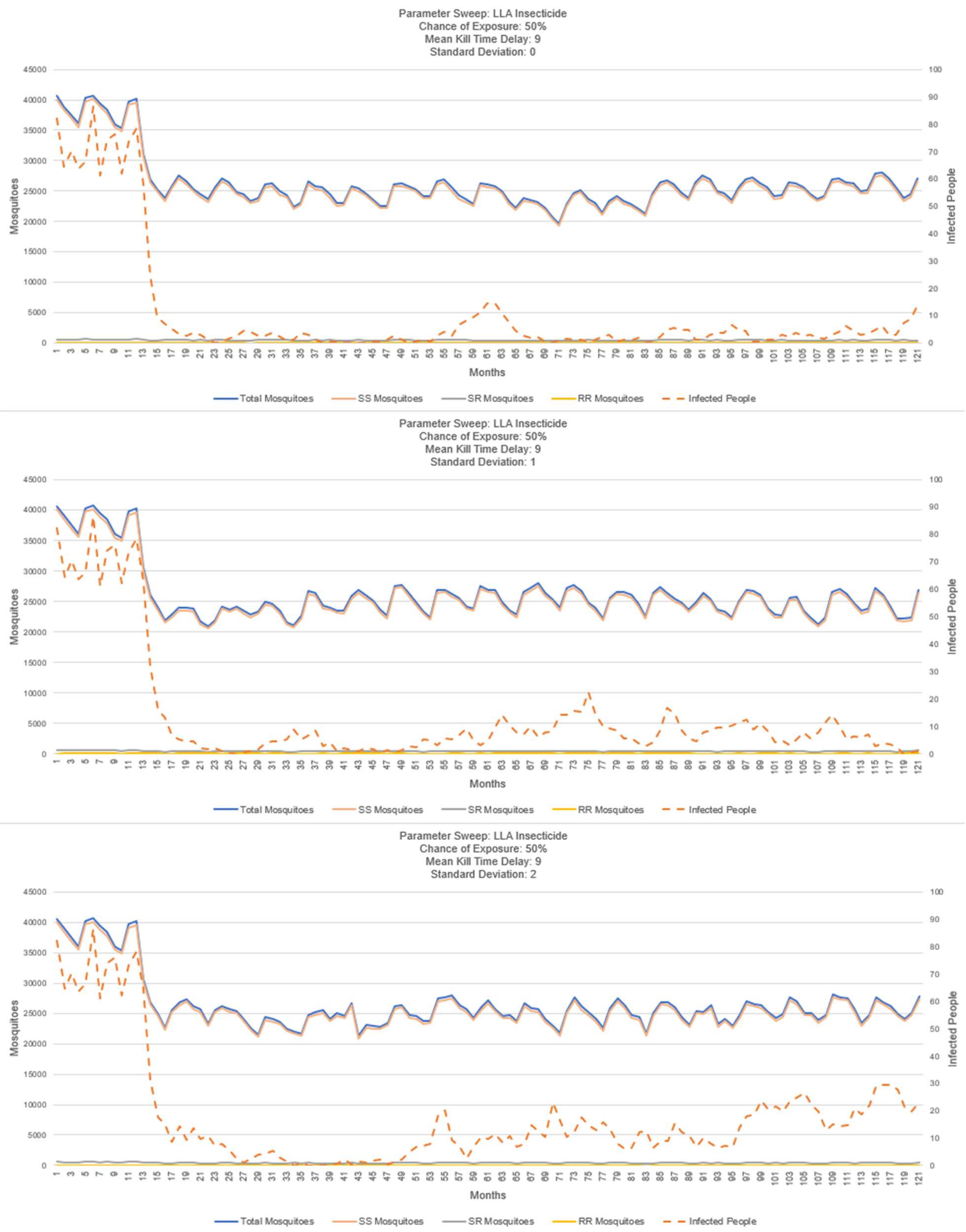

**S4-Fig. 15. LLA insecticide: sweep on SD – mean kill-delay 9 days, exposure chance 50%.**

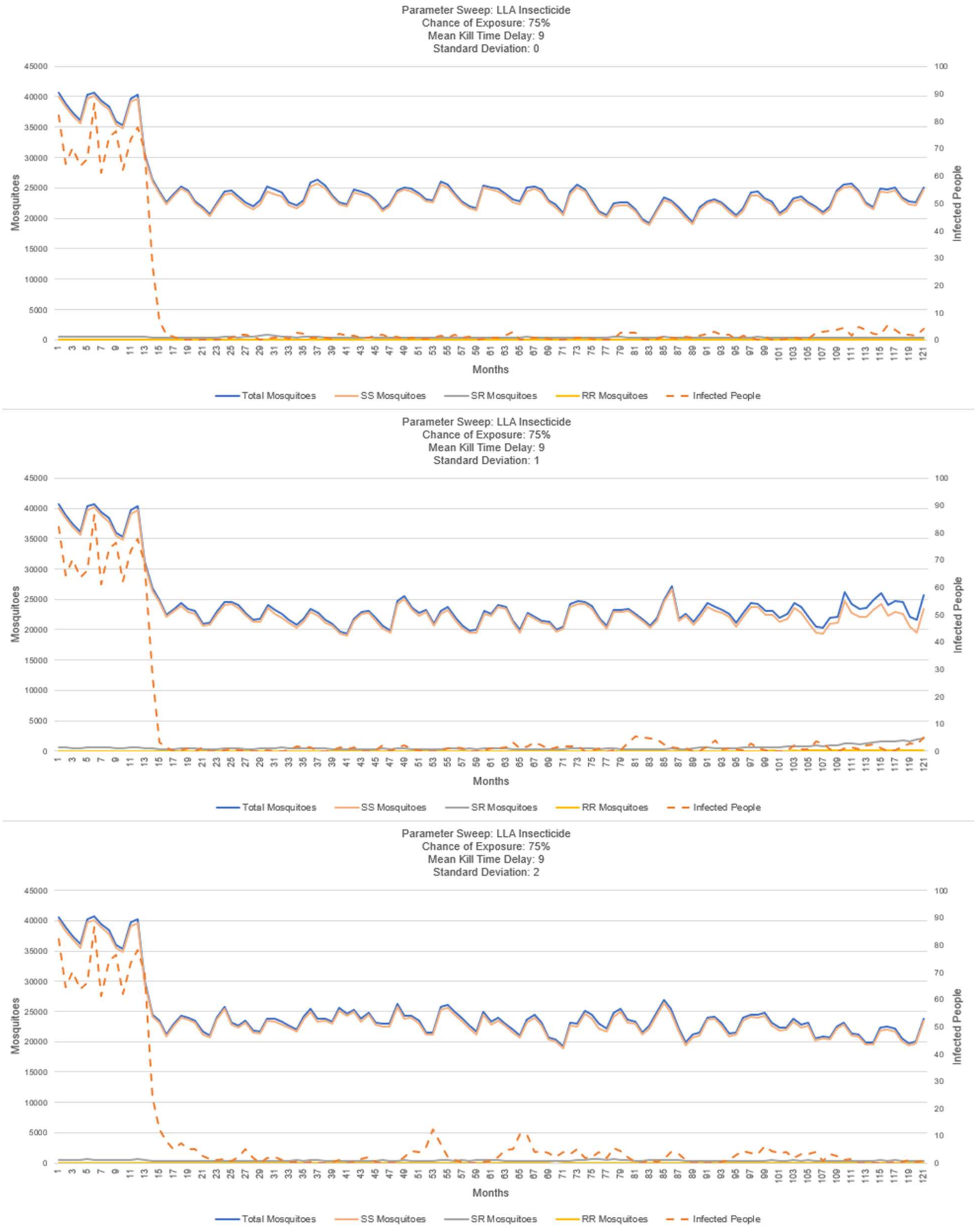

**S4-Fig. 16. LLA insecticide: sweep on SD – mean kill-delay 9 days, exposure chance 75%.**

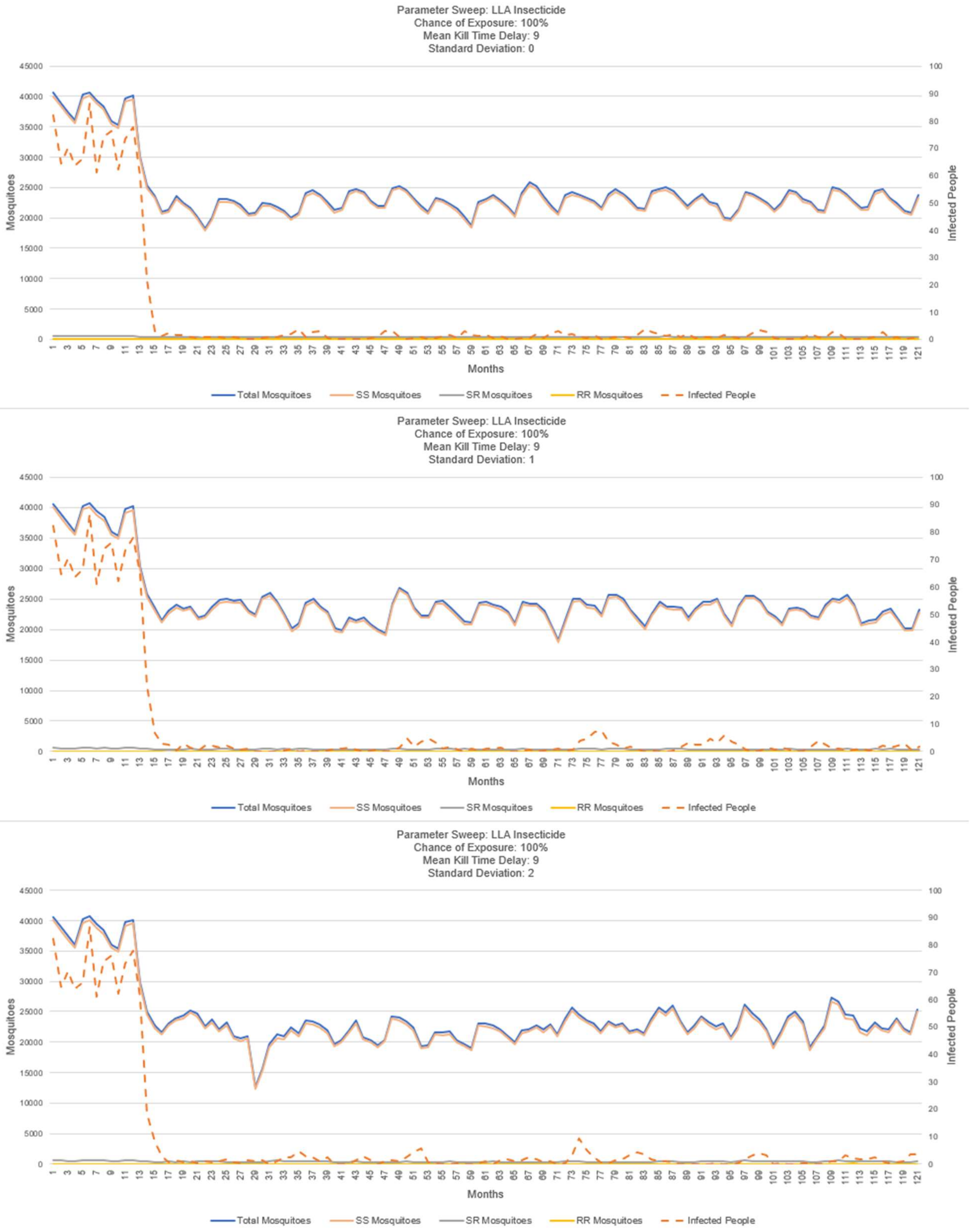

**S4-Fig. 17. LLA insecticide: sweep on SD – mean kill-delay 9 days, exposure chance 100%.**

#### Sensitivity Analysis and Verification & Validation: LLA-Coverage% Sweep

These figures depict the results of a parameter sweep on the amount of the village area that was treated with the LLA insecticide. The insecticide was distributed randomly across the village patches at the rate defined by the "LLA-Coverage%" variable. The BehaviorSpace experimental settings are displayed in S4-Fig. 18. These figures show that the amount of the village that is treated by the insecticide is significantly linked to the performance of the insecticide for controlling transmission. As the coverage increases, the insecticide's ability to reduce malaria transmission increases because more of the infectious mosquitoes are coming into contact with the disease and dying. These experiments were run with a kill time mean of 9 days and a standard deviation of 1 on the LLA insecticide.

The plots showing the number of mosquitoes, the number of each genotype (SS, SR, RR) and the number of infected humans is shown in figures S4-Fig. 19 through S4-Fig. 22.

Observations:

- Across the figures, as the LLA coverage-% increases, the number of mosquitoes decreases and the human infection decreases; this is expected and supports the verification & validation of the simulation.
- Across the figures, as the LLA coverage-% increases, a slight emergence of the resistant genotype appears in one case (coverage 30%); this can be attributed to the simulation's dependence on the starting random seed, and suggests additional repetitions with different seeds. See the experiment and results presented in figures S4-Fig. 29 and S4-Fig. 30.

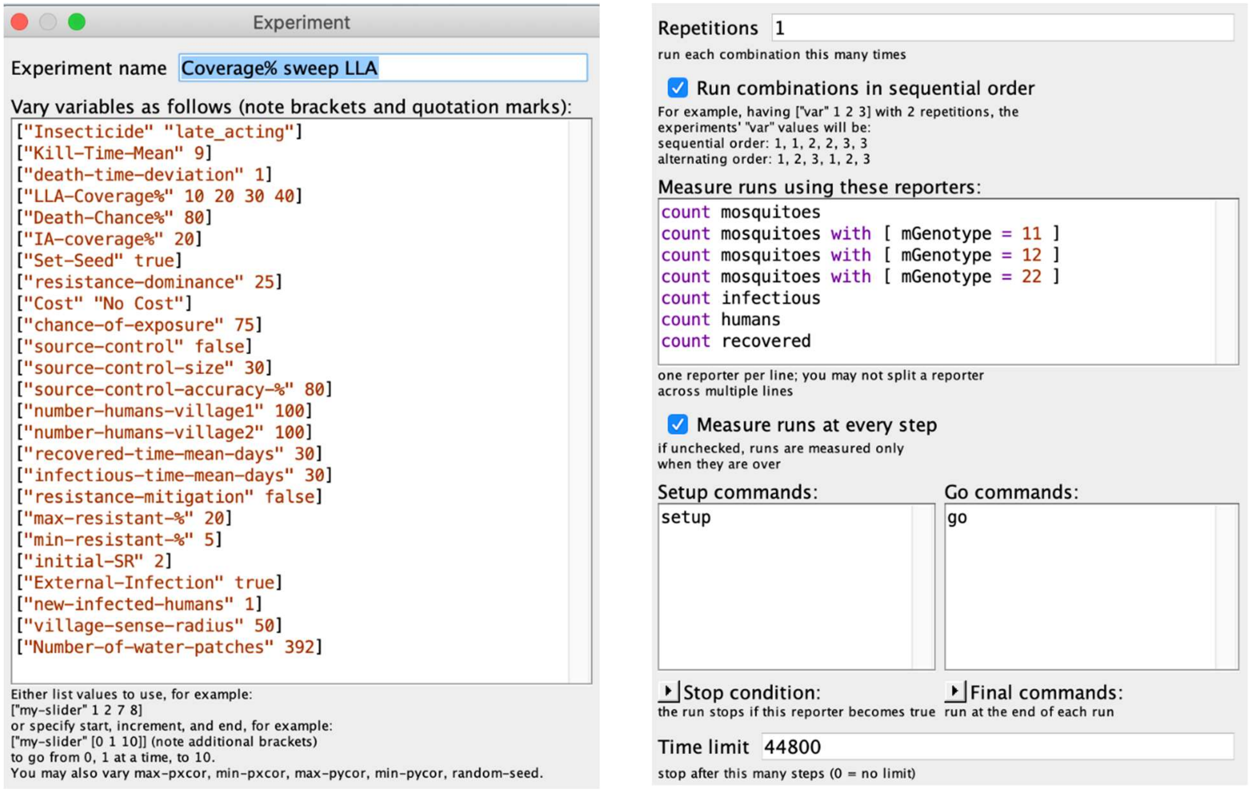

**S4-Fig. 18. LLA. Experimental settings for LLA-Coverage runs.**

Note: sweep over LLA-coverage values: 10%, 20%, 30%, 40%

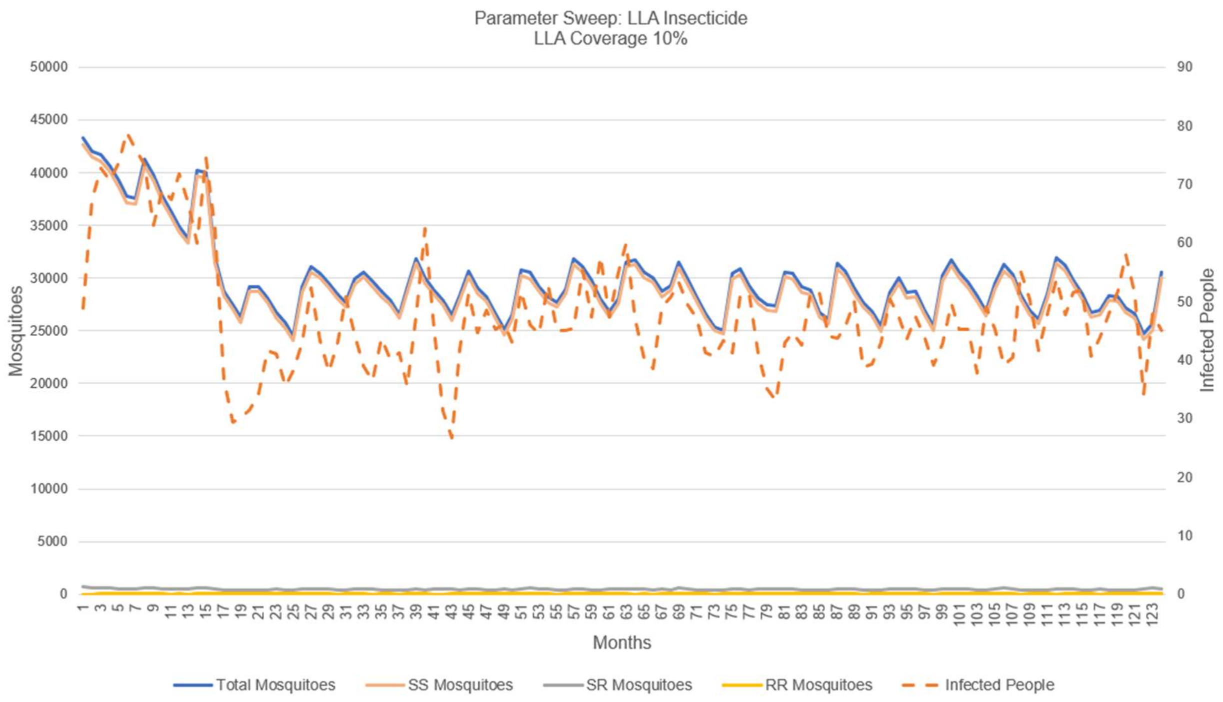

**S4-Fig. 19. LLA insecticide coverage 10%.**
Note: modest reduction in human infections, no emergence of the resistant genotype.

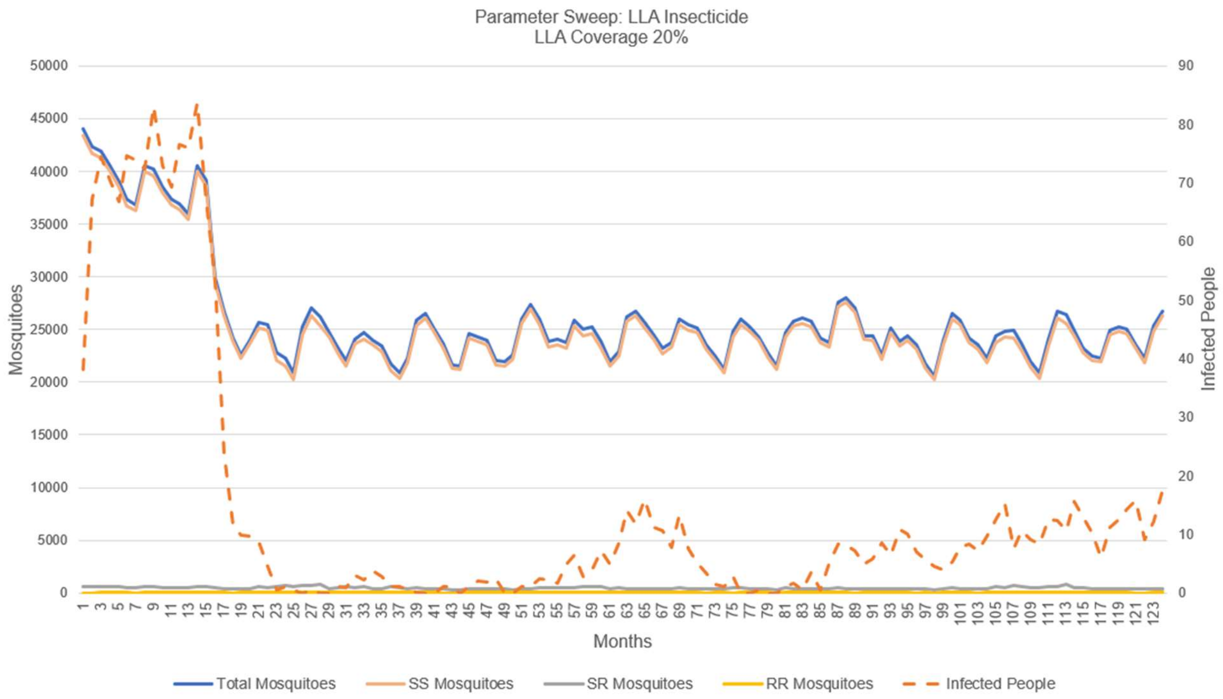

**S4-Fig. 20. LLA insecticide coverage 20%.**
Note: significant reduction in human infections, no emergence of the resistant genotype.

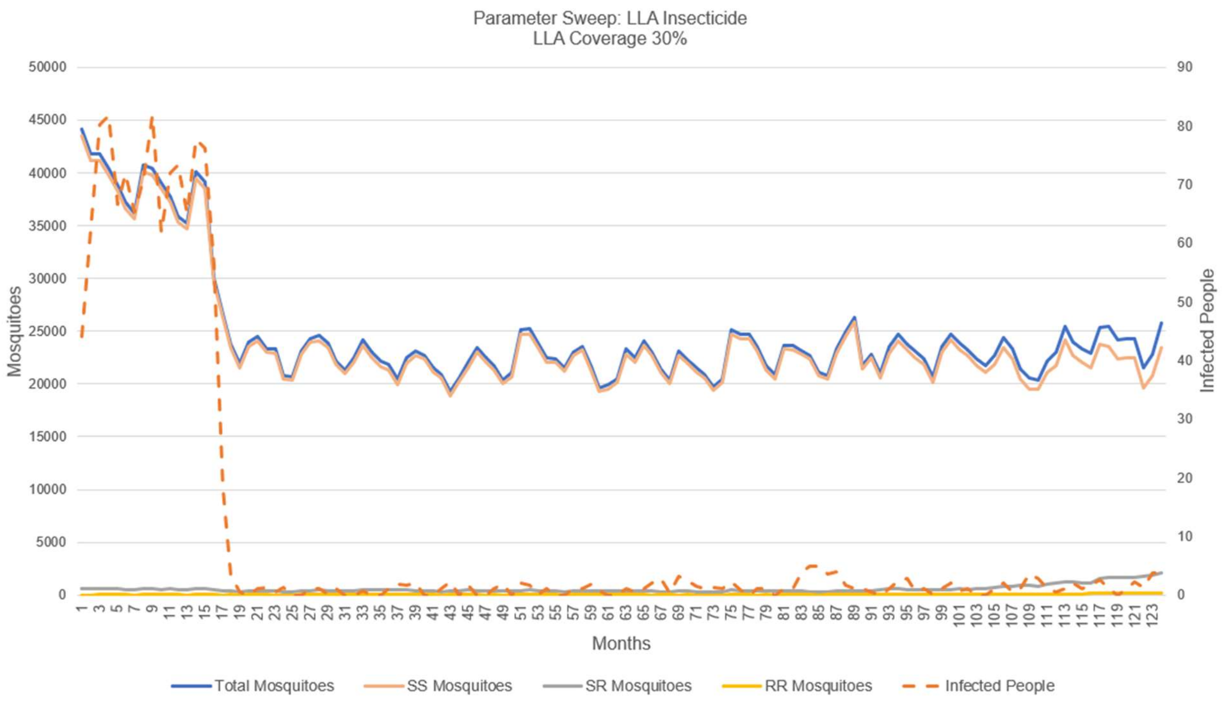

**S4-Fig. 21. LLA insecticide coverage 30%.**

Note: almost total reduction in human infections, slight emergence of the resistant genotype.

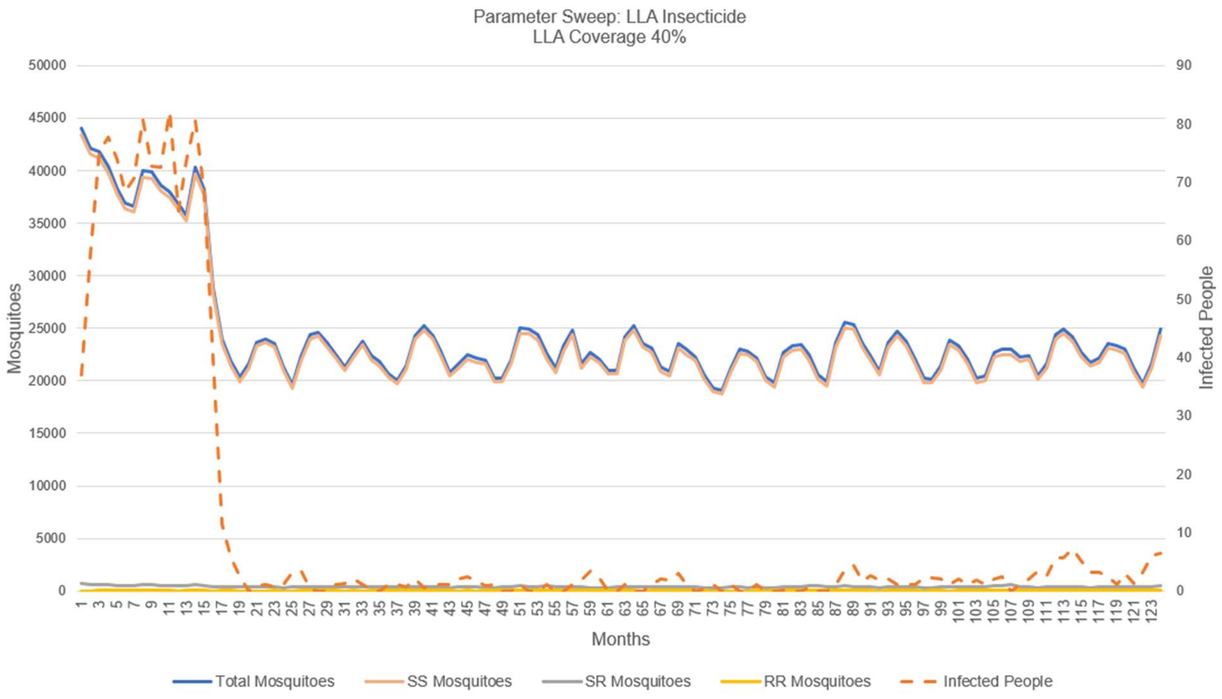

**S4-Fig. 22. LLA insecticide coverage 40%.**

Note: almost total reduction in human infections, no emergence of the resistant genotype

#### Evaluation of the Importance of LLA Insecticide's Mean Kill-Time-Delay and Standard Deviation of the Kill-Time-Delay

These figures show the results of a parameter sweep on two of the key performance variables for the LLA insecticide: the mean and the standard deviation on the kill-time-delay from exposure to mortality. The BehaviorSpace experimental settings are displayed in S4-Fig. 23. The results are plotted in figures S4-Fig. 24 through S4-Fig. 28.

Notes:

- The timing discussed here is based on assumptions about the durations of the mosquitoes' gonotrophic cycle, the assumed hourly death rate of the mosquitoes from natural causes, the hourly distance traveled by each mosquito when searching for a blood meal or a breeding site, etc. These are calibration parameters that would need to be "tuned" to represent the malaria transmission patterns in a specific locality.
- A few of the plots do not appear to be entirely consistent, raising verification & validation concerns. An example can be seen in figure S4-Fig. 24 where the timing of the emergence of IR does not trend as expected, e.g., a SD of two days appears later (88 months) than a SD of one day (50 months); we expected the opposite. In the subsequent section we conclude that this apparent anomaly is not a verification & validation issue, but simply caused by viewing one run and the potential randomness of a stochastic simulation. See figures S4-Fig. 29 and S4-Fig. 30. These experiments support the verification and validation of the simulation.
- The observations above suggest the potential value of the simulation in support of the design of a LLA insecticide.

Observations:

- These results demonstrate the significance of precision in the mean kill-time-delay of an LLA insecticide. It can be seen that, while shorter kill-time-delays are more effective at reducing the malaria transmission in the model, the LLA insecticides with low kill-time-delays also are more likely to produce resistance after a few years. However, it can also be seen that as the kill-time-delay increases, the ability for the insecticide to control the malaria transmission decreases. There is, therefore, a balance in the middle of the kill-time-delays which we tested where the insecticide both acts late enough to avoid incurring resistance and early enough to be able to control the malaria transmission to a reasonable degree. We see this balance to be most expressed in the kill-time-delay with mean of 9 days.
- We also observed, as expected, that larger standard deviations produced worse results in both the deterrence of resistance and control of the malaria transmission. We relate this to

the decrease in accuracy that raising the standard deviation produces which therefore causes the insecticide to kill more mosquitoes outside of the target age group.

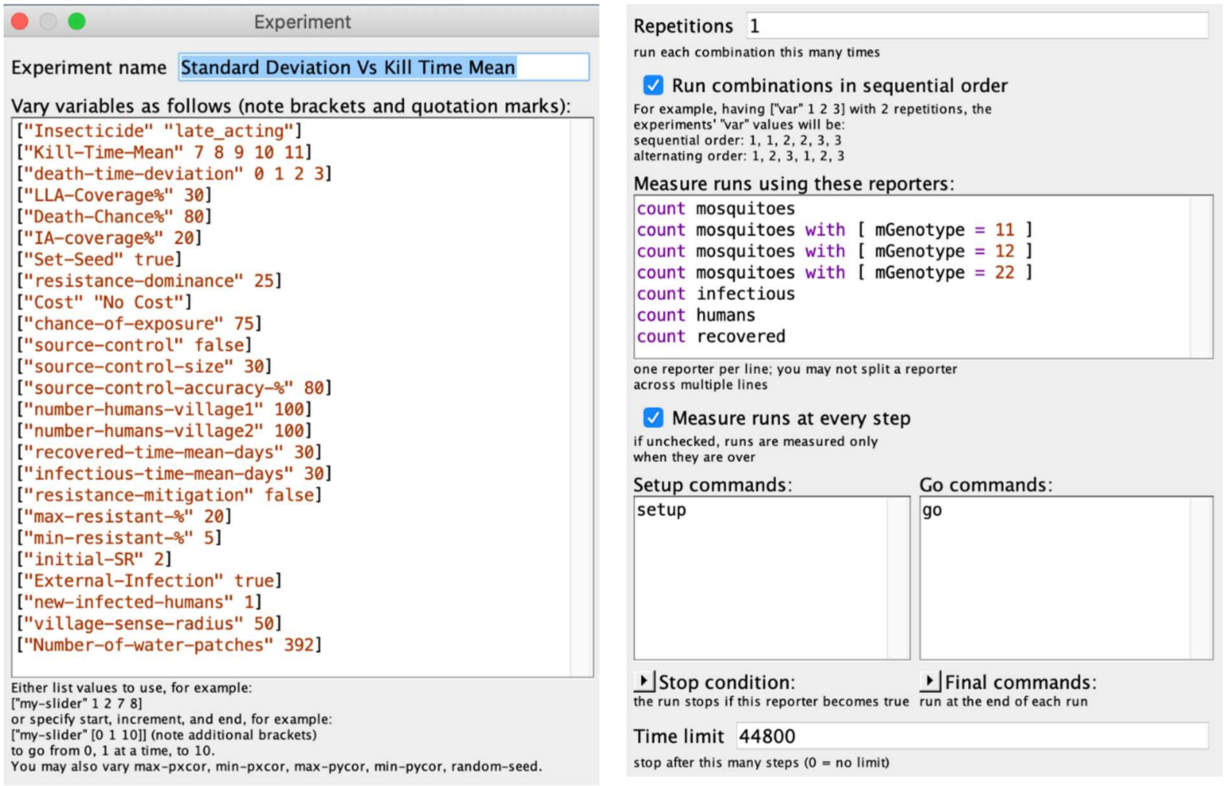

**S4-Fig. 23. Experimental settings for kill-time-delay.**
Sweep on mean kill-time-delay and standard deviation around kill-time-delay.

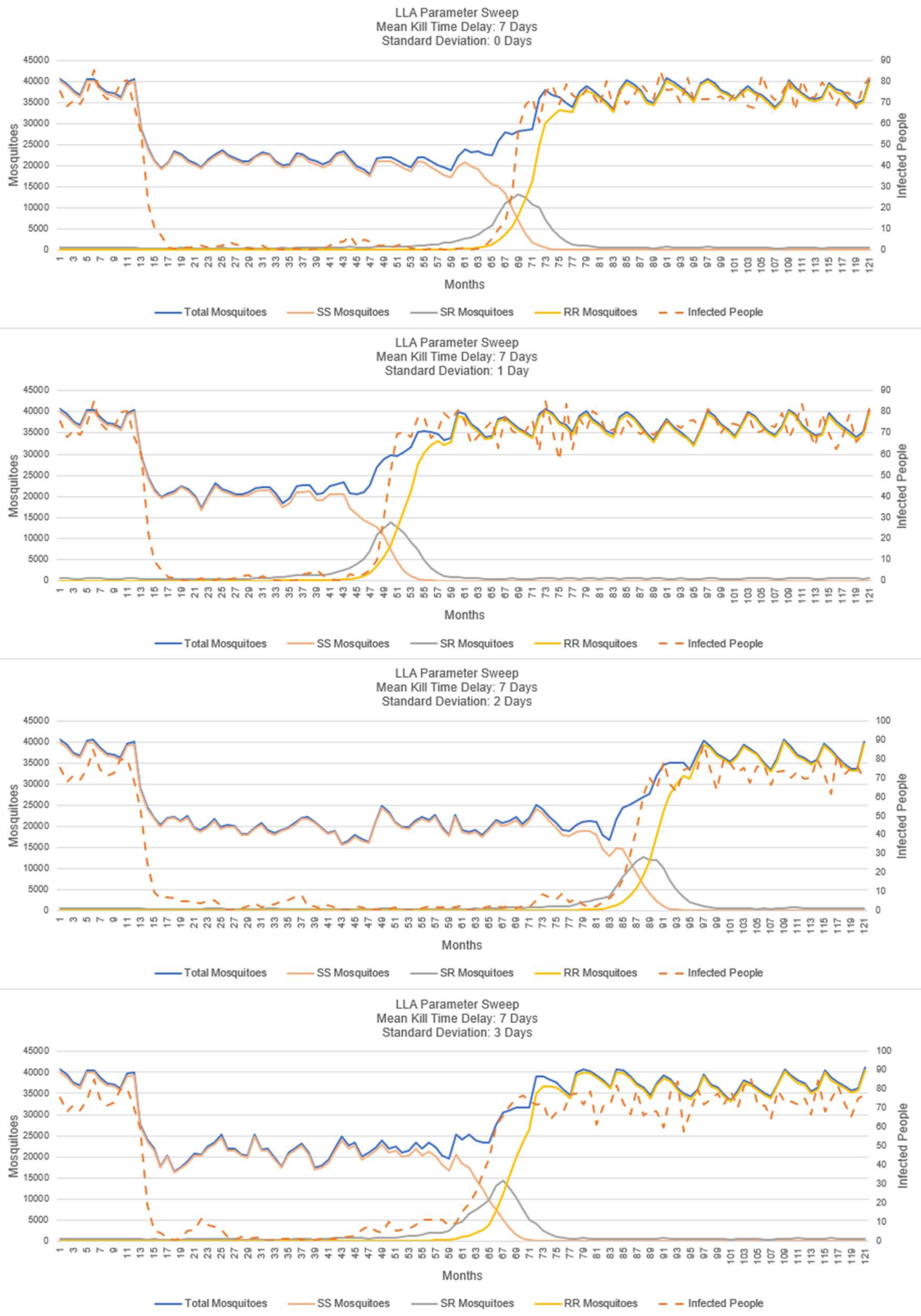

**S4-Fig. 24. LLA insecticide: sweep on SD – mean kill-time-delay 7 days.**

Note: apparent anomaly caused by only one run of the simulation. See next section.

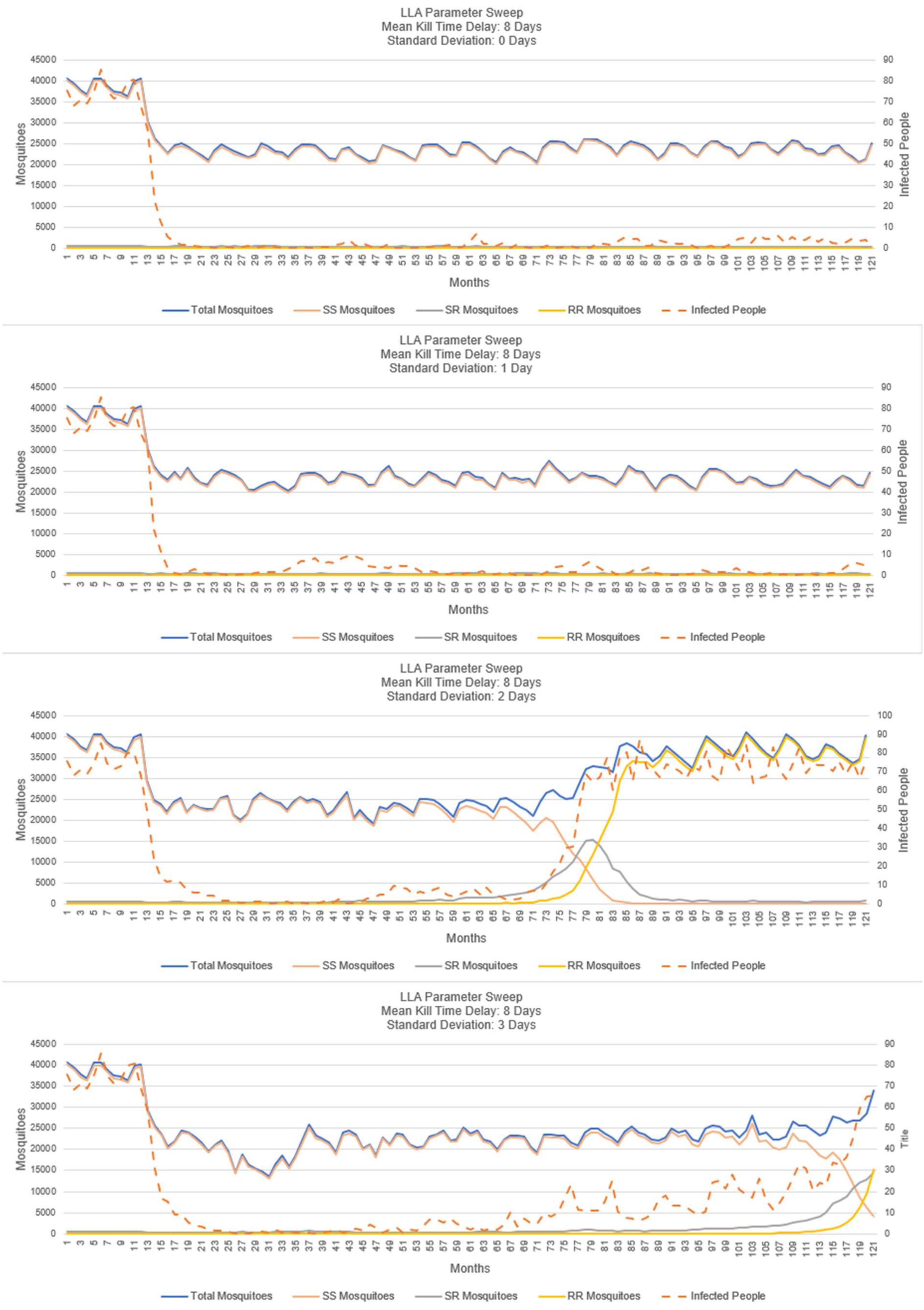

**S4-Fig. 25. LLA insecticide: sweep on SD – mean kill-time-delay 8 days.**

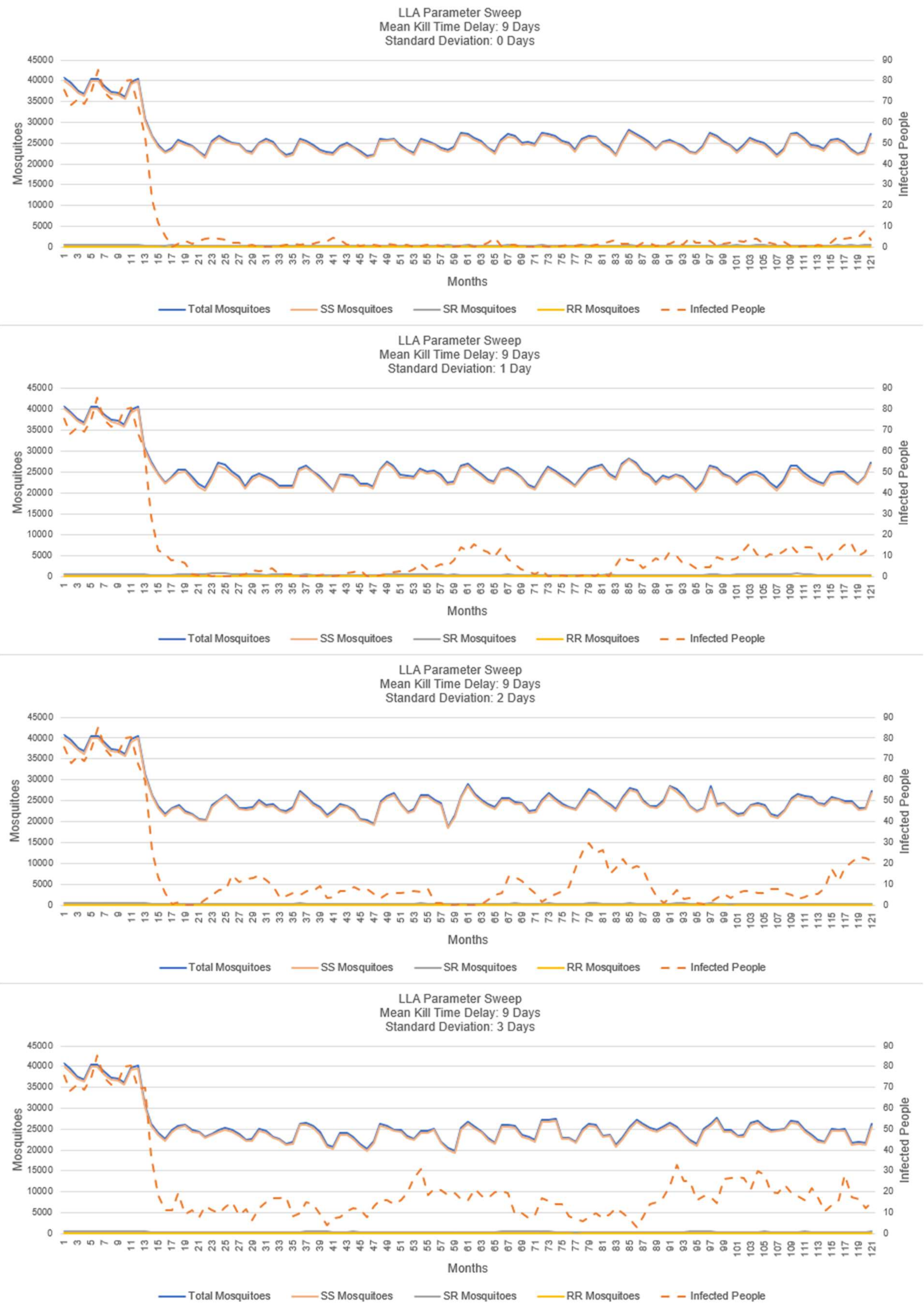

**S4-Fig. 26. LLA insecticide: sweep on SD – mean kill-time-delay 9 days.**

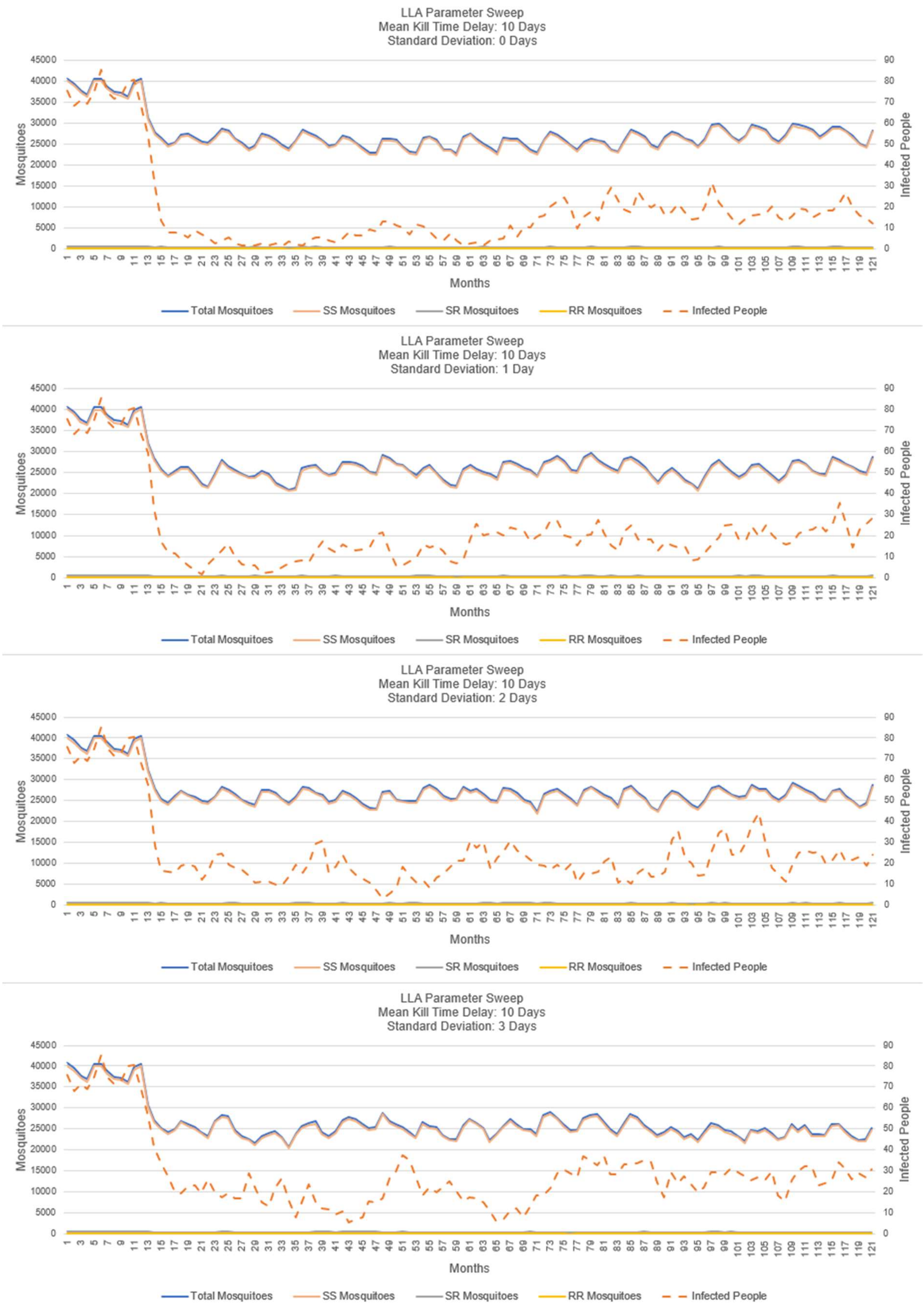

**S4-Fig. 27. LLA insecticide: sweep on SD – mean kill-time-delay 10 days.**

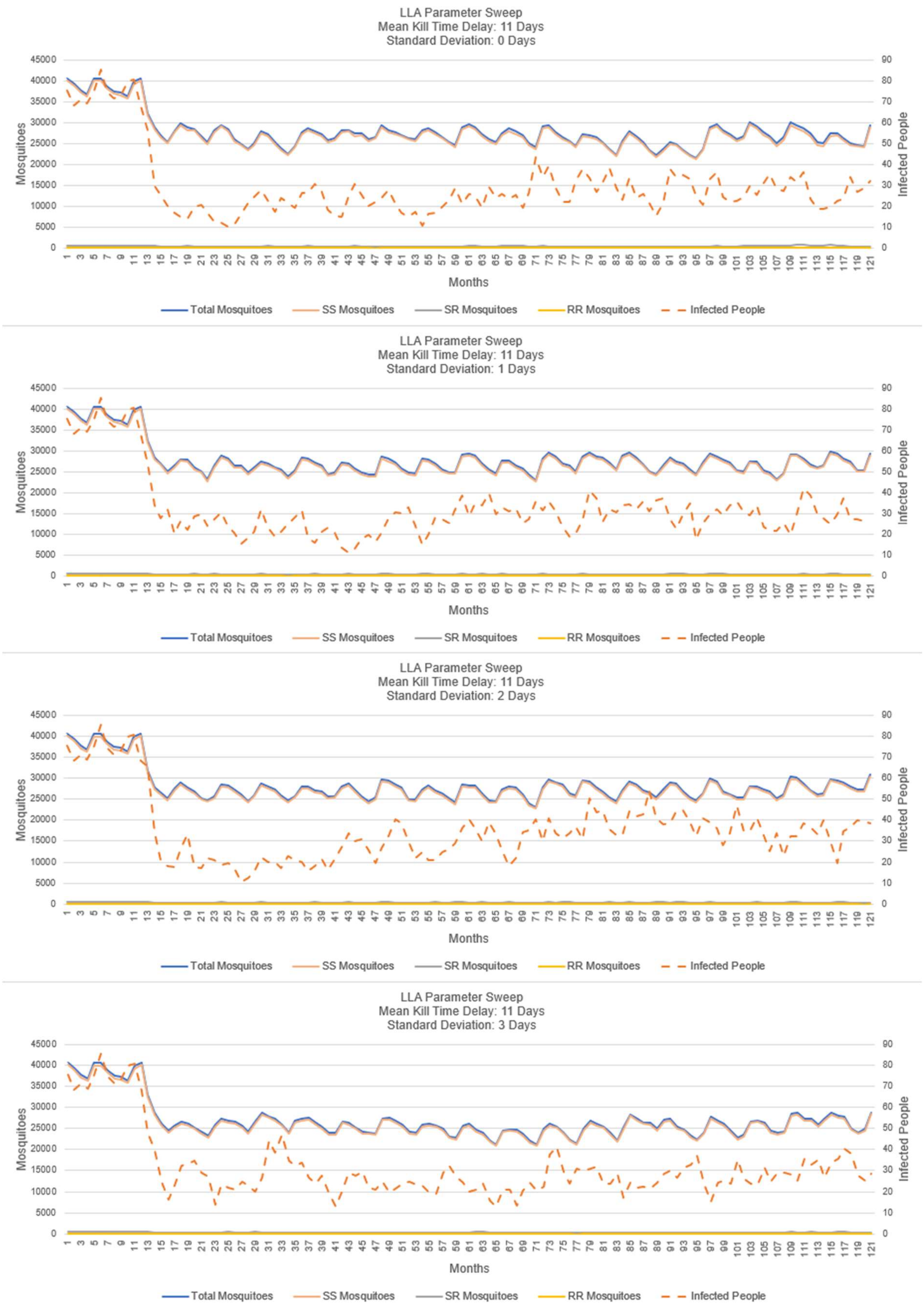

**S4-Fig. 28. LLA insecticide: sweep on SD – mean kill-time-delay 11 days.**

#### Sensitivity Analysis – LLA Insecticide with Random Seed Repetitions

In figure S4-Fig. 24, we observed an apparent anomaly in the lack of a trend in the time it took for insecticide resistance to peak so we investigated further by testing multiple random seeds to see if this was caused by the randomness associated with the starting seed, or was a verification or validation issue with the simulation. In this experiment, three repetitions were simulated (using different random seeds) so that we could test the randomness of the time before the development of resistance. We discovered that this time was actually quite random and was highly dependent on the seed for this experiment. The BehaviorSpace setting for this experiment are displayed in figure S4-Fig. 29. Plots of the results are displayed in figure S4-Fig. 30. Additional sensitivity tests of the choice of initial random number generator are presented in figures S4-Fig. 7, S4-Fig. 40, and S4-Fig. 43

Notes:

- All runs use kill-time-mean of 7 days; three repetitions each with a different starting random seed.

Observations:

- We observe that choice of starting random seed generates rather different times for the emergence of IR. In figure S4-Fig. 30 for SD of 0 days, we see two replications with IR emergence close to 55 months, while the third replication has no IR emergence for the duration of the simulation. For SD of 1 day, emergence appears around 45, 54, and 102 days. The pattern continues confirming that there is a sensitivity to the choice of the starting random seed.
- Proper analysis for this particular investigation will require multiple repetitions of the simulation and the results will need to be interpreted and presented as frequency distributions, as in the typical nature of many stochastic simulations. This supports the verification and validation of the simulation vis-à-vis the apparent anomaly observed in figure S4-Fig. 24.

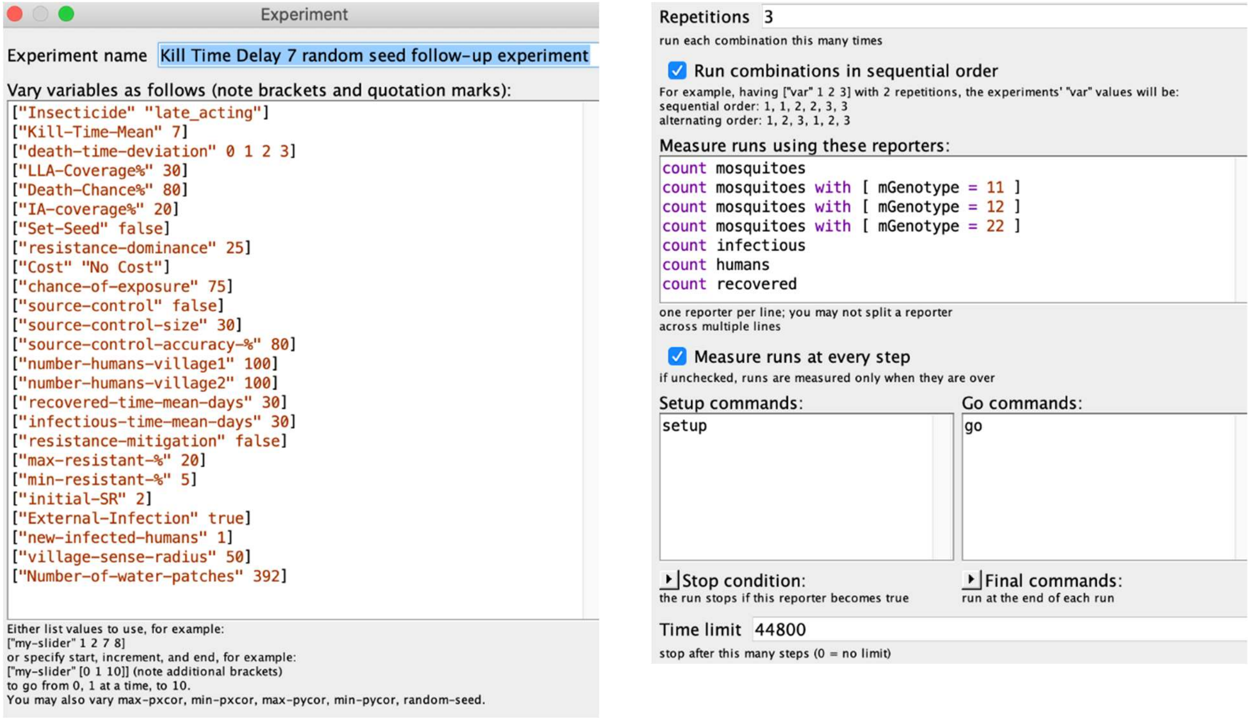

**S4-Fig. 29. Experimental settings, random seed sensitivity analysis, LLA Insecticide.**
Note: All runs use kill-time-mean of 7 days; three repetitions each with a different random seed.

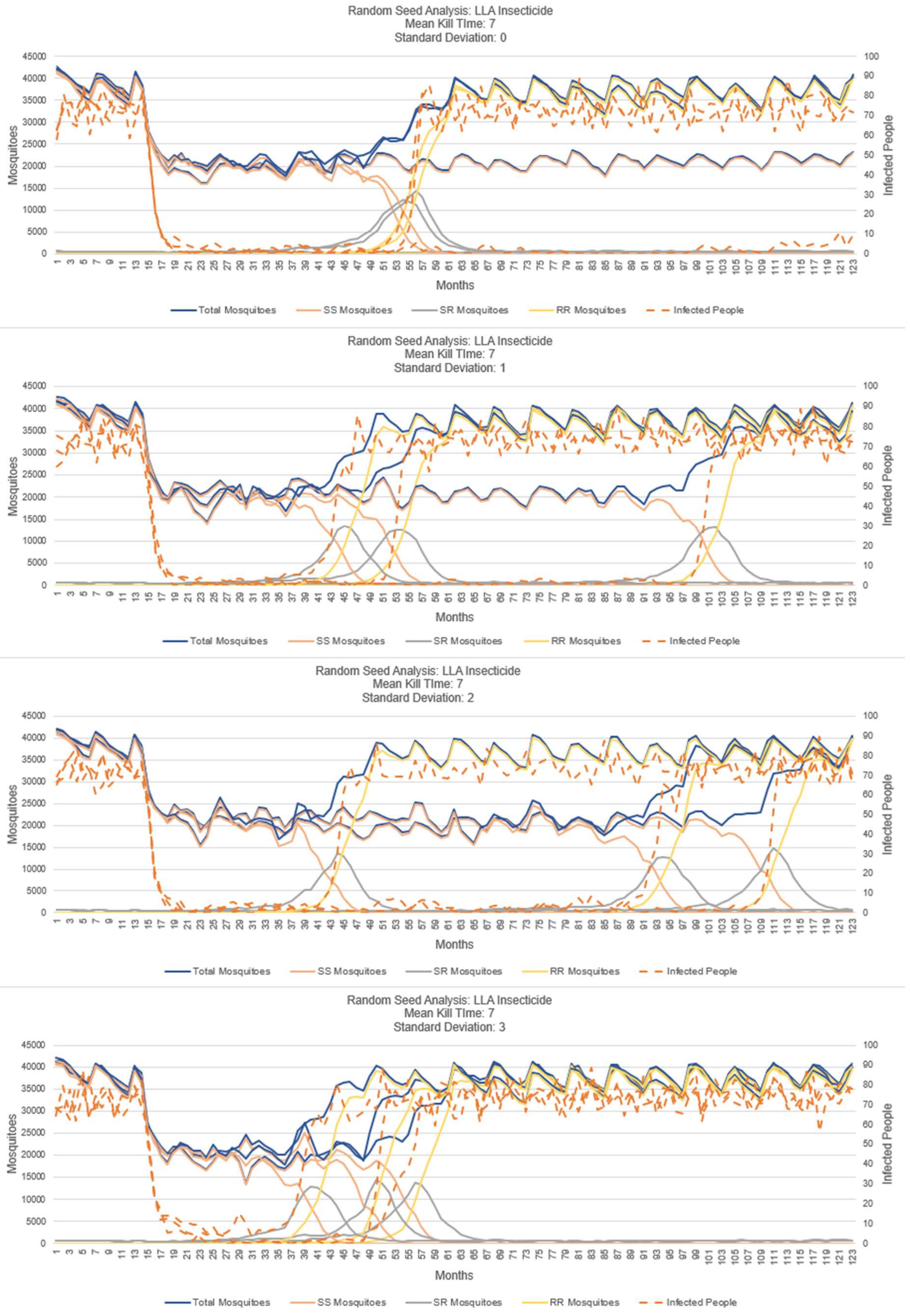

**S4-Fig. 30. Random seed sensitivity analysis, LLA Insecticide.**

#### Sensitivity Analysis and Verification & Validation of Resistance Dominance for LLA Insecticide

These figures display the result of a parameter sweep on the "Resistance Dominance" variable for a LLA insecticide. This variable controls how resistant the heterozygous genotype (SR) mosquitoes are to the insecticide. A higher vale for Resistance Dominance means that the SR mosquitoes are more resistant. A Resistance dominance value of 100 means that there is no difference between the SR mosquitoes and the RR mosquitoes in terms of IR while a value of 0 would mean that the SR mosquitoes have no difference from the SS mosquitoes in terms of resistance. The BehaviorSpace settings for this experiment are displayed in figure S4-Fig. 31. Plots of the results are presented in figures S4-Fig. 32 though S4-Fig. 39.

Notes:

- Only one run of each set of intervention values is presented to keep plots from becoming overly complex. A complete analysis would require multiple repetitions with different ransom seeds.
- Each figure also shows three plots for three values of mean-delay-time to death after exposure for values 8, 9, and 10 days. We observe the sensitivity of the mean-delay-time to death; too short a mean-delay-time to death and IR emerges quickly, while too long a mean-delay-time to death IR is not observed but the effectiveness at controlling malaria transmission is reduced. As an example, see figure S4-Fig. 34. For a kill-time-mean of 8 days IR emerges in the presented run around 70 months, but for mean values of 9 and 10 days, no emergence appears. When IR emerges (mean 8 days) transmission control fails as seen in the number of infected humans returning to the baseline (first pre-intervention 12-month period). When comparing the mean times of 9 and 10 days, we see an increase in transmission for the 10 day kill-time-mean over the 9 day mean. This sensitivity analysis suggests the potential value of the simulation in support of the design of a LLA insecticide, i.e., the importance of the mean-kill-time of the insecticide.

Observations:

- These plots show that the higher the resistance dominance, the more quickly that IR emerges. For example, compare figures S4-Fig. 32, S4-Fig. 34, S4-Fig. 36, and S4-Fig. 38. This supports the validation of the simulation.
- In figures S4-Fig. 38 and S4-Fig. 39, an interesting result can be seen when resistance dominance is 100%. At that level, there is no selective advantage of the RR genotype over the SR genotype. Thus, unlike lower values of resistance dominance where the SS and SR genotypes in the population eventually decrease to zero (see figure S4-Fig. 36), for the 100% resistance dominance level the SS and SR genotypes persist in the population. The SS genotype mosquitoes remain in the simulation because of the

constant presence of the SR mosquitoes which produce a proportion of SS mosquitoes when they mate with another SR mosquito.

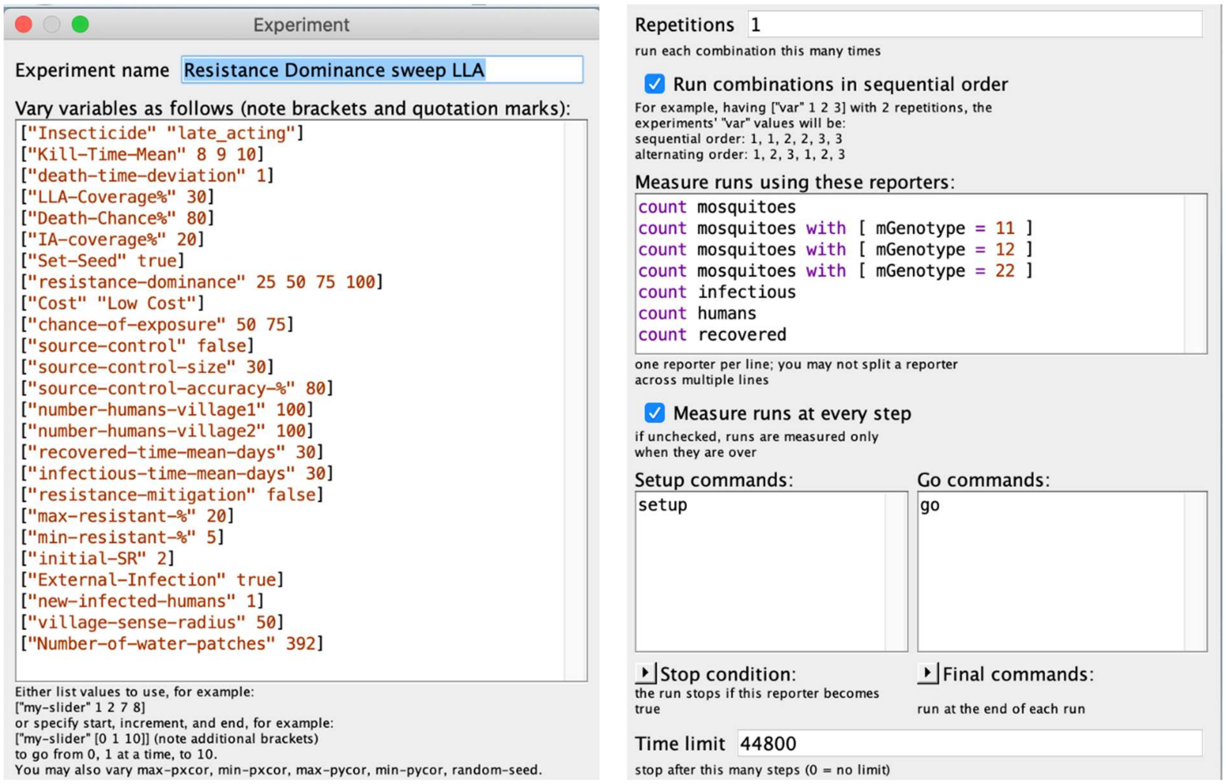

**S4-Fig. 31. Settings – Parameter sweep: Resistance Dominance for LLA Insecticide**

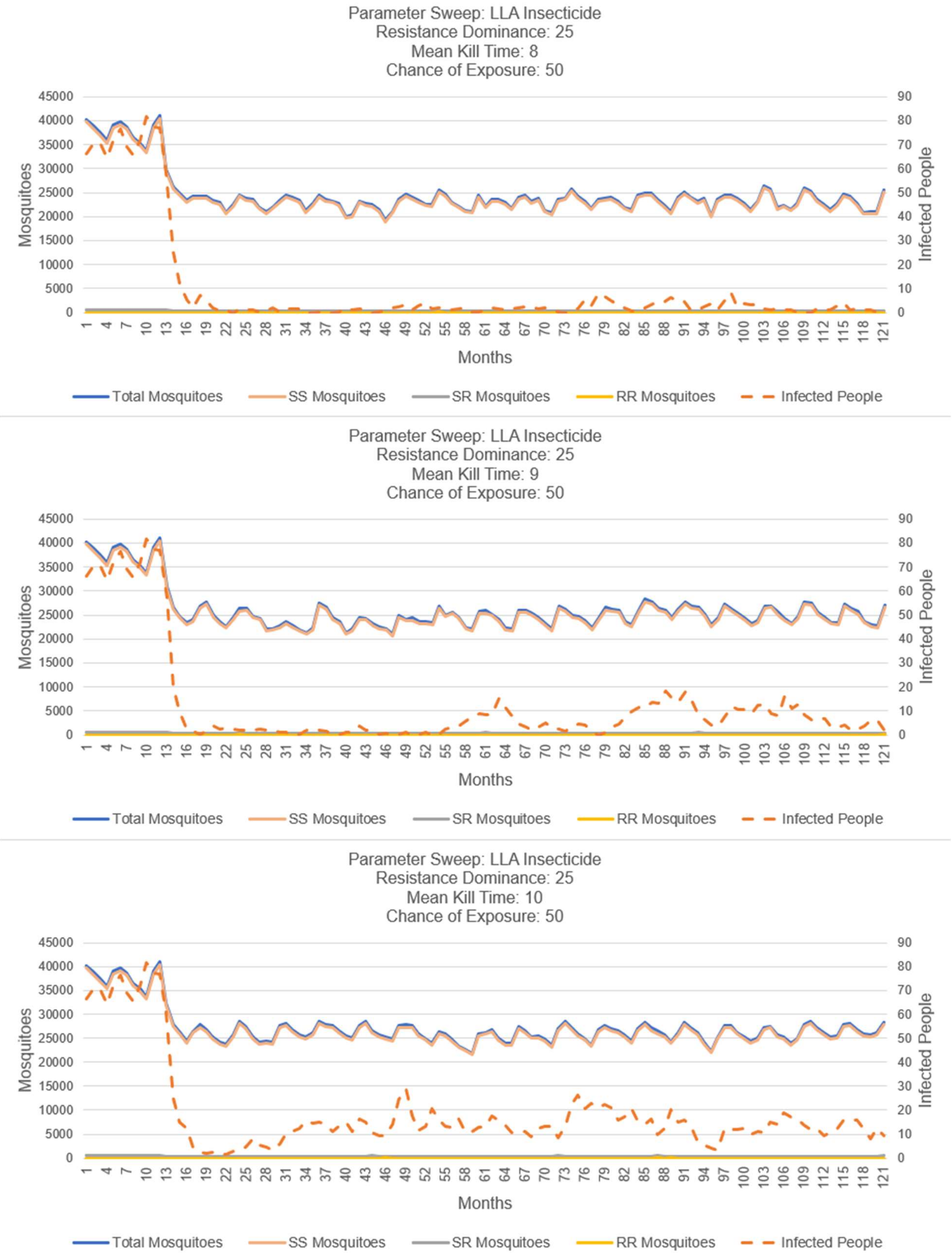

**S4-Fig. 32. LLA insecticide, resistance dominance 25%, chance-of-exposure 50%.**
Note: sweep over mean-kill-time.

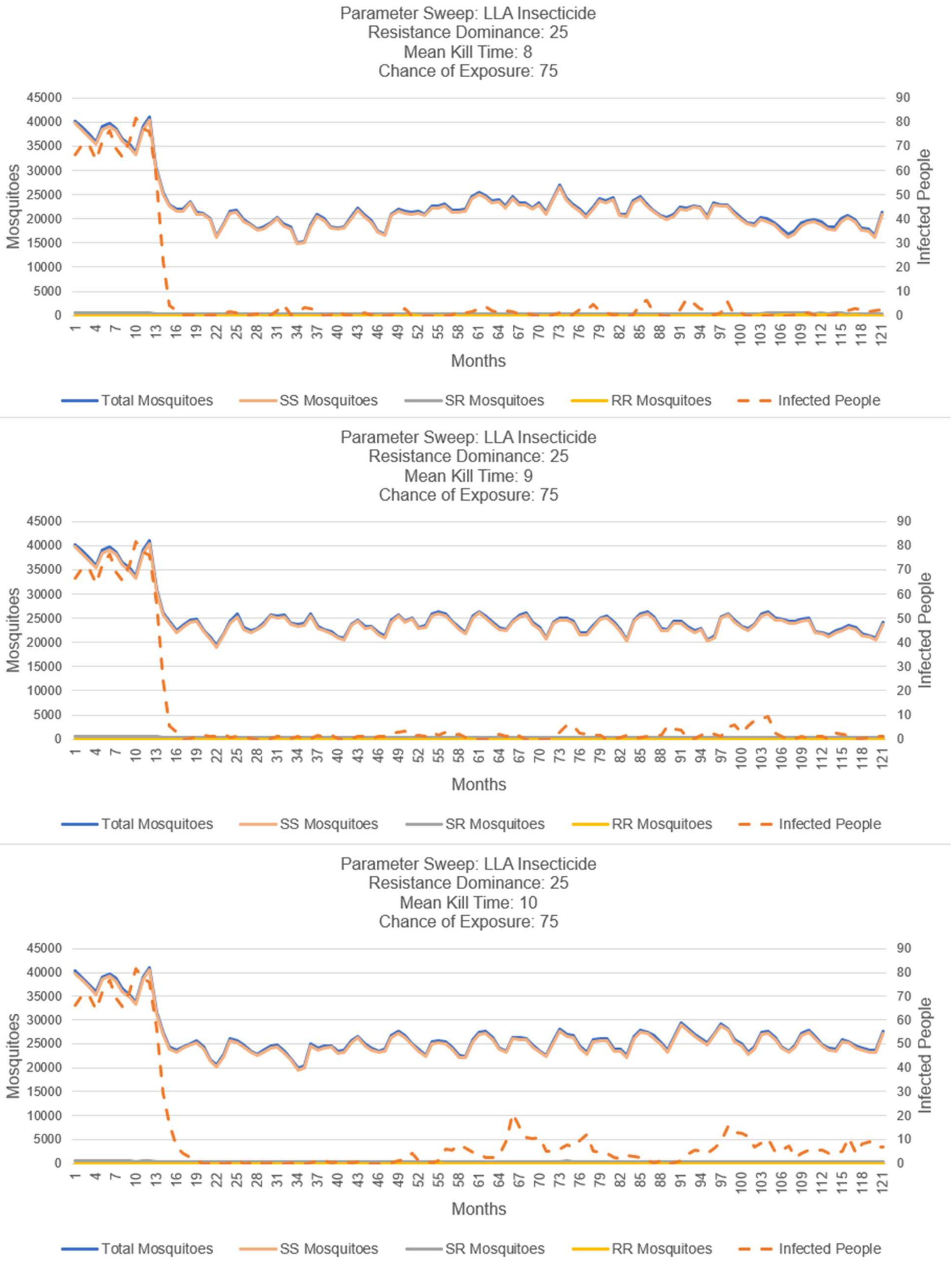

**S4-Fig. 33. LLA insecticide, resistance dominance 25%, chance-of-exposure 75%.**

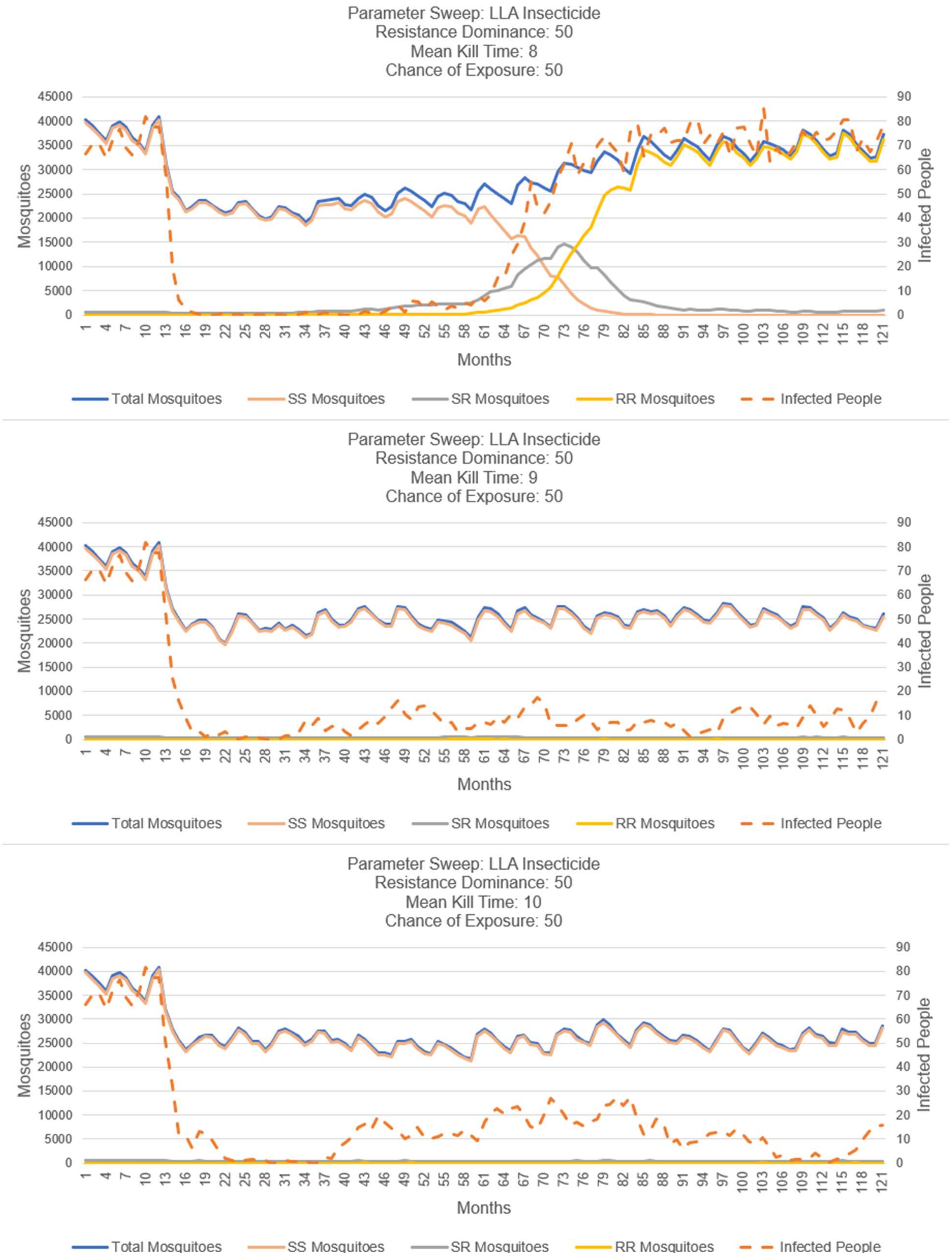

**S4-Fig. 34. LLA insecticide, resistance dominance 50%, chance-of-exposure 50%.**

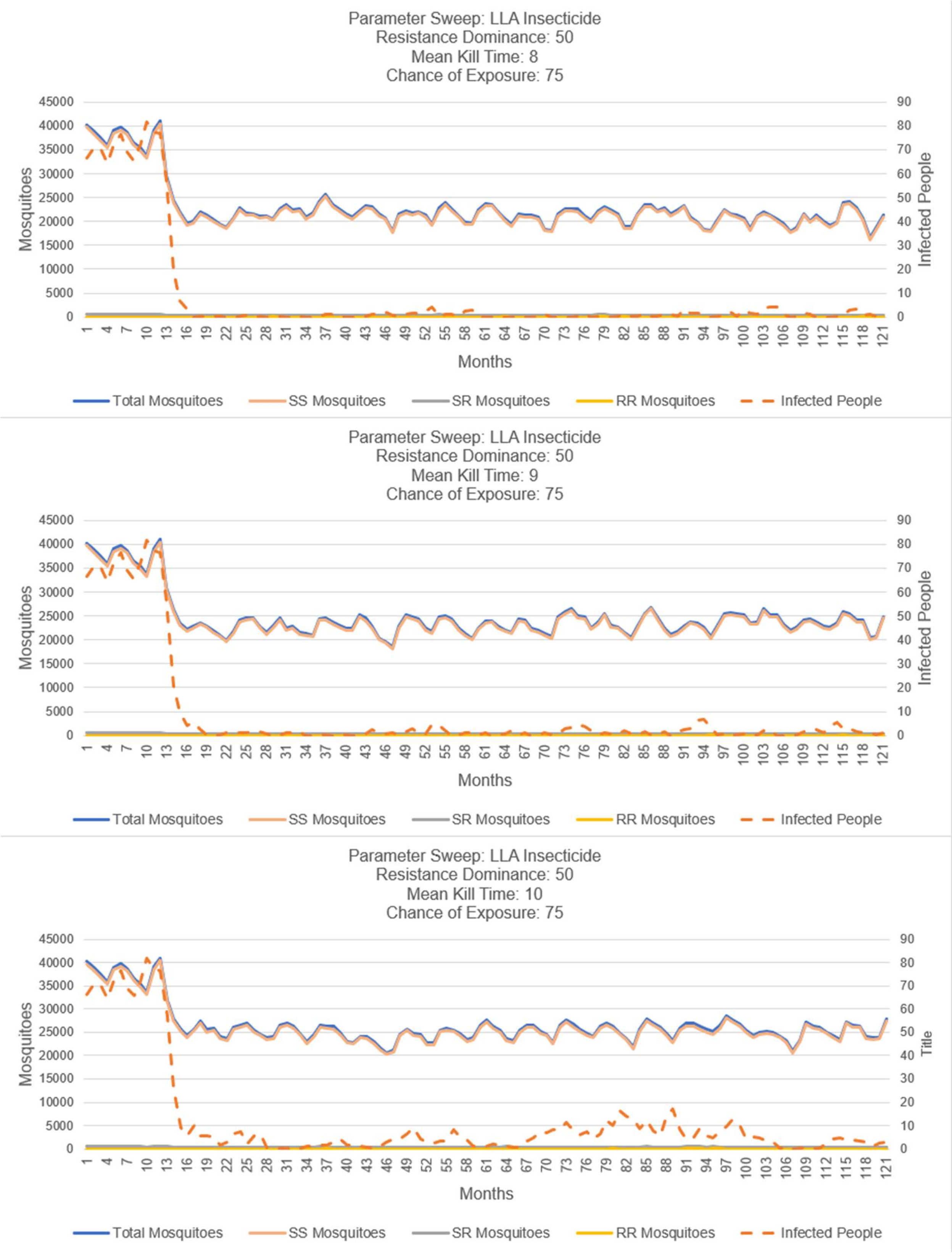

**S4-Fig. 35. LLA insecticide, resistance dominance 50%, chance-of-exposure 75%.**

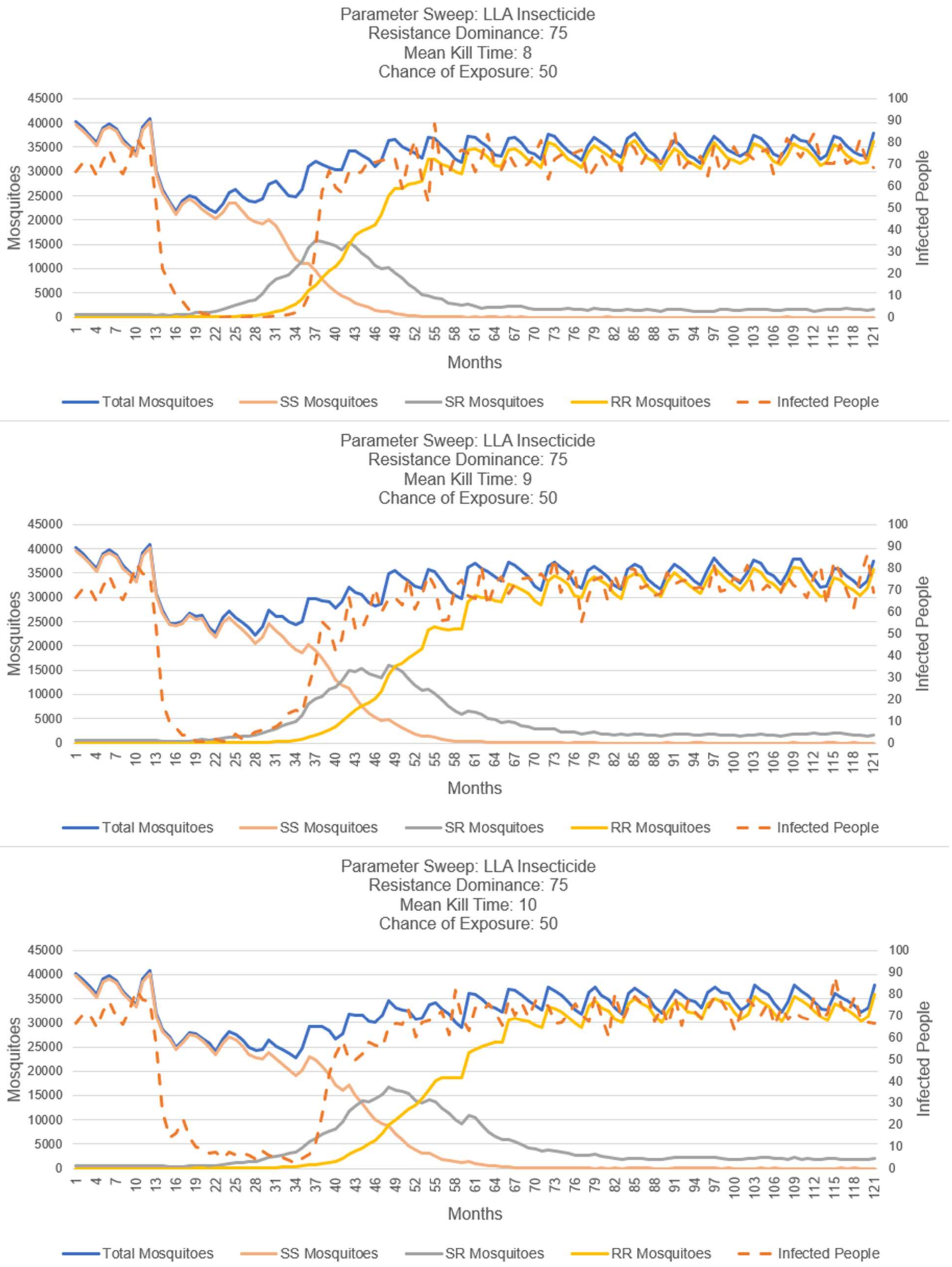

**S4-Fig. 36. LLA insecticide, resistance dominance 75%, chance-of-exposure 50%.**

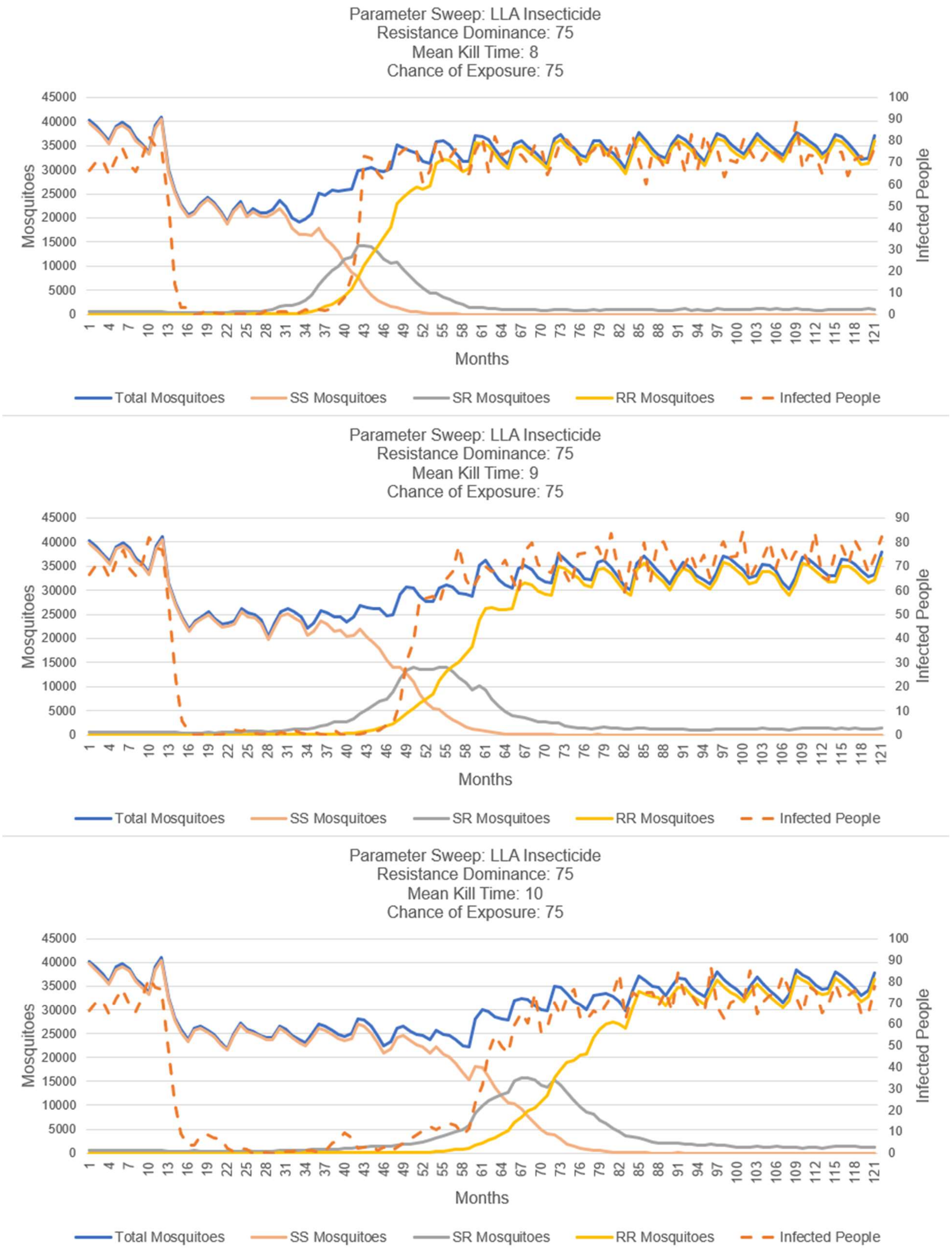

**S4-Fig. 37. LLA insecticide, resistance dominance 75%, chance-of-exposure 75%.**

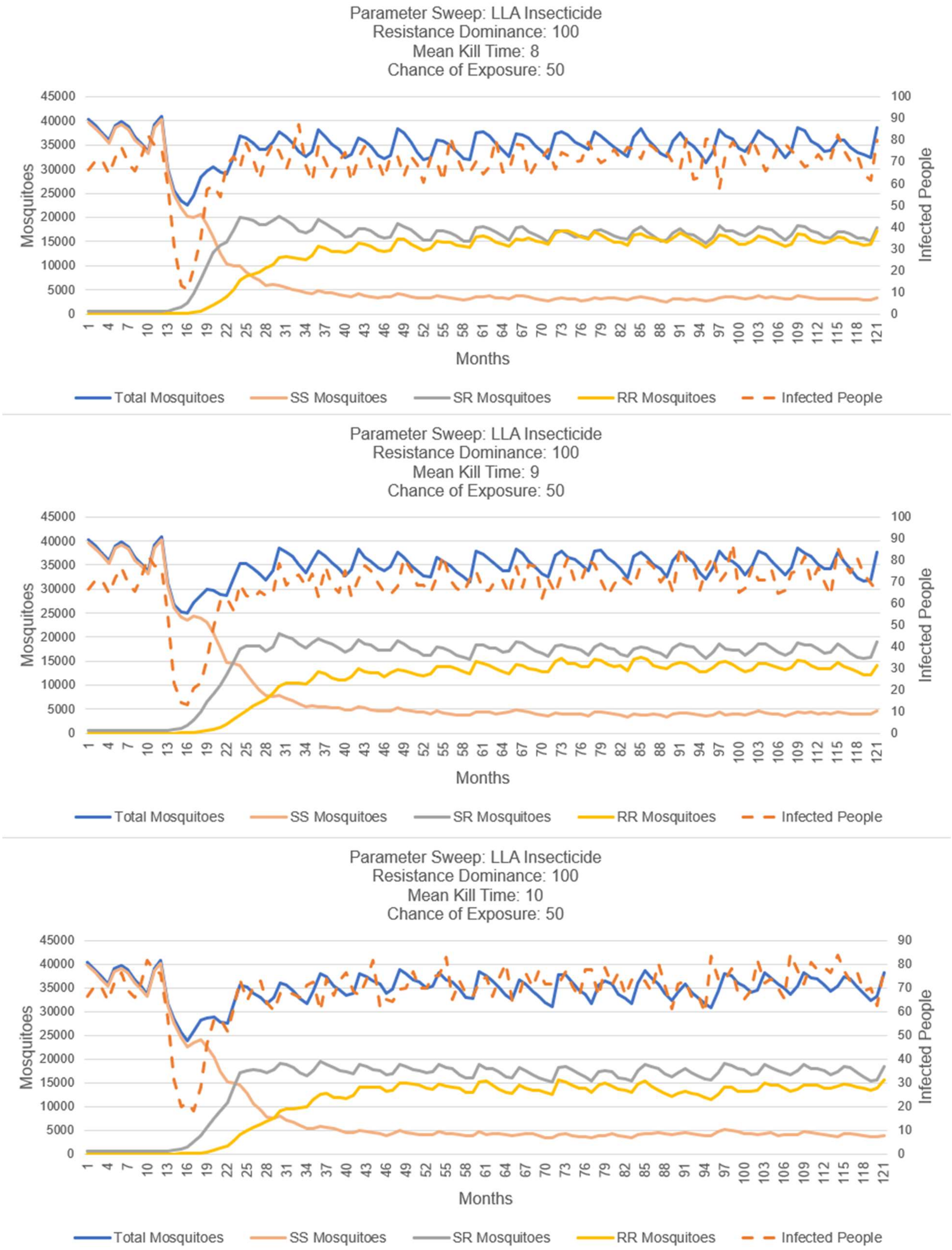

**S4-Fig. 38. LLA insecticide, resistance dominance 100%, chance-of-exposure 50%.**

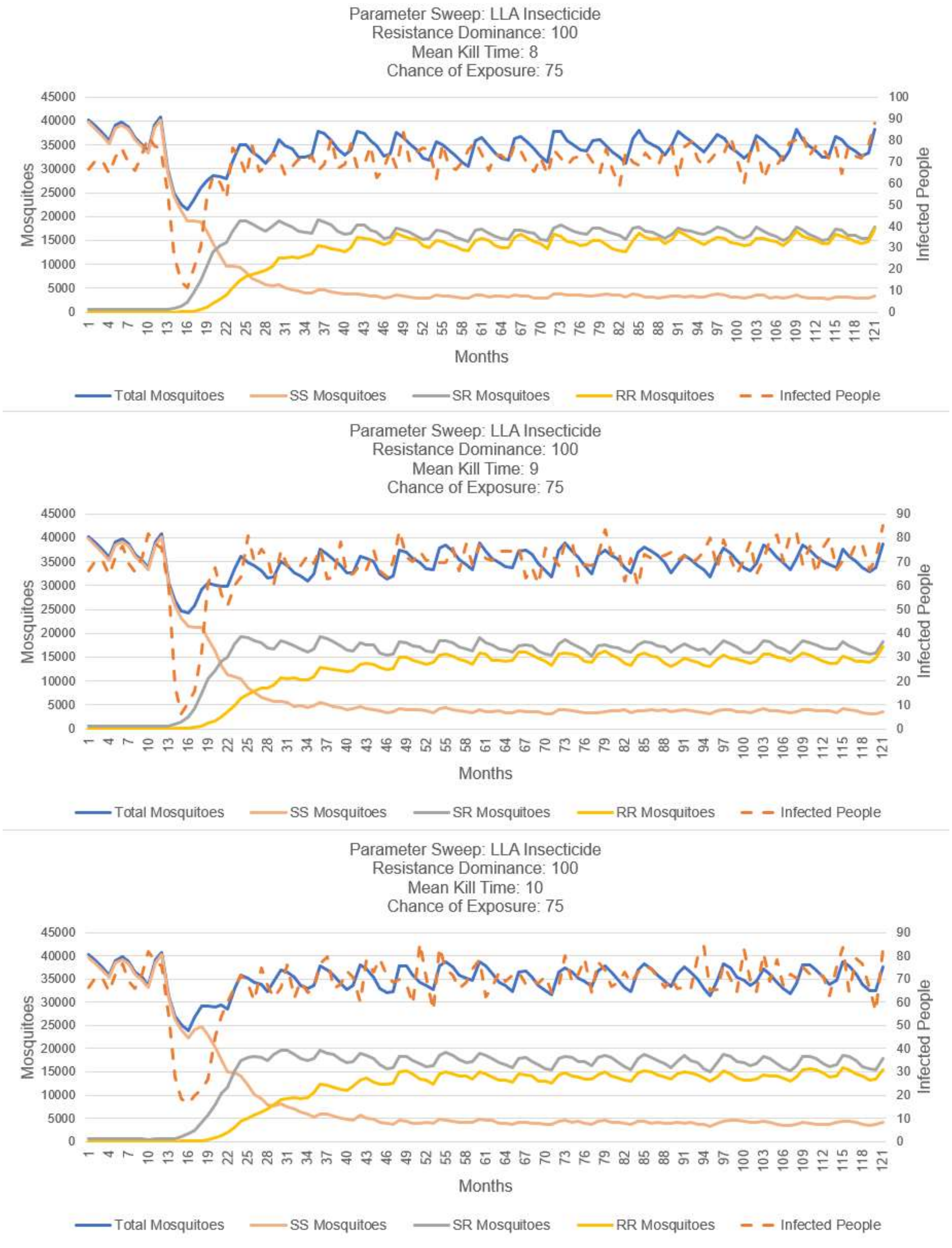

**S4-Fig. 39. LLA insecticide, resistance dominance 100%, chance-of-exposure 75%.**

#### Sensitivity Analysis on Random Number Generator Seed

These experiments display the sensitivity of a simulation run to the value of the starting random number generator seed. The experimental settings are displayed in figure S4-Fig. 40 and the simulation results are displayed in figure S4-Fig. 41. The thin lines show the results of individual runs and the thick lines represent the average of each category of the mosquito and human populations. Additional sensitivity tests of the choice of initial random number generator are presented in figures S4-Fig. 6, S4-Fig. 29 and S4-Fig. 43.

Notes:

- The analysis of the variability of simulation output caused by the initial value of the random number generator's seed is presented.
- The simulation represents the randomness of many events in the transmission of malaria (e.g., probabilities of exposure to an insecticide, the probability that a mosquito blood meal results in an infection, the distribution of mortality of an LLA insecticide, etc.) with calls to a random number generator. As with most such random number generators, an initial seed is required to start a sequence of values which then are used to compute probabilities in the simulation.

Observations:

- The total mosquito population is largely insensitive to the choice of random number generator seed.
- The emergence of the SR genotype is sensitive to the choice of starting random number generator seed, as early as 30 months and as late as 50 months. A similar sensitivity is seen for the RR genotype.
- A corresponding sensitivity of human infections is also observed.
- The display is complex because it displays 10 repetitions plus an average for the mosquito and human categories. For this reason, most the plots presented in this supplement, for clarity reasons, only use one repetition of each experimental setting on the topic being investigated.
- Because of the stochastic nature of the simulation, proper use will require multiple replications and statistical summaries of the results. For an actual application, if more precise values of simulation parameters can be determined, the corresponding stochastic variables can be replaced by fixed values reducing the standard deviations around simulation results.
- We thus conclude that a valid interpretation of the simulation results will require such sensitivity analysis to avoid the possible situation of any one simulation run being an outlier and approximately 10 replications provide a good sample size for estimating the mean and variability of the simulation's output sequences.

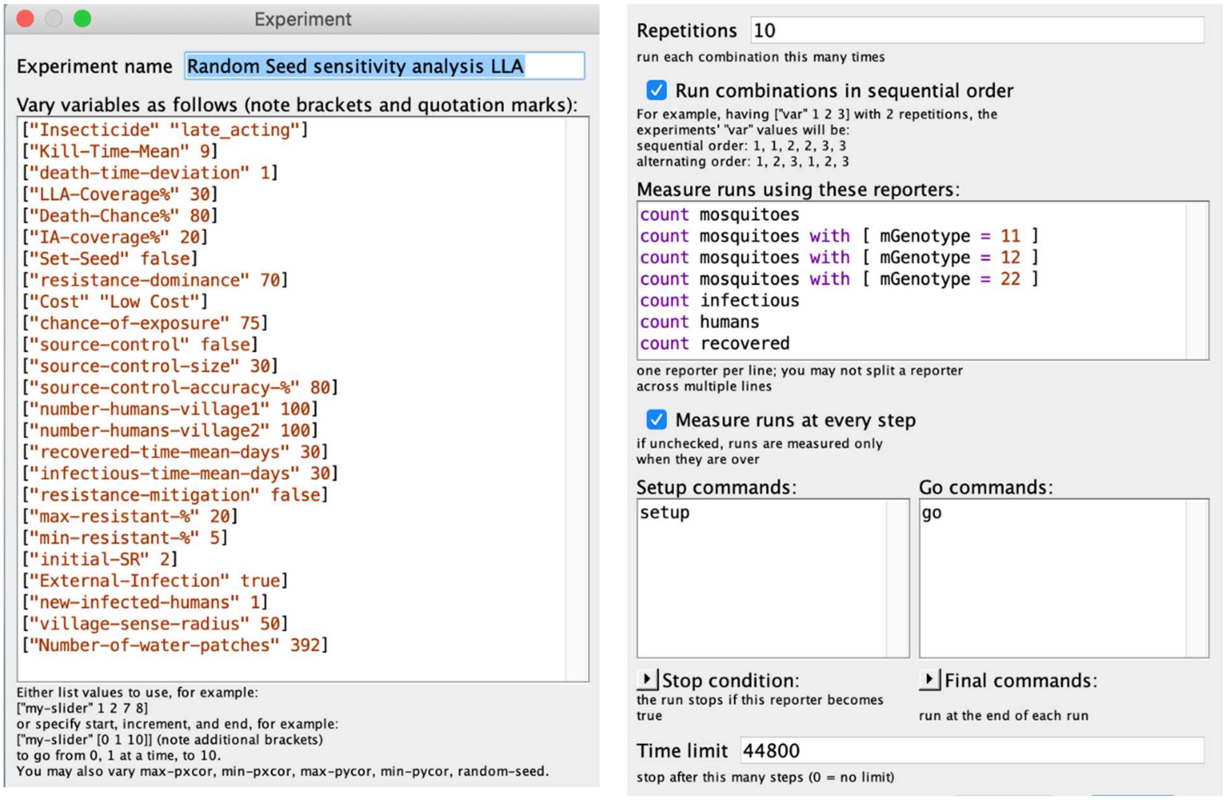

**S4-Fig. 40. Experimental settings determining sensitivity of random number seed.**

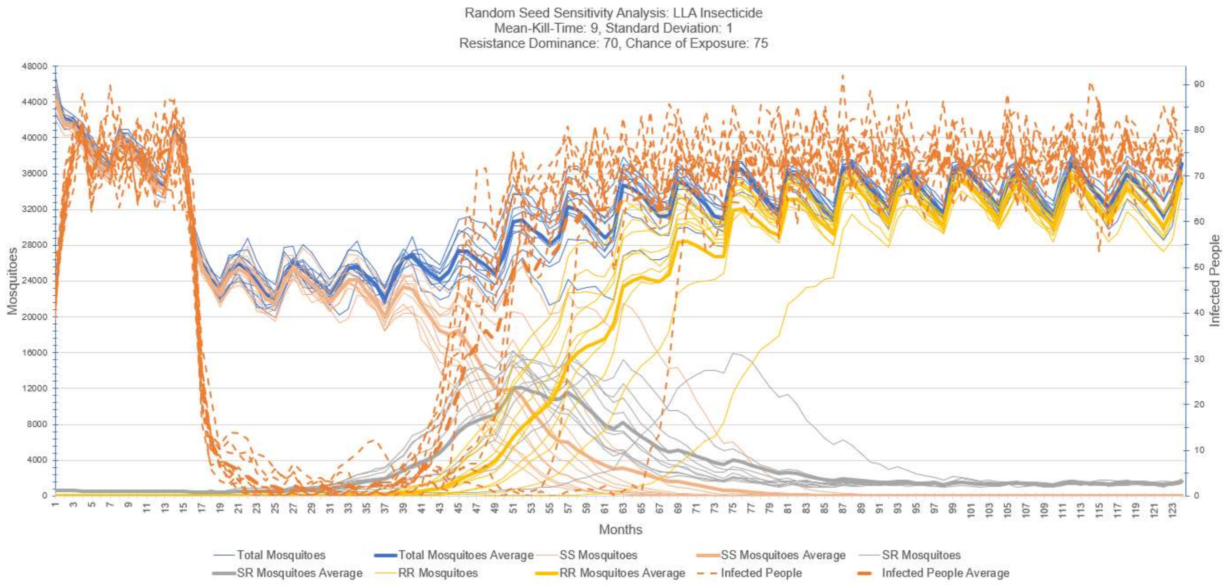

**S4-Fig. 41. Sensitivity analysis – random number generator seed, 10 repetitions**
Displayed are 10 runs of the simulation for one LLA insecticide scenario, all model parameters the same with only the starting seed changing. The sensitivity of simulation outputs based on the starting seed can be observed in the wide range of output time series. The thin graph lines present the simulation output for individual runs. The thick lines report the mean of the output time series over all the 10 runs of the simulation.

### Sensitivity of Standard Deviation of LLA Insecticide Time-to-Death

This section contains the same Fig 7 of the main manuscript that this file accompanies. To assist with replication of results, we add the Behavior Space settings for this experiment. The experimental settings are displayed in S4-Fig. 42 and Fig 7 from main manuscript is here labeled S4-Fig. 43.

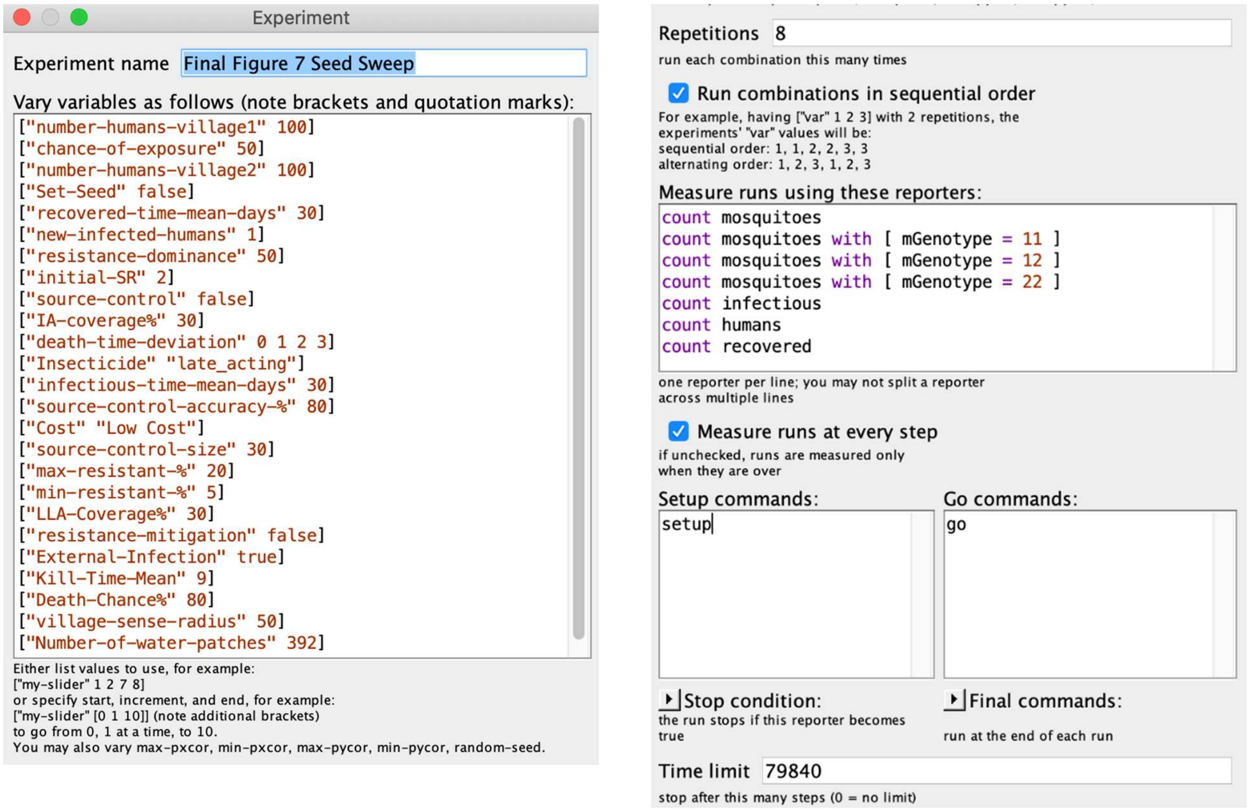

**S4-Fig. 42. Experimental Settings – LLA insecticide Standard Deviation Sweep**

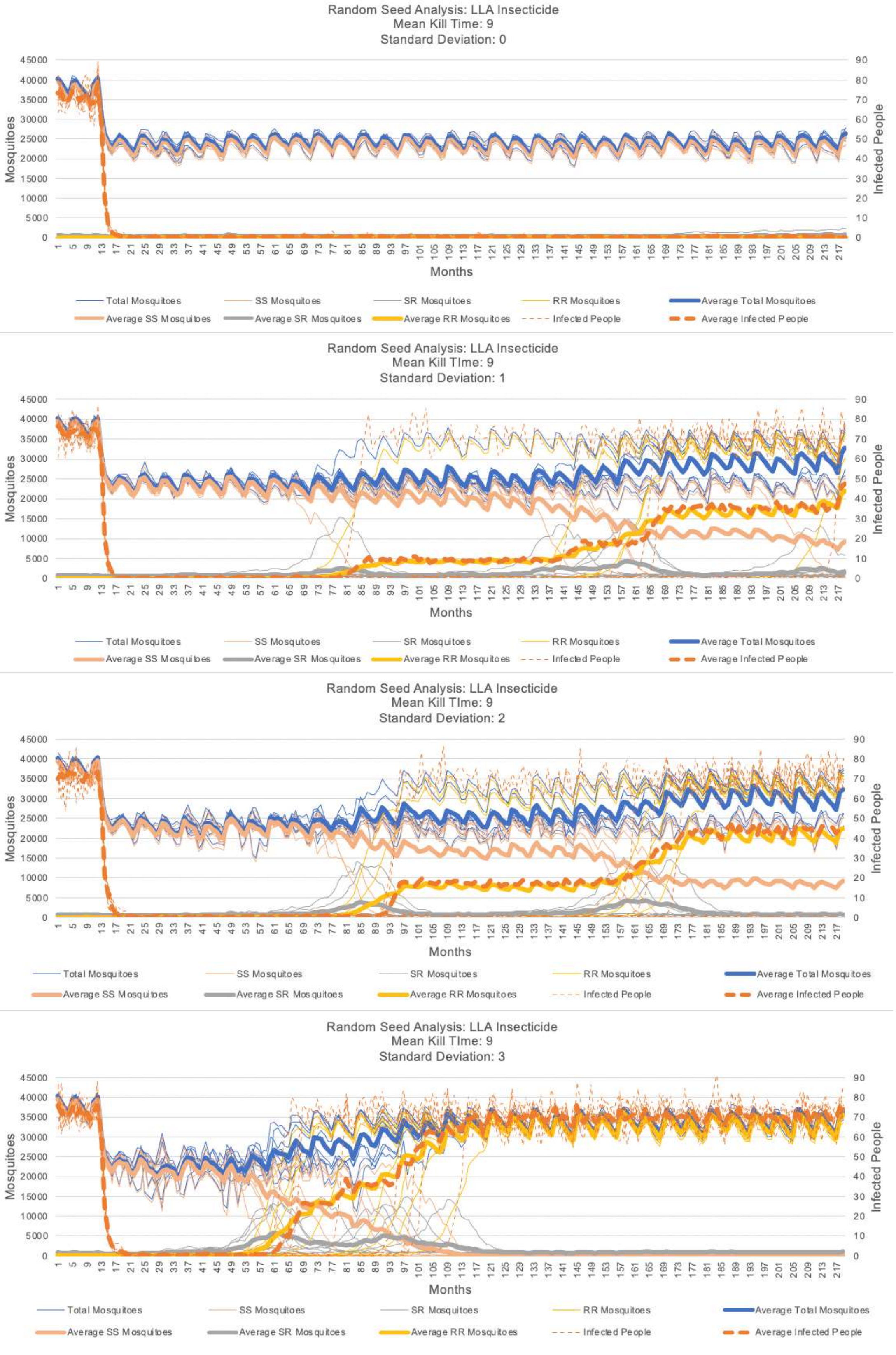

**S4-Fig. 43. Sweep – standard deviation for kill-time-delay for the LLA insecticide.**

### Demonstration of the Extensibility of the Simulation – Larval Source Control Interventions

To demonstrate the extensibility of the simulation, this section provides an example of an added malaria control intervention that was not part of its original design. These plots show the results of a parameter sweep on the distance from the villages where the larval source control feature was applied. When this feature is on, all patches within a specified distance of each village's perimeter (i.e., the annulus around each village) are excluded as oviposition sites (with a specified probability) to simulate the draining and removal of mosquito breeding sites near the villages. The BehaviorSpace experimental settings are displayed in S4-Fig. 44. The area around each village for which larval source control (breeding site reduction) is specified as a distance from the edge of each circular village measured in units of patches: 10, 20, 30, 40, and 50 patches. Plots of each run are displayed in figures S4-Fig. 45 through S4-Fig. 49.

Notes:

- Only one run of the model is presented for each source control coverage value to simplify each plot.
- For each simulated intervention, breeding sites in the target area are removed with 80% accuracy (meaning 20% are not removed) as specified with variable "source-control-accuracy-%".

Observations:

- As the size of source control increases (from 10 patches to 50 patches) we observe: 1) the total mosquito population decreases, 2) the number of infected humans decreases, 3) the number of susceptible humans increases, and 4) the number of recovered humans decreases.
- As the coverage increases the effect is greater; this is the expected trend and supports verification and validation of this component of the simulation.

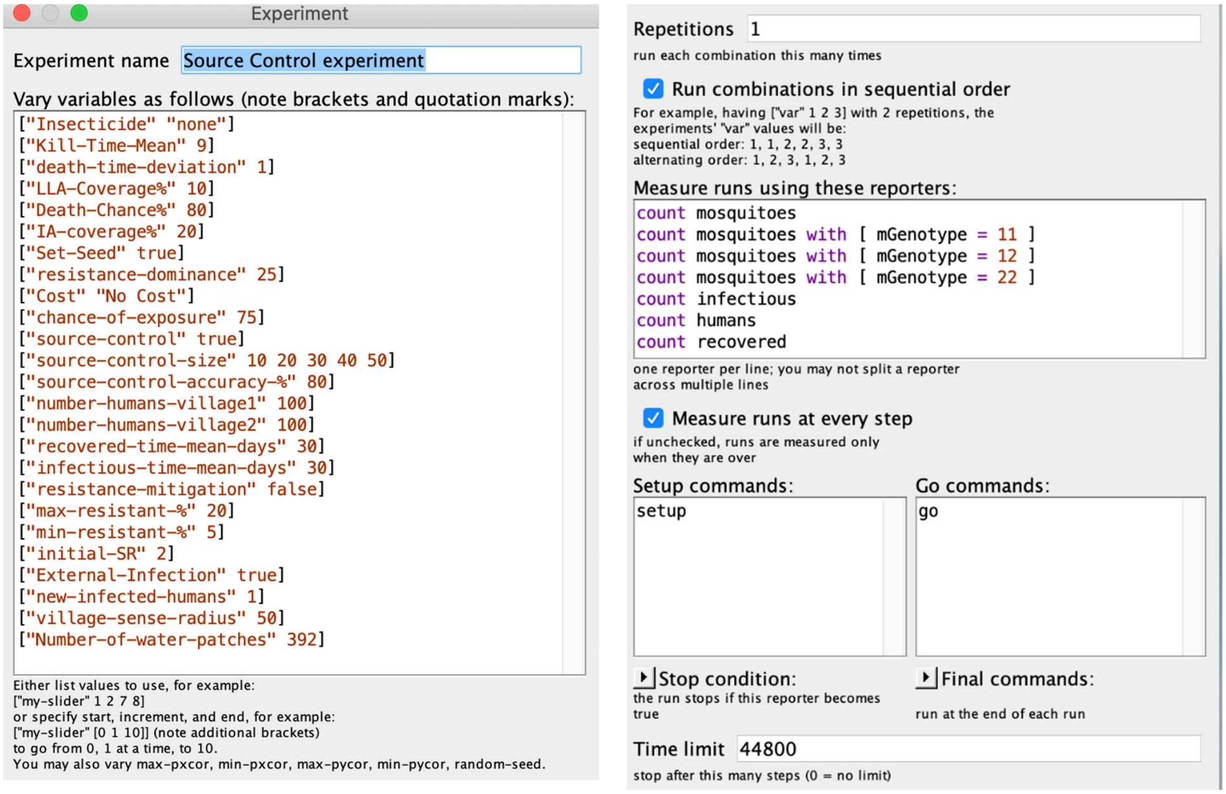

**S4-Fig. 44. Experiment setting for sweep on source control interventions.**

Note: No insecticide applied. Sweep is on distance of the intervention from  the village perimeter

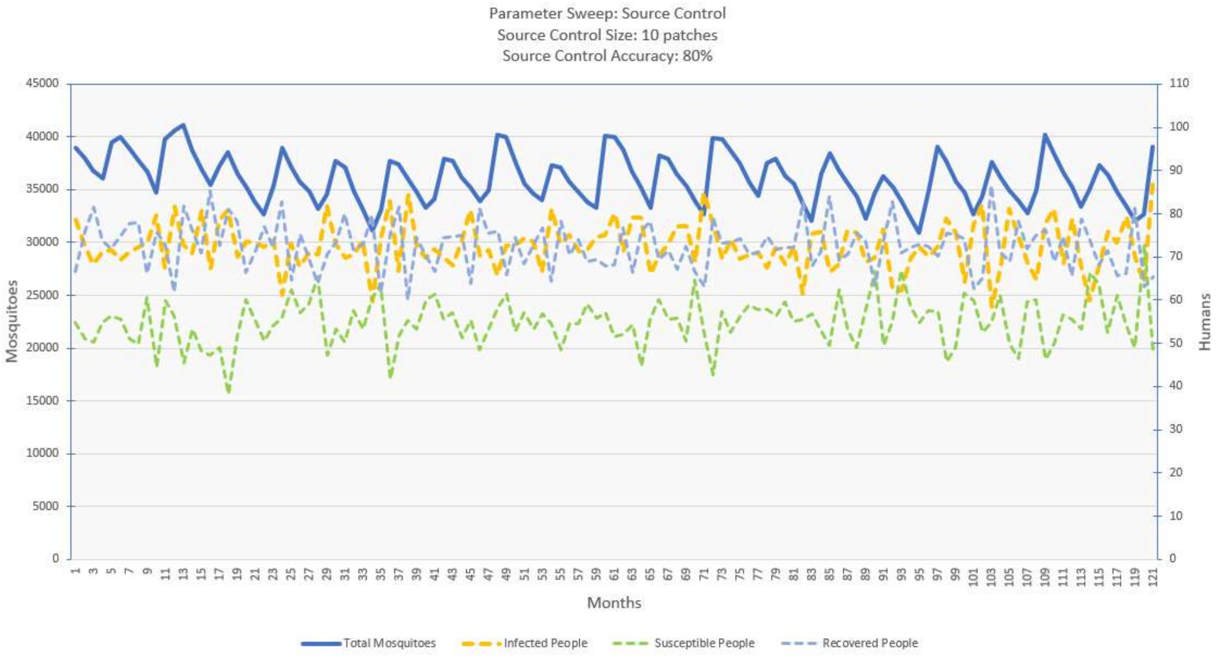

**S4-Fig. 45. Source control intervention – 10 patches around the villages.**
Note: treatment area is an annulus around each circular village.

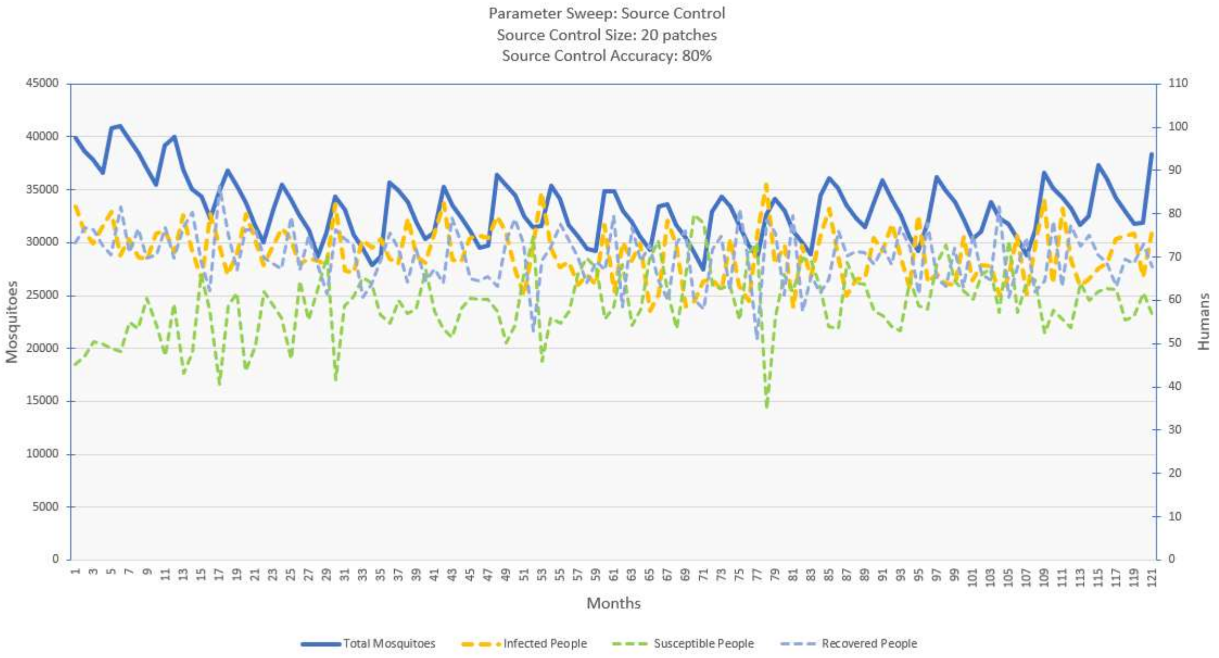

**S4-Fig. 46. Source control intervention – 20 patches around the villages.**
Note: treatment area is an annulus around each circular village.

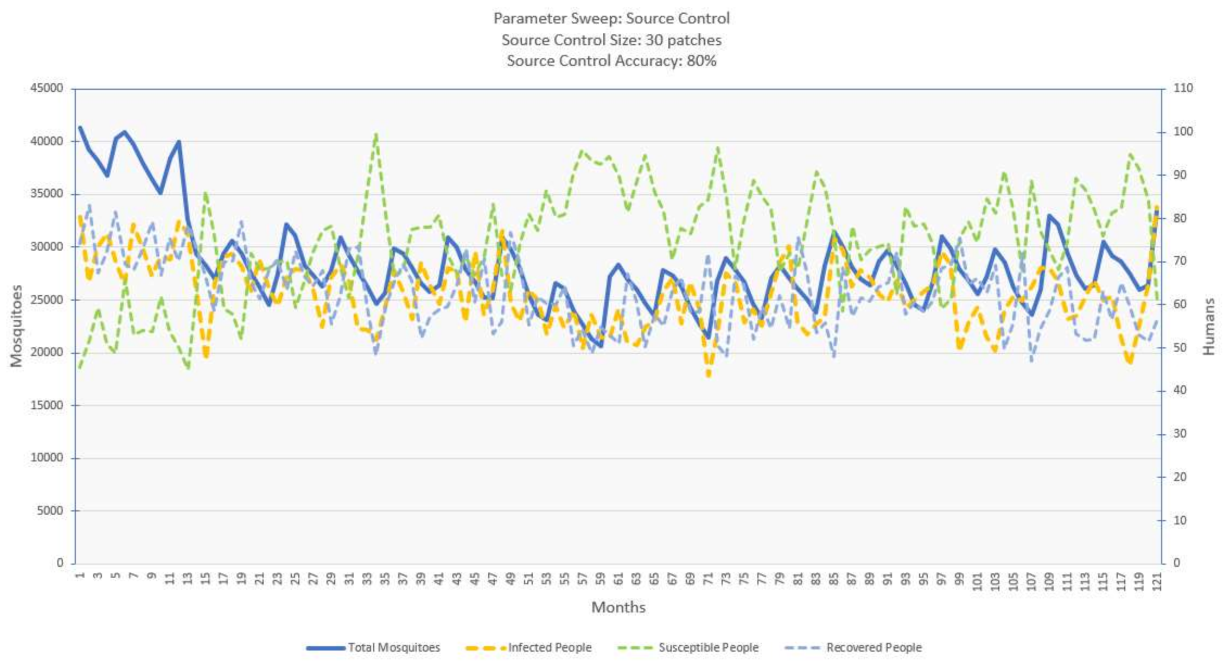

**S4-Fig. 47. Source control intervention – 30 patches around the villages.**
Note: treatment area is the annulus around each circular village.

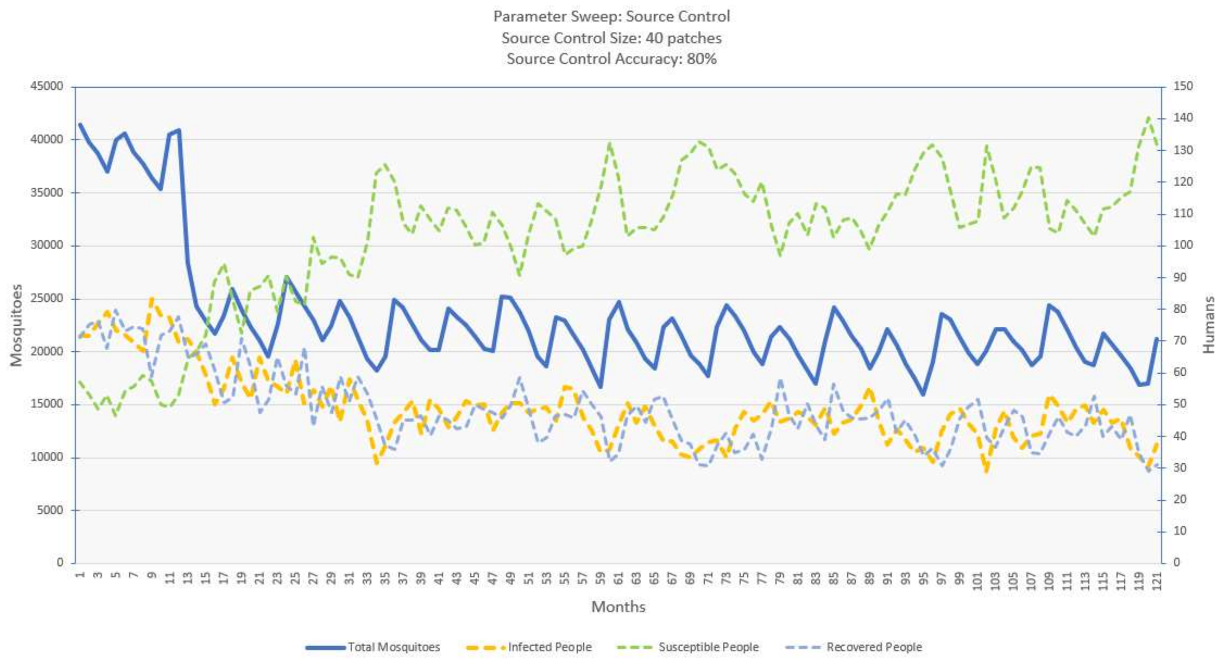

**S4-Fig. 48. Source control intervention – 40 patches around the villages.**
Note: treatment area is an annulus around each circular village.

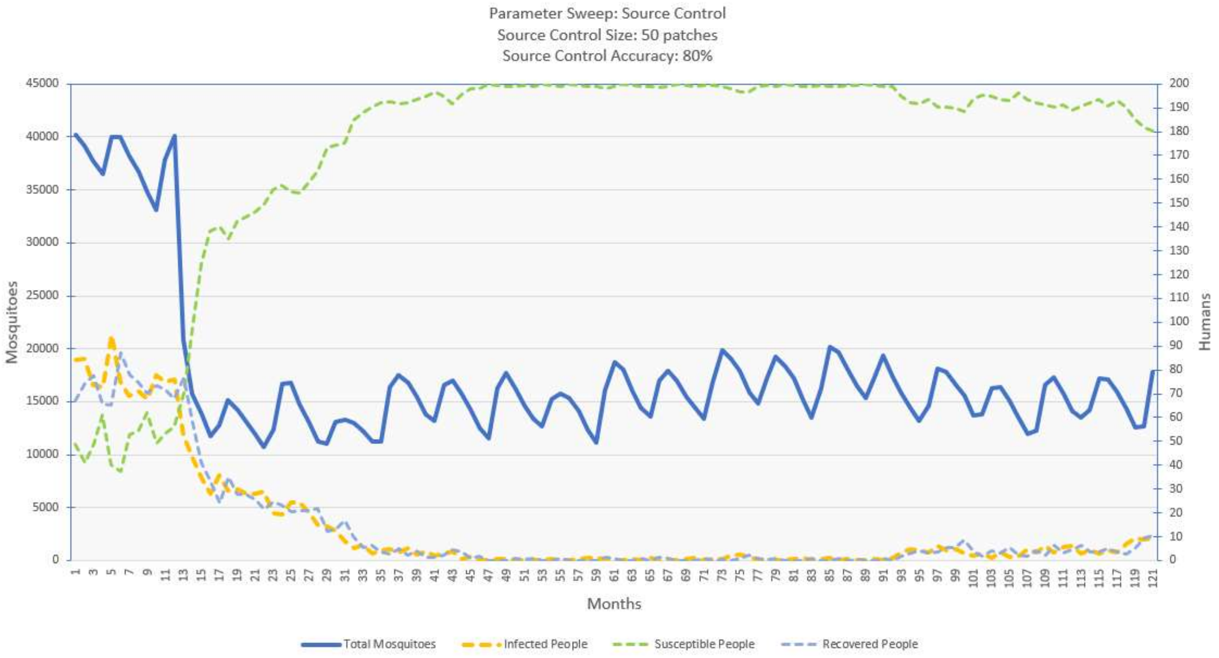

**S4-Fig. 49. Source control intervention – 50 patches around the villages.**
Note: treatment area is an annulus around each circular village.

### Demonstration of the Extensibility of the Simulation – Simulation of an IR Mitigation

To demonstrate the extensibility of the simulation, this section provides an example of an added malaria control intervention that was not part of its original design. Here we have implemented a new feature simulating a policy of "resistance mitigation" which sets a limit on the amount of resistant mosquitoes allowed in the model before the insecticide is removed to prevent the further encouragement of resistance. This intervention requires the cost of the resistant genotype to be high so that the resistant population will decrease after the insecticide's use is discontinued. Once the resistant population reaches a baseline, the insecticide is reintroduced at the original level. To control this process there are two interface controls we added which set the maximum percent of resistance allowed (max-resistant-%) and the baseline percent of resistance (min-resistant-%) which the feature seeks to restore. To demonstrate this process, we have plotted the simulation results using a high resistance dominance to guarantee that there will be resistant mosquitoes to which this feature can respond. We ran these simulations with the max-resistant-% ranging from 20% to 50% and the min-resistant-% set at 5%. See the experimental settings in figure S4-Fig. 50, and the results plotted in figures S4-Fig. 51 and S4-Fig. 52 for the IA and LLA insecticides, respectively. It is important to see how as the max-resistant-% increases the insecticide is applied for longer but then takes longer for the resistant mosquito population to return to the baseline. Additional testing is required to determine the settings for this feature which produce the greatest amount of time with no infected people in the simulation. This feature was added through only a few hours of programming showing the ease and speed of augmenting our simulation.

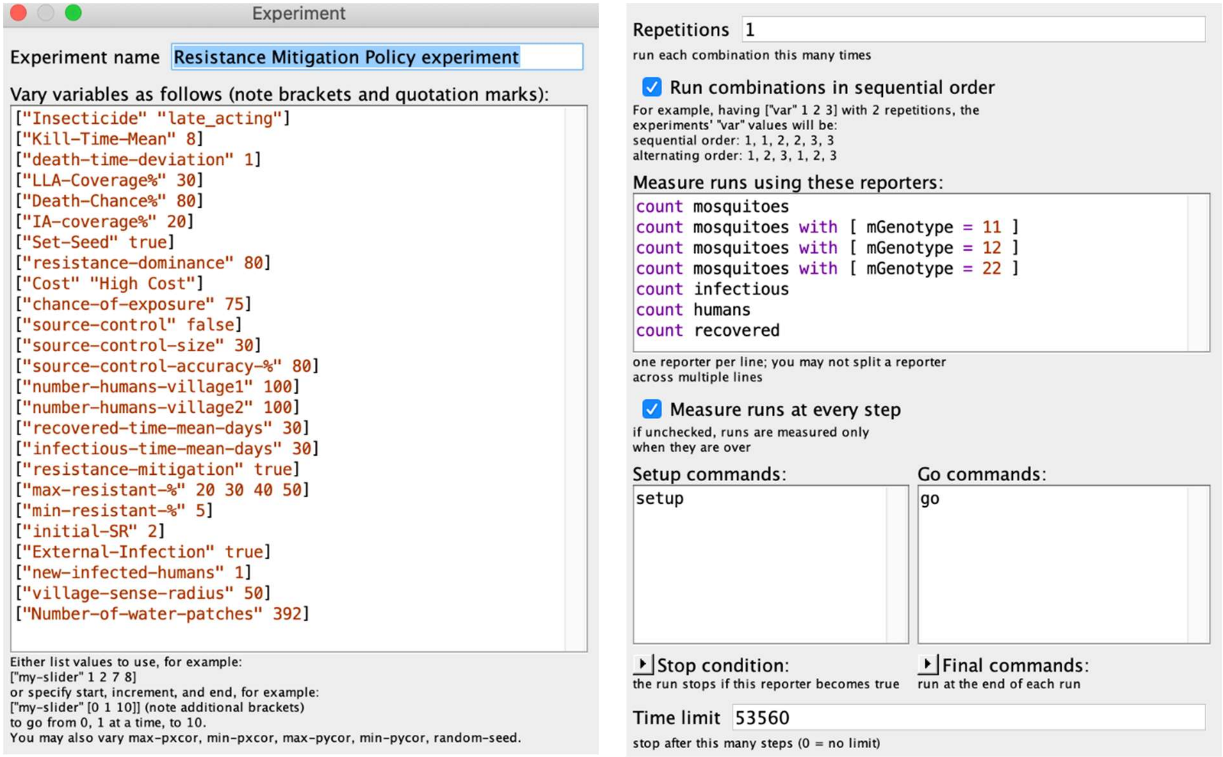

**S4-Fig. 50. Experimental Settings – Simulation of an IR Mitigation.**
Demonstration of simulation extensibility.

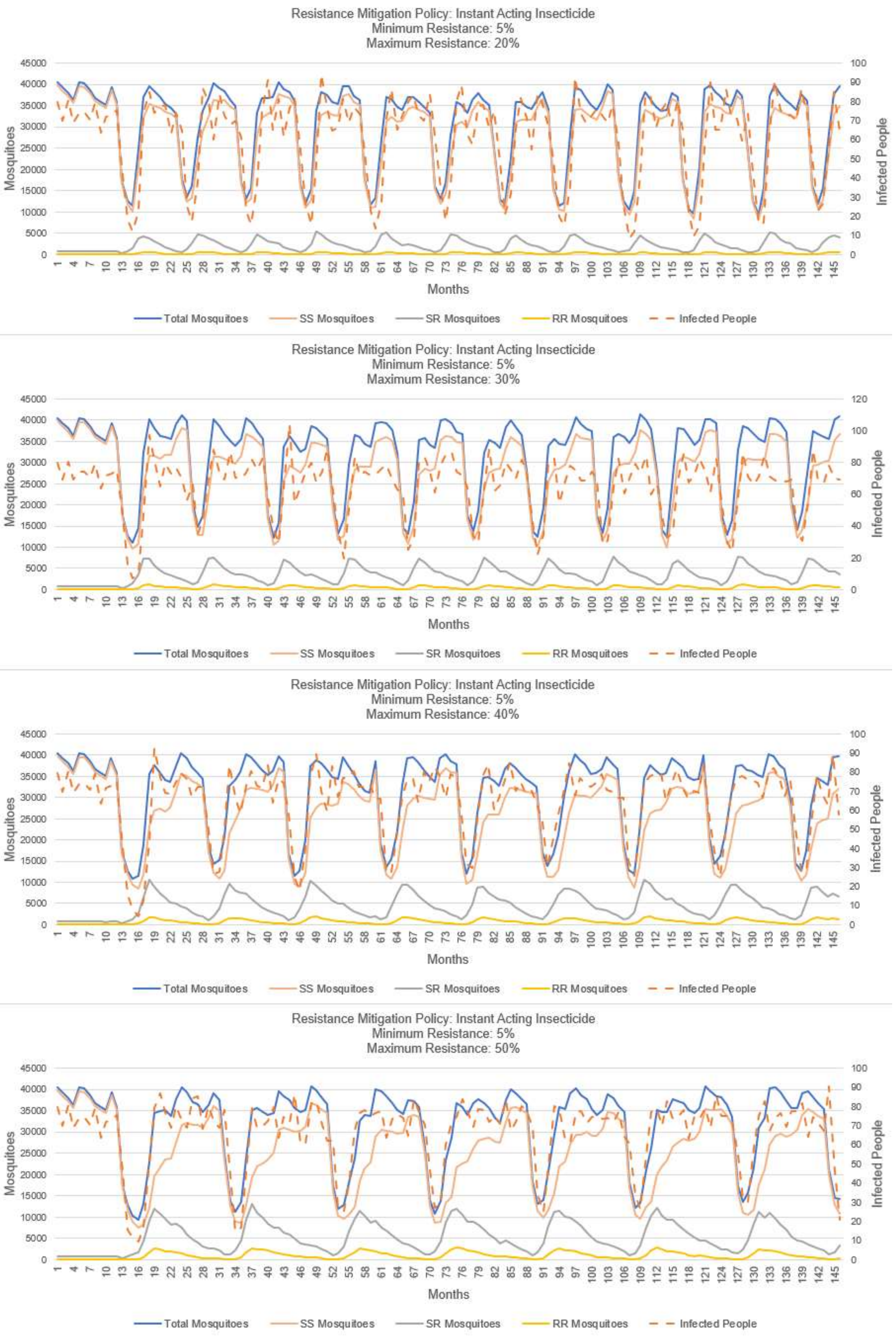

**S4-Fig. 51. Resistance mitigation policy – IA insecticide.**

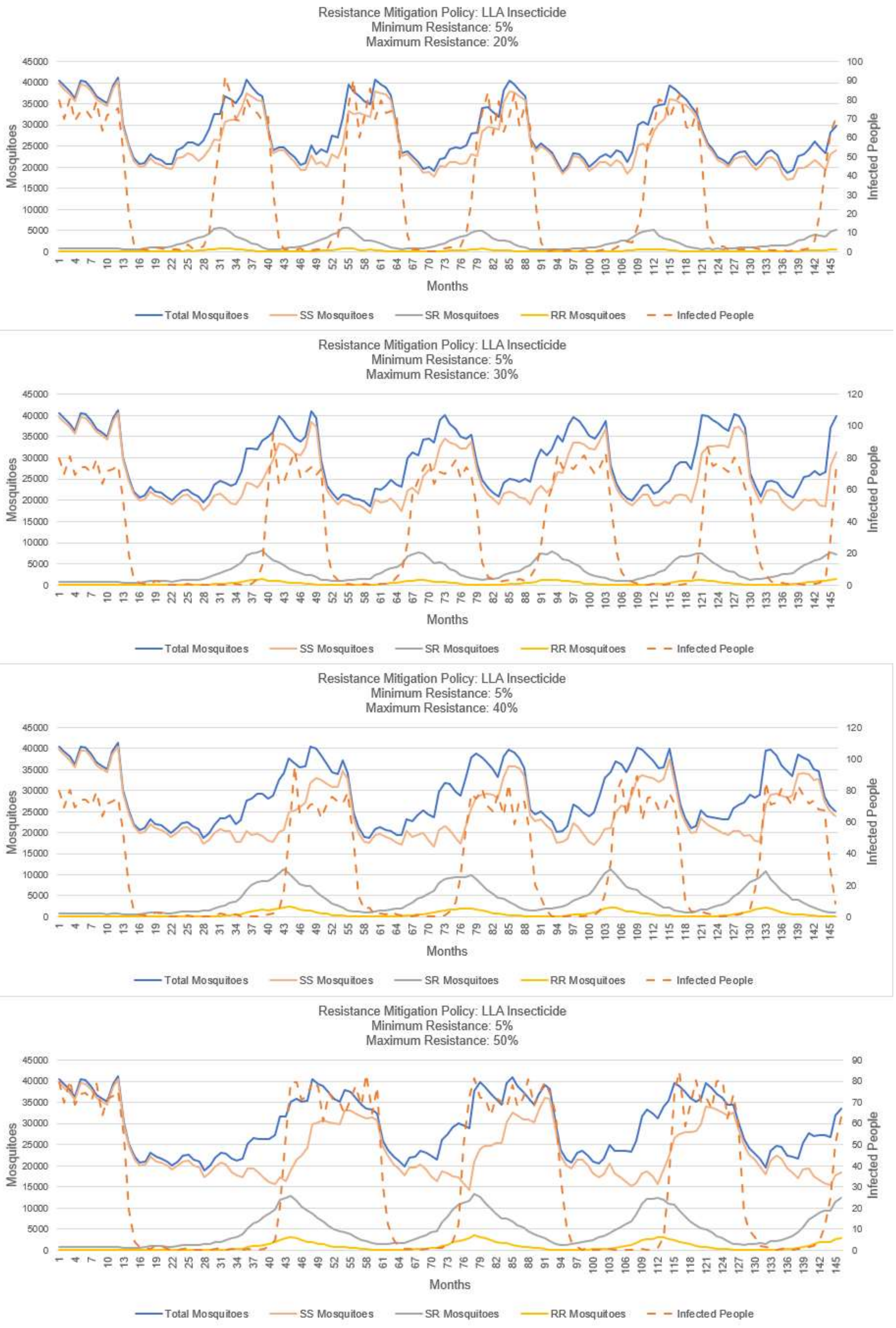

**S4-Fig. 52. Resistance mitigation policy – LLA insecticide.**
